# Creation of a ustekinumab external control arm for Crohn’s disease using electronic health records data: a pilot study

**DOI:** 10.1101/2021.11.12.21266064

**Authors:** Vivek A. Rudrapatna, Yao-Wen Cheng, Colin Feuille, Arman Mosenia, Jonathan Shih, Yongmei Shi, Olivia Roberson, Benjamin Rubin, Atul J. Butte, Uma Mahadevan, Nicholas Skomrock, Ngozi Erondu, Christel Chehoud, Saquib Rahim, David Apfel, Mark Curran, Najat S. Khan, Christopher O’Brien, Natalie Terry, Benjamin D Martini

## Abstract

**Objectives:** The use of external control arms to study treatment effects is growing in interest among drug sponsors and regulators. However, experience with performing these kinds of studies for complex, immune-mediated diseases is limited. We sought to analyze a retrospective cohort of Crohn’s patients to predict the outcome of a prospective cohort.

**Methods:** We queried electronic health records databases and screened records at the University of California, San Francisco to identify patients meeting the eligibility criteria of TRIDENT, a concurrent trial involving ustekinumab as a reference arm. Timepoints were defined to balance the tradeoff between missing disease activity and bias. We compared two imputation models by their impacts on cohort membership and outcomes. We compared the results of ascertaining disease activity using structured data algorithms against manual review. We used these data to estimate ustekinumab’s real-world effectiveness.

**Results:** Screening identified 183 patients. 30% of the cohort had missing baseline data. Two imputation models were tested and had similar effects on cohort definition and outcomes. Algorithms for ascertaining non-symptom-based elements of disease activity were similar in accuracy to manual review. The final cohort consisted of 56 patients. 34% of the cohort was in steroid-free clinical remission by week 24.

**Conclusions:** We predict that a third of the ustekinumab-treated cohort in TRIDENT will be in steroid-free remission by week 24. However, our prediction is limited by substantial missing data. Efforts to improve real-world data capture and align trial design with clinical practice may enable more robust future studies and improve trial efficiency.

**STUDY HIGHLIGHTS:** *WHAT IS KNOWN:*

- External control arm studies are receiving growing interest from drug sponsors and regulators as a potential source of real-world evidence
- However, the feasibility and robustness of this approach is currently limited
- Ustekinumab is an FDA-approved treatment of moderately to severely active Crohn’s disease

*WHAT IS NEW HERE:*

- We derived a retrospective cohort of patients designed to resemble the participants of TRIDENT, a concurrent phase 2b trial using ustekinumab as a reference arm
- We predict that about 34% of the ustekinumab-assigned participants in TRIDENT will be in steroid-free clinical remission by week 24.
- EHR structured data algorithms may be an accurate and less laborious alternative to manual abstraction of non-symptom-based components of the Crohn’s Disease Activity Index
- Real-world practice differs from controlled studies in important ways, including the treatment goals and the timing of encounters
- These differences can pose challenges to the feasibility of external control arm studies, and must be addressed to enable this novel study design
- The methods, data, and code used in this pilot study are shared here for reproducibility and enhancement by others

## INTRODUCTION

The term *external control arm* (ECA) commonly refers to the use of retrospective cohorts to estimate treatment effects by indirect comparison to other cohorts, including those in interventional trials. Recent years have seen a growing interest in this approach with regulatory bodies like the FDA evaluating the use of these data to support claims of treatment efficacy and safety^1,2^. ECAs have some advantages over conventional randomized trials, including lowering the cost of executing trials, greater feasibility for rare diseases, smaller patient populations, or situations involving strong treatment preferences (lack of clinical equipoise)^3^.

Despite this interest, experience in the conduct of ECAs remains limited. To date, most ECAs with regulatory intent have occurred in the space of oncology and rare diseases^3^. These are contexts where treatment paradigms and outcome measures are relatively well-defined, and thus may align well with prospective study designs. However, the feasibility and robustness of ECAs for more common and complex diseases such as inflammatory bowel disease (IBD) remain unknown. There is an unmet need to better understand ECAs and evaluate their viability as an alternative or supplement to prospective trials, which continue to grow in expense and remain challenging to recruit for.

Our own prior work has used electronic health records (EHR) data to estimate the effectiveness of tofacitinib for treating IBD^4^. While this study suggested that EHR data may be useful for this purpose, it was conducted on a cohort whose baseline characteristics substantially differed from the cohorts under study in the pre-approval trials of this drug.

The primary objective of this pilot study was to define and evaluate a method for creating an external control arm for Crohn’s disease. We sought to derive a real-world patient cohort matching the ustekinumab arm of TRIDENT, a concurrent phase 2b trial^5^. Secondary objectives included 1) to evaluate the use of EHR algorithms as an alternative to manual ascertainment of some disease activity variables, 2) to assess the robustness of imputation methods for handling missing data, and 3) to assess ustekinumab’s effectiveness.

## METHODS

### Eligibility

Four inclusionary and seven exclusionary criteria were adapted from the TRIDENT protocol and retrospectively applied to patients at UCSF who received ustekinumab as part of the standard of care (Supplemental Methods). These criteria select for adults with a Crohn’s Disease Activity Index (CDAI) between 220 and 450 and who were stably exposed to other treatments for Crohn’s disease.

All major criteria from TRIDENT were applied except for the exclusion of patients with recent exposure to tumor necrosis factor inhibitors (8 weeks) and integrin inhibitors (16 weeks). This decision was undertaken for two reasons: 1) real-world patients who fail to benefit from one biologic are typically switched to another with minimal delay to avoid the risk of a flare, and 2) patients who sustained long biologic washout periods without being hospitalized or meeting other exclusionary criteria tend to have a lower disease severity and thus tended to be excluded due to a low baseline CDAI.

### Cohort identification

Cohort identification proceeded in six sequential phases.

<u>Phase 1</u>: Using a database of structured EHR data at the University of California, San Francisco (UCSF; 2012-2020), patient records were queried to find those with a medication order for ustekinumab and a Crohn’s disease diagnosis code (ICD-9-CM 555*; ICD-10-CM K50*) anywhere in their care timeline.

<u>Phase 2</u>: Eligible records were then divided among four trained chart abstractors for additional manual screening. Chart abstraction was performed under the supervision of the principal investigator using a pre-defined protocol (Supplemental Methods).

Initial manual screening was performed to identify patients meeting all inclusion and exclusion criteria except for that pertaining to the baseline CDAI. The process of manual abstraction included confirmation of the informatics-based criteria used for initial screening (i.e. confirmation of the diagnosis, treatment with ustekinumab, etc.).

<u>Phase 3</u>: Patients who met all study criteria prior to the assessment of the baseline CDAI were identified. From this cohort, the baseline patient-reported outcome elements of the CDAI (abdominal pain, diarrhea, wellbeing; PRO3) were abstracted using a time window of up to 16 weeks prior to the date of the first dose of ustekinumab. The data were analyzed to empirically define the baseline period. This corresponded to the narrowest window of time prior to week 0 that was not associated with a substantial increase in missing data.

<u>Phase 4</u>: The non-PRO3 elements of the CDAI were abstracted by two methods: manual abstraction and informatics criteria (Supplemental Methods). This process was done by one of the four chart abstractors, corresponding to roughly one quarter of the still eligible cohort. All non-PRO3 CDAI elements were abstracted at the baseline period (week -12 to 0) and compared to the informatic approach.

Manual abstraction was performed on a subset of these variables at post-baseline time periods to assess the accuracy of informatics-based ascertainment across time. These variables included the use of antidiarrheals or opiates (binary), the use of steroids (binary), and hematocrit (numeric). Post-baseline time periods corresponded to the times of the primary and secondary endpoints of TRIDENT.

<u>Phase 5</u>: Informatics-based criteria were used to ascertain all non-PRO3 CDAI elements at the baseline period among the 183 patients (25% of the initial cohort) who met all other criteria.

Given missing CDAI elements at baseline, imputation was required to determine which individuals met the baseline CDAI requirement and thus could be confirmed as members of the cohort. Two random forest-based imputation routines (*MissForest, MissRanger*) were used to assess the sensitivity of cohort membership to the choice of imputation model.

<u>Phase 6</u>: The *MissForest* algorithm was used to finalize cohort membership using the same method for calculating the CDAI as was used in TRIDENT (Supplemental Methods).

### Number of subjects

The sample size calculation in the TRIDENT protocol specified 50 subjects per arm. This study analyzed 56 patients.

### Endpoints

Endpoints included the mean reduction in CDAI at week 12 (TRIDENT primary endpoint) and week 24. Of note, subjects in TRIDENT who entered the study on glucocorticoids were required to remain on them during the induction period. Because real-world clinical practice involves tapering these medications earlier, steroid use and steroid-free clinical remission (CDAI _≤_ 150) at weeks 12 and 24 were also included as efficacy endpoints.

Time windows were used to approximate the true week 12 (weeks 10-14) and week 24 (weeks 20-28) relative to the date of ustekinumab initiation. These windows decided upon and fixed a priori, guided by prior work^4^.

Manual review was used to abstract PRO3 elements corresponding to these time windows, and informatics-based criteria were used for the non-PRO3 elements of the CDAI. A second model using *MissForest* was used to impute any missing values across time. These data were then analyzed to estimate the efficacy of ustekinumab according to the above endpoints.

### Safety

Safety was not assessed. No incidental findings of possible adverse drug reactions were identified.

### Statistics

This was an estimation study; no statistical hypotheses were tested. Binary outcomes are reported numerically and as a proportion. Numeric outcomes are reported by the mean and standard error. Statistical computing was performed in *R*.

### Ethics/Compliance

This retrospective study was approved by the UCSF Institutional Review Board (#20-31760).

## RESULTS

In this study, we used a combination of informatics methods and manual review to identify a real-world cohort with Crohn’s disease who met the eligibility criteria of TRIDENT and were treated with ustekinumab. From this cohort, we abstracted and analyzed their post-treatment outcomes according to a number of endpoints including the TRIDENT primary endpoint.

### Cohort identification

The cohort selection process is outlined in Figure 1, and further described according to the phase of the study.

**Figure 1:**
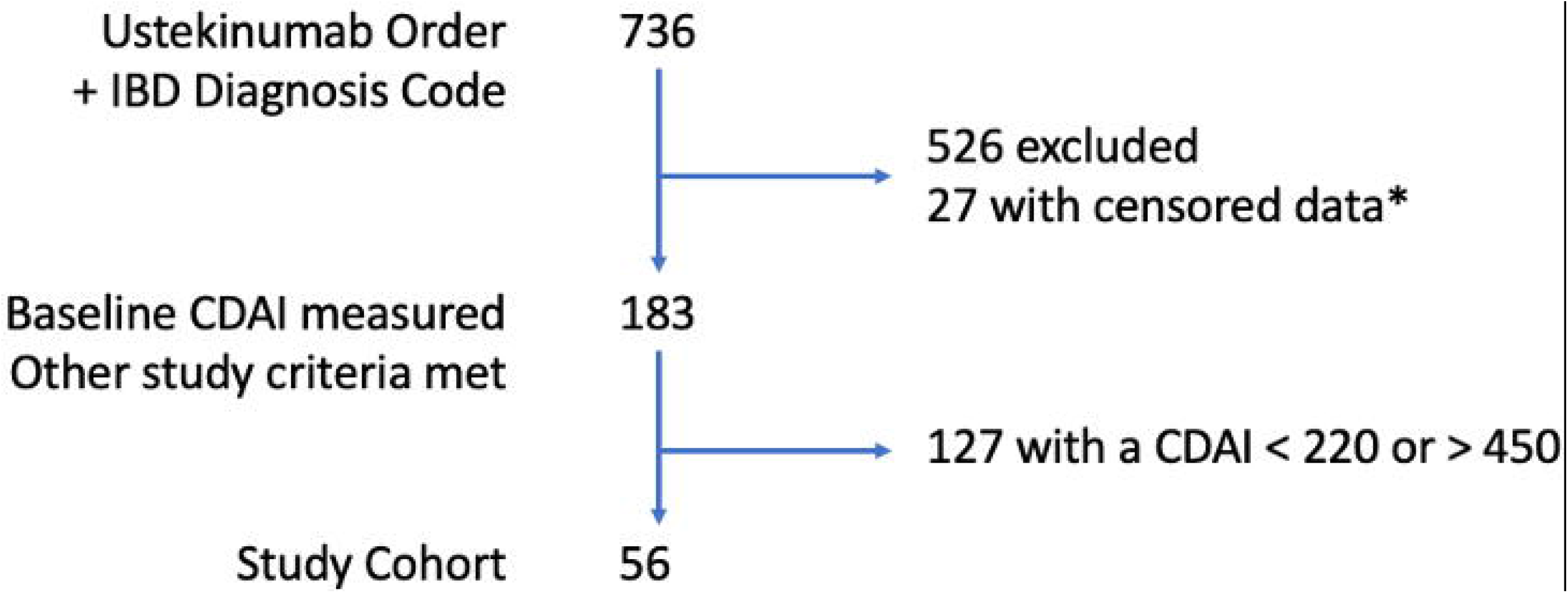
Study flow diagram. 736 patient records were screened for inclusion, 56 records met all study criteria. *: Informatically-censored patients at unique risk for re-identification or who withdrew consent for EHR-based research.

*Phases 1 and 2*: At the time of our EHR database query (April 2020), 736 patients were identified as having an ustekinumab medication order and a Crohn’s disease diagnosis code. These records were then manually screened according to the study eligibility criteria. 526 patients failed to meet at least one TRIDENT eligibility criterion. These included the presence of a low baseline CDAI (prevention of post-operative Crohn’s recurrence), as well as the initiation of steroids to treat active disease while awaiting ustekinumab induction.

*Phase 3*: PRO3 elements were manually abstracted at timepoints ranging from -16 to 0 weeks relative to the date of ustekinumab initiation. An analysis of data availability according to differently sized time windows at baseline supported the use of a 12-week window (Figure 2). Data was commonly missing at time points close to the date of ustekinumab induction, reflecting gaps of time between clinic visits where treatments were decided upon and the patient receipt of intravenous ustekinumab. When clinic visits occurred, we found that all three PRO elements tended to be documented together. These were more commonly available than lab testing such as c-reactive protein.

**Figure 2:**
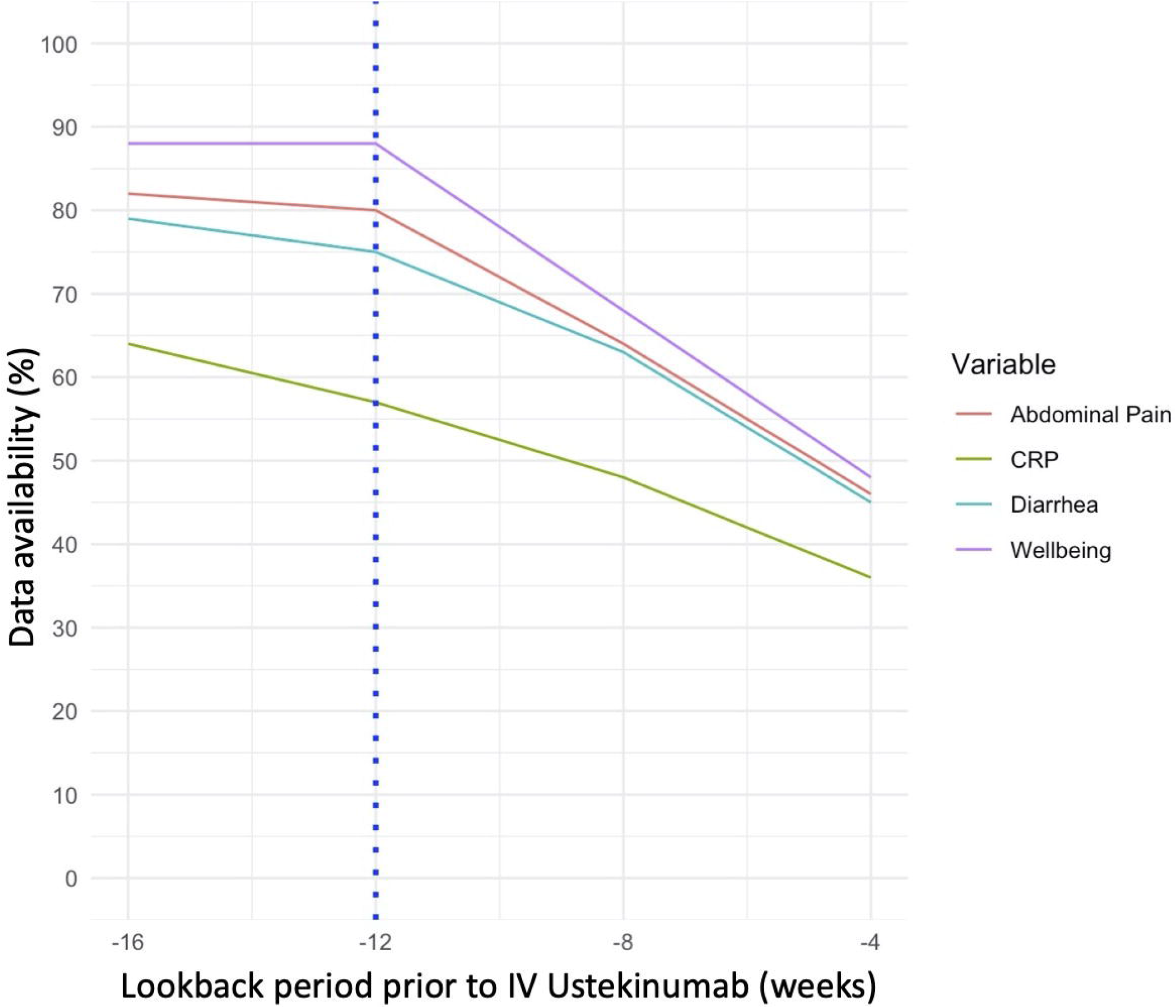
Impact of different definitions for the baseline time period on variable availability. These results correspond to the 183 patients who were not excluded by non-CDAI related study criteria. All of these patient records underwent manual review to ascertain the presence of PROs occurring during the 16 week period prior to the first dose of ustekinumab (week 0). The blue dotted line corresponds to the cut point (12 week lookback) that was selected following a visual review of the trends depicted here.

*Phase 4*: Non-PRO3 elements were abstracted by manual and informatics-based approaches on a sample of the still eligible cohort (N=183). The accuracy of CDAI components as abstracted informatically as well as the CDAI itself was generally high compared to manual review (Figure 3). The degree of agreement, measured by Pearson’s r^2^, ranged from 0.91 to 0.96 across timepoints (Table 1, Supplementary Data Tables 1 and 2). This high correlation appeared to be driven by the fact that for most patients, the major contributor to the total CDAI came from PRO3 elements, not the other elements ascertained by informatics query.

**Figure 3:**
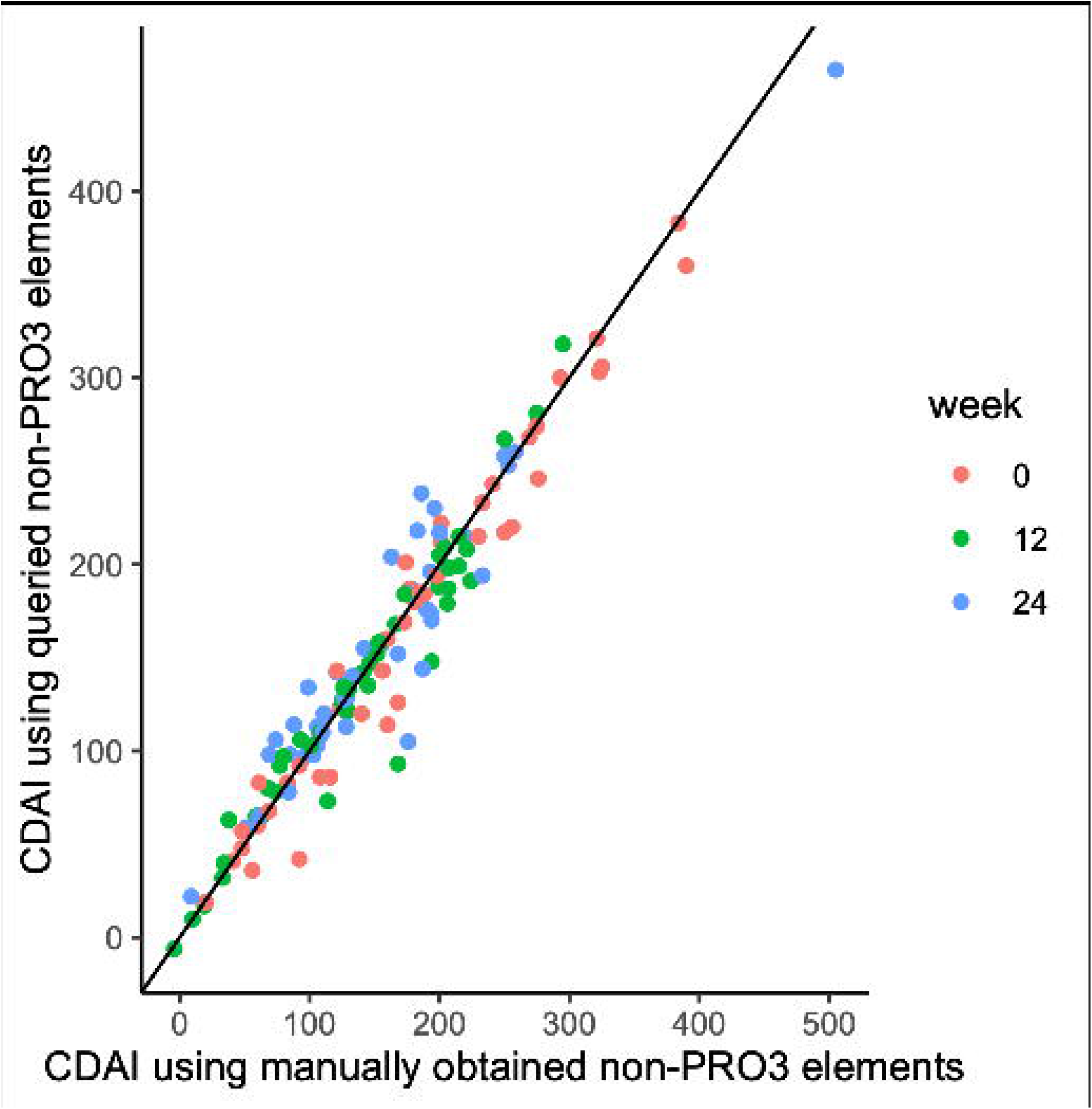
Comparison of total CDAI as computed using CDAI elements ascertained informatically vs manually. The y-axis corresponds to the CDAI calculated by database query of non-PRO3 elements. The x-axis corresponds to the CDAI calculated by manual abstraction.

**Table 1:** Comparison of total CDAI as computed using CDAI elements ascertained informatically vs manually. All results correspond to the results of manual abstraction by one chart reviewer (45 patients).

| Time | Mean Absolute Error | Pearson's $r^2$ | Steroid-free Remission (manual) | Steroid-free Remission (informatics) |
| --- | --- | --- | --- | --- |
| Baseline | 13 | 0.96 | 0/14 | 0/14 |
| Week 12 | 12 | 0.93 | 5/14 | 6/13 |
| Week 24 | 17 | 0.91 | 6/14 | 5/13 |

**Table 2:** Characterization of missing PRO3 elements at baseline. Proportions correspond to the 183 patients who met all eligibility criteria prior to application of the baseline CDAI requirement.

| Variable | Missing data |
| --- | --- |
| Abdominal Pain | 27% |
| Diarrhea | 30% |
| Wellbeing | 20% |

*Phase 5*: 30% of the cohort was missing at least one PRO3 element at baseline (Table 2). To handle this missing data and determine cohort membership, we compared two methods (*MissRanger, MissForest*) for performing single imputation. These random forest-based methods differ in their modes of optimization as well as their final models, which are fit according to a stochastic process. The two models gave very similar results relevant to the selection of the baseline cohort and their outcomes (Table 3).

**Table 3:**
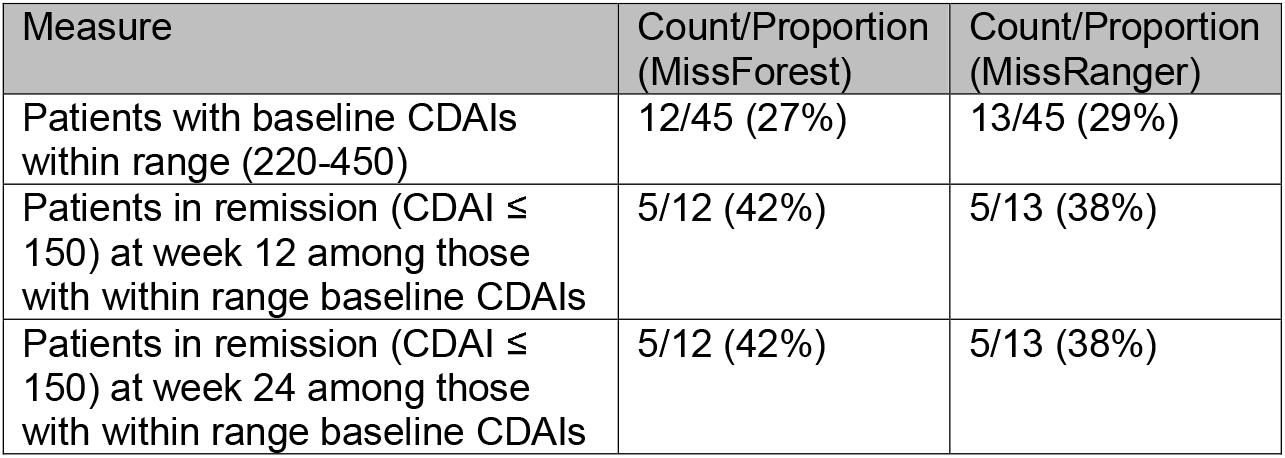
Comparison of the results of two imputation models. Imputation models were applied to all of the otherwise eligible patients (less the baseline CDAI requirement) that had been assigned to one chart abstractor (45 patients).

*Phases 6*: The *MissForest* algorithm was used to identify 56 patients meeting all study criteria (Table 4). Post-baseline PRO3 abstraction was selectively performed in just these patients, and the previously described informatics algorithm was used to abstract other post-baseline variables. To handle post-baseline missing data (Table 5), a second imputation model using *MissForest* was applied to complete the dataset across timepoints and enable an assessment of patient outcomes.

**Table 4:** Characterization of the Study Cohort. *: Reported as median, other continuous variables reported as mean ± standard deviation.

|  | UCSF External Control Arm (N=56) |
| --- | --- |
| Age in years | 34 ± 14 |
| Female Sex | 59% |
| Disease Duration | 10 ± 11 |
| C-Reactive Protein* | 12.3 |
| History of Biologic | 89% |
| Intolerance or Refractoriness |  |
| Disease Location: |  |
| • Ileal | 13% |
| • Ileocolonic | 52% |
| • Colonic | 30% |
| • Other | 5% |
| Baseline CDAI | 305 ± 51 |
| Baseline steroid use | 38% |

**Table 5:** Characterization of missing PRO3 elements across all time periods. Proportions correspond to all 56 members of the external cohort.

| Variable | Time | Missing data |
| --- | --- | --- |
| Abdominal Pain | Baseline | 25% |
| Diarrhea | Baseline | 27% |
| Wellbeing | Baseline | 13% |
| Abdominal Pain | Week 12 | 59% |
| Diarrhea | Week 12 | 77% |
| Wellbeing | Week 12 | 46% |
| Abdominal Pain | Week 24 | 61% |
| Diarrhea | Week 24 | 66% |
| Wellbeing | Week 24 | 43% |

### Efficacy

Ustekinumab was associated with a 95 point mean reduction in the CDAI by week 12, and a 133 point reduction by week 24 (N=56; Figure 4, Table 6). The proportions of patients in steroid-free clinical remission were 23% and 34% at Weeks 12 and 24 respectively. 38% of the cohort had been using steroids at the baseline timepoint. Out of the cohort of 56, 7 (13%) and 9 (16%) remained on steroids at weeks 12 and 24 respectively (Table 7).

**Figure 4:**
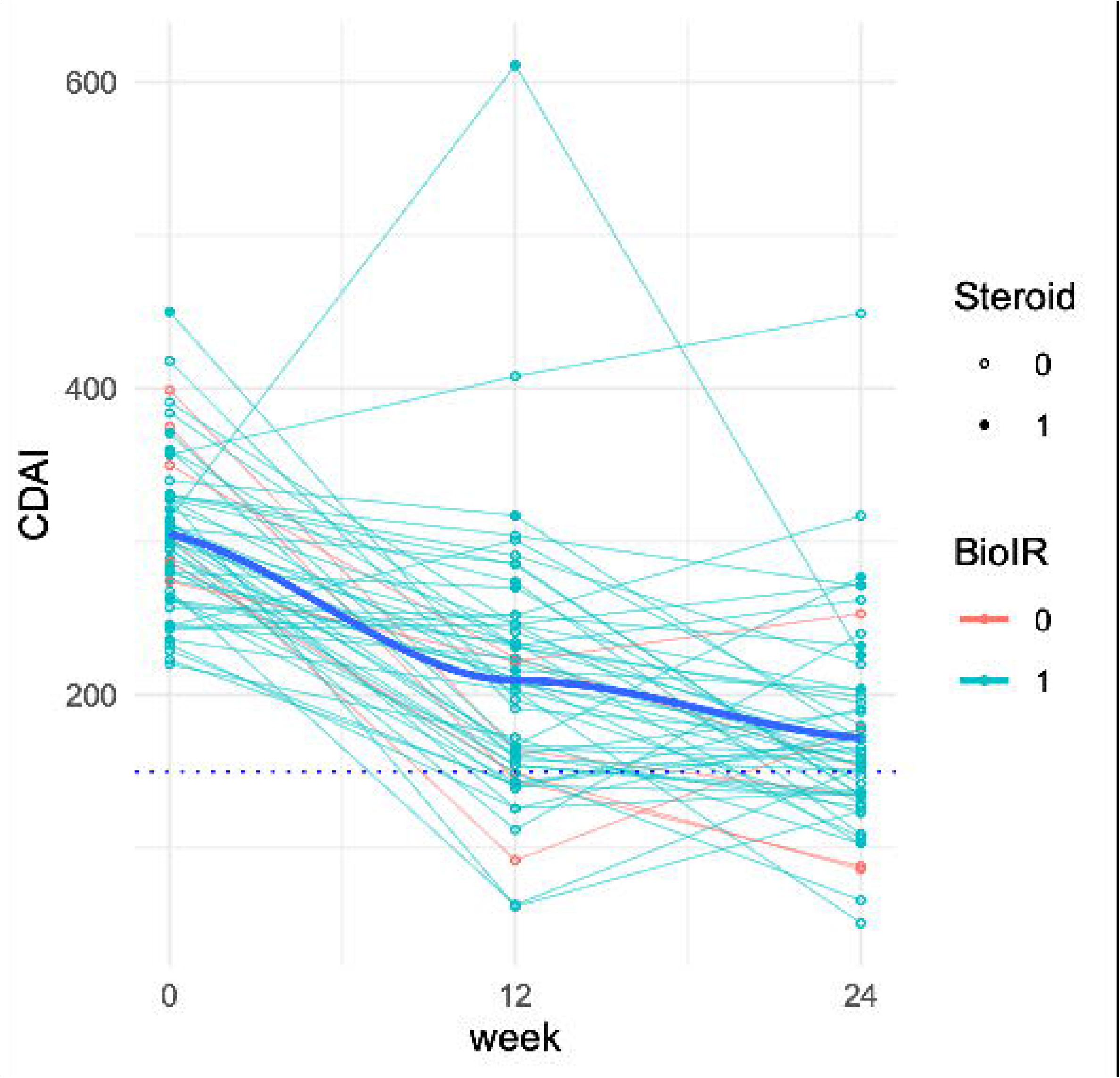
Change in CDAI over time. Thin lines correspond to individual CDAI trajectories over time. These are colored turquoise if the patient had a history of biologic intolerance or refractoriness at the time of ustekinumab induction and is colored red otherwise. At each timepoint, closed circles correspond to the use of steroids at a given point of time, open circles otherwise. The thicker blue line depicts the results of LOESS regression.

**Table 6:** Efficacy Endpoints.

| Outcome | Numerical Result [Mean ± SE, N/D (%)] |
| --- | --- |
| CDAI Reduction by Week 12 | 95 ± 13 |
| CDAI Reduction by Week 24 | 133 ± 11 |
| Proportion in steroid-free remission (CDAI ≤ 150) at Week 12 | 13/56 (23%) |
| Proportion in steroid-free remission (CDAI ≤ 150) at Week 24 | 19/56 (34%) |

**Table 7:** Steroid use across timepoints.

| Time | Proportion (%) |
| --- | --- |
| Baseline | 21/56 (37%) |
| Week<br>12 | 7/56 (13%) |
| Week<br>24 | 9/56 (16%) |

## DISCUSSION

In this pilot study, we used a combination of manual review, EHR data informatics, and imputation modeling to establish a method for creating external control arms for Crohn’s disease. We applied this method to identify a real-world cohort resembling the ustekinumab arm in TRIDENT, a concurrent phase 2b trial. We found that algorithms utilizing EHR structured data were accurate at ascertaining the CDAI (both components and in aggregate) and may be a favorable alternative to manual review for non-PRO3 components. We found a substantial amount of missing data in the context of retrospective use for this study design. However, our results suggested that different imputation models may be equivalent in their impacts on cohort definition and outcome measurement. Lastly, this observational cohort appeared to demonstrate a plausible improvement in disease activity by several measures, consistent with the well-established efficacy of ustekinumab^6^. Our results suggest that roughly a third of the ustekinumab-treated cohort of TRIDENT will be in steroid-free remission at week 24.

Interest in external control arm studies has continued to grow in recent years. This has directly followed from several trends: 1) the increasing availability of large clinical datasets such as from medical claims and EHRs, 2) advances in methods for organizing and extracting information from these data, 3) the large and rising costs of randomized trials^7^, 4) increasingly favorable attitudes by regulators towards their use^2^.

If done well, these studies have the potential to transform the way we generate clinical evidence. They can be used to inform the safety and efficacy of existing therapies, as well as new ones by indirect comparison. They may also help answer questions about comparative effectiveness, cost-benefits, and precision medicine, particularly in cohorts who might not have been studied in registrational trials.

Despite this potential, our pilot study underscores several important differences between this variety of retrospective research and their prospective counterparts. It is more difficult to create a high-quality external control arm for protocols that 1) specify exact study visit timing, 2) prioritize specific outcome measurements that are not commonly obtained in ordinary practice, and 3) constrain what treatments a patient may or may not receive (deviating from clinical care).

The principal limitation of our approach to creating an external control arm was missing data. This problem can be understood as the natural consequence of retrospective deviation from prospective study design in each of these three ways.

### Study Timing

The timing of real-world clinic visits follows clinical necessity as well as the individual preferences of providers and patients. It is not uncommon for patients to have one clinic visit that determines the need to start a new therapy, and another to evaluate treatment response. Delays between the decision to start treatment and the receipt of treatment in the real-world essentially guarantees missing data at the study visit equivalent of week 0. These delays are particularly magnified in IBD, where patients with active disease commonly need expensive biologics that often require payor authorization and scheduling of infusions.

This situation is similar at the time of follow-up. The precise timing of a follow-up visit can differ based on several factors, including provider availability, patient preferences, and even what drug a patient receives (which in turn informs the most reasonable time to expect a response).

Our study attempted to overcome these differences. We noted the high presence of missing clinic visits at week 0, and therefore used an empirical calibration approach to identify the time window for estimating a patient’s actual CDAI at the time of ustekinumab induction. We use predefined windows of ± 2 and 4 weeks for the outcome assessment for similar reasons, to avoid the retrospective miscalibration of clinic visit timings with that of TRIDENT. Future studies are needed to explore the use of other patient-interactive technologies, such as timed patient surveys embedded into EHR systems, to better address this limitation.

### Outcome measures

We found that the informal capture of clinical data can deviate substantially from that of common clinical trial instruments. The CDAI requires a significant amount of data collection across a wide variety of domains – PROs, vitals, laboratories, and extraintestinal diagnoses. It also requires week-long symptom diaries. Unsurprisingly, the CDAI has had poor uptake in actual clinical practice. This was a large driver of missing data in this study.

Our findings suggest substantial potential for simplifying these indices, or more generally, approaches to better align them to the realities of real-world practice. A large part of the reason why our algorithms were as accurate as they were for ascertaining the CDAI was because of class imbalance. That is, most patients did not have any EHR-based evidence for extraintestinal manifestations, and thus their CDAIs were strongly driven by the PROs (all of which were abstracted manually). A further simplification of the PROs from multi-level ordinal variables to even a binary variable might make for a good tradeoff between precision and suitability for routine clinical capture. Future work is needed to develop ‘real-world ready’ instruments that maintain responsiveness and validity.

### Constrained treatments

The final point, getting at the heart of the difference between clinical care and controlled studies, resulted in a different kind of missing data problem: one of diminished sample size rather than missing values. Although we began this study with 736 potential candidates, we excluded about 75% of this population after sequentially applying the major eligibility criteria used to screen subjects in TRIDENT. This study was not designed to measure what proportion of candidates were excluded by different criteria. We applied a ‘greedy’ selection approach (eliminating candidates at the earliest evidence of disqualification) to avoid the labor of otherwise capturing 7,360 data points (10 non-CDAI eligibility criteria).

However, our impression was that many patients were disqualified due to changes in therapy during the washout period prior to the date of ustekinumab induction. This of course is quite natural: patients with active Crohn’s disease who are under clinical care are highly likely to undergo changes in treatment, whether rapid transitions from prior treatment to new ones, or the addition of adjunctive/bridging agents like steroids. This was our reason for removing the biologic washout requirement as an eligibility criterion in this study.

This misalignment between experiments designed to measure treatment effects and clinical practice designed to treat patients results in a significant loss in study efficiency. One solution might involve using larger clinical datasets, to find many more of those rare symptomatic patients who ordinarily would have been treated but by chance were not. However, this approach may increase the risks of unmeasured confounding and residual bias. A potentially better solution would be that of an EHR-enabled registry. The use of phenotyping algorithms to screen patients prior to manual review might be a more cost-effective way to efficiently recruit patients, guarantee the timing and capture of relevant data, and involve more under-represented patients in studies that culminate in practice-changing evidence.

Strengths of this study include the use of predefined chart review protocol, validation of algorithms against that of manual review, sensitivity analyses to explore the effects of various design decisions on outcomes, the release of our raw data and code, and the identification of treatment effects that are broadly consistent the literature. Weaknesses as described above pertain primarily to missing data and the resulting inability to make stronger inferences about treatment effectiveness.

In conclusion, we have piloted an approach for performing an external control arm study in Crohn’s disease. Future studies are needed to improve data capture and bridge the gap between retrospective and prospective clinical research.

## Data Availability

The clinical data used in this study were deidentified to comply with US Department of Health and Human Services 'Safe Harbor' guidance and applicable laws and regulations concerning privacy and/or security of personal information. A deidentified table containing abstracted data and accompanying code have been made available for public use in the supplementary data file accompanying this manuscript.

## ACKNOWLEDGEMENTS

The authors thank Jennifer Creasman, UCSF Academic Research Services, and Clinical Data Research Consultation services for clinical informatics support.

