## Supplemental Methods for "Creation of a ustekinumab external control arm for Crohn’s disease using electronic health records data: a pilot study"

Table of Contents:

1) Study Eligibility Criteria
2) Definitions of informatically-ascertained CDAI elements
3) Calculation of the CDAI
4) Abstraction definitions
5) STROBE Checklist
6) TRIDENT Protocol, Amendment 6

Study Eligibility Criteria

*Inclusion criteria*:
1) Age ≥ 18
2) A diagnosis of Crohn's disease as determined by the treating clinician
3) Receipt of ustekinumab at the FDA-approved route and dose
4) A baseline Crohn's Disease Activity Index (CDAI) within the range of 220 to 450 inclusive

*Exclusion criteria*:
1) Receipt of IV steroids within the 3 weeks prior to starting ustekinumab
2) A change to the dose of an oral corticosteroid (e.g. prednisone, budesonide) within the 3 weeks prior to starting ustekinumab
3) A change in the dose of methotrexate, 6-mercaptopurine, or azathioprine within the 12 weeks prior to starting ustekinumab
4) A change in the dose of antibiotics as used to treat Crohn's disease within the 3 weeks prior to starting ustekinumab
5) A change in the dose of 5-ASA compounds within the 3 weeks prior to starting ustekinumab
6) The occurrence of bowel surgery (segmental colectomy, ileocecectomy) within the 6 months prior to starting ustekinumab
7) A history of major bowel surgery (resection of >50% of the colon, multiple small bowel surgeries) at any point in time prior to starting ustekinumab

Definitions of informatically-ascertained CDAI elements:

For each of the below extraintestinal manifestation variables, the diagnosis was considered present (and corresponding points were assigned) if the below codes were found within time windows corresponding to baseline, week 12, and week 24.

*Arthritis/arthralgias*:
- ICD-9-CM: 710*-719*
- ICD-10-CM: M04*-M14.*

*Iritis/uveitis:*
- ICD-9-CM: 364*
- ICD-10-CM: H20*

*Erythema nodosum, pyoderma gangrenosum, aphthous stomatitis:*
- ICD-9-CM: 686.01, 695.2, 528.2
- ICD-10-CM: L88*, L52*, K12.0

*Anal fissure, perianal fistula, perianal abscess*:
- ICD-9-CM: 565*/566*
- ICD-10-CM: K60*/K61*

*Other fistula*:
- ICD-9-CM 569.81
- ICD-10-CM K63.2

Abdominal mass was assumed absent for all patients. Other variables were obtained from structured data as follows:

*Weight, fever/temperature > $100^o$ F:* Ascertained by query of patient vitals tables
*Hematocrit*: Ascertained by query of patient labs tables
*Antidiarrheals or Opiates*: Ascertained by query of patient medications tables corresponding to the following medications: diphenoxylate-atropine (Lomotil), loperamide (Imodium), Oxycodone, Oxycontin, Percocet, Norco, Vicodin, Hydrocodone

For each of the diagnoses above, the absence of any corresponding diagnosis code was treated as positive evidence of absence (i.e. no missing data in these variables by definition). Similarly, the absence of a medication listing for an antidiarrheal or opiate was treated as positive evidence of absence.

*Ideal body weight*: Patient heights were ascertained by query of the patient vitals tables. This height and gender were used to determine the ideal body weight according to the following table provided by the study sponsor.

| Actual Height (cm) | Men | Women |
|---|---|---|
| 137.2 to 138.3 | 54.4 | 48.5 |
| 138.4 to 139.6 | 54.9 | 49 |
| 139.7 to 140.9 | 55.3 | 49.4 |
| 141.0 to 142.1 | 55.8 | 49.9 |
| 142.2 to 143.4 | 56.2 | 50.3 |
| 143.5 to 144.7 | 56.7 | 50.8 |

| | | |
|---|---|---|
| 144.8 to 146.0 | 57.1 | 51.2 |
| 146.1 to 147.2 | 57.6 | 51.7 |
| 147.3 to 148.5 | 58 | 52.2 |
| 148.6 to 149.8 | 58.5 | 52.6 |
| 149.9 to 151.0 | 59 | 53.1 |
| 151.1 to 152.3 | 59.4 | 53.6 |
| 152.4 to 153.6 | 59.9 | 54.2 |
| 153.7 to 154.8 | 60.3 | 54.8 |
| 154.9 to 156.1 | 60.8 | 55.3 |
| 156.2 to 157.4 | 61.2 | 56 |
| 157.5 to 158.7 | 61.7 | 56.7 |
| 158.8 to 159.9 | 62.1 | 57.4 |
| 160.0 to 161.2 | 62.6 | 58 |
| 161.3 to 162.5 | 63 | 58.7 |
| 162.6 to 163.7 | 63.5 | 59.4 |
| 163.8 to 165.0 | 64.1 | 60.1 |
| 165.1 to 166.3 | 64.6 | 60.8 |
| 166.4 to 167.5 | 65.2 | 61.4 |
| 167.6 to 168.8 | 65.8 | 62.1 |
| 168.9 to 170.1 | 66.4 | 62.8 |
| 170.2 to 171.4 | 67.1 | 63.5 |
| 171.5 to 172.6 | 67.8 | 64.2 |
| 172.7 to 173.9 | 68.5 | 64.9 |
| 174.0 to 175.2 | 69.2 | 65.5 |
| 175.3 to 176.4 | 69.8 | 66.2 |
| 176.5 to 177.7 | 70.5 | 66.9 |
| 177.8 to 179.0 | 71.2 | 67.6 |
| 179.1 to 180.2 | 71.9 | 68.3 |
| 180.3 to 181.5 | 72.6 | 68.9 |
| 181.6 to 182.8 | 73.4 | 69.9 |
| 182.9 to 184.1 | 74.1 | 70.3 |
| 184.2 to 185.3 | 75 | 71 |
| 185.4 to 186.6 | 75.7 | 71.7 |
| 186.7 to 187.9 | 76.6 | 72.3 |
| 188.0 to 189.1 | 77.5 | 73 |
| 189.2 to 190.4 | 78.4 | 73.7 |
| 190.5 to 191.7 | 79.1 | 74.4 |
| 191.8 to 192.9 | 80.2 | 75.1 |
| 193.0 to 194.2 | 81.2 | 75.7 |

| | | |
|---|---|---|
| 194.3 to 195.5 | 82.1 | 76.4 |
| 195.6 to 196.8 | 83 | 77.1 |
| 196.9 to 198.0 | 83.9 | 77.8 |
| 198.1 to 199.3 | 84.8 | 78.5 |
| 199.4 to 200.6 | 85.7 | 79.1 |
| 200.7 to 201.8 | 86.6 | 79.8 |
| 201.9 to 203.1 | 87.5 | Not applicable |
| 203.2 to 204.4 | 88.5 | Not applicable |
| 204.5 to 205.6 | 89.4 | Not applicable |
| 205.7 to 206.9 | 90.3 | Not applicable |
| 207.0 to 208.2 | 91.2 | Not applicable |
| 208.3 to 209.5 | 92.1 | Not applicable |
| 209.6 to 210.7 | 93 | Not applicable |
| 210.8 | 93.9 | Not applicable |

Calculation of the CDAI: Calculation generally followed the formula as originally published by Best et al (1976) with the following modifications to better match the statistical analysis plan of TRIDENT:

- Weight loss was calculated as a percentage relative to the ideal body weight. Weight loss calculated to be less than -10% were truncated to -10%
- 30 points were assigned if the patient had any evidence of antidiarrheal therapy or opiate use within a given time window

Abstraction definitions: The below protocol was used to support Phases 2-6 as described in the methods section. Prior to its use, it was distributed in advance to the study sponsor and to all annotators for comment and revision.

**Ustekinumab SCA**
**Data Dictionary and Record Review Criteria**

**General Comments**

This chart review process requires that the reviewer understand the principles of clinical medicine and gastroenterology, the medical management of Inflammatory Bowel Disease (IBD), and various practical aspects of healthcare delivery. This knowledge is needed to properly interpret data found in the electronic health records (EHR). Although this expertise is typically found among gastroenterology clinicians with exposure to IBD patient management, it may also be developed in individuals with a less technical background following an initial period of close supervision.

In the review process, annotators should make use of the complete information available within the EHR. This includes notes, encounter-level data, scanned documents, endoscopy reports, global search, and patient-authorized clinical data shared from other sites where healthcare has been provisioned (i.e. Health Information Exchange data via *Care Everywhere* or equivalent).

Annotators are encouraged to take a '360 degree' approach that relies on several independent sources of information rather than relying on a single source of truth. They are encouraged to anticipate the possibility of mis-annotation from 'copy forwarding', and to recognize sections of the clinical note more likely to be meaningfully annotated (e.g. the assessment/plan or impression section, or a de novo, free-texted nursing note) than others. They are encouraged to be conservative in the face of uncertainty. This may involve documenting certain fields as missing rather than available.

Annotators are recommended to use heuristics that incorporate existing knowledge about clinical medicine, common practice of medication usage in real-world practice settings, and healthcare delivery. These heuristics include understanding common frequencies of dosing, locations of where medication are delivered and whom might document them, as well as common and uncommonly prescribed combinations of IBD treatments among others. The use of this process makes the record review process somewhat more 'organic' and less 'algorithmic' or 'deterministic', but in our view substantially increases the relevant information content available within the EHR without sacrificing robustness or reproducibility.

This document does not aspire to be exhaustive and cover every edge case; we view this task as being essentially impossible and/or one that may bias the annotation process in unanticipated and undesirable ways. Rather, we anticipate that a combination of this guidance with individuals of sufficient training will lead to the consistent identification and annotation of medical records corresponding to Ustekinumab-treated Crohn's Disease patients.

**Part 1 Annotations**:

Variables should be annotated in order; if a patient already merits exclusion from the target study based on an earlier variable, subsequent variables may be annotated NA at the discretion of the reviewer.

**MRN** (Integer):
Medical Record Number of the patient obtained by structured database query

**Annotated by** (character):
Initials of the chart annotator

**Confirmed dx** (Y, N, NA):
Annotate as Y if the patient was confirmed to have a diagnosis of Crohn's disease at the time of Ustekinumab initiation, N if there is positive evidence for a different diagnosis (including IBD-unclassified), and NA if insufficient data.

The sources of this diagnosis should preferentially come from the gastroenterologist who either recommended or prescribed the drug, typically documented in a note from the clinic encounter associated with the prescription. Provisional diagnoses can also be taken from endoscopy reports done near the time of treatment initiation. Of note, this field corresponds to the diagnosis assigned by the clinician at the time of drug initiation and should not incorporate any subsequent diagnostic revision.

**Received UST** (Y, N, NA):
Annotate as Y if the patient was confirmed to have received Ustekinumab, N if not, and NA if there is insufficient information to confirm or deny receipt. In general the absence of information to support Ustekinumab initiation should be annotated as an N (i.e. use NA sparingly).

Confirmation of Ustekinumab receipt should typically be accompanied by the date of initiation (see next field). The gold-standard for receipt is an infusion center note (typically authored by a nurse) that provides a date and the details of this infusion encounter. It is sometimes accompanied by a timestamp in the form of a 'medication administration record' entry that is linked to these notes within the electronic health record.

An acceptable alternative is multiple sources of clinical documentation, typically by GI providers, that indicate a month and year of initiation.

**Date of UST** (YYYY-MM-DD, NA):
Provide the date that Ustekinumab was initiated. If a month and year are available but not the day, the day may be imputed to the $15^{th}$ day of that month (so that it is correct on average). If the month is unavailable (e.g. only the year), annotate this as NA.

As above, preferentially use infusion encounter or pharmacy notes to determine this. When unavailable can use documentation from the primary GI's office, especially from clinic administration staff (who facilitate payor approval and schedule infusion appointments). Notes from non-GI clinicians (e.g. primary care, rheumatology, dermatology) can increase confidence in making this annotation. Rarely the reviewer may have to use the knowledge that this drug is typically dosed every 8 weeks in order to back calculate the precise day of infusion within a given month or week.

**Exact Date** (Y, N, NA): Annotate this as Y if the prior field corresponds to a precise date, or N if the day was imputed to '15' due to inadequate documentation within the EHR. NA if the patient has been excluded by prior variables (i.e. patient does not have Crohn's).

**Age >= 18** (Y, N, NA): Annotate a Y if the patient was aged 18 or older *at the time they received Ustekinumab*. Annotate as N otherwise, or NA if unable to determine (rare) or patient excluded by prior variables.

**NoRecentOrMajorSurgery** (Y, N, NA):
Annotate N if the patient had any small or large bowel resection within the 6 months prior to Ustekinumab infusion, or alternatively if they have had a history of >50% resection of the colon, or a clinical diagnosis of 'short bowel syndrome.' Annotate Y otherwise. If the patient record is deemed to have inadequate documentation related to these surgical events, annotate as NA.

**History of Biologic Intolerance/Refractoriness** (Y, N, NA):
Annotate as a Y if the patient has previously experienced
1) one of the following biologics (Infliximab, Adalimumab, Certolizumab, or Vedolizumab; Natalizumab, Golimumab, and other biologics are not listed in the TRIDENT trial protocol Attachment 1), and
2) either was not thought to have responded or lost response from the standpoint of Crohn's disease

(does not count if the patient received it for a different indication and Crohn's disease activity were not explicitly evaluated or requiring of medical treatment).

Annotate as Y if the patient received one of the above biologics and experienced any intolerance to the medication that was thought to be related to the medication by any documenting clinician.

Document as N otherwise, or document as NA if insufficient data is available to make this determination.

**No TNFi<8wks** (Y, N, NA):
Annotate as N if the patient received a dose of a tumor necrosis factor inhibitor (TNFi) – for example Infliximab, Adalimumab, Certolizumab, or Golimumab – within 56 days of the date of Ustekinumab induction. Annotate Y if the patient has no history of prior TNFi use or if the last date of use was over 56 days prior to Ustekinumab induction.

Annotate as NA if the information within the EHR is inadequate to annotate as Y or N. At the reviewers discretion such patients may alternatively be imputed to N (so as to exclude them from any further consideration).

**No IV Steroids<3wks** (Y, N, NA):
Annotate as N if the patient received a dose of an IV corticosteroid, typically methylprednisolone or (less commonly) hydrocortisone within 21 days of Ustekinumab induction.

The annotator is reminded that these medications are almost exclusively administered in an inpatient/hospitalized setting. If the patient had a significant number of available primary care or gastroenterology clinician encounters within the 3 week window preceding or following Ustekinumab induction, this variable may be annotated as a Y. The logic underlying this is that the patient would be expected to have had documentation of such an event by a treating physician, and as such the absence of this documentation is treated as positive evidence of no hospitalization.

If the numbers of clinical encounters during this period are insufficient to confidently ascertain that the patient was not hospitalized, annotate this field as NA.

**No Vedo<16wks** (Y, N, NA):
Annotate as N if the patient received Vedolizumab within 112 days of the date of Ustekinumab initiation.

The annotator is reminded at as of the date of annotation for the present project (6/1/2020), Vedolizumab is only FDA approved as an infused drug and is not commercially available in a subcutaneous formulation – thus it is typically delivered in an infusion center and commonly accompanied by an note from an infusion nurse. He or she is also reminded that the drug is typically administered in 4 or 8 week intervals, and that the dosing frequency can typically be ascertained from clinical notes and/or documentation related to payor approval of dosing frequency.

Heuristics as suggested in the preamble of this document may be used to calculate the last date of Vedolizumab administration in order to annotate this variable.

If the patient has no history of Vedolizumab use, or received it outside of this window, annotate as Y. Annotate as NA if the EHR provides insufficient information to otherwise annotate this variable.

**Stable 5ASA 3wks** (Y, N, NA):
Annotate as Y if the patient has either not been on an oral 5-Aminosalicylate or derivative (e.g. mesalamine, olsalazine, balsalazide, sulfasalazine) in the 21 days prior to Ustekinumab, or has been on them at a stable dose for this period. Annotate as N if the patient has evidence of a fluctuating dose of this medication during the lead-in period. Annotate as NA if there is insufficient evidence of annotate otherwise.

Annotators are advised that this use of this medication is often poorly documented in clinical notes and may be captured properly during the medication reconciliation process. However, gastroenterologists (and occasionally rheumatologists) are the only clinicians who prescribe this medication and therefore the absence of specific mention in the clinical note or new medication order can be taken as evidence that the dose was stable (whether at a dose of 0mg or greater).

**Stable Abx 3wks** (Y, N, NA):
Annotate as Y if the patient has been on a stable dose of antibiotics <u>as used for the primary treatment of Crohn's Disease itself</u> in the 21 days prior to Ustekinumab induction. If the patient has not been on antibiotics, annotate as Y. If the patient has been on antibiotics to treat an infection (infectious diarrhea, abscess, sepsis) this does not disqualify the patient. If the patient has been on antibiotics to treat a draining fistula, this would could as treatment of Crohn's disease. Primary does not imply that it is the only treatment, but rather the purpose of that treatment. Annotate as N if the patient has received antibiotics to treat Crohn's within the 21 days prior to Ustekinumab, or NA if insufficient data to determine this.

**Stable PO Steroids 3wks** (Y, N, NA):
Annotate as Y if the patient has either not been on an oral steroid (e.g. prednisone) in the 21 days prior to Ustekinumab, or has been on them at a stable dose under 40mg prednisone or 9mg budesonide (or equivalent) for this period.

If patients are on a higher dose than 40mg predisone equivalent, they should be annotated as a N. If they have been on either an increasing or decreasing dose during this period, they should be annotated as an N.

Annotators are advised that this medication is unevenly documented in the clinical record, and are advised to rely more strongly on active documentation of  use or non-use as well as supporting documentation from prior GI visits as well as from other non-GI providers and/or providers outside of the host institution to obtain a more accurate picture.

In the absence of sufficient information, annotate this as NA.

**Stable MTX/6MP/AZA 12wks** (Y, N, NA):
Annotate as Y if the patient has either not been on an oral immunomodulator in the 84 days prior to Ustekinumab use, or N if so. If a patient has not been on immunomodulators previously and the GI who prescribes Ustekinumab does not indicate his or her intention to initiate this as co-treatment, then annotate this as N. If the patient has previously been on an immunomodulator and the notes are unclear as to whether or not the medication is being continued, annotate this as NA.

**NoOtherExclusion** (Y, N):
Annotate as Y if the patient should be excluded from the study for a reason not otherwise covered in the major criteria. Otherwise annotate as N. Blanks values and NAs will be assumed to represent an N.

**Likely Exclude** (Y, N, NA):
The purpose of this 'flag' is to potentially prioritize future, subsequent annotations in the event that this first pass results in too many cases.

Annotate this as Y if the patient has already been 'excluded' based on a major criterion from the variables covered thus far, or if he or she is likely to be excluded based on a variable not yet captured (but likely to be captured on the next round of review). Annotate as N if the chart reviewer has not identified any clear red flags suggesting exclusion. Annotate as NA for all other reasons.

**Borderline** (Y, N, NA):
The purpose of this flag is to prioritize future subsequent annotations in the event that this first pass results in too few cases.

Annotate as Y if the patient is currently listed as excluded but may be excluded for a relatively minor reason unlikely to substantially affect the outcome measurement (e.g. disease improvement on Ustekinumab at 12 or 24 weeks). These may be candidates for inclusion in a sensitivity analysis. Examples include fluctuating 5ASA dose (comparatively weak), exclusion based on recent drug exposure that was thought to be weakly effective or ineffective, etc. Annotate as N if the patient is clearly included or excluded, or NA otherwise.

**Part 1.1 annotations**:

**MRN** (Integer): Medical Record Number. This variable will be captured by an informatic pull

**Annotator** (character): Initials of the annotator

**Birthdate** (MM/DD/YYYY): Indicate the birthdate of the patient here

**Gender** (M, F, O, NA): Indicate the gender of the patient here, M for male, F for female, O for other, NA for missing data

**BioIR** (Y, N, NA):
Annotate as a Y if the patient has previously experienced
1) one of the following biologics (Infliximab, Adalimumab, Certolizumab, or Vedolizumab; Natalizumab, Golimumab, and other biologics are not listed in the TRIDENT trial protocol Attachment 1), and
2) either was not thought to have responded or lost response from the standpoint of Crohn's disease (does not count if the patient received it for a different indication and Crohn's disease activity were not explicitly evaluated or requiring of medical treatment).

Annotate as Y if the patient received one of the above biologics and experienced any intolerance to the medication that was thought to be related to the medication by any documenting clinician.

Document as N otherwise, or document as NA if insufficient data is available to make this determination.

**First Dose IV** (Y, N, NA):
Annotate Y if the patient received the first dose as an IV, N if the patient did not (i.e. subcutaneous), or NA if unable to determine.

<u>If the patient is N or NA, no further annotations are necessary; the patient is to be excluded from the study</u>.

**NoRecentOrMajorSurgery** (Y, N, NA):
Annotate N if the patient had any small or large bowel resection within the 6 months prior to Ustekinumab infusion, or alternatively if they have had a history of >50% resection of the colon, or a clinical diagnosis of 'short bowel syndrome.' Annotate Y otherwise. If the patient record is deemed to have inadequate documentation related to these surgical events, annotate as NA.

**Date of UST** (MM/DD/YYYY): Date the patient received the first dose of Ustekinumab. This can essentially be copied from your phase 1 annotations; but is only here to assist the annotator with coding other variables corresponding to an 8 week period lookback from this date.

**Bsln at UCSF** (Y, N, NA):
Annotate as Y if the patient was prescribed his or her first dose by a UCSF physician, N if not, or NA if unclear. Note that a prescription by a UCSF physician does not imply that the patient actually received the dose at a UCSF facility.

These patients typically are evaluated in an office visit or at endoscopy at the time of this prescription; as such this variable serves as a proxy for patients who are likely to have baseline and follow-up data available.

<u>This criteria is not to be considered exclusionary at this time, so do continue to annotate other fields even if this is an N or NA</u>. Rather, it serves as more of an indicator variable that will tell us how good the data capture is or isn't for these patients. Based on this we will consider whether or not we will make this a hard eligibility criteria when we consider revising the first pass annotations to determine patient selection (e.g. include patients who did not meet the biologic washout period but did have good baseline data capture at UCSF).

**Bsln Daily Abd Pain** (Numeric, NA):
Within the -16 to 0 week period prior to the first dose of Ustekinumab, indicate patient's level of daily abdominal pain according to the following scale:
0 for none, 1 for mild, 2 for moderate, 3 for severe.
If the notes indicate a range, report the mean (e.g. mild-moderate pain as a 1.5).
If otherwise not well documented, annotate as NA.

For situations in which there are multiple sources of truth within the 8 week period relevant to this data, prioritize reporting the one that is felt to be most reliable, followed by the data element closest to week 0.

**Date Bsln Daily Abd Pain** (MM/DD/YYYY):

Indicate the date of the note that the daily abdominal pain score was abstracted from. In the event that multiple dates contributed to a score, select the date closest to the date of infusion.

**Bsln Diarrhea** (Numeric, NA):
Within the -16 to 0 week period prior to the first dose of Ustekinumab, indicate the <u>total number of bowel movements per day</u> as reported at any point during the baseline period (-8 weeks to 0 weeks of the first Ustekinumab dose). If the notes report a range, indicate the average of the range. For instance, if the notes indicate 3-4 bowel movements daily, report 3.5 here.

<u>Of note, this definition slightly defers from the trial protocol definition, which indicates the number of liquid or very soft stools daily</u>. The rationale for this difference is that clinical notes tend to better capture total bowel movement frequency than they do characterize the quality of the bowel movement. Many patients with active Crohn's disease tend to have loose bowel movements. Lastly this distinction is unlikely to impact downstream analyses, especially those that look for within-subject differences over time.

For situations in which there are multiple sources of truth within the 8 week period relevant to this data, prioritize reporting the one that is felt to be most reliable, followed by the data element closest to week 0.

If the notes do not explicitly indicate number of bowel movements during this period, but if notes from before and after this period indicate a stable frequency of bowel movements, then can use heuristics to report this during the baseline period.

If the patient has an ostomy, then annotate this as NA and flag this chart (these may ultimately be excluded). If the patient has an ileal pouch-anal anastomosis (also known as a 'J-Pouch', a surgery following complete removal of the rectum and colon), ok to report the number of bowel movements but also flag this chart. If otherwise not well documented, annotate as NA.

**Date Bsln Diarrhea** (MM/DD/YYYY):
Indicate the date of the note that the stool frequency was abstracted from. In the event that multiple dates contributed to a score, select the date closest to the date of infusion.

**Bsln WellBeing** (Numeric, NA):
Within the -16 to 0 week period prior to the first dose of Ustekinumab, score the patient's apparent well-being on the following scale:
0 = generally well, 1 = slightly under par, 2 = poor, 3 = very poor, 4 = terrible

This variable is generally more difficult to annotate and may require some degree of annotator 'reading between the lines' in order to perform mapping to this scale. This variable is associated with the existing variables (abdominal pain, diarrhea) but also encompasses other inputs such as fatigue, subjective fevers, anxiety, loss of appetite, and other symptoms.

Patients who are symptom-free should be annotated as a 0. The presence of mild symptoms (occasional abdominal cramping, some loose stools) likely reflects a 1. Patients with significant symptoms, chronic use of opiates, steroids, seeing multiple specialists (mental health, nutrition/dieticians, rheumatology/dermatology) may be annotated as a 2. Patients who are hospitalized should be considered at least a 3, with patients who are suffering concomitant infection/sepsis and/or weight loss considered for a 4. For patients who appear to fit a range (e.g. between a 2 and 3), report the average (e.g. 2.5).

For situations in which there are multiple sources of truth within the 8 week period relevant to this data, prioritize reporting the one that is felt to be most reliable, followed by the data element closest to week 0.

**Date Bsln Wellbeing** (MM/DD/YYYY):
Indicate the date of the note that the wellbeing was abstracted from. In the event that multiple dates contributed to a score, select the date closest to the date of infusion.

**Bsln CRP** (Numeric, NA):
Within the -16 to 0 week period prior to the first dose of Ustekinumab, report the serum c-reactive protein level (in mg/L). Report the value closest to week 0. <u>Note the units</u>: this lab is sometimes reported as mg/dl, and under these scenarios has two decimal places rather than one.

Note the longer period of time for this baseline value; the rationale for this is that 1) this is a high-importance data element, 2) many patients end up having a prolonged period of prior treatment failure before receiving Ustekinumab so it might be more legitimate to carry forward this value, and 3) we can use the date field to refine this field by proximity to week 0 after the fact.

**Date Bsln CRP** (MM/DD/YYYY):
Indicate the date of the test (received, not ordered or resulted) corresponding to the crp reported in the prior field.

**Bsln Calprotectin** (Numeric, NA):
Within the -16 to 0 week period prior to the first dose of Ustekinumab, report the fecal calprotectin (in mcg/g). Report the value closest to week 0.

As with crp, we are using a longer window of time here in order to maximize the capture of this less commonly ordered (but highly informative) variable. If the variable proves to be too sparse we may decide to exclude it.

**Date Bsln Calprotectin** (MM/DD/YYYY):
Indicate the date of the test (received, not ordered or resulted) corresponding to the fecal calprotectin reported in the prior field.

**Bsln Steroid use** (Y, N, NA):
Indicate Y if the patient was taking steroids at the time of Ustekinumab induction, N if the patient was not, or NA if insufficient data to determine this. Use the Phase 1.1 document as appropriate to annotate this.

**Disease Location** (I, IC, C, O, NA):
Indicate if the patient has ileal disease (I), ileocolonic (IC), colonic (C), or other (O, e.g. isolated perianal). This designation is inherently ambiguous in part due to the lack of universal agreement regarding the interpretation of ileocecal valve involvement, lack of clarity regarding need for (and interpretation of) histological specimens as compared to gross endoscopic involvement, etc. The study sponsors have indicated that they intend to combine ileal and ileocolonic groups into a single category (to be

distinguished from colonic).

As such, it is acceptable to simply use the designation indicated in the clinical notes at the time of Ustekinumab treatment without a need to refer to the original documentation (typical referral notes from other providers, with or without supporting endoscopy/pathology data). This is most likely to be consistent with how study clinicians are annotating this in the setting of prospective trials.

**Diagnosis Year** (YYYY, NA):
Indicate the year that the diagnosis was confirmed by a treating clinician. This is typically done by colonoscopy with biopsies or less commonly surgical explant histopathology. Many patients are diagnosed elsewhere and referred in so search for the earliest GI note and consider looking in the scanned clinical documents tab to identify outside docs sent in around the time of the referral to UCSF.

**NoOtherExclusion** (Y, N):
Annotate as N if the patient should be excluded from the study for a reason not otherwise covered in the major criteria. Otherwise annotate as Y. Blanks values and NAs will be assumed to represent an N.

**Likely Exclude** (Y, N):
Annotate as a Y if the patient fails eligibility criteria based on the reassessment done here (including recent or major bowel surgeries, or first dose not given IV).

**Part 2 Annotations**:

**W12AbdPain** (Numeric, NA):
Within the week 10-14 period with respect to the patient's dose of IV Ustekinumab, indicate patient's level of daily abdominal pain according to the following scale:
0 for none, 1 for mild, 2 for moderate, 3 for severe.
If the notes indicate a range, report the mean (e.g. mild-moderate pain as a 1.5).
If otherwise not well documented, annotate as NA.

For situations in which there are multiple sources of truth within this window, select the one closest to week 12.

**Date W12AbdPain** (MM/DD/YYYY):
Indicate the date of the note that the daily abdominal pain score was abstracted from. In the event that multiple dates contributed to a score, select the date closest to the date of infusion.

**W12Diarrhea** (Numeric, NA):
Within the week 10-14 period with respect to the patient's dose of IV Ustekinumab, indicate the total number of bowel movements per day as reported at any point. If the notes report a range, indicate the average of the range. For instance, if the notes indicate 3-4 bowel movements daily, report 3.5 here.

Of note, this definition slightly defers from the trial protocol definition, which indicates the number of liquid or very soft stools daily. The rationale for this difference is that clinical notes tend to better capture total bowel movement frequency than they do characterize the quality of the bowel movement. Many patients with active Crohn's disease tend to have loose bowel movements. Lastly this distinction is unlikely to impact downstream analyses, especially those that look for within-subject differences over time.

For situations in which there are multiple sources of truth within this window, select the one closest to week 12.

If the notes do not explicitly indicate number of bowel movements during this period, but if notes from before and after this period indicate a stable frequency of bowel movements, then can use heuristics to report this during the baseline period.

If the patient has an ostomy, then annotate this as NA and flag this chart (these may ultimately be excluded). If the patient has an ileal pouch-anal anastomosis (also known as a ‘J-Pouch’, a surgery following complete removal of the rectum and colon), ok to report the number of bowel movements but also flag this chart. If otherwise not well documented, annotate as NA.

**Date W12Diarrhea** (MM/DD/YYYY):
Indicate the date of the note that the stool frequency was abstracted from. In the event that multiple dates contributed to a score, select the date closest to the date of infusion.

**W12WellBeing** (Numeric, NA):
Within the week 10-14 period with respect to the patient’s dose of IV Ustekinumab, score the patient’s apparent well-being on the following scale:
0 = generally well, 1 = slightly under par, 2 = poor, 3 = very poor, 4 = terrible

This variable is generally more difficult to annotate and may require some degree of annotator ‘reading between the lines’ in order to perform mapping to this scale. This variable is associated with the existing variables (abdominal pain, diarrhea) but also encompasses other inputs such as fatigue, subjective fevers, anxiety, loss of appetite, and other symptoms.

Patients who are symptom-free should be annotated as a 0. The presence of mild symptoms (occasional abdominal cramping, some loose stools) likely reflects a 1. Patients with significant symptoms, chronic use of opiates, steroids, seeing multiple specialists (mental health, nutrition/dieticians, rheumatology/dermatology) may be annotated as a 2. Patients who are hospitalized should be considered at least a 3, with patients who are suffering concomitant infection/sepsis and/or weight loss considered for a 4. For patients who appear to fit a range (e.g. between a 2 and 3), report the average (e.g. 2.5).

For situations in which there are multiple sources of truth within this window, select the one closest to week 12.

**Date W12Wellbeing** (MM/DD/YYYY):
Indicate the date of the note that the wellbeing was abstracted from.

**W12Antidiarrheal** (Y, N, NA): Annotate as Y if the patient has been reported to use either loperamide (i.e. Imodium) or diphenoxylate-atropine (i.e. Lomotil) within the week 10-14 period with respect to the patient’s dose of IV Ustekinumab. If so, annotate as Y. If no, annotate as N. If no explicit comment on the use or absence of antidiarrheals, annotate as NAs. You may find text-searching helpful; consider use of the intentional misspelling ‘immodium’ when searching.

In the event of multiple reports indicating antidiarrheal use, select the one closest to week 12.

Of note, we are anticipating a substantial amount of missing data for this field.

**W12AntidiarrhealDate** (MM/DD/YYYY, NA): Date corresponding to the above.

**W12DailyOpiate** (Y, N, NA): Annotate as Y if the patient had ever used any opiate (oral, subcutaneous, or IV) on a daily basis within the week 10-14 period with respect to the patient's dose of IV Ustekinumab. These include, but are not limited to: Hydrocodone, Oxycodone, Hydromorphone, Morphine and Codeine. The first two on this list are most common in the outpatient IBD population, including in their formulations that include Acetaminophen (most common brand names Vicodin, Norco).

In the event of multiple reports indicating on opiate use, select the one closest to week 12.

**W12DailyOpiateDate** (MM/DD/YYYY, NA): Date corresponding to the above.

**W12Hct** (Numeric, NA): Within the week 10-14 period with respect to the patient's dose of IV Ustekinumab, indicate that value here. Use the date that the test was performed, not ordered or resulted. Consider examining scanned clinical documents and Care Everywhere to identify this data, as well as the clinic note where outside lab data is often summarized.

In the event of multiple reports indicating antidiarrheal use, select the one closest to week 12.

If this data is unavailable during this timeframe, indicate NA.

**W12HctDate** (MM/DD/YYYY, NA): Date corresponding to the above.

**W12SteroidUse** (Y, N, NA):
Indicate Y if the patient was taking steroids in the week 10-14 period, N if the patient was not, or NA if insufficient data to determine this.

**W24AbdPain** (Numeric, NA):
Within the week 20-28 period with respect to the patient's dose of IV Ustekinumab, indicate patient's level of daily abdominal pain according to the following scale:
0 for none, 1 for mild, 2 for moderate, 3 for severe.
If the notes indicate a range, report the mean (e.g. mild-moderate pain as a 1.5).
If otherwise not well documented, annotate as NA.

For situations in which there are multiple sources of truth within this window, select the one closest to week 24.

**Date W24AbdPain** (MM/DD/YYYY):
Indicate the date of the note that the daily abdominal pain score was abstracted from. In the event that multiple dates contributed to a score, select the date closest to the date of infusion.

**W24Diarrhea** (Numeric, NA):

Within the week 20-28 period with respect to the patient's dose of IV Ustekinumab, indicate the total number of bowel movements per day as reported at any point. If the notes report a range, indicate the average of the range. For instance, if the notes indicate 3-4 bowel movements daily, report 3.5 here.

Of note, this definition slightly defers from the trial protocol definition, which indicates the number of liquid or very soft stools daily. The rationale for this difference is that clinical notes tend to better capture total bowel movement frequency than they do characterize the quality of the bowel movement. Many patients with active Crohn's disease tend to have loose bowel movements. Lastly this distinction is unlikely to impact downstream analyses, especially those that look for within-subject differences over time.

For situations in which there are multiple sources of truth within this window, select the one closest to week 24.

If the notes do not explicitly indicate number of bowel movements during this period, but if notes from before and after this period indicate a stable frequency of bowel movements, then can use heuristics to report this during the baseline period.

If the patient has an ostomy, then annotate this as NA and flag this chart (these may ultimately be excluded). If the patient has an ileal pouch-anal anastomosis (also known as a 'J-Pouch', a surgery following complete removal of the rectum and colon), ok to report the number of bowel movements but also flag this chart. If otherwise not well documented, annotate as NA.

**Date W24Diarrhea** (MM/DD/YYYY):
Indicate the date of the note that the stool frequency was abstracted from. In the event that multiple dates contributed to a score, select the date closest to the date of infusion.

**W24WellBeing** (Numeric, NA):
Within the week 20-28 period with respect to the patient's dose of IV Ustekinumab, score the patient's apparent well-being on the following scale:
0 = generally well, 1 = slightly under par, 2 = poor, 3 = very poor, 4 = terrible

This variable is generally more difficult to annotate and may require some degree of annotator 'reading between the lines' in order to perform mapping to this scale. This variable is associated with the existing variables (abdominal pain, diarrhea) but also encompasses other inputs such as fatigue, subjective fevers, anxiety, loss of appetite, and other symptoms.

Patients who are symptom-free should be annotated as a 0. The presence of mild symptoms (occasional abdominal cramping, some loose stools) likely reflects a 1. Patients with significant symptoms, chronic use of opiates, steroids, seeing multiple specialists (mental health, nutrition/dieticians, rheumatology/dermatology) may be annotated as a 2. Patients who are hospitalized should be considered at least a 3, with patients who are suffering concomitant infection/sepsis and/or weight loss considered for a 4. For patients who appear to fit a range (e.g. between a 2 and 3), report the average (e.g. 2.5).

For situations in which there are multiple sources of truth within this window, select the one closest to week 24.

**Date W24Wellbeing** (MM/DD/YYYY):
Indicate the date of the note that the wellbeing was abstracted from.

**W24Antidiarrheal** (Y, N, NA): Annotate as Y if the patient has been reported to use either loperamide (i.e. Imodium) or diphenoxylate-atropine (i.e. Lomotil) within the week 20-28 period with respect to the patient's dose of IV Ustekinumab. If so, annotate as Y. If no, annotate as N. If no explicit comment on the use or absence of antidiarrheals, annotate as NAs. You may find text-searching helpful; consider use of the intentional misspelling 'immodium' when searching.

In the event of multiple reports indicating antidiarrheal use, select the one closest to week 24.

Of note, we are anticipating a substantial amount of missing data for this field.

**W24AntidiarrhealDate** (MM/DD/YYYY, NA): Date corresponding to the above.

**W24DailyOpiate** (Y, N, NA): Annotate as Y if the patient had ever used any opiate (oral, subcutaneous, or IV) on a daily basis within the week 20-28 period with respect to the patient's dose of IV Ustekinumab. These include, but are not limited to: Hydrocodone, Oxycodone, Hydromorphone, Morphine and Codeine. The first two on this list are most common in the outpatient IBD population, including in their formulations that include Acetaminophen (most common brand names Vicodin, Norco).

In the event of multiple reports indicating on opiate use, select the one closest to week 24.

**W24DailyOpiateDate** (MM/DD/YYYY, NA): Date corresponding to the above.

**W24Hct** (Numeric, NA): Within the week 20-28 period with respect to the patient's dose of IV Ustekinumab, indicate that value here. Use the date that the test was performed, not ordered or resulted. Consider examining scanned clinical documents and Care Everywhere to identify this data, as well as the clinic note where outside lab data is often summarized.

In the event of multiple reports indicating antidiarrheal use, select the one closest to week 24.

If this data is unavailable during this timeframe, indicate NA.

**W24HctDate** (MM/DD/YYYY, NA): Date corresponding to the above.

**W24SteroidUse** (Y, N, NA):
Indicate Y if the patient was taking steroids in the week 20-28 period, N if the patient was not, or NA if insufficient data to determine this.

STROBE Statement—Checklist of items that should be included in reports of ***cohort studies***

| | Item No | Recommendation |
|---|---|---|
| **Title and abstract** | 1 | (*a*) Indicate the study's design with a commonly used term in the title or the abstract<br>**Title** |
| | | (*b*) Provide in the abstract an informative and balanced summary of what was done and what was found<br>**Abstract** |
| **Introduction** | | |
| Background/rationale | 2 | Explain the scientific background and rationale for the investigation being reported<br>**Introduction, Paragraphs 1-3** |
| Objectives | 3 | State specific objectives, including any prespecified hypotheses<br>**Introduction, Paragraph 4** |
| **Methods** | | |
| Study design | 4 | Present key elements of study design early in the paper<br>**Introduction Paragraph 4, Methods section Eligibility Criteria** |
| Setting | 5 | Describe the setting, locations, and relevant dates, including periods of recruitment, exposure, follow-up, and data collection<br>**Methods sections Eligibility Criteria and Cohort Identification** |
| Participants | 6 | (*a*) Give the eligibility criteria, and the sources and methods of selection of participants. Describe methods of follow-up<br>**Methods section Eligibility Criteria** |
| | | (*b*) For matched studies, give matching criteria and number of exposed and unexposed<br>**NA** |
| Variables | 7 | Clearly define all outcomes, exposures, predictors, potential confounders, and effect modifiers. Give diagnostic criteria, if applicable<br>**Methods section Eligibility Criteria** |
| Data sources/ measurement | 8* | For each variable of interest, give sources of data and details of methods of assessment (measurement). Describe comparability of assessment methods if there is more than one group<br>**Supplemental Methods, section Definitions of informatically-ascertained CDAI elements** |
| Bias | 9 | Describe any efforts to address potential sources of bias<br>**Methods section Cohort Identification (Phases 3 through 6)** |
| Study size | 10 | Explain how the study size was arrived at<br>**Methods sections Cohort Identification, Number of Subjects (planned and analyzed)** |
| Quantitative variables | 11 | Explain how quantitative variables were handled in the analyses. If applicable, describe which groupings were chosen and why<br>**Supplemental Methods, section Definitions of informatically-ascertained CDAI elements** |
| Statistical methods | 12 | (*a*) Describe all statistical methods, including those used to control for confounding<br>**Methods sections Cohort Identification, Analytical Methods** |
| | | (*b*) Describe any methods used to examine subgroups and interactions<br>**NA** |
| | | (*c*) Explain how missing data were addressed<br>**Methods section Cohort Identification (Phase 5)** |
| | | (*d*) If applicable, explain how loss to follow-up was addressed |

| | | |
|---|---|---|
| | | **Methods section Cohort Identification (Phase 6)** |
| | | (*e*) Describe any sensitivity analyses<br>**Methods section Cohort Identification** |
| **Results** | | |
| Participants | 13* | (a) Report numbers of individuals at each stage of study—eg numbers potentially eligible, examined for eligibility, confirmed eligible, included in the study, completing follow-up, and analysed<br>**Figure 1, Results** |
| | | (b) Give reasons for non-participation at each stage<br>**Figure 1, Results** |
| | | (c) Consider use of a flow diagram<br>**Figure 1** |
| Descriptive data | 14* | (a) Give characteristics of study participants (eg demographic, clinical, social) and information on exposures and potential confounders<br>**Table 4** |
| | | (b) Indicate number of participants with missing data for each variable of interest<br>**Table 2, Table 5** |
| | | (c) Summarise follow-up time (eg, average and total amount)<br>**Tables 1, 5, 6** |
| Outcome data | 15* | Report numbers of outcome events or summary measures over time<br>**Table 6** |
| Main results | 16 | (*a*) Give unadjusted estimates and, if applicable, confounder-adjusted estimates and their precision (eg, 95% confidence interval). Make clear which confounders were adjusted for and why they were included<br>**Table 6** |
| | | (*b*) Report category boundaries when continuous variables were categorized<br>**Table 6** |
| | | (*c*) If relevant, consider translating estimates of relative risk into absolute risk for a meaningful time period<br>**NA** |
| Other analyses | 17 | Report other analyses done—eg analyses of subgroups and interactions, and sensitivity analyses<br>**Figure 2** |
| **Discussion** | | |
| Key results | 18 | Summarise key results with reference to study objectives<br>**Discussion, Paragraph 1** |
| Limitations | 19 | Discuss limitations of the study, taking into account sources of potential bias or imprecision. Discuss both direction and magnitude of any potential bias<br>**Discussion** |
| Interpretation | 20 | Give a cautious overall interpretation of results considering objectives, limitations, multiplicity of analyses, results from similar studies, and other relevant evidence<br>**Discussion** |
| Generalisability | 21 | Discuss the generalisability (external validity) of the study results<br>**Discussion** |
| **Other information** | | |
| Funding | 22 | Give the source of funding and the role of the funders for the present study and, if applicable, for the original study on which the present article is based<br>**Title Page, section Financial Support** |

**Janssen Research & Development***

**Clinical Protocol**

### A Phase 2b, Randomized, Double-blind, Placebo-controlled, Parallel-group, Multicenter Protocol to Evaluate the Safety and Efficacy of JNJ-64304500 in Subjects with Moderately to Severely Active Crohn's Disease

**TRIDENT**

**Protocol 64304500CRD2001; Phase 2b**
**AMENDMENT 6**

**JNJ-64304500**

*Janssen Research & Development is a global organization that operates through different legal entities in various countries. Therefore, the legal entity acting as the sponsor for Janssen Research & Development studies may vary, such as, but not limited to Janssen Biotech, Inc.; Janssen Products, LP; Janssen Biologics, BV; Janssen-Cilag International NV; Janssen, Inc; Janssen Infectious Diseases BVBA; Janssen Sciences Ireland UC; or Janssen Research & Development, LLC. The term "sponsor" is used throughout the protocol to represent these various legal entities; the sponsor is identified on the Contact Information page that accompanies the protocol.

This study will be conducted under US Food & Drug Administration IND regulations (21 CFR Part 312).

**EudraCT NUMBER:** 2016-000634-21

**Status:** Approved
**Date:** 4 February 2020
**Prepared by:** Janssen Research & Development, LLC
**EDMS number:** EDMS-ERI-117774163:8.0

**GCP Compliance:** This study will be conducted in compliance with Good Clinical Practice, and applicable regulatory requirements.

**Confidentiality Statement**
The information in this document contains trade secrets and commercial information that are privileged or confidential and may not be disclosed unless such disclosure is required by applicable law or regulations. In any event, persons to whom the information is disclosed must be informed that the information is privileged or confidential and may not be further disclosed by them. These restrictions on disclosure will apply equally to all future information supplied to you that is indicated as privileged or confidential.

5.2. Blinding ... 106

**6. DOSAGE AND ADMINISTRATION ... 108**
6.1. Part I ... 108
6.2. Part II ... 108
6.3. Part II Long-term Extension ... 109

**7. TREATMENT COMPLIANCE ... 110**

**8. PRESTUDY AND CONCOMITANT THERAPY ... 110**
8.1. Concomitant Medications ... 111
8.1.1. Corticosteroid Tapering ... 112
8.2. Prohibited Medications ... 112

**9. STUDY EVALUATIONS ... 113**
9.1. Study Procedures ... 113
9.1.1. Overview ... 113
9.1.2. Screening Period ... 113
9.1.3. Double-Blind Treatment Period ... 114
9.1.4. Final Efficacy and Safety Follow-up Visit ... 115
9.2. Efficacy Evaluations ... 115
9.2.1. Crohn's Disease Activity Index ... 115
9.2.2. Bristol Stool Form Scale ... 115
9.2.3. Abdominal Pain Numerical Rating Scale ... 115
9.2.4. Patient's Global Impression of Severity of Crohn's Disease ... 116
9.2.5. Patient's Global Impression of Change of Severity of Crohn's Disease ... 116
9.2.6. C-Reactive Protein ... 116
9.2.7. Fecal Lactoferrin and Calprotectin ... 116
9.2.8. Inflammatory Bowel Disease Questionnaire ... 116
9.2.9. 36-Item Short Form Health Survey ... 117
9.2.10. Fistula Assessment ... 117
9.2.11. Endoscopic Endpoints ... 117
9.2.12. Histologic Assessment ... 118
9.3. Pharmacokinetics and Immunogenicity Evaluations ... 118
9.3.1. Evaluations ... 118
9.3.2. Serum Concentration ... 118
9.3.3. Immunogenicity Assessments (Antibodies to Study Agent) ... 119
9.4. Biomarker and Other Pharmacodynamic Evaluations ... 119
9.4.1. Serum-based Biomarkers ... 119
9.4.2. Whole Blood-based Biomarkers ... 119
9.4.3. Biopsy-based Biomarkers ... 119
9.4.4. NKG2D Receptor Occupancy ... 119
9.4.5. Immunophenotyping ... 120
9.5. Pharmacogenomic (DNA) Evaluations ... 120
9.6. Medical Resource Usage ... 120
9.7. Safety Evaluations ... 120
9.8. Sample Collection and Handling ... 124

**10. SUBJECT COMPLETION/DISCONTINUATION OF STUDY TREATMENT/ WITHDRAWAL FROM THE STUDY ... 124**
10.1. Completion ... 124
10.2. Discontinuation of Study Treatment ... 125
10.3. Withdrawal From the Study ... 127
10.4. Withdrawal From the Use of Research Samples ... 127
10.5. Withdrawal From Pharmacogenomic Research Only ... 128

**11. STATISTICAL METHODS ... 128**
11.1. Sample Size Determination ... 128
11.1.1. Sample Size in Part I ... 129

17.1. Protocol Amendments ... 150
17.2. Regulatory Documentation ... 151
17.2.1. Regulatory Approval/Notification ... 151
17.2.2. Required Prestudy Documentation ... 151
17.3. Subject Identification, Enrollment, and Screening Logs ... 152
17.4. Source Documentation ... 152
17.5. Case Report Form Completion ... 153
17.6. Data Quality Assurance/Quality Control ... 153
17.7. Record Retention ... 154
17.8. Monitoring ... 154
17.9. Study Completion/Termination ... 155
17.9.1. Study Completion/End of Study ... 155
17.9.2. Study Termination ... 155
17.10. On-Site Audits ... 155
17.11. Use of Information and Publication ... 156

**REFERENCES ... 158**

**ATTACHMENTS ... 159**

**INVESTIGATOR AGREEMENT ... 172**

#### LIST OF ATTACHMENTS

Attachment 1: Definition of Inadequate Initial Response, Loss of Response, or Intolerance to TNF-Antagonist Therapies (Infliximab, Adalimumab, or Certolizumab Pegol) or Vedolizumab ... 159
Attachment 2: Definitions of Inadequate Response to or Intolerance of Corticosteroids or AZA/6-MP/MTX and Corticosteroid Dependence ... 164
Attachment 3: Tuberculin Skin Testing ... 166
Attachment 4: Crohn's Disease Activity Index ... 167
Attachment 5: Liver Safety Monitoring and Assessment ... 168
Attachment 6: Anticipated Events ... 171

### LIST OF IN-TEXT TABLES AND FIGURES

#### TABLES

Table 1: Time and Events Schedule, Part I ... 62
Table 2: Time and Events Schedule, Part II ... 66
Table 3: Time and Events Schedule, Long-term Extension Before Unblinding for JNJ-64304500, Ustekinumab, and Placebo and After Unblinding for JNJ-64304500[j] ... 71
Table 4: Time and Events Schedule, Long-term Extension Phase After Unblinding in Countries Where Ustekinumab Is Not Commercially Available ... 73
Table 5: Predicted median serum JNJ-64304500 concentrations and NKG2D receptor occupancy after administration of the selected dose regimens of JNJ-64304500 ... 95
Table 6: Power to detect a treatment difference and sample size combinations at an overall Type 1 error of 0.05 (2-sided) for Part I ... 129
Table 7: Power to detect a treatment difference and sample size combinations at an overall Type 1 error of 0.05 (2-sided) for Part II ... 130

#### FIGURES

Figure 1: Schematic representation of 64304500CRD2001 ... 84
Figure 2: Schematic Overview of Part I of Study 64304500CRD2001 ... 86
Figure 3: Schematic Overview of the 64304500CRD2001 Part II (Including the Long-term Extension) ... 87

### PROTOCOL AMENDMENTS

| Protocol Version | Issue Date |
|---|---|
| Original Protocol | 17 March 2016 |
| Amendment 1 | 14 June 2016 |
| Amendment UK-1 | 22 September 2016 (superceded by Amendment 2) |
| Amendment 2 | 09 November 2016 |
| Amendment 3 | 12 December 2017 |
| Amendment 4 | 24 April 2018 |
| Amendment 5 | 17 January 2019 |
| Amendment 6 | 04 February 2020 |

Amendments below are listed beginning with the most recent amendment. The changes to the protocol are indicated with an underline.

**Amendment 6** (04 February 2020)

**The overall reason for the amendment:** To adjust the maximum proportions of Bio-NF subjects enrolled in Part II.

| Applicable Section(s) | Description of Change(s) |
|---|---|
| **Rationale:** The maximum proportion of Bio-NF subjects will be adjusted to 60% (from 50%), which will allow for additional enrollment flexibility, based upon observed enrollment patterns in Part II. The maximum proportion of Bio-IR subjects will remain at 60%. The proposed change should pose no risk to statistical power for Part II, as the power is based on overall population. | |
| 3.1, Overview of Study Design | The following subsection was revised:<br>It is anticipated that approximately 370 to 420 subjects will be randomized overall across Part I and Part II:<br>• **Part I** will study the safety and efficacy of a high dose regimen of JNJ-64304500 compared with placebo and will enroll a minimum of 120 subjects and a maximum of 170 subjects (Bio-IR and Bio-NF).<br>• **Part II** will study the safety and efficacy of multiple-dose regimens of JNJ-64304500 compared with placebo, with ustekinumab (STELARA®) as a reference arm. Part II will enroll approximately 250 subjects (the maximum proportion of either Bio-NF or Bio-IR subjects will be 60%). |
| 3.1.2, Part II | The last sentence in the first paragraph was revised:<br>In Part II, 250 additional Bio-IR or Bio-NF subjects will be randomly assigned to receive placebo or 1 of 3 dose levels of JNJ-64304500 or ustekinumab in a ratio of 1:1:1:1:1 using permuted block randomization, stratified by baseline CDAI score (≤300 or >300), SNP-positive status (yes or no), and Bio-IR status (yes or no). The maximum proportion of either Bio-NF or Bio-IR subjects will be 60%. |

| Applicable Section(s) | Description of Change(s) |
|---|---|
| **Rationale:** Footnotes in the Time and Events Schedules for the Part II Long-term Extension were revised to clarify that PK and immunogenicity samples will be collected at the timepoints shown in the schedules; PK and immunogenicity samples collected at Week 24 are part of the main study, not part of the LTE. | |
| Table 3, Time and Events Schedule, Long-term Extension Before Unblinding for JNJ-64304500, Ustekinumab, and Placebo and After Unblinding for JNJ 64304500[j] | Footnote k was revised:<br>k. Pharmacokinetic and immunogenicity samples will be obtained only at the specified timepoints and when SAEs are reported by study subjects. |
| Table 4, Time and Events Schedule, Long-term Extension Phase After Unblinding in Countries Where Ustekinumab is Not Commercially Available | Footnote i was revised:<br>i. Pharmacokinetic and immunogenicity samples will be obtained only at the specified timepoints and when SAEs are reported by study subjects. |
| **Rationale:** The specified time period for treatment with apheresis as an exclusion criterion (5h) was inadvertently omitted in Protocol Amendment 5 and has been reinstated. | |
| 4.2, Exclusion Criteria | 5. Has received any of the following prescribed medications or therapies within the specified period:<br>…<br>h. Treatment with apheresis (eg, Adacolumn apheresis) <3 weeks before baseline. |
| **Rationale:** Minor errors were corrected. | |
| Throughout the protocol | Minor grammatical, formatting, or spelling changes were made. |

**Amendment 5** (17 January 2019)

**The overall reason for the amendment:** The 3 main reasons for updating this protocol were to reduce the sample size for Part II from 275 to 250 subjects, change the primary endpoint timing for Part II from Week 8 to Week 12, based on the results from the previous Part I Week 12 analysis, and add a Part II long-term extension (LTE) to provide longer term study drug access to eligible subjects for up to 52 weeks. Additionally, the exclusion and discontinuation criteria have been refined for clarification.

| Applicable Section(s) | Description of Change(s) |
|---|---|

**Rationale:** A 52-week LTE is being added to Part II to provide longer-term study drug access to eligible subjects who are benefitting from therapy, and to obtain longer-term safety data for JNJ-64304500.

| | |
|---|---|
| Synopsis; Section 1.2.2, Crohn's Disease; | The following sentence was added:<br><br>A long-term extension (LTE) for Part II of the study was added to the protocol as part of Protocol Amendment 5. After Protocol Amendment 5 is implemented, subjects who complete Part II Week 24 assessments and who may benefit from continued treatment in the opinion of the investigator, are eligible to enter the Part II LTE to receive up to 52 weeks of additional treatment. Subjects who have completed Week 24 assessments prior to the implementation of Protocol Amendment 5 will not be eligible to enroll in the Part II LTE. |
| Section 3.1, Overview of Study Design; Section 3.2, Study Design Rationale | The following sentence was added:<br><br>After Protocol Amendment 5 is implemented, subjects who complete Part II Week 24 assessments and who may benefit from continued treatment in the opinion of the investigator, are eligible to enter the Part II LTE to receive up to 52 weeks of additional treatment. Subjects who have completed Week 24 prior to the implementation of Protocol Amendment 5 will not be eligible to enroll in the Part II LTE. |
| Synopsis, Overview of Study Design | The 2nd paragraph was revised:<br><br>The duration of the study will be 38 weeks in Part I and 36 weeks in Part II <u>for subjects who do not enter the Part II LTE. The study duration includes</u> study agent administration visits and a final efficacy and safety follow-up visit. Eligible subjects will only participate in Part I <u>or</u> Part II of the study.<br><br>The following was added as the 3rd paragraph:<br><br>After Protocol Amendment 5 is implemented, subjects who complete Part II of the study through Week 24 and who may benefit from continued treatment in the opinion of the investigator, will be eligible to enter the Part II LTE to receive up to 52 weeks of additional treatment. |
| Synopsis, Dosage and Administration, Long-term Extension; Section 6.3, Part II Long-term Extension | The following content was added:<br><br>After Protocol Amendment 5 is implemented, subjects who complete Part II Week 24 assessments, and who may benefit from continued treatment in the opinion of the investigator, are eligible to enter the Part II LTE. In the Part II LTE, each subject will be eligible to receive up to 52 weeks of additional study drug (for a total of up to 72 weeks of study drug in Part II). The first dose of study drug within the Part II LTE will be administered after the Week 24 assessments have been completed. The dose may be administered on the same day as the Week 24 assessments, or may be administered on a later date, as long as the dose is administered within the Week 24 visit window.<br><br>Subjects will continue to receive the same treatment regimen during the Part II LTE that they were receiving between Week 12 and Week 20 in the main study phase of Part II (placebo, high, middle, low dose JNJ-64304500 or ustekinumab). To maintain the study blind, all patients, investigators, and sites will remain blinded to treatment allocation during the Part II LTE until the last subject in the Part II main study phase has completed the Week 24 assessments and the Week 24 data analysis has been completed.<br><br>The timing of the Week 24 data analysis is dependent upon the timing and completion of Part II enrollment. An individual subject's unblinding during the Part II LTE will depend upon his/her Part II enrollment date. Therefore, a portion of subjects will complete the entire Part II LTE in a blinded fashion before study unblinding, while other subjects could |

be unblinded to treatment allocation during their participation in the Part II LTE.

Any subject who withdraws from the Part II LTE prior to study unblinding will return for a final safety follow-up visit 16 weeks after the last dose of study drug. For subjects that remain in the Part II LTE at the time of study unblinding, continuation of study drug during the Part II LTE will be dependent upon treatment allocation as follows:

- **JNJ-64304500:** Subjects will continue to receive the same dose of JNJ-64304500 administered during the Part II main study phase. Subjects will continue to receive study drug q4w, although any additional placebo injections used to maintain the study blind during the blinded phase of the Part II LTE will be discontinued after study unblinding. Following the last scheduled dose of study drug at Week 72, a final safety follow-up visit will be performed at approximately Week 88. For subjects who discontinue study participation prior to Week 72, a final safety follow-up visit will be performed 16 weeks after the last dose of study drug.
- **Placebo**: Subjects receiving placebo who remain in the Part II LTE at the time of study unblinding will have their final safety follow-up visit subsequent to unblinding and will be discontinued from the study. No further follow-up visits will be performed.
- **Ustekinumab**: Subjects receiving ustekinumab will continue to receive it during the Part II LTE until unblinding occurs. After unblinding, based on the treating physician's clinical judgment, subjects may receive ustekinumab in a manner dependent on the country in which they are located:
  - If a subject is not continuing on ustekinumab after unblinding, the subject will need to return for a final safety follow-up visit that will be performed 16 weeks after the final dose of study ustekinumab.
  - If a subject continues on ustekinumab in a country where commercial ustekinumab is available and approved for the treatment of adult Crohn's disease, the treating physician is responsible for providing the subject with commercial ustekinumab, which should be administered according to the approved dosing regimen in each country. The final safety follow-up visit should be performed after unblinding but before receiving the first dose of commercial ustekinumab.
  - If a subject continues on ustekinumab in countries where commercial ustekinumab is not available or approved for adult Crohn's disease, ustekinumab will continue to be provided to subjects through Week 72, followed by a final safety follow-up visit 16 weeks after the last dose of ustekinumab.

For eligible Part II subjects, participation in the Part II LTE is entirely voluntary. Eligible subjects who do not wish to enter the Part II LTE will complete the Part II Week 24 safety and efficacy assessments, followed by the final efficacy and safety assessments at Week 36.

During the Part II LTE, all concomitant medications, including Crohn's disease-specific medications (except for the prohibited medications listed in Section **8.2**) may be administered at the discretion of the investigator.

All subjects in the Part II LTE will be assessed according to the Time & Events Schedule (Table 3, Table 4), which includes assessments, adverse events (AEs), laboratory analyses,

| | |
|---|---|
| | and PK and immunogenicity samples. |
| Synopsis, Efficacy Evaluations; Section 3.1, Overview of Study Design; Section 3.2.2, Efficacy Assessments | The following sentence was added:<br>Efficacy evaluations will not be performed during the Part II LTE. |
| Table 2, Time and Events Schedule: Part II Main Study Phase | Urine pregnancy test removed from Table 2 (Time and Events Schedule: Part II Main Study Phase) and added to Table 3 (Part II Long-term extension).<br>Footnotes b, c, d, and v updated:<br>b. Visit windows should be ±4 days for each visit up to and including Week 12; from Week 16 to end of study, visit window should be ± 7 days.<br>c. Subjects who terminate study participation should complete an early termination visit. If a subject completed assessments at Week 24 and terminated after Week 24, the only assessments that should be performed at the early termination visit are those planned for the final efficacy and safety follow-up visit. Subjects who discontinue study agent administration before Week 12 (but have not terminated study participation) should complete the Week 12 assessments and return for a final efficacy and safety follow-up visit approximately 16 weeks after their last study agent administration. Subjects who discontinue study agent administration after Week 12 (but have not terminated study participation) should complete the Week 24 assessments and return for a final efficacy and safety follow-up visit approximately 16 weeks after their last study agent administration.<br>d. Subjects should complete a final efficacy and safety follow-up visit approximately 16 weeks after their last study agent administration. For subjects who complete all visits and are not enrolling in the Part II LTE, this will occur at Week 36.<br>v. At all visits when study intervention will be administered, 1 blood sample should be collected prior to study intervention administration for evaluation of serum concentrations and/or antibodies to the study interventions. For the IV infusion related visit (Week 0), the SC study intervention should be administered first, followed by IV infusion; another blood draw should be taken approximately 60 minutes after completion of the infusion for serum concentration measurement.<br>Footnote f added to Table 2. |
| Table 3, Time and Events Schedule: Part II Long-Term Extension | Time and Events Schedule: Time and Events Schedule, Long-term Extension Before Unblinding for JNJ-64304500, Ustekinumab, and Placebo and After Unblinding for JNJ-64304500 was added. |
| Table 4, Time and Events Schedule: Part II Long-Term Extension | Time and Events Schedule: Long-term Extension Phase After Unblinding in Countries Where Ustekinumab is Not Commercially Available was added. |

| | |
|---|---|
| Section 3.1, Overview of Study Design | The following sentence was added to the eleventh paragraph:<br>After Protocol Amendment 5 is implemented, all subjects who complete Part II of the study through Week 24, and who may benefit from continued treatment in the opinion of the investigator, are eligible to enter the Part II LTE and continue to receive treatment for up to 52 weeks (Week 24 to Week 72), as described in Section 3.1.4. A schematic representation of the Part II LTE is shown in Figure 3. |
| Section 3.1, Overview of Study Design | Figure 1, Schematic representation of 64304500CRD2001 was updated to include the Part II LTE. |
| Section 3.1, Overview of Study Design | Figure 3, Schematic Overview of the 64304500CRD2001 Part II (Including the Long-term Extension) was added. |
| Section 3.1.4, Long-Term Extension | The following content was added:<br>After Protocol Amendment 5 is implemented, subjects who complete Part II Week 24 assessments, and who may benefit from continued treatment in the opinion of the investigator, are eligible to enter the Part II LTE. In the Part II LTE, each subject will be eligible to receive up to 52 weeks of additional study drug (for a total of up to 72 weeks of study drug in Part II). The first dose of study drug within the Part II LTE will be administered after the Week 24 assessments have been completed. The dose may be administered on the same day as the Week 24 assessments, or may be administered on a later date, as long as the dose is administered within the Week 24 visit window.<br><br>Subjects will continue to receive the same treatment regimen during the Part II LTE that they were receiving between Week 12 and Week 20 in the main study phase of Part II (placebo, high, middle, low dose JNJ-64304500 or ustekinumab). To maintain the study blind, all patients, investigators, and sites will remain blinded to treatment allocation during the Part II LTE until the last subject in the Part II main study phase has completed the Week 24 assessments and the Week 24 data analysis has been completed.<br><br>The timing of the Week 24 data analysis is dependent upon the timing and completion of Part II enrollment. Whether an individual subject will receive his/her treatment allocation during the Part II LTE will depend upon his/her Part II enrollment date. Therefore, a portion of subjects will complete the entire Part II LTE in a blinded fashion before study unblinding, while other subjects could be unblinded to treatment allocation during their participation in the Part II LTE.<br><br>Any subject who withdraws from the Part II LTE prior to study unblinding will return for a final safety follow-up visit 16 weeks after the last dose of study drug. For subjects that remain in the Part II LTE at the time of study unblinding, continuation of study drug during the Part II LTE will be dependent upon treatment allocation as follows:<br><br>• **JNJ-64304500**: Subjects will continue to receive the same dose of JNJ-64304500 administered during the Part II main study phase. Subjects will continue to receive study drug q4w, although any additional placebo injections used to maintain the study blind during the blinded phase of the Part II LTE will be discontinued after study unblinding. Following the last scheduled dose of study drug at Week 72, a final safety follow-up visit will be performed at approximately Week 88. For subjects who discontinue study participation prior to Week 72, a final safety follow-up visit will be performed 16 weeks after the last dose of study drug.<br><br>• **Placebo**: Subjects receiving placebo who remain in the Part II LTE at the time of study unblinding will have their final safety follow-up visit subsequent to unblinding |

and will be discontinued from the study. No further follow-up visits will be performed.

- **Ustekinumab**: Subjects receiving ustekinumab will continue to receive it during the Part II LTE until unblinding occurs. After unblinding, based on the treating physician's clinical judgment, subjects may receive ustekinumab in a manner dependent on the country in which they are located:
  - If a subject is not continuing on ustekinumab after unblinding, the subject will need to return for a final safety follow-up visit that will be performed 16 weeks after the final dose of study ustekinumab.
  - If a subject continues on ustekinumab in a country where commercial ustekinumab is available and approved for adult Crohn's disease, the treating physician is responsible for providing the subject with commercial ustekinumab, which should be administered according to the approved dosing regimen in each country. The final safety follow-up visit should be performed after unblinding but before receiving the first dose of commercial ustekinumab.
  - If a subject continues on ustekinumab in countries where commercial ustekinumab is not available or approved for adult Crohn's disease, ustekinumab will continue to be provided to subjects through Week 72, followed by a final safety follow-up visit 16 weeks after the last dose of study drug.

For eligible Part II subjects, participation in the Part II LTE is entirely voluntary. Eligible subjects who do not wish to enter the Part II LTE will complete the Part II Week 24 safety and efficacy assessments, followed by the final efficacy and safety assessments at Week 36.

During the Part II LTE, all concomitant medications, including Crohn's disease-specific medications (except for the prohibited medications listed in Section 8.2) may be administered at the discretion of the investigator.

All subjects in the Part II LTE will be assessed according to the Time & Events Schedule (Table 3, Table 4), which includes assessments, AEs, laboratory analyses, and PK and immunogenicity samples.

Section 4.2, Exclusion Criteria

The following content was added to the end of this section:

**Long-term Extension Phase**

After Protocol Amendment 5 is implemented, subjects who complete Part II Week 24 assessments and who may benefit from continued treatment in the opinion of the investigator, are eligible to enter the Part II LTE to receive up to 52 weeks of additional treatment. Subjects must be excluded from LTE if they have developed Crohn's disease complications which may be anticipated to require surgery in the next 12 months, if they have clinical evidence of a serious infection at Week 24, or if they intend to participate in any other study using an investigational agent or procedure during participation in the LTE. Investigators who have questions regarding subject eligibility for the LTE should contact the sponsor.

Section 5.2, Blinding

The third bullet point was added:

- **Final DBL for Part I:** Occurs when all Part I subjects have completed their final efficacy and safety visit or have terminated their study participation before the final efficacy and safety visit.

The fourth bullet point was revised:

- **Week 12 DBL for Part II ~~(final DBL for Part I occurs at the same time)~~:** Occurs when all Part II subjects have completed their Week 12 visit or have terminated their study participation before Week 12.

The sixth bullet point was revised:

- **Final DBL for Part II:** Occurs when all Part II subjects who entered the Part II LTE have completed their final safety visit in the Part II LTE or have terminated their study participation before the final safety visit in the Part II LTE.

The 3rd paragraph was revised:

At the time of the interim analysis lock for Part I, a limited number of sponsor personnel will become unblinded to treatment assignment. At the time of the Week 12 DBL for Part I and the Week 12 DBL for Part II, the sponsor, except for site monitors, who have interactions with the investigative sites, will become unblinded to treatment assignment. Identification of sponsor personnel who will have access to subject-level data for the interim analysis lock for Part I will be documented before the unblinding. The study blind will be maintained for investigators, site personnel, subjects, and ~~sponsor~~ site monitors until the ~~final~~ Week 24 analyses have been completed for ~~all subjects in Part I and~~ Part II. This measure will mitigate the potential bias in the remaining investigator and subject assessments.

Section 8.1, Concomitant Medications

The following paragraph was added as the sixth paragraph in the section:

Subjects participating in the Part II LTE may continue receiving concomitant medications they were receiving at the time of Part II Week 24, including aminosalicylates, immunomodulatory agents (including 6-MP/AZA, MTX), enteral therapy, and corticosteroids. During the Part II LTE, these medications may be initiated, discontinued, or dose adjusted at the investigator's discretion.

Section 8.1.1, Corticosteroid Tapering

The following sentence was added as the 3rd paragraph:

During the Part II LTE, corticosteroids may be initiated, discontinued, or dose adjusted at the investigator's discretion

Section 8.2, Prohibited Medications

The first sentence in the section was revised:

Randomized subjects must not initiate any of the following prohibited medications at any time during Part I, Part II, or the Part II LTE:

Section 9.1.1, Overview

Added reference to Table 3, Time and Events Schedule, Long-term Extension Before Unblinding for JNJ-64304500, Ustekinumab, and Placebo and After Unblinding for JNJ-64304500 and Table 4: Time and Events Schedule, Long-term Extension Phase After Unblinding in Countries Where Ustekinumab is Not Commercially Available.

The last paragraph was updated:

The maximum amount of blood drawn from each subject in this study will be approximately 330 mL.

**Section 9.1.3, Double-Blind Treatment Period**

The last sentence in this section was revised:

Each subject must be instructed to complete the CDAI diary card daily during the study, except during the Part II LTE.

**Section 9.1.4, Final Efficacy and Safety Follow-up Visit**

This section was revised:

Subjects in Part I ~~and Part II~~ should complete the final efficacy and safety follow-up visit approximately 16 weeks after the last study agent administration (Table 1~~, Table 2~~). Subjects in Part II who are not continuing into the Part II LTE should complete the final efficacy and safety follow-up visit approximately 16 weeks after receiving the last dose of study drug (Table 2). Subjects in the Part II LTE should complete the final safety follow-up visit approximately 16 weeks after receiving the last dose of study drug (Table 3, Table 4). Placebo subjects who remain in the Part II LTE at the time of study unblinding will have their final safety follow-up visit subsequent to unblinding and will be discontinued from the study.

**Section 10.1, Completion**

The original section was updated to become a subsection: Main Study Phase (Part I and Part II)

The bullet points in the Main Study Phase (Part I and Part II) section were revised:

- Subjects who discontinue study agent in Part I before Week 8 should return for the Week 8 and Week 12 visit and a final efficacy and safety follow-up visit, which should occur 16 weeks after receiving the last dose of study drug.
- Subjects who discontinue study agent at or after Week 8 in Part I should complete the Week 12 and Week 24 visit and a final efficacy and safety follow-up visit, which should occur 16 weeks after receiving the last dose of study drug.
- Subjects who discontinue study agent in Part II before Week 12 should return for the Week 12 visit and a final efficacy and safety follow-up visit, which should occur 16 weeks after receiving the last dose of study drug.
- Subjects who discontinue study agent at or after Week 12 in Part II should complete the Week 24 visit and a final efficacy and safety follow-up visit, which should occur 16 weeks after receiving the last dose of study drug.

A new subsection was added:

Long-term Extension Phase

A subject will be considered to have completed the long-term extension phase of the study if he or she has completed the final safety follow-up visit in the Part II LTE.

The final safety follow-up visit will occur approximately 16 weeks after last study agent administration for all subjects who received JNJ-64304500 or ustekinumab. If the investigator decides that the patient should continue ustekinumab treatment after completing the Part II LTE, the investigator or the subject's treating physician is responsible for providing the subject with commercial ustekinumab, which should be administered according to the approved dosing regimen in each country. Refer to Section 3.1.4, Part II Long-Term Extension, for details regarding the ustekinumab follow-up visit.

Subjects who received placebo will be considered to have completed the extension phase of the study if they complete 52 weeks of the Part II LTE and complete the final safety

| | |
|---|---|
| | follow-up visit in the Part II LTE at before study unblinding or if they have been discontinued from the study prior to the final safety visit due to study unblinding. After unblinding of the Part II Week 24 data, subjects who received placebo during the Part II LTE will have their final safety follow-up visit at the time of study unblinding and will then be discontinued from the study. Subjects who prematurely discontinue study treatment for any reason, other than study unblinding, before completion of the study extension will not be considered to have completed the extension phase of the study. |

**Rationale:** The assumptions for the sample size calculations in Part II have been refined based on observed efficacy data from the Part I Week 12 analysis. The sample size for Part II has been reduced from 275 to 250 (reduction of 25 subjects, or 5 subjects per arm).

| | |
|---|---|
| Synopsis, Overview of Study Design | The first paragraph, 4th sentence revised:<br><br>An additional 250 subjects will be randomized in Part II. |
| Synopsis, Statistical Methods, Sample Size Determination | The first paragraph was revised:<br><br>Sample size calculations for Part I (Bio-IR subjects and Bio-NF subjects combined) were determined by the power to detect a significant difference in the change from baseline in the CDAI score at Week 8 between JNJ-64304500 and placebo using a 2-sample t-test. Sample size calculations for Part II (Bio-IR and Bio-NF subjects) were determined by the power to detect a dose-response signal for the change from baseline in the CDAI score at Week 12 (primary endpoint in Part II) using the Multiple Comparison Procedures with modeling techniques (MCP-Mod) method.<br><br>The third paragraph was revised:<br><br>For Part II, assuming the mean CDAI change from baseline at Week 12 is -111 in the JNJ-64304500 group and -51 in the placebo group with a common SD of 102, 50 subjects per treatment group will provide a mean power of 85% to detect a dose-response signal based on 7 candidate dose-response models at an overall Type 1 error rate of 0.05. |
| Section 3.1, Overview of Study Design | The first sentence and the second bullet point of the seventh paragraph was revised:<br><br>It is anticipated that approximately 370-420 subjects will be randomized overall across Part I and Part II:<br><br>• **Part II** will study the safety and efficacy of multiple-dose regimens of JNJ-64304500 compared with placebo, with ustekinumab (STELARA®) as a reference arm. Part II will enroll approximately 250 subjects (Bio-NF subjects will be no more than 50% and Bio-IR subjects will be no more than 60%). |
| Section 3.1.2, Part II | The first sentence of the first paragraph was revised:<br><br>In Part II, 250 additional Bio-IR or Bio-NF subjects will be randomly assigned to receive placebo or 1 of 3 dose levels of JNJ-64304500 or ustekinumab in a ratio of 1:1:1:1:1 using permuted block randomization, stratified by baseline CDAI score (≤300 or >300), SNP-positive status (yes or no), and Bio-IR status (yes or no). |

| | |
|---|---|
| Section 11.1, Sample Size Determination | This section was revised:<br><br>Sample size calculations for Part I ~~and Part II~~ were determined by the power to detect a significant difference in the change from baseline in the CDAI score at Week 8 (primary endpoint for Part I) between JNJ-64304500 and placebo using a 2-sample t-test. Sample size calculations for Part II were determined by the power to detect a dose-response signal for the change from baseline in CDAI at Week 12 (primary endpoint for Part II) using the Multiple Comparison Procedures with modeling techniques (MCP-Mod) method. |
| Section 11.1.1, Sample Size in Part I | The title of Table 6 was revised to reflect the sample size of Part I. |
| Section 11.1.2, Sample Size in Part II | The entire section was revised:<br><br>The assumptions for the sample size calculations in Part II were based on the Part I Week 12 analysis results. For Bio-IR subjects in Part I, the mean CDAI change from baseline at Week 12 was -30.9 (SD=85.15) and -77.4 (SD=103.68) for the placebo and JNJ-64304500 groups, respectively. For Bio-NF subjects in Part I, the mean CDAI change from baseline at Week 12 was -70.9 (SD=93.59) and -144.7 (SD=92.50) for the placebo and JNJ-64304500 groups, respectively.<br><br>For Part II, assuming the mean CDAI change from baseline at Week 12 is -111 in the JNJ-64304500 high dose group and -51 in the placebo group (these values are derived assuming a 1:1 ratio of Bio-IR and Bio-NF subjects) with a common SD of 102, 50 subjects per treatment group will provide a mean power of 85% to detect a dose-response signal for the change from baseline in CDAI at Week 12 based on 7 candidate dose-response models (to be detailed in the SAP) at an overall Type 1 error rate of 0.05 (2-sided). Fifty subjects per treatment group will also provide 83% power to detect a treatment difference between the JNJ-64304500 treatment group with the highest dose and the placebo treatment group for change from baseline in CDAI at Week 12 based on a 2-sample t-test at a Type I error rate of 0.05 (2-sided; Table 7). This will result in a total sample size of 250 subjects in Part II (incorporating an additional 50 subjects for the ustekinumab treatment group).<br><br>Table 7 added to reflect the sample size in Part II. |

**Rationale:** The timing for the primary endpoint and major secondary endpoints for Part II (except for the endoscopy endpoint) will be modified from Week 8 to Week 12, based on the results from the previous Part I Week 12 analysis. This change will also align the timing of the primary endpoint and major secondary endpoints to the planned Week 12 Part II endoscopy endpoint.

| | |
|---|---|
| Synopsis, Primary Endpoints; Section 2.1.2, Endpoints; Section 3.1, Overview of Study Design (9th paragraph); Section 11.2.2.1, Primary Endpoint Analyses | The primary endpoint was revised:<br><br>The primary endpoint for Part I ~~and Part II~~ is: Change from baseline in the CDAI score at Week 8.<br><br>The primary endpoint for Part II is: Change from baseline in the CDAI score at Week 12. |

Synopsis, Major Secondary Endpoints; Section 2.1.2, Endpoints; Section 11.2.2.2, Major Secondary Endpoint Analyses

The major secondary endpoints were revised:

- Clinical remission at Week 12 as measured by CDAI (CDAI <150).
- Clinical response at Week 12 as measured by CDAI (≥100-point reduction from baseline in CDAI or CDAI <150).
- Change in PRO-2 (the sum of the abdominal pain and stool frequency subscores of the CDAI score) from baseline at Week 12.
- Clinical remission at Week 12 as measured by PRO-2 (PRO-2 <75).
- Clinical response at Week 12 as measured by PRO-2 (≥50-point reduction from baseline in PRO-2 or PRO-2 <75).

Synopsis, Efficacy Analyses

The 4th paragraph was revised:

The primary endpoint for Part II is ~~also~~ the change from baseline in the CDAI score at Week 12. A unified strategy that combines Multiple Comparison Procedures with modeling techniques, MCP-Mod, will be used to analyze the dose-response data over the JNJ-64304500 doses. This approach consists of 2 major steps. The first step consists of testing the dose-response signal via multiple contrast tests while controlling the overall Type 1 error. If a dose-response signal is detected, the second step is to select a model that best describes the observed data and use it to estimate adequate doses with associated precision. The study will be considered positive if a dose-response signal for the primary endpoint is detected. In addition to the dose-response analysis, pairwise comparisons of the JNJ-64304500 treatment groups versus the placebo group will be performed for the change from baseline in the CDAI score at Week 12; these comparisons will not be adjusted for multiplicity.

Section 2.1.2, Endpoints; Section 11.2.1.2, Other Efficacy Endpoint Analyses

Three efficacy endpoints were updated:

- Clinical remission based on CDAI at Week 24 among subjects in clinical response at Week 12.
- Clinical remission based on CDAI at Week 24 among subjects in clinical remission at Week 12.
- Clinical remission based on CDAI at Week 12 by SNP status. Subjects who are positive in at least 1 of 2 SNPs (NKG2D or MICB) will be considered to be SNP-positive.

Section 3.1, Overview of Study Design

Figure 3 was updated to reflect that the primary endpoint analysis occurs at Week 12 instead of Week 8. It was also updated with a note that the Final Safety and Efficacy Visit will occur at Week 36 if the subject is not enrolling in the Part II LTE.

| | |
|---|---|
| Section 3.2.2, Efficacy Assessments | This section was revised to:<br><br>The efficacy evaluations selected for both parts of the study (eg, CDAI, CRP, fecal biomarkers; Section 9.2) are well-established measures that are accepted by regulatory agencies as primary or supportive of clinically relevant effect of disease activity in Crohn's disease studies.<br><br>The primary endpoint for Part I is change from baseline in CDAI at Week 8. The primary endpoint for Part II is change from baseline in CDAI at Week 12.<br><br>Change in the CDAI is being used as the primary endpoint because this measure is more sensitive than remission (ie, the change in CDAI provides greater power than remission for the same sample size). Therefore, the study can be more efficient for Phase 2 using the change in CDAI.<br><br>To evaluate the level of efficacy after prolonged discontinuation of study drug, CDAI will be assessed at the final efficacy and safety visit in Part I and Part II for those subjects who do not enter the Part II LTE. The Crohn's Disease Activity Index will be not assessed during the Part II LTE.<br><br>It is anticipated that endoscopic improvement will occur by Week 12. In order to have an appropriate comparison of JNJ-64304500 to placebo, the placebo-controlled period will continue to Week 12.<br><br>Efficacy evaluations will not be performed during the Part II LTE. |
| Section 9.1.3, Double-Blind Treatment Period | The first sentence in the first paragraph was revised to:<br><br>In Part I, the visit window should be ±4 days for each visit. In Part II, the visit window should be ±4 days, from the Week 0 visit up to and including the Week 12 visit, and ±7 days from Week 16 until the end of the study. |
| Section 11.2.2.1, Primary Endpoint Analysis; Section 11.2.2.2, Major Secondary Endpoint Analyses | All references to Week 8 were changed to Week 12. |

**Rationale:** The Part II Week 24 optional endoscopy was removed. The optional Week 24 endoscopy in Part II was intended to obtain additional biomarker data. Similar to Part I, mandatory baseline and Week 12 endoscopies will be performed on all subjects in Part II, which will be used to assess efficacy in conjunction with the primary efficacy endpoint (change in CDAI from baseline at Week 12). Elimination of the optional Week 24 endoscopy in Part II will therefore not impact the primary Part II endpoint and is expected to reduce the burden upon the patients, sites, and investigators at the Week 24 visit.

| | |
|---|---|
| Table 2, Time and Events Schedule: Part II | Video ileocolonoscopy and ileocolonoscopy biopsy sample collection for RNA and histology were removed from the Week 24 visit.<br><br>Footnote t was removed; the Week 24 video ileocolonoscopy and related endoscopic assessments are optional. |

| | |
|---|---|
| Section 9.2.11, Endoscopic Endpoints | The first paragraph was revised:<br>Endoscopic improvement will be assessed during endoscopy (ileocolonoscopy). A video ileocolonoscopic examination will be performed to determine the presence or absence of mucosal inflammation and ulceration at screening and Week 12, and Week 24 (optional in Part I), according to the Study Reference Manual provided to each site; if the video ileocolonoscopic examination is not performed on the day of the visit, it must be performed at least 8 days before the Week 0 visit and no more than 8 days before the Week 12 visit. The Week 24 video ileocolonoscopy in Part I is suggested but not required; if performed, it should occur not more than 8 days before the Week 24 visit. The video ileocolonoscopy will not be performed at Week 24 in Part II. Video endoscopies will be assessed by a central facility that will be blinded to treatment group and study visit. A complete video endoscopic examination does not require assessment of the terminal ileum if it cannot be visualized. |

**Rationale:** Guidelines for administration of parenteral nutrition and enteral nutrition were updated. As a safety mitigation measure, exclusion of total parenteral nutrition has now been broadened to exclusion of both total and partial parenteral nutrition administered through any indwelling catheter. The exclusion of subjects recently initiating enteral therapy is added because initiation of enteral therapy may confound the efficacy endpoint evaluation in this study.

| | |
|---|---|
| Section 4.2, Exclusion Criteria | Footnote 5i was added:<br>Initiation of total (complete) or partial (supplemental) parenteral nutrition administered through any indwelling catheter <3 weeks before baseline or anticipated to require parenteral nutrition administered through an indwelling catheter during enrollment in the study.<br>Footnote 5j was added:<br>Initiation of enteral therapy for Crohn's disease (defined as liquid nutritional formula comprising ≥80% of total caloric intake administered through the gastrointestinal tract) <3 weeks before baseline. Subjects who are on a stable regimen of enteral feeds ≥3 weeks before baseline may be considered for enrollment if they plan to continue enteral feeds as treatment for Crohn's disease through the duration of the study (Week 24, Part I or Part II). |
| Section 8.1, Concomitant Medications | The first paragraph was revised to add enteral therapy:<br>Subjects who are receiving oral 5-ASA compounds, oral corticosteroids, conventional immunomodulators (ie, AZA, 6-MP, or MTX), enteral therapy, and/or antibiotics for the treatment of Crohn's disease at baseline should maintain a stable dose for a specified period before baseline, as defined in the Inclusion Criteria (Section 4.1).<br>The 3rd paragraph, 5th bullet point was revised to:<br>• Total (complete) or partial (supplemental) parenteral nutrition administered through an indwelling catheter as a treatment for Crohn's disease.<br>A 6th bullet point was added to the 3rd paragraph:<br>• Enteral therapy (liquid nutritional formula comprising ≥80% of total caloric intake) as treatment for Crohn's disease. |

**Rationale:** Discontinuation of study treatment guidelines were updated for clarification.

<table>
<tr><td>Section 10.2, Discontinuation of Study Treatment</td><td>The 5th bullet point was revised:<br><br>Adverse events of NCI-CTCAE grade ≥3 ~~potentially related to worsening of Crohn's disease~~ will be evaluated by the investigator and the study medical monitor to make a determination on discontinuation of study agent. Discontinuation of study agent should be considered in subjects with worsening Crohn's disease where continuation of the study drug is not in the best interest of the subject.<br><br>The following was added as the 7th bullet point:<br><br>Total (complete) or partial (supplemental) parenteral nutrition is initiated through an indwelling catheter at any time during the study.<br><br>The 2nd paragraph was revised from:<br><br>Discontinuation of study agent administration must be considered for subjects who develop a severe injection-site or infusion reaction or who develop a serious infection.<br><br>To:<br><br>In the event of any serious infection, severe injection-site or infusion reaction, the study drug must be held and dosing may not be resumed until the investigator has discussed the case with the study medical monitor.</td></tr>
</table>

**Rationale:** Attachment 2: Definitions of Inadequate Response to or Intolerance of Corticosteroids or AZA/6-MP/MTX and Corticosteroid Dependence was updated to include Methotrexate (MTX). There is no change to the originally intended protocol criteria. The specified criteria for MTX were erroneously omitted in the original protocol.

<table>
<tr><td>Attachment 2: Definitions of Inadequate Response to or Intolerance of Corticosteroids or AZA/6-MP/MTX and Corticosteroid Dependence</td><td>The content in this attachment was revised:<br><br>CORTICOSTEROIDS<br><br>Subjects have failed to respond to corticosteroids if they have had evidence of an initial inadequate response, recurrent disease, or a relapse despite receiving at least 0.75 mg/kg/day or ≥40 mg/day of prednisone (or corticosteroid equivalent, given orally or intravenously) for 2 weeks; or ≥9 mg/day of budesonide or ≥5 mg/day of beclomethasone dipropionate given orally for at least 4 weeks.<br><br>Subjects are intolerant of corticosteroids if:<br><br>• They have developed clinically significant adverse events (eg, osteonecrosis or osteoporosis, psychosis, uncontrolled diabetes) unresponsive to dose reduction that, in the judgment of the investigator, precluded the use of corticosteroids to treat Crohn's disease.<br><br>OR<br><br>• They have a medical condition that precludes the use of corticosteroids as a treatment for Crohn's disease.<br><br>Subjects are corticosteroid dependent if they have failed to successfully taper their corticosteroid (ie, had a flare of disease) within 3 months of starting therapy, or if a relapse occurs within 3 months after stopping corticosteroids or if they are unable to discontinue these agents without flare within 3 months after starting them.<br><br>6-MERCAPTOPURINE (6-MP), AZATHIOPRINE (AZA), OR METHOTREXATE (MTX):<br><br>Subjects have failed to respond to 6-MP, AZA, or MTX if they have had evidence of an</td></tr>
</table>

| | |
|---|---|
| | initial inadequate response, recurrent disease, or a relapse despite receiving:<br>• At least 3 months of therapy with 1 mg/kg/day of 6-MP, 2 mg/kg/day of AZA, or 25 mg/week (intramuscular or subcutaneous) of MTX.<br>**OR**<br>• A lower dosage of 6-MP, AZA, or MTX when country or local guidelines specify a different treatment regimen. (In such an event, the country or local guidelines needs to be included in the source document).<br>**OR**<br>• The dosage of 6-MP, AZA, or MTX confirmed to be therapeutic for the subject with whole blood thioguanine nucleotide levels >200 pmole/8x10[8] red blood cells.<br>**OR**<br>• The highest tolerated dosage due to leukopenia, elevated liver enzymes, or nausea.<br>**Subjects are intolerant of 6-MP, AZA, or MTX if:**<br>• They have developed clinically significant adverse events (eg, pancreatitis, arthritis accompanied by high fever and/or rash, leukopenia, or persistently elevated liver enzymes) unresponsive to dose reduction that, in the judgment of the investigator, precluded the use of 6-MP, AZA, or MTX to treat Crohn's disease within the past 5 years.<br>**OR**<br>• They have a medical condition that precludes the use of 6-MP, AZA, or MTX. |
| **Rationale:** Attachment 3: QuantiFERON-TB Gold Testing was removed since most of this information is included in the Site Laboratory Manual and is not necessary to be included in the protocol. QuantiFERON-TB Gold test has been updated to QuantiFERON-TB test since there may be regional or country-specific variability in the type of QuantiFERON-TB test used based on changes in performance of the test by the central lab. | |
| Table 1, Time and Events Schedule: Part I; Table 2, Time and Events Schedule: Part II; Section 4.1, Inclusion Criteria #11.d.1; Section 9.1.2, Screening Period; Section 9.7, Safety Evaluations; Section 10.2, Discontinuation of Study Treatment | QuantiFERON-TB Gold test revised to QuantiFERON-TB test. |
| Attachment 3: QuantiFERON-TB Gold Testing | Removed from protocol. |
| **Rationale:** Minor errors were noted. | |
| Throughout the protocol | Minor grammatical, formatting, or spelling changes were made. |

**Amendment 4** (24 April 2018)

**The overall reason for the amendment:** To revise the study drug concentrations in Section 6.2 for the JNJ-64304500 low and middle doses in Part II of the study to correspond to the Investigational Product Preparation Instructions dilution regimen.

| Applicable Section(s) | Description of Change(s) |
|---|---|
| **Rationale:** To clarify the study drug concentrations of JNJ-64304500 in Part II of the study. | |
| 6.2 Part II | The 3rd bullet is revised from:<br>JNJ-64304500 middle dose: 150 mg SC at Week 0 and 75 mg SC at Weeks 2, 4, 8, 12, 16, and 20. (Study drug concentration=100 mg/mL and 50 mg/mL at Week 0, 75 mg/mL for subsequent doses)<br>To:<br>JNJ-64304500 middle dose: 150 mg SC at Week 0 and 75 mg SC at Weeks 2, 4, 8, 12, 16, and 20 (study drug concentration=75 mg/mL).<br>The 4th bullet is revised from:<br>JNJ-64304500 low dose: 50 mg SC at Week 0 and 25 mg SC at Weeks 2, 4, 8, 12, 16, and 20. (Study drug concentration=50 mg/mL at Week 0, 25 mg/mL for subsequent doses)<br>To:<br>JNJ-64304500 low dose: 50 mg SC at Week 0 and 25 mg SC at Weeks 2, 4, 8, 12, 16, and 20 (study drug concentration=25 mg/mL). |
| **Rationale:** Minor errors were noted | |
| Throughout the protocol | Minor grammatical, formatting, or spelling changes were made. |

**Amendment 3** 12 December 2017

This amendment is considered to be substantial based on the criteria set forth in the Article 10(a) of Directive 2001/20/EC of the European Parliament and the Council of the European Union.

The overall reason for the amendment: To change the timing and subject population for the Interim Analysis in Part I; change the overall sample size of Part I; and change the subject population and sample size in Part II.

Clarification of specific study procedures is also included in this amendment as described below.

| Applicable Section(s) | Description of Change(s) |
|---|---|
| **Rationale (To change the timing and subject population for the Interim Analysis in Part I):** The original planned interim analysis was solely based on the Bio-IR population. It was projected that this study would enroll the Bio-IR population before the Bio-NF population. In actuality, both populations are enrolling at a similar rate. The change in the subject population reflects the current recruitment rate and will enable a decision based upon both populations. This is important considering the population in Part II will now also include Bio-NF subjects. In addition, a change from Week 8 to Week 12 for the interim analysis allows endoscopic data to be incorporated into the decision on whether to initiate Part II. | |

| Applicable Section(s) | Description of Change(s) |
|---|---|

| | |
|---|---|
| Synopsis | Under the Endpoints section, the following sentence was added:<br>The data from Study 1 and Study 2 in Part I will be pooled for analysis.<br>In the Overview of Study Design section, the fourth paragraph was revised to read:<br>An interim analysis is planned in Part I when 100 randomized Part I subjects (Study 1: Bio-IR and Study 2: Bio-NF) have completed their Week 12 visit or have terminated their study participation before Week 12. If acceptable safety and efficacy are established in the combined Bio-IR and Bio-NF populations, Part II of the protocol, which consists of a dose-ranging study in subjects who are Bio-IR or Bio-NF, will be initiated. The interim analysis will allow for timely evaluation of safety and efficacy in Part I and uninterrupted patient enrollment through a seamless transition from Part I to Part II of the trial. Enrollment for Part I will continue until the sponsor makes a decision on whether to start Part II based on the interim analysis or when a maximum of 170 subjects have been randomized, whichever occurs first. Part II will be initiated if results from Part I demonstrate acceptable safety and efficacy, either at the interim analysis or when all Part I subjects have completed their Week 12 visit (or<br>have terminated study participation prior to Week 12). If Part II is not initiated based on the interim analysis results, then the results through Week 12 for all subjects in Part I (ie, when all randomized subjects in Part I have either completed the Week 12 visit or terminated study participation prior to Week 12) will be examined to determine whether to start Part II. Under this scenario, a pause in enrollment between Part I and Part II will occur.<br>In the Dosage and Administration Part I section, the first paragraph was revised to read:<br>In Part I, a minimum of 120 subjects and a maximum of 170 subjects (Bio-IR and Bio-NF) will be randomly assigned to receive placebo or the JNJ-64304500 high dose in a 1:1 ratio using permuted block randomization, stratified by baseline CDAI score (≤300 or >300) and SNP-positive status (yes or no). The Bio-IR and Bio-NF populations will be randomized separately. |
| 2.1.2. Endpoints | The following sentence was added:<br>The data from Study 1 and Study 2 in Part I will be pooled for analysis. |
| 3.1. Overview of Study Design | The fourth paragraph was revised to read:<br>The 2 studies in Part I serve to build on the original Phase 2a study findings by employing dedicated populations of both Bio-IR and Bio-NF subjects. If acceptable safety and efficacy are established in the combined Bio-IR and Bio-NF populations, Part II of the protocol, which consists of a dose-ranging study (Study 3) in subjects who are Bio-IR or Bio-NF, will be initiated.<br>The following sentence was removed from the tenth paragraph:<br>Each of the 3 studies will be analyzed separately. |
| 3.1.1. Part I | This section was revised to read:<br>In Part I, a minimum of 120 subjects and a maximum of 170 subjects (Bio-IR and Bio-NF) will be randomly assigned to receive placebo or the JNJ-64304500 high dose in a 1:1 ratio using permuted block randomization, stratified by baseline CDAI score (≤300 or >300) and SNP-positive status (yes or no). Separate randomizations will be used for the Bio-IR and Bio-NF populations.<br>The treatment groups for each study in Part I will be as follows:<br>• Placebo SC at Weeks 0, 2, 4, 6, 8, and 10; from Week 12, these subjects will receive |

additional doses as follows:

- Placebo-treated subjects who <u>are</u> in clinical response at Week 12 (≥100-point reduction from baseline in CDAI or CDAI <150) will continue to receive placebo SC injections q2w from Week 12 through Week 22.
- Placebo-treated subjects who <u>are not</u> in clinical response at Week 12 will receive JNJ-64304500 400 mg SC at Week 12 and then 200 mg SC q2w from Week 14 through Week 22.

- JNJ-64304500 400 mg SC at Week 0 then 200 mg SC q2w through Week 22.

An interim analysis is planned in Part I when 100 randomized subjects (Bio-IR and Bio-NF) have completed their Week 12 visit or have terminated their study participation before Week 12. The interim analysis will allow for uninterrupted patient enrollment through a seamless transition from Part I to Part II of the trial.

To allow for uninterrupted patient enrollment through a seamless transition from Part I to Part II of the trial, enrollment for Part I will continue until the sponsor makes a decision on whether to start Part II based on the interim analysis or when a maximum of 170 subjects have been randomized, whichever occurs first. If Part II is not initiated based on the interim analysis results, then the results through Week 12 for all subjects in Part I (ie, when all randomized subjects in Part I have either completed the Week 12 visit or terminated study participation prior to Week 12) will be examined to determine whether to start Part II. Under this scenario, a pause in enrollment between Part I and Part II will occur.

**3.1.3. Interim Analysis**

The entire section was revised to read:

An interim analysis is planned in Part I when 100 randomized Part I subjects (Study 1: Bio-IR and Study 2: Bio-NF) have completed their Week 12 visit or have terminated their study participation before Week 12.

This interim analysis will allow for uninterrupted patient enrollment through a seamless transition from Part I to Part II of the trial. As this interim analysis does not affect the conduct or completion of Part I it will be considered administrative and will not require multiplicity adjustment for the final Part I analysis.

A sponsor committee independent of the study team will be established to review the interim data and formulate recommended decisions/actions in accordance with predefined decision rules (to be defined in the Interim Analysis Plan).

An interim analysis is not planned for Part II.

Other planned DBLs are described in Section 5.2.

**3.2. Study Design Rationale**

The first paragraph was revised to read:

This protocol is comprised of 2 parts (Part I [Study 1: Bio-IR subjects and Study 2: Bio-NF subjects] and Part II [Bio-IR and Bio-NF subjects]) that are designed to evaluate the safety and efficacy of JNJ-64304500 in subjects with moderately to severely active Crohn's disease. Study 1 and Study 2 constitute Part I of the protocol. In this part, the safety and efficacy of a single dosing regimen of JNJ-64304500 in Bio-IR and Bio-NF subjects with moderately to severely active Crohn's disease is evaluated. If acceptable efficacy is established in Part I (for the combined Bio-IR and Bio-NF subjects), Part II (a dose-ranging study in Bio-IR and Bio-NF subjects) will be initiated.

| | |
|---|---|
| 3.2.1. Study Population | The entire section was removed:<br>The target population for each of the 3 studies consists of men or women ≥18 years of age at the time of informed consent with moderately to severely active Crohn's disease (of at least 3 months' duration), defined as a CDAI score ≥220 and ≤450, with elevated CRP >0.3 mg/dL (>3.0 mg/L) and/or calprotectin >250 mg/kg.<br><br>Additionally, subjects in these studies must have previously failed or been intolerant to 1 or more approved biologic agents (ie, TNFα-antagonists or vedolizumab) **or** have demonstrated an inadequate response to or failed to tolerate corticosteroids or immunomodulators (ie, 6-mercaptopurine [6-MP], azathioprine [AZA], and MTX) but not a biologic agent.<br><br>The Bio-IR population, comprising subjects who have failed to respond, lost response, or have been intolerant to one or more biologic therapies, is the primary population of interest for this protocol because it has the highest unmet need with current therapies. Responses to therapies are generally lower in the Bio-IR population than in the Bio-NF population.<br><br>The cohort of Bio-NF subjects is included in Part I (Study 2) to obtain additional information about the effect of JNJ-64304500 in this population early in the development program. |
| 3.2.3. Efficacy Assessments | The last sentence of the third paragraph was removed:<br>The clinical remission endpoint is being used for the interim analysis, however, as it is a more stringent endpoint and provides a more conservative decision rule to determine whether to start Part II early. |
| 11.10 Interim Analysis | The entire section was revised to read:<br>An interim analysis is planned in Part I when 100 randomized subjects (Study 1: Bio-IR and Study 2: Bio-NF) have completed their Week 12 visit or have terminated their study participation before Week 12. This interim analysis will allow for uninterrupted patient enrollment through a seamless transition from Part I to Part II of the trial. As this interim analysis does not affect the conduct or completion of Part I, it will be considered administrative and will not require multiplicity adjustment for the final Part I analysis.<br><br>The primary efficacy evaluation is the comparison between JNJ-64304500 and placebo with respect to the change in CDAI from baseline (in the combined Bio-IR and Bio-NF subjects). Other selected efficacy analyses (eg, clinical remission, clinical response, and change in SES-CD) will also been performed; details will be provided in the Interim Analysis Plan.<br><br>A sponsor committee independent of the study team will be established to review the interim data and formulate recommended decisions/actions in accordance with predefined decision rules that will be defined in the Interim Analysis Plan. |

**Rationale (change the overall sample size of Part I):** The Part I analysis is now focused on all subjects (Bio-IR vs. Bio-NF) due to the faster than anticipated enrollment of the Bio-NF population. This analysis may be more reflective of the patient population in need of therapy. The overall sample size of Part I was changed to a minimum of 120 subjects and a maximum of 170 subjects (Bio-IR and Bio-NF).

| Applicable Section(s) | Description of Change(s) |
|---|---|
| Synopsis | The Overview of Study Design section was revised to read: |

This protocol is comprised of 2 parts (Part I [Study 1: Bio-IR subjects and Study 2: Bio-NF subjects] and Part II [Bio-IR and Bio-NF subjects]). Each study is a randomized, double-blind, placebo-controlled, parallel-group, multicenter study of JNJ-64304500 in subjects with moderately to severely active Crohn's disease. A minimum of 120 subjects and a maximum of 170 subjects (Bio-IR and Bio-NF) will be randomized in Part I. An additional 275 subjects will be randomized in Part II. Part I will study the safety and efficacy of a high dose regimen of JNJ-64304500 compared with placebo in subjects who have failed biologic or conventional therapy (Bio-IR or Bio-NF subjects, respectively). Part II will study the safety and efficacy of multiple-dose regimens of JNJ-64304500 compared with placebo, with ustekinumab (STELARA®) as a reference arm, in Bio-IR and Bio-NF subjects.

The duration of the study will be 38 weeks in Part I and 36 weeks in Part II for a subject who completes all scheduled visits, including study agent administration visits and a final efficacy and safety follow-up visit. Eligible subjects will only participate in Part I <u>or</u> Part II of the study.

The Time and Events Schedules summarize the frequency and timing of assessments for efficacy, PK, biomarkers, immunogenicity, PD, and safety in Part I and Part II of the study.

An interim analysis is planned in Part I when 100 randomized Part I subjects (Study 1: Bio-IR and Study 2: Bio-NF) have completed their Week 12 visit or have terminated their study participation before Week 12. If acceptable safety and efficacy are established in the combined Bio-IR and Bio-NF populations, Part II of the protocol, which consists of a dose-ranging study in subjects who are Bio-IR or Bio-NF, will be initiated. The interim analysis will allow for timely evaluation of safety and efficacy in Part I and uninterrupted patient enrollment through a seamless transition from Part I to Part II of the trial. Enrollment for Part I will continue until the sponsor makes a decision on whether to start Part II based on the interim analysis or when a maximum of 170 subjects have been randomized, whichever occurs first. Part II will be initiated if results from Part I demonstrate acceptable safety and efficacy, either at the interim analysis or when all Part I subjects have completed their Week 12 visit (or have terminated study participation prior to Week 12). If Part II is not initiated based on the interim analysis results, then the results through Week 12 for all subjects in Part I (ie, when all randomized subjects in Part I have either completed the Week 12 visit or terminated study participation prior to Week 12) will be examined to determine whether to start Part II. Under this scenario, a pause in enrollment between Part I and Part II will occur.

An external Data Monitoring Committee (DMC) will review unblinded safety data from all subjects periodically to monitor subject safety.

The end of the 64304500CRD2001 study is defined as the date on which the last subject completes the last efficacy and safety follow-up visit.

In the Sample Size Determination section, the first and second paragraphs were revised to read:

Sample size calculations for Part I (Bio-IR subjects and Bio-NF subjects combined) were determined by the power to detect a significant difference in the change from baseline in the CDAI score at Week 8 between JNJ-64304500 and placebo using a 2-sample t-test. Sample size calculation for Part II (Bio-IR and Bio-NF subjects) was determined by the power to detect a dose-response signal in the change from baseline in the CDAI score at Week 8 (primary endpoint in Part II) using the Multiple Comparison Procedures with modeling techniques (MCP-Mod) method.

For Part I, assuming the mean CDAI change from baseline at Week 8 is -98 in the JNJ-64304500 group and -46 in the placebo group with a common SD of 96, 60 subjects per treatment group will provide 84% power to detect a treatment difference between JNJ-

64304500 and placebo at an overall Type 1 error of 0.05.

In the Efficacy Analyses section, the first 3 paragraphs were revised to read:

This protocol is comprised of 2 separate parts (Part I [Study 1: Bio-IR subjects and Study 2: Bio-NF subjects] and Part II [Bio-IR and Bio-NF subjects]). Part I and Part II will be analyzed separately with separate Type I error control for the primary endpoint (at the 0.05 level of significance). The other endpoints within each part will not be controlled for multiplicity.

For each part, the analysis set is all randomized subjects who received study agent. Efficacy analyses will be based on a modified intent-to-treat principle. Therefore, the efficacy data for each subject who received study agent will be analyzed according to the assigned treatment regardless of the actual treatment received.

For Part I, the primary endpoint of change from baseline in the CDAI score at Week 8 will be compared between the JNJ-64304500 treatment group and the placebo treatment group by an analysis of covariance model on the van der Waerden normal scores with treatment as a fixed factor and baseline CDAI score, SNP-positive status (yes or no), and Bio-IR status (yes or no) as covariates. Part I will be considered to be positive if a significant improvement is detected in the change from baseline in the CDAI score at Week 8 in the JNJ-64304500 group compared with the placebo group at the 0.05 level of significance.

3.1. Overview of Study Design

The seventh paragraph was revised to read:

It is anticipated that approximately 395-445 subjects will be randomized overall across Part I and Part II:

- **Part I** will study the safety and efficacy of a high dose regimen of JNJ-64304500 compared with placebo and will enroll a minimum of 120 subjects and a maximum of 170 subjects (Bio-IR and Bio-NF).

3.1.1. Part I

The first paragraph was revised to read:

In Part I, a minimum of 120 subjects and a maximum of 170 subjects (Bio-IR and Bio-NF) will be randomly assigned to receive placebo or the JNJ-64304500 high dose in a 1:1 ratio using permuted block randomization, stratified by baseline CDAI score (≤300 or >300) and SNP-positive status (yes or no). Separate randomizations will be used for the Bio-IR and Bio-NF populations.

11.1. Sample Size Determination

The first paragraph was revised to read:

Sample size calculations for Part I and Part II were determined by the power to detect a significant difference in the change from baseline in the CDAI score at Week 8 (primary endpoint in each part) between JNJ-64304500 and placebo using a 2-sample t-test.

11.1.1. Sample Size in Part I (Study 1 and Study 2)

The section header was updated to Sample Size in Part I.

Subheaders 11.1.1.1. Bio-IR Subjects (Study 1), 11.1.1.1.1. All Bio-IR Subjects, 11.1.1.1.2 Bio-IR Subjects Who Are SNP-Positive (Bio-IR/SNP+), 11.1.1.1. Bio-NF Subjects (Study 2), 11.1.1.2.1. All Bio-NF Subjects, and 11.1.1.2.2. Bio-NF Subjects Who Are SNP-Positive (Bio-NF/SNP+) were removed.

The entire section was revised to read:

Part I is powered based on the combined data from Study 1 and Study 2. Study 1 and Study 2 were not individually powered to detect differences between JNJ-64304500 and placebo.

The assumptions for the sample size calculations in Part I were based on data from

| | |
|---|---|
| | CNTO1275CRD3001 (a study conducted by the sponsor in subjects with Crohn's disease who had failed or were intolerant to TNF-antagonist therapy) and CNTO1275CRD3002 (a study conducted by the sponsor in subjects with Crohn's disease who had failed or were intolerant to corticosteroids or immunomodulators but who had not failed TNF-antagonist therapy). In CNTO1275CRD3001, the mean CDAI change from baseline at Week 8 was -25.1 (SD=91.41) and -78.7 (SD=91.79) for the placebo and ustekinumab 6 mg/kg groups, respectively. These assumptions incorporated the impact of 6% of subjects being noncompleters (in CNTO1275CRD3001). In CNTO1275CRD3002, the mean CDAI change from baseline at Week 8 was -66.3 (SD=97.81) and -116.3 (SD=102.88) for the placebo and ustekinumab 6 mg/kg groups, respectively. These assumptions incorporated the impact of 4% of subjects being noncompleters (in CNTO1275CRD3002).<br><br>For Part I, assuming the mean CDAI change from baseline at Week 8 is -98 in the JNJ-64304500 group and -46 in the placebo group with a common SD of 96 (these values are derived assuming 1:1 ratio of Bio-IR and Bio-NF subjects) 60 subjects per treatment group (a total of 120 subjects) will provide 84% power to detect a treatment difference between JNJ-64304500 and placebo at an overall Type 1 error of 0.05 (2-sided).<br><br>Table 4 was updated. |
| Section 11.2. Efficacy Analyses | This section was revised to read:<br><br>This protocol is comprised of 2 separate parts (Part I [Study 1: Bio-IR subjects and Study 2: Bio-NF subjects] and Part II [Bio-IR and Bio-NF subjects]). Part I and Part II will be analyzed separately with separate Type I error control for the primary endpoint (at the 0.05 level of significance). The other endpoints within each part will not be controlled for multiplicity.<br><br>For each part, the analysis set is all randomized subjects who received study agent. Efficacy analyses will be based on a modified intent-to-treat principle. Therefore, the efficacy data for each subject who received study agent will be analyzed according to the assigned treatment regardless of the actual treatment received. |
| 11.2.1. Part I (Study 1 and Study 2) | Subheader title updated to Part I |
| 11.2.1.1 Primary Endpoint Analysis | Entire section revised to read:<br><br>The primary endpoint for Part I is the change from baseline in the CDAI score at Week 8.<br><br>The change from baseline in the CDAI score will be compared between the JNJ-64304500 treatment group and the placebo treatment group for all subjects in Part I. For the comparison, an ANCOVA model on the van der Waerden normal scores will be used with treatment as a fixed factor and baseline CDAI score, SNP-positive status (yes or no), and Bio-IR status (yes or no) as covariates. For this analysis, treatment failure rules and missing data rules as specified in Section 11.2.2.1 will be applied.<br><br>Part I will be considered to be positive if a significant improvement is detected in the change from baseline in the CDAI score at Week 8 in the JNJ-64304500 group compared with the placebo group at the 0.05 level of significance. |
| 11.2.1.2 Other Efficacy Endpoint Analyses | First sentence revised to read:<br><br>The following endpoints will be compared between the JNJ-64304500 treatment group and the placebo treatment group for all subjects in Part I and by Bio-IR status (yes or no): |

| | |
|---|---|
| 11.2.2. Study 2 (Part I Bio-NF Subjects) | This section was removed. |

**Rationale (change the subject population in Part II):** To evaluate the efficacy and safety of JNJ-64304500 in a broader population, the subject population in Part II was changed to include both Bio-IR and Bio-NF subjects.

| Applicable Section(s) | Description of Change(s) |
|---|---|
| Synopsis | In the first section, the bullet regarding Part II, Study 3 was revised to read:<br>Study 3: A dose-ranging study in subjects who are Bio-IR or Bio-NF<br>In the Dosage and Administration Part II section, the first paragraph was revised to read:<br>In Part II of the study, subjects (Bio-IR and Bio-NF) will be randomly assigned in equal proportions to receive placebo, 1 of 3 dose regimens of JNJ-64304500, or ustekinumab, using permuted block randomization, stratified by baseline CDAI score (≤300 or >300), SNP-positive status (yes or no), and Bio-IR status (yes or no). The treatment groups are as follows:<br>In the Efficacy Analyses section, the fourth and fifth paragraphs were revised to read:<br>The primary endpoint for Part II is also the change from baseline in the CDAI score at Week 8. A unified strategy that combines Multiple Comparison Procedures with modeling techniques, MCP-Mod, will be used to analyze the dose-response data over the JNJ-64304500 doses. This approach consists of 2 major steps. The first step consists of testing the dose-response signal via multiple contrast tests while controlling the overall Type 1 error. If a dose-response signal is detected, the second step is to select a model that best describes the observed data and use it to estimate adequate doses with associated precision. The study will be considered positive if a dose-response signal for the primary endpoint is detected. In addition to the dose-response analysis, pairwise comparisons of the JNJ-64304500 treatment groups versus the placebo group will be performed for the change from baseline in the CDAI score at Week 8; these comparisons will not be adjusted for multiplicity.<br>The major secondary endpoints are only specified in Part II. The major secondary endpoints that are dichotomous endpoints will be analyzed by the Cochran-Mantel-Haenszel chi-square test (2-sided) stratified by baseline CDAI score (≤300 or >300), SNP-positive status (yes or no), and Bio-IR status (yes or no); the major secondary endpoints that are continuous endpoints will be analyzed by an analysis of covariance model on the van der Waerden normal scores with treatment as a fixed factor and appropriate baseline level of the variable, SNP-positive status (yes or no), and Bio-IR status (yes or no) as covariates. The major secondary endpoints will not be controlled for multiplicity. |
| 1.2.2. Crohn's Disease | The eighth paragraph was revised to read:<br>In Part II, the following study will be conducted:<br>• Study 3: A dose-ranging study in subjects who are biologic intolerant or refractory (Bio-IR) or Biologic nonfailure (Bio-NF)<br>The following paragraph was removed from this section:<br>In order to provide an efficient means of rapidly moving to formal Phase 2 dose-ranging, this protocol is designed to be a comprehensive Phase 2 program in which the sponsor will evaluate safety and efficacy in subjects who are biologic intolerant or refractory (ie, Bio-IR population or Bio-NF). |

| | |
|---|---|
| | The ninth paragraph was revised to read:<br><br>In Study 1, a higher dose regimen than was used in the Phase 2a study will be employed supported by the acceptable safety margins demonstrated in Phase 1 (the Novo Nordisk Phase 1 single ascending/multiple ascending dose study NN8555-3618), and the high exposure margins relative to NOAEL exposure in cynomolgus monkeys. This higher dose will be used to provide additional information on safety and efficacy for the Bio-IR population, as proof of concept was not established in this population in the Novo Nordisk Phase 2a study (NN8555-3797). In addition, this higher dose will also be evaluated in Bio-NF subjects to determine if a more robust efficacy outcome than was shown in the Phase 2a study can be achieved while maintaining an acceptable safety profile. Combined with data from Phase 2a in Bio-NF subjects, Part I and Part II would provide valuable information for the assessment of dose/exposure-response and thus the selection of the optimal dose in Bio-NF population for the Phase 3 program. Study 1 and Study 2 comprise Part I of the protocol and these 2 components will be initiated in parallel. The decision to initiate the dose-ranging portion of the protocol (Part II, Bio-IR and Bio-NF) is dependent on demonstration of acceptable safety and efficacy in Part I (combined Bio-IR subjects and Bio-NF subjects). A ustekinumab reference arm will be included in Part II to provide context in the interpretation of results from the JNJ-64304500 and placebo groups and to assist in the overall selection of dose(s) to carry forward into the Phase 3 program. |
| 3.1. Overview of Study Design | The first bullet in the third paragraph was revised to read:<br><br>• Study 3: A dose-ranging study in subjects who are Bio-IR or Bio-NF<br><br>Figure 1 was revised to include Bio-IR and Bio-NF subjects in Part II.<br><br>The 2nd bullet in the sixth paragraph was revised to read:<br><br>• **Biologic nonfailure (Bio-NF) subjects** are defined as those who have demonstrated an inadequate response to or have failed to tolerate corticosteroids or the immunomodulators 6-MP, AZA, or MTX (Attachment 2). Subjects who have demonstrated corticosteroid dependence (ie, an inability to successfully taper corticosteroids without a return of the symptoms of Crohn's disease) are also eligible. Bio-NF subjects may also have received biologic therapy but only if it was discontinued for reasons other than lack of efficacy or intolerance (eg, drug holiday).<br><br>The second bullet in the seventh paragraph was revised to read:<br><br>• **Part II** will study the safety and efficacy of multiple-dose regimens of JNJ-64304500 compared with placebo, with ustekinumab (STELARA®) as a reference arm. Part II will enroll approximately 275 subjects (Bio-NF subjects will be no more than 50% and Bio-IR subjects will be no more than 60%). |
| 3.1.2. Part II | The first paragraph was revised to read:<br><br>In Part II, 275 additional Bio-IR or Bio-NF subjects will be randomly assigned to receive placebo or 1 of 3 dose levels of JNJ-64304500 or ustekinumab in a ratio of 1:1:1:1:1 using permuted block randomization, stratified by baseline CDAI score (≤300 or >300), SNP-positive status (yes or no), and BIO-IR status (yes or no). Bio-NF subjects will be no more than 50% and Bio-IR subjects will be no more than 60%. |
| 4.1. Inclusion Criteria | Inclusion Criterion 6 was revised to read:<br><br>6. **In Part II,** meet the following requirement for prior or current medications for Crohn's disease:<br><br>a. Has previously demonstrated inadequate response, loss of response, or intolerance to 1 or more approved biologic therapies (eg, infliximab, adalimumab, certolizumab pegol, or vedolizumab) as outlined in Attachment 1. |

| | |
|---|---|
| | **OR**<br>b. Has failed conventional therapy:<br>1) Is currently receiving corticosteroids and/or immunomodulators (ie, AZA, 6-MP, or MTX) at adequate therapeutic doses (Attachment 2).<br>**OR**<br>2) Has a history of failure to respond to or tolerate an adequate course of corticosteroids and/or immunomodulators (ie, AZA, 6-MP, or MTX) (Attachment 2).<br>**OR**<br>3) Is corticosteroid dependent or has had a history of corticosteroid dependency (Attachment 2). |
| 5.1. Treatment Allocation | The first paragraph was revised to read:<br>Central randomization will be implemented in this study. Subjects will be randomly assigned to 1 of 2 treatment groups (1:1 ratio) in Part I and to 1 of 5 treatment groups (1:1:1:1:1 ratio) in Part II, based on a computer-generated randomization schedule prepared before the study by or under the supervision of the sponsor. Each of the 3 studies will have separate randomizations. Each randomization will be balanced by using randomly permuted blocks and will be stratified by baseline CDAI score (≤300 or >300) and SNP-positive status (yes or no; Part II will also be stratified by Bio-IR status [yes or no]). The interactive web response system (IWRS) will assign a unique treatment code, which will dictate the treatment assignment and matching study drug kit for the subject. The requestor must use his or her own user identification and personal identification number when contacting the IWRS, and will then give the relevant subject details to uniquely identify the subject. |
| 11.2.2. Study 3 (Part II) | Subheader title revised to Part II. |

**Rationale (change the sample size in Part II):** The sample size in Part II was increased to 275 subjects in order to have sufficient power based on the amended Part II population.

| Applicable Section(s) | Description of Change(s) |
|---|---|
| Synopsis | In the Sample Size Determination section, the entire section was revised to read:<br>Sample size calculations for Part I (Bio-IR subjects and Bio-NF subjects combined) were determined by the power to detect a significant difference in the change from baseline in the CDAI score at Week 8 between JNJ-64304500 and placebo using a 2-sample t-test. Sample size calculation for Part II (Bio-IR and Bio-NF subjects) was determined by the power to detect a dose-response signal in the change from baseline in the CDAI score at Week 8 (primary endpoint in Part II ) using the Multiple Comparison Procedures with modeling techniques (MCP-Mod) method.<br>For Part I, assuming the mean CDAI change from baseline at Week 8 is -98 in the JNJ-64304500 group and -46 in the placebo group with a common SD of 96, 60 subjects per treatment group will provide 84% power to detect a treatment difference between JNJ-64304500 and placebo at an overall Type 1 error of 0.05.<br>For Part II, assuming the mean CDAI change from baseline at Week 8 is -98 in the JNJ-64304500 group and -46 in the placebo group with a common SD of 96, 55 subjects per treatment group will provide a mean power of 83% to detect a dose-response signal based on 7 candidate dose-response models at an overall Type 1 error of 0.05. |

| | |
|---|---|
| 3.1.2. Part II | The first paragraph was revised to read:<br>In Part II, 275 additional Bio-IR or Bio-NF subjects will be randomly assigned to receive placebo or 1 of 3 dose levels of JNJ-64304500 or ustekinumab in a ratio of 1:1:1:1:1 using permuted block randomization, stratified by baseline CDAI score (≤300 or >300), SNP-positive status (yes or no), and BIO-IR status (yes or no). Bio-NF subjects will be no more than 50% and Bio-IR subjects will be no more than 60%. |
| 11.1.2. Sample Size in Part II (Study 3) | Subheader title changed to Sample Size in Part II |
| 11.1.2.1 All Bio-IR Subjects | This subheader title was removed. The content for this section is now included in section 11.1.2. Sample Size in Part II. |
| 11.1.2.2. Bio-IR Subjects Who Are SNP-Positive(Bio-IR/SNP+) | This section was removed. |

**Rationale:** Contents for blinding and planned database locks were updated based on the changes to the study design.

| Applicable Section(s) | Description of Change(s) |
|---|---|
| 5.2. Blinding | The second and third paragraphs were revised to read:<br>• **Interim analysis lock for Part I:** Occurs when 100 randomized subjects (Bio-IR and Bio-NF) in Part I have completed their Week 12 visit or have terminated their study participation before Week 12.<br>• **Week 12 DBL for Part I:** Occurs when all subjects in Part I have completed their Week 12 visit or have terminated their study participation before Week 12.<br>• **Week 12 DBL for Part II (final DBL for Part I occurs at the same time):** Occurs when all Part II subjects have completed their Week 12 visit or have terminated their study participation before Week 12.<br>• **Week 24 DBL for Part II:** Occurs when all Part II subjects have completed their Week 24 visit or have terminated their study participation before Week 24.<br>• **Final DBL for Part II:** Occurs when all Part II subjects have completed their final efficacy and safety visit or have terminated their study participation before the final efficacy and safety visit.<br>At the time of the interim analysis lock for Part I, a limited number of sponsor personnel will become unblinded to treatment assignment. At the time of the Week 12 DBL for Part I and the Week 12 DBL for Part II, the sponsor, except for site monitors (who have interactions with the investigative sites), will become unblinded to treatment assignment. Identification of sponsor personnel who will have access to subject-level data for the interim analysis lock for Part I will be documented before the unblinding. The study blind will be maintained for investigators, site personnel, subjects, and sponsor site monitors until the final analyses have been completed for all subjects in Part I and Part II. This measure will mitigate the potential bias in the remaining investigator and subject assessments. |

**Rationale:** Minor wording changes to the numerical rating scale (NRS) content based on feedback from a Health Authority.

| Applicable Section(s) | Description of Change(s) |
|---|---|
| 9.2.3. Abdominal Pain Numerical Rating Scale | This section was updated to read:<br>The NRS for pain is a unidimensional measure of pain intensity in adults.[5] An 11-point (0-10) NRS will be used to evaluate abdominal pain. The score of 0 represents "no abdominal pain" |

| | |
|---|---|
| | and the score of 10 represents the "worst possible abdominal pain", with greater scores indicating greater pain severity and intensity. Subjects will select only one number that best reflects their pain at its worst in the past 24 hours. The abdominal pain NRS will be assessed daily. Subjects will complete the NRS at approximately the same time each day before going to bed and bring the diary to each visit. |

**Rationale:** Minor wording changes to Patient's Global Impression of Severity (PGIS) content based on feedback from a Health Authority.

| Applicable Section(s) | Description of Change(s) |
|---|---|
| 9.2.4. Patient's Global Impression of Severity of Crohn's Disease | This section was updated to read:<br>The PGIS of Crohn's disease is a 5-point scale ("None", "Mild", "Moderate", "Severe" and "Very Severe") to rate Crohn's disease intensity. Subjects will rate their PGIS of Crohn's disease at Weeks 0, 4, 8, 12, and 24. |

**Rationale:** Minor edits were noted.

| Applicable Section(s) | Description of Change(s) |
|---|---|
| Throughout the protocol | Minor grammatical, formatting, or spelling changes were made. |

**Amendment 2** (09 November 2016)

This amendment is considered to be substantial based on the criteria set forth in Article 10(a) of Directive 2001/20/EC of the European Parliament and the Council of the European Union.

**The overall reason for the amendment:** The overall reason for the amendment was to address health authority requests.

Clarification of specific study procedures is also included in this amendment, as described below.

| Applicable Sections | Description of Changes |
|---|---|
| **Rationale:** The 12-week follow-up period after the last administration of study agent was changed to a 16-week follow-up period in both Part I and Part II. Requirements for the duration of contraception use and the duration of avoidance of live vaccinations were also changed to 16 weeks after the last administration of study agent.<br>Due to the addition of 4 weeks to the follow-up period after the last study agent administration in both Part I and Part II of the study, the study duration was increased by 4 weeks in both Part I and Part II. | |
| Synopsis | In the Overview of Study Design section, the second paragraph was revised to read:<br>The duration of the study will be 38 weeks in Part I and 36 weeks in Part II for a subject who completes all scheduled visits, including study agent administration visits and a final efficacy and safety follow-up visit. Eligible subjects will only participate in Part I or Part II of the study. |

| | |
|---|---|
| Time and Events Schedule: Part I | Footnote c was revised to read:<br><br>c. Subjects who <u>terminate study participation</u> should complete an early termination visit. If a subject completed assessments at Week 24 and terminated after Week 24, the only assessments that should be performed at the early termination visit are those planned for the final efficacy and safety follow-up. Subjects who <u>discontinue study agent administration</u> before Week 8 (but have not terminated study participation) should complete the Week 8 and Week 12 assessments and return for a final efficacy and safety follow-up visit approximately 16 weeks after their last study agent administration. Subjects who <u>discontinue study agent administration</u> after Week 8 (but have not terminated study participation) should complete the Week 12 and Week 24 assessments and return for a final efficacy and safety follow-up visit approximately 16 weeks after their last study agent administration.<br><br>Footnote d was revised to read:<br><br>d. Subjects should complete a final efficacy and safety follow-up visit approximately 16 weeks after their last study agent administration. For subjects who complete all visits, this will occur at Week 38. |
| Time and Events Schedule: Part II | Footnote d was revised to read:<br><br>d. Subjects should complete a final efficacy and safety follow-up visit approximately 16 weeks after their last study agent administration. For subjects who complete all visits, this will occur at Week 36. |
| 3.1 Overview of Study Design | Figure 2 was updated to indicate Week 38 as the final efficacy and safety visit in Part I, and Figure 3 was updated to indicate Week 36 as the final efficacy and safety visit in Part II. |
| 3.2.9.1. JNJ-64304500 | The following paragraph was added after the second paragraph:<br><br>A 16-week follow-up interval after the last administration of study agent has been employed based on the observed safety profile and PK profile of JNJ-64304500 in the completed clinical studies in RA and Crohn's disease subjects. The average serum concentration of JNJ-64304500 at 16 weeks after the last administration of study agent is predicted to be approximately 0.01 µg/mL, which is considered unlikely to produce drug-related effects or safety concerns. |
| 4.1 Inclusion Criteria | Inclusion Criterion 13.b.2 was revised to read:<br><br>Agrees to remain on a highly effective method of contraception throughout the study and for at least 16 weeks after the last administration of study agent.<br><br>Inclusion Criterion 14 was revised to read:<br><br>A woman must agree not to donate eggs (ova, oocytes) for the purposes of assisted reproduction during the study and for a period of 16 weeks after the last administration of study agent.<br><br>The first part Inclusion Criterion 15 was revised to read:<br><br>During the study and for at least 16 weeks after the last administration of study agent, a man |
| 4.2 Exclusion Criteria | Exclusion Criterion 28 was revised to read:<br><br>Is a woman who is pregnant, or breast-feeding, or planning to become pregnant or is a man who plans to father a child while enrolled in this study or within 16 weeks after the last administration of study agent. |

| | |
|---|---|
| 4.3. Prohibitions and Restrictions | Restriction 3 was revised to read:<br>Subjects must agree not to receive a live virus or live bacterial vaccination, including a BCG vaccination, during the study and for 12 months after receiving the last dose of study agent for BCG vaccination or 16 weeks for other live vaccines. |
| 9.1.2 Screening Period | The fourth paragraph was revised to read:<br>Women of childbearing potential must have a negative serum β-hCG pregnancy test result at screening. Subjects must be reminded that they are required to use a highly effective method of contraception during the study (as described in Inclusion Criterion 13) and must continue taking such precautions for 16 weeks after receiving the last administration of study agent. The method(s) of contraception used by each subject must be documented. |
| 9.1.4. Final Efficacy and Safety Follow-up Visit | This section was revised to read:<br>Subjects in Part I and Part II should complete the final efficacy and safety follow-up visit approximately 16 weeks after the last study agent administration (Table 1, Table 2). |
| 10.1. Completion | The first paragraph was revised to read:<br>A subject will be considered to have completed the study if he or she has completed assessments through the final efficacy and safety visit (ie, Week 38 in Part I and Week 36 in Part II), as specified in the Time and Events Schedules for Part I or Part II.<br>The third paragraph was revised to read:<br>The final efficacy and safety follow-up visit for subjects who discontinue study agent is approximately 16 weeks after the last administration of study agent in both Part I and Part II. |
| 10.2. Discontinuation of Study Treatment | The second bullet of the second paragraph was revised to read:<br>The subject becomes pregnant or plans a pregnancy within the study period or within 16 weeks after the last study agent administration. |
| 12.3.1. All Adverse Events | The first paragraph was revised to read:<br>All AEs and special reporting situations, whether serious or nonserious, will be reported from the time a signed and dated ICF is obtained until completion of the subject's last study-related procedure, which may include contact for follow-up of safety. Serious adverse events, including those spontaneously reported to the investigator within 16 weeks after the last dose of study drug, must be reported using the Serious Adverse Event Form. The sponsor will evaluate any safety information that is spontaneously reported by an investigator beyond the time frame specified in this protocol. |

**Rationale:** In response to a Health Authority request, the Patient’s Global Impression of Severity (PGIS) of Crohn’s disease and the Patient’s Global Impression of Change (PGIC) of severity of Crohn’s disease were added as efficacy assessments in Part II of the study.

| | |
|---|---|
| Synopsis | In the Efficacy Evaluations section, the following were added:<br>• Patient’s Global Impression of Severity (PGIS) of Crohn’s disease<br>• Patient’s Global Impression of Change (PGIC) of severity of Crohn’s disease |
| Time and Events Schedule: Part II | The following efficacy assessments were added:<br>PGIS of Crohn’s disease assessment at Weeks 0, 4, 8, 12, 24, and Early termination.<br>PGIC of severity of Crohn’s disease assessment at Weeks 4, 8, 12, 24, and Early termination. |

| | |
|---|---|
| 2.1.2. Endpoints | The third paragraph was revised to read:<br>The following efficacy endpoints will be evaluated in each of the 3 studies (except for the Patient's Global Impression of Severity [PGIS] of Crohn's disease and the Patient's Global Impression of Change [PGIC] of severity of Crohn's disease, which will only be evaluated in Study 3):<br>The following were added to the list of efficacy endpoints:<br>• Change in PGIS of Crohn's disease from baseline at Weeks 4, 8, 12, and 24.<br>• A ≥1-point improvement in PGIS of Crohn's disease from baseline at Weeks 4, 8, 12, and 24.<br>• Improvement in PGIC of severity of Crohn's disease at Weeks 4, 8, 12, and 24. |
| 9.2. Efficacy Evaluations | This section was revised to read:<br>The CDAI will be the primary tool for assessing disease activity response to JNJ-64304500, along with PRO-2, PRO-3, Bristol stool form scale, abdominal pain based on NRS 0-10 scale, PGIS of Crohn's disease, and PGIC of severity of Crohn's disease. The degree of inflammation will be assessed by measuring serum CRP concentrations. Stool samples will be collected and analyzed to evaluate changes in markers that may reflect JNJ-64304500 or ustekinumab treatment. The well-being of subjects will be measured using the IBDQ and the SF-36. Mucosal healing will be assessed by ileocolonoscopy. For subjects with fistulizing disease, fistula closure will also be assessed. |
| 9.2.4 Patient's Global Impression of Severity of Crohn's Disease | In Section 9.2 Efficacy Evaluations, a new section 9.2.4 Patient's Global Impression of Severity of Crohn's Disease was added:<br>The PGIS of Crohn's disease is a 5-point scale ("Absent", "Mild", "Moderate", "Severe" and "Very Severe") to rate Crohn's disease intensity. Subjects will rate their PGIS of Crohn's disease at Weeks 0, 4, 8, 12, and 24. |
| 9.2.5 Patient's Global Impression of Change of Severity of Crohn's Disease | In Section 9.2 Efficacy Evaluations, a new section 9.2.5 Patient's Global Impression of Change of Severity of Crohn's Disease was added:<br>The PGIC of severity of Crohn's disease is a 7-point scale ranging from "a lot better now" to "a lot worse now" with a neutral center point ("neither better nor worse") to measure perceived change (improvement or deterioration) in severity of Crohn's disease. Subjects will assess their PGIC of severity of Crohn's disease at Weeks 4, 8, 12, and 24. |
| 11.2.3.3. Other Efficacy Endpoint Analyses | This section was revised to read:<br>Comparisons between each of the JNJ-64304500 treatment groups and the placebo treatment group will also be made for each of the endpoints specified in Section 11.2.1.2 and for the following endpoints.<br>• Change in PGIS of Crohn's disease from baseline at Weeks 4, 8, 12, and 24.<br>• A ≥1-point improvement in PGIS of Crohn's disease from baseline at Weeks 4, 8, 12, and 24.<br>• Improvement in PGIC of severity of Crohn's disease at Weeks 4, 8, 12, and 24.<br>Pairwise comparisons of the ustekinumab treatment group with the JNJ-64304500 treatment groups or with placebo are not planned for these endpoints, however summary statistics will be provided for the ustekinumab group. |
| 15. Study-Specific Materials | The following was added to the list of supplies provided to the investigator:<br>• Worksheets for data collection |

**Rationale:** In response to a Health Authority request, further clarification of the study design and protocol structure

| | |
|---|---|
| was added. | |
| 1.2.2. Crohn's Disease | The second paragraph was revised to read:<br><br>Subjects were evaluated for the primary endpoint of change from baseline in the CDAI score at Week 4. Safety and efficacy evaluations were performed through Week 24. The observed 16-point greater reduction in CDAI in the JNJ-64304500 group at Week 4 compared with the placebo group was not significant (p=0.403). Based on a predefined significance level of 0.10, however, the reduction in CDAI score was significantly higher in the JNJ-64304500 group compared with the placebo group at Week 12 (55-point greater reduction in CDAI was observed in JNJ-64304500 compared with placebo, p=0.056). Based on the same predefined significance level of 0.10, reductions in CDAI scores were significantly higher in the predefined subgroup of "no prior failure to biologics" (Bio-NF, 71% of the study population) in the JNJ-64304500 group compared with placebo at all post baseline visits through Week 12 (Week 1, p=0.068; Week 2, p=0.048; Week 4, p=0.095; Week 8, p=0.015; and Week 12, p=0.025). The biologic intolerant or refractory population (Bio-IR) in the Novo Nordisk Phase 2a study constituted 29% of the study population (N=12 in the active treatment arm and N=11 in the placebo arm). In this subpopulation, a 45-point greater reduction in CDAI was observed at Week 12 in JNJ-64304500 compared with placebo.<br><br>The last paragraph was replaced with the following 3 paragraphs at the end of the section:<br><br>In order to provide an efficient means of rapidly moving to formal Phase 2 dose-ranging, this protocol is designed to be a comprehensive Phase 2 program in which the sponsor will evaluate safety and efficacy in subjects who are biologic intolerant or refractory (the Bio-IR population, Study 1).<br><br>In Study 1, a higher dose regimen than was used in the Phase 2a study will be employed supported by the acceptable safety margins demonstrated in Phase 1 (the Novo Nordisk Phase 1 single ascending/multiple ascending dose study NN8555-3618), and the high exposure margins relative to NOAEL exposure in cynomolgus monkeys. This higher dose will be used to establish safety and efficacy (Proof of Concept) for the Bio-IR population, as this was not satisfactorily established in this population in the Novo Nordisk Phase 2a study (NN8555-3797). In addition, this higher dose will also be evaluated in subjects who have not previously failed a biologic therapy (Study 2, Bio-NF) to determine if a more robust efficacy outcome than was shown in the Phase 2a study can be achieved while maintaining an acceptable safety profile. Combined with data from Phase 2a in Bio-NF subjects, Study 2 would provide valuable information for the assessment of dose/exposure-response and thus the selection of the optimal dose in Bio-NF population for the Phase 3 program. Study 1 and Study 2 comprise Part I of the protocol and these 2 components will be initiated in parallel. It is anticipated that Bio-NF subjects will enroll slower than Bio-IR subjects. Therefore, initiation of Study 2 in parallel with Study 1 will facilitate the completion of Study 2 in a timely fashion to be able to inform Phase 3 study planning for the Bio-NF population. The decision to initiate the dose-ranging portion of the protocol (Study 3, Bio-IR) is dependent on demonstration of acceptable safety and efficacy in the Bio-IR subjects in Study 1. A ustekinumab reference arm will be included in Study 3 to provide context in the interpretation of results from the JNJ-64304500 and placebo groups and to assist in the overall selection of dose(s) to carry forward into the Phase 3 program.<br><br>The inclusion of the 3 component studies in a single protocol will facilitate both efficient trial conduct and program decision-making based on planned interim DBLs as described in Section 3.1.1, Section 3.1.3, and Section 5.2. |
| **Rationale:** Inclusion criteria related to prior or current medications for Crohn's disease were updated to clarify the requirements for Part I (Study 1 and Study 2) and Part II (Study 3). | |
| 4.1. Inclusion Criteria | Inclusion Criterion 5 was revised to read: |

| | |
|---|---|
| | 5. Criterion modified per Amendment 2:<br><br>5.1 **In Study 1**, meet the following requirements for prior or current medications for Crohn's disease:<br><br>a. Has previously demonstrated inadequate response, loss of response, or intolerance to 1 or more approved biologic therapies (eg, infliximab, adalimumab, certolizumab pegol, or vedolizumab) as outlined in Attachment 1.<br><br>**In Study 2**, meet the following requirements for prior or current medications for Crohn's disease:<br><br>b. Has failed conventional therapy as follows:<br>1) Is currently receiving corticosteroids and/or immunomodulators (ie, AZA, 6-MP, or MTX) at adequate therapeutic doses (Attachment 2).<br>**OR**<br>2) Has a history of failure to respond to or tolerate an adequate course of corticosteroids and/or immunomodulators (ie, AZA, 6-MP, or MTX) (Attachment 2).<br>**OR**<br>3) Is corticosteroid dependent or has had a history of corticosteroid dependency (Attachment 2).<br><br>Inclusion Criterion 6 was revised to read:<br><br>6. **In Study 3**, meet the following requirement for prior or current medications for Crohn's disease: has previously demonstrated inadequate response, loss of response, or intolerance to 1 or more approved biologic therapies (eg, infliximab, adalimumab, certolizumab pegol, or vedolizumab) as outlined in Attachment 1. |
| **Rationale:** To accurately assess potential injection-site reactions, additional instructions regarding injection-site location were added. | |
| Time and Events Schedule: Part I | Under Study Procedures, Administer study agent was revised to read:<br><br>Administer study agent [e] (SC study agent at a different location at each visit [Section 6]) |
| Time and Events Schedule: Part II | Under Study Procedures, Administer study agent was revised to read:<br><br>Administer study agent [e] (SC study agent at a different location at each visit [Section 6]) |
| 6. Dosage and Administration | The following was added at the beginning of the section:<br><br>Subcutaneous injections of study agent should be administered at different locations at each visit at which study agent is administered to allow for assessment of potential injection-site reactions. |
| **Rationale:** The inclusion criterion related to contraception was revised to be consistent with the core safety information for JNJ-64304500. | |
| 4.1. Inclusion Criteria | The following was added as the last sentence of Inclusion Criterion 13.b.1:<br><br>If using a hormone birth control method, a second method of birth control, such as a condom or diaphragm, must be used. |
| **Rationale:** The footnote for video ileocolonoscopy in the Time and Events Schedule Part II was updated to be consistent with the footnote for video ileocolonoscopy in the Time and Events Schedule Part I. | |

| | |
|---|---|
| Time and Events Schedule Part II | Footnote r was revised to read:<br>r. Because these procedures must not interfere with the collection of CDAI data, if performed on the day of these visits, the 7 days before the initiation of the colonoscopy preparation should be used to calculate CDAI scores for these visits. If the video ileocolonoscopic examination is not performed on the day of the visit, it must be performed at least 8 days before the Week 0 visit and no more than 8 days before the Week 12 visit. |

**Rationale:** The numbers of subjects exposed to JNJ-64304500 in clinical studies were updated to be consistent with the Investigator's Brochure.

| | |
|---|---|
| 1.2. Clinical Studies | This section was revised to read:<br>As of 11 November 2015, a total of 128 subjects had been exposed to JNJ-64304500 in 3 clinical studies: 88 subjects in 2 studies in rheumatoid arthritis (RA) and 40 subjects in a Phase 2a study in Crohn's disease. |
| 1.2.1. Rheumatoid Arthritis | This section was revised to read:<br>Two studies with JNJ-64304500 were conducted in subjects with active RA.<br>In **NN8555-3618**, a first-in-humans, Phase 1, single ascending dose/multiple ascending dose study that included single-dose (0.0002 to 7.5 mg/kg) and multiple-dose (0.02 to 4 mg/kg every 2 weeks for 4 administrations) parts, 13 dose levels were evaluated in 24 subjects exposed to JNJ-64304500 in the single ascending dose portion and 23 subjects exposed to JNJ-64304500 in the multiple ascending dose portion. Subcutaneous administration of JNJ-64304500 was well tolerated at the dose ranges investigated and no safety signals were associated with either the single- or multiple-dose regimens.<br>In **NN8555-3796**, a Phase 2a, randomized, single-dose, double-blind, placebo-controlled, parallel-group study, clinical efficacy was assessed in subjects with active RA concomitantly treated with methotrexate (MTX). In this study, a single SC injection of 4 mg/kg JNJ-64304500 in 41 subjects exposed to JNJ-64304500 did not result in a statistically significant reduction in disease activity at Weeks 6, 12, or 24 after treatment compared with placebo. JNJ-64304500 was well tolerated and no safety concerns were raised during the study.<br>Additional details about these studies are provided in the latest version of the IB. |

**Rationale:** The timing for the collection of the random population pharmacokinetic (PK) samples was clarified.

| | |
|---|---|
| Time and Events Schedule: Part I | Footnote x was revised to read:<br>x. Each random population PK sample should be collected at any time between Weeks 0 and 2 (after the Week 0 study agent administration) and between Weeks 22 and 24 (after the Week 22 study agent administration). The date for the random population PK sample collection will be at the subject's discretion. However, samples should not be drawn on or within 1 day of a scheduled visit (ie, the Week 0, 2, 22, or 24 visit) in this time period. |
| Time and Events Schedule: Part II | Footnote x was revised to read:<br>x. Each random population PK sample should be collected at any time between Weeks 0 and 2 (after the Week 0 study agent administration) and between Weeks 20 and 24 (after the Week 20 study agent administration). The date for the random population PK sample collection will be at the subject's discretion. However, samples should not be drawn on or within 1 day of a scheduled visit (ie, the Week 0, 2, 20, or 24 visit) in this time period. |

| | |
|---|---|
| 9.3.1. Evaluations | The first paragraph was revised to read:<br><br>Serum samples used to evaluate the PK and immunogenicity of JNJ-64304500 and ustekinumab (antibodies to JNJ-64304500 and antibodies to ustekinumab) will be collected according to the Time and Events Schedules. Samples collected for analyses of serum concentration of JNJ-64304500 and ustekinumab and antibodies to JNJ-64304500 or ustekinumab may additionally be used to evaluate safety or efficacy aspects that address concerns arising during or after the study period, for further characterization of immunogenicity or for the evaluation of relevant biomarkers. Genetic analyses will not be performed on these serum samples. Subject confidentiality will be maintained. The exact dates and times of sample collection must be recorded on the laboratory requisition form. |
| **Rationale:** The instructions regarding the components of the CDAI were clarified. | |
| Time and Events Schedule: Part I | Footnote e was revised to read:<br><br>e. All assessments are to be completed before study agent administration, unless otherwise specified. It is recommended that PRO assessments be completed first, starting with the PRO components of the CDAI.<br><br>Footnote n was revised to read:<br><br>n. The hematocrit value obtained during screening will be used to calculate CDAI at Week 0. For other visits, the most recent hematocrit value obtained before a visit will be used to calculate the CDAI for that visit. |
| Time and Events Schedule: Part II | Footnote e was revised to read:<br><br>e. All assessments are to be completed before study agent administration, unless otherwise specified. It is recommended that PRO assessments be completed first, starting with the PRO components of the CDAI.<br><br>Footnote n was revised to read:<br><br>n. The hematocrit value obtained during screening will be used to calculate CDAI at Week 0. For other visits, the most recent hematocrit value obtained before a visit will be used to calculate the CDAI for that visit. |
| **Rationale:** The instructions regarding the procedures/assessments to be completed at early termination were clarified. | |
| Time and Events Schedule: Part I | Footnote c was revised to read:<br><br>c. Subjects who <u>terminate study participation</u> should complete an early termination visit. If a subject completed assessments at Week 24 and terminated after Week 24, the only assessments that should be performed at the early termination visit are those planned for the final efficacy and safety follow-up. Subjects who <u>discontinue study agent administration</u> before Week 8 (but have not terminated study participation) should complete the Week 8 and Week 12 assessments and return for a final efficacy and safety follow-up visit approximately 16 weeks after their last study agent administration. Subjects who <u>discontinue study agent administration</u> after Week 8 (but have not terminated study participation) should complete the Week 12 and Week 24 assessments and return for a final efficacy and safety follow-up visit approximately 16 weeks after their last study agent administration. |

| | |
|---|---|
| Time and Events Schedule: Part II | Footnote c was revised to read:<br>c. Subjects who terminate study participation should complete an early termination visit. If a subject completed assessments at Week 24 and terminated after Week 24, the only assessments that should be performed at the early termination visit are those planned for the final efficacy and safety follow-up. Subjects who discontinue study agent administration before Week 8 (but have not terminated study participation) should complete the Week 8 and Week 12 assessments and return for a final efficacy and safety follow-up visit approximately 16 weeks after their last study agent administration. Subjects who discontinue study agent administration after Week 8 (but have not terminated study participation) should complete the Week 12 and Week 24 assessments and return for a final efficacy and safety follow-up visit approximately 16 weeks after their last study agent administration. |
| **Rationale:** Exploratory histologic assessments were added. | |
| Synopsis | In the Efficacy Evaluations section, the following was added:<br>• Histologic assessment |
| 2.1.2 Endpoints | The following was added:<br>• Endpoint(s) for histologic assessment based on the Global Histology Activity Score (GHAS; to be detailed in the Statistical Analysis Plan [SAP]). |
| 9.2.12 Histologic Assessment | In Section 9.2 Efficacy Evaluations, a new section 9.2.12 Histologic Assessment was added:<br>Histologic assessment will be performed using biopsy samples collected during endoscopy. Biopsy samples will be collected at screening, Week 12, and Week 24 from each of 3 predefined anatomic locations: the terminal ileum, splenic flexure, and rectum. The biopsy samples collected at Week 12 and Week 24 will be obtained near where the biopsy samples at screening were collected from each of the 3 predefined locations. Histologic assessment will be conducted by a central reader who is blinded to treatment groups. The GHAS will be used to evaluate histologic improvement. Additional details will be provided in the Study Reference Manual. |
| 11.2.1.2 Other Efficacy Endpoint Analyses | The following was added:<br>• Endpoint(s) for histologic assessment based on the GHAS (to be detailed in the SAP). |
| 11.5. Biomarker Analyses | The last sentence was revised to read:<br>Results of serum, whole blood analyses, stool, and mucosal biopsy analyses will be reported in separate technical reports. |
| **Rationale:** The requirement for an ileocolonoscopy biopsy sample collection at early termination was added as it was erroneously omitted from the protocol. | |
| Time and Events Schedule: Part II | In the Pharmacodynamics/Biomarkers Assessments section, an X was added in the Early Termination column of the Ileocolonoscopy biopsy sample collections for RNA and histology row. |
| **Rationale:** To clarify the efficacy assessment calculations for change in abdominal pain score and change in stool frequency score, mean daily average was revised to daily average. | |

| | |
|---|---|
| 2.1.2 Endpoints | The following were revised to read:<br>• Change in abdominal pain score (daily average based on the CDAI assessment) from baseline at all postbaseline visits.<br>• Change in stool frequency score (daily average based on the CDAI assessment) from baseline at all postbaseline visits. |
| 11.2.1.2. Other Efficacy Endpoint Analyses | The following were revised to read:<br>• Change in abdominal pain score (daily average based on the CDAI assessment) from baseline at all postbaseline visits.<br>• Change in stool frequency score (daily average based on the CDAI assessment) from baseline at all postbaseline visits. |
| **Rationale:** The rationale for an interim analysis was clarified. | |
| Synopsis, Overview of Study Design | The fourth paragraph was revised to read:<br>An interim analysis is planned in Study 1 of Part I when the first 80% of the randomized Bio-IR subjects in Study 1 of Part I (at least 40 Bio-IR subjects and at least 30 Bio-IR/SNP-positive subjects per treatment group) have completed their Week 8 visit or have terminated their study participation before Week 8. This interim analysis will allow for an earlier start of Part II (ie, Study 3, the dose-ranging part) if a sufficient effect is observed. Part II will be initiated if results from Study 1 of Part I demonstrates acceptable safety and efficacy, either at the interim analysis or when 100% of the Bio-IR subjects have completed their Week 12 visit (or have terminated study participation prior to this time). |
| 3.1.3. Interim Analysis | The second paragraph was revised to read:<br>This interim analysis will allow for an earlier start of Part II (ie, Study 3, the dose-ranging part) if a sufficient effect is observed (see Section 11.10). As this interim analysis does not affect the conduct or completion of Study 1, it will be considered administrative and will not require multiplicity adjustment for the final Study 1 analysis. |
| 11.10. Interim Analysis | The first paragraph was revised to read:<br>An interim analysis is planned in Study 1 when the first 80% of the randomized Part I Bio-IR subjects (at least 40 Bio-IR subjects and at least 30 Bio-IR/SNP-positive subjects per treatment group) have completed their Week 8 visit or have terminated their study participation before Week 8. This interim analysis will allow for an earlier start of Part II (ie, Study 3, the dose-ranging part) if a sufficient effect is observed. As this interim analysis does not affect the conduct or completion of Study 1, it will be considered administrative and will not require multiplicity adjustment for the final Study 1 analysis. |
| **Rationale:** Safety data for dose selection were clarified. | |
| 3.2.9.1. JNJ-64304500 | The first paragraph was revised to read:<br>The results of PK, PD and efficacy analyses and safety data from previous clinical studies of JNJ-64304500 in subjects with RA and Crohn's disease were used to guide the dose selection for the 3 studies in this protocol. In study NN8555-3618, the highest single SC dose investigated was 7.5 mg/kg and the highest multiple SC dose regimen investigated was 4 mg/kg q2w for a total of 4 doses in RA subjects. JNJ-64304500 was well tolerated and no safety concerns were identified in subjects with RA or Crohn's disease from the previous clinical studies. In addition, a 52-week repeat-dose toxicology study has demonstrated a no-observed-adverse-effect level (NOAEL) of 100 mg/kg SC once weekly in cynomolgus monkeys. The exposures achieved in the last dosage interval (100 mg/kg) of the 52-week study were 10,900 and 8,480 µg/mL for maximum concentration ($C_{max}$), and 1,520,000 and 1,250,000 hr·µg/mL for $AUC_{0\text{-}168hr}$, for male and female monkeys, respectively. Based on these safety and toxicology findings, it is expected that the |

| | |
|---|---|
| | proposed dose regimens of JNJ-64304500 would have acceptable safety profiles.<br>The last paragraph was revised to read:<br>The selection of the 4 different dosing regimens of JNJ-64304500 in Part I and Part II was based on all available PK, efficacy and safety data from the previous clinical studies and from a 52-week repeat-dose toxicology study. It should be noted that the current available information on JNJ-64304500 has not established the relationship between the NKG2D RO and clinical effects of the drug. In addition, the currently reported antibody distribution coefficients for intestinal tissues may have limitations and the JNJ-64304500 concentrations in the intestine may not be accurately predicted. Nevertheless, the use of 4 different dose regimens of JNJ-64304500 in Part I and Part II, which will provide a wide range of drug exposures while remaining within the acceptable safety margins, is anticipated to provide a robust characterization of the exposure-response relationship of JNJ-64304500 in the treatment of Crohn's disease. |
| **Rationale:** The definitions of inadequate initial response, loss of response, or intolerance to TNF-antagonist therapies or vedolizumab were updated. | |
| Attachment 1 | Attachment 1 was replaced with the updated definitions. |
| **Rationale:** Minor errors were noted. | |
| Throughout the protocol | Minor grammatical, formatting, or spelling changes were made. |

**Amendment 1** (14 June 2016)

This amendment is considered to be substantial based on the criteria set forth in Article 10(a) of Directive 2001/20/EC of the European Parliament and the Council of the European Union.

**The overall reason for the amendment:** The overall reason for the amendment was to address health authority requests.

Clarification of specific study procedures is also included in this amendment as described below.

| Applicable Sections | Description of Changes |
|---|---|
| **Rationale:** At the request of a health authority, collection of stool consistency data using the Bristol stool form scale and assessment of abdominal pain based on the Numerical Rating Scale (NRS) were added, as well as exploratory analyses of these data. | |
| Synopsis | In the Efficacy Evaluations section, the following bullets on Bristol stool form scale and assessment of abdominal pain were added:<br>• Bristol stool form scale<br>• Abdominal pain Numerical Rating Scale (NRS) |
| Time and Events Schedule: Part I and Part II | In the Efficacy Assessments section, the Bristol stool form scale was added as an assessment with data collection at Weeks 0, 2, 4, 6, 8, 10, and 12. Also, a new footnote o was added and subsequent footnotes were renumbered:<br>o. Subject will complete the Bristol stool form scale as a daily diary entry and bring the diary to each visit. Daily diary information should be collected for every day prior to each visit.<br><br>In the Efficacy Assessments section, abdominal pain NRS was added as an assessment with diary data collection at all visits. Also, a new footnote p was added and subsequent footnotes were renumbered: |

| Applicable Sections | Description of Changes |
|---|---|
| | p. Subject will complete the abdominal pain NRS assessment as a daily diary entry and bring the diary to each visit. For all visits up to Week 24, daily diary information should be collected for every day prior to the visits. For the final efficacy and safety visit, daily diary information should be collected for 14 days prior to the visit. |
| 2.1.2 Endpoints | The following efficacy endpoints bullets were added to each of the 3 studies:<br>• Change in abdominal pain score (mean daily average based on the CDAI assessment) from baseline at all postbaseline visits.<br>• Change in stool frequency score (mean daily average based on the CDAI assessment) from baseline at all postbaseline visits.<br>• Endpoint(s) based on Bristol stool form scale (to be detailed in the Statistical Analysis Plan).<br>• Change in abdominal pain from baseline at all postbaseline visits based on a 0-10 Numerical Rating Scale (NRS). |
| 9.2 Efficacy Evaluations | The section was revised to read:<br>The CDAI will be the primary tool for assessing disease activity response to JNJ-64304500, along with PRO-2, PRO-3, Bristol stool form scale, and abdominal pain based on NRS 0-10 scale. The degree of inflammation will be assessed by measuring serum CRP concentrations. Stool samples will be collected and analyzed to evaluate changes in markers that may reflect JNJ-64304500 or ustekinumab treatment. The well-being of subjects will be measured using the IBDQ and the SF-36. Mucosal healing will be assessed by ileocolonoscopy. For subjects with fistulizing disease, fistula closure will also be assessed. |
| 9.2.2 Bristol Stool Form Scale | In Section 9.2 Efficacy Evaluations, a new Section 9.2.2 Bristol Stool Form Scale was added:<br>The Bristol stool form scale is a medical aid to classify the form (or consistency) of human feces into 7 categories.[8] It has been used as a research tool to evaluate the effectiveness of treatments for various diseases of the bowel (eg, irritable bowel syndrome).[4] Subjects will complete the Bristol stool form scale as a daily diary entry and bring the diary to each visit up to Week 12. |
| 9.2.3 Abdominal Pain Numerical Rating Scale | In Section 9.2 Efficacy Evaluations, a new Section 9.2.3 Abdominal Pain Numerical Rating Scale was added:<br>The NRS for pain is a unidimensional measure of pain intensity in adults.[5] An 11-point (0-10) NRS will be used to evaluate abdominal pain. The score of 0 represents “no pain” and the score of 10 represents the “pain as bad as you can imagine”, with greater scores indicating greater pain severity and intensity. Subjects will select only one number that best reflects their pain at its worst in the past 24 hours. The abdominal pain NRS will be assessed daily. Subjects are to complete a daily diary entry and bring the diary to each visit. |

| Applicable Sections | Description of Changes |
|---|---|
| 11.2.1.2 Other Efficacy Endpoint Analyses | The following new bullets were added:<br>• Change in abdominal pain score (mean daily average based on the CDAI assessment) from baseline at all postbaseline visits.<br>• Change in stool frequency score (mean daily average based on the CDAI assessment) from baseline at all postbaseline visits.<br>• Endpoint(s) based on Bristol stool form scale (to be detailed in the SAP).<br>• Change in abdominal pain from baseline at all postbaseline visits based on a 0-10 NRS. |
| 15 Study-Specific Materials | Study-specific materials pertaining to the Bristol stool form scale and assessment of abdominal pain were added:<br>• Bristol stool form scale diary<br>• Abdominal pain NRS diary |
| References | Two references for the Bristol stool form scale (Dove et al 2013 and Lewis/Heaton 1997) were added to the References and other references were renumbered accordingly.<br>One reference for the abdominal pain NRS assessment (Hawker et al 2011) was added to the References and other references were renumbered accordingly. |

**Rationale:** Text was revised to clarify that the sample size for Study 3 was based on the power to detect a dose-response signal and the sample size/power considerations for the pairwise comparisons of the JNJ-64304500 groups with placebo were based on the comparison of the high dose group with placebo.

| | |
|---|---|
| Synopsis | In the Statistical Methods section, the text under Sample Size Determination was revised to read:<br>Sample size calculations for Study 1 (Part I Bio-IR subjects) and Study 2 (Part I Bio-NF subjects) were determined by the power to detect a significant difference in the change from baseline in the CDAI score at Week 8 (primary endpoint in each study) between JNJ-64304500 and placebo using a 2-sample t-test. Sample size calculation for Study 3 (Bio-IR subjects) was determined by the power to detect a dose-response signal in the change from baseline in the CDAI score at Week 8 (primary endpoint in Study 3) using the Multiple Comparison Procedures with modeling techniques (MCP-Mod) method.<br>For Study 1, assuming the mean CDAI change from baseline at Week 8 is -79 in the JNJ-64304500 group and -25 in the placebo group with a common SD of 92, 50 subjects per treatment group will provide approximately 80% power to detect a treatment difference between JNJ-64304500 and placebo at an overall Type 1 error of 0.05.<br>For Study 2, assuming the mean CDAI change from baseline at Week 8 is -152 in the JNJ-64304500 group and -66 in the placebo group with a common SD of 100, 50 subjects per treatment group will provide approximately 99% power to detect a treatment difference between JNJ-64304500 and placebo at an overall Type 1 error of 0.05.<br>For Study 3, assuming the mean CDAI change from baseline at Week 8 is -79 in the JNJ-64304500 group and -25 in the placebo group with a common SD of 92, 50 subjects per treatment group will provide a mean power of 85% to detect a dose-response signal based on 7 candidate dose-response models at an overall Type 1 error of 0.05. |

| Applicable Sections | Description of Changes |
|---|---|
| 11.1.2.1 All Bio-IR Subjects | The section was revised to read:<br><br>Using the same assumptions as were used for the Bio-IR population in Study 1, 50 subjects per treatment group will provide a mean power of 85% to detect a dose-response signal for change from baseline in CDAI at Week 8 based on 7 candidate dose-response models (to be detailed in the SAP) at an overall Type 1 error of 0.05 (2-sided). Fifty subjects per treatment group will also provide approximately 80% power to detect a treatment difference between the JNJ-64304500 treatment group with the highest dose and the placebo treatment group for change from baseline in CDAI at Week 8 at a Type I error of 0.05 (2-sided; Table 4). This will result in a total sample size of 250 subjects in Part II (incorporating an additional 50 subjects for the ustekinumab treatment group).<br><br>Fifty Bio-IR subjects per treatment group in Part II will also provide 90% power to detect a difference between the JNJ-64304500 treatment group with the highest dose and the placebo treatment group in the proportion of subjects in clinical remission at Week 8 (the first major secondary endpoint) at a Type 1 error of 0.05 (2-sided; Table 4), assuming JNJ-64304500 has a remission rate of 31%, which is 10% greater than the ustekinumab remission rate in CNTO1275CRD3001. |
| 11.1.2.2 Bio-IR Subjects Who Are SNP-Positive (Bio-IR/SNP+) | The section was revised to read:<br><br>Based on the assumption that 75% of the Crohn's disease population will be SNP-positive, 50 Bio-IR subjects will provide approximately 38 Bio-IR/SNP+ subjects. Thirty-eight Bio-IR/SNP+ subjects per group will provide 80% power to detect a difference between the JNJ-64304500 treatment group with the highest dose and the placebo treatment group in the proportion of subjects in clinical remission at Week 8 at a Type 1 error of 0.05 (2-sided; Table 4), assuming JNJ-64304500 has a remission rate of 31%, which is 10% greater than the ustekinumab remission rate in CNTO1275CRD3001. |
| **Rationale:** In order to clarify the Efficacy Assessments (CDAI assessments, Fistula exam, and Collect and review diary cards) and the Clinical Laboratory Assessments (Hematology) in the Time and Events Schedules, the frequency of completing these events was increased to all visits of the Main Study phase (and to the Final efficacy and safety follow-up visit for the Fistula Exam). | |
| Time and Events Schedule: Part I | In the Efficacy Assessments section (CDAI assessments, Fistula exam, and Collect and review diary cards) and the Clinical Laboratory Assessments section (Hematology), an X was added at Weeks 14, 18, and 22 for completion of these events. An X was also added at the Final Efficacy and Safety Follow-Up Visit for completion of the Fistula Exam. A new footnote m was added for the collection and review of diary cards, and subsequent footnotes were renumbered:<br><br>m. For all visits up to Week 24, daily diary information should be collected for every day prior to each visit. For the final efficacy and safety visit, daily diary information should be collected for 14 days prior to the visit. |
| Time and Events Schedule: Part II | In the Efficacy Assessments section (CDAI assessments, Fistula exam, and Collect and review diary cards) and the Clinical Laboratory Assessments section (Hematology), an X was added at Week 14 for completion of these events. An X was also added at the Final Efficacy and Safety Follow-Up Visit for completion of the Fistula Exam. A new footnote m was added for the collection and review of diary cards, and subsequent footnotes were renumbered:<br><br>m. For all visits up to Week 24, daily diary information should be collected for every day prior to each visit. For the final efficacy and safety visit, daily diary information should be collected for 14 days prior to the visit. |

| Applicable Sections | Description of Changes |
|---|---|
| **Rationale:** At the request of a health authority to identify early immunogenic responses, an additional sample for anti-drug antibody analysis was added at Week 2 in Part I (Studies 1 and 2) and Part II (Study 3). | |
| Time and Events Schedule: Part I and Part II | Added an X for Week 2 collection of samples to measure antibodies to JNJ-64304500 in Part I, and Assessment for antibody to study agent in Part II. |
| **Rationale:** In order to clarify instructions on sample handling for site investigators, additional footnotes were added to the Time and Events Schedules, and new text was added to Section 9.3.1 Evaluations. | |
| Time and Events Schedule: Part I and Part II | To the events of 'JNJ-64304500 serum concentration', 'JNJ-64304500 random population PK sample' and 'Antibodies to JNJ-64304500' in Part I, and 'Study agent serum concentration', 'JNJ-64304500 random population PK sample' and 'Assessment for antibody to study agent' in Part II, a new footnote w was added:<br><br>w. At visits where only serum concentration of study agent will be evaluated (ie, no antibodies to study agent will be evaluated), 1 venous blood sample of sufficient volume should be collected, and each serum sample should be divided into 2 aliquots (1 for serum concentration of study agent, and a back-up). At visits where serum concentration of study agent and antibodies to study agent will be evaluated, 1 venous blood sample of sufficient volume should be collected. Each serum sample will be divided into 3 aliquots (1 each for serum concentration of study agent, antibodies to study agent, and a back-up). |
| 9.3.1 Evaluations | New text was added to the second paragraph in order to clarify instructions on sample handling for site investigators, as follows:<br><br>At visits where only serum concentration of study agent will be evaluated (ie, no antibodies to study agent will be evaluated), 1 venous blood sample of sufficient volume should be collected, and each serum sample should be divided into 2 aliquots (1 for serum concentration of study agent, and a back-up). |
| **Rationale:** The requirement for a video ileocolonoscopy at early termination was added as it was erroneously omitted from the protocol. | |
| Time and Events Schedule: Part II | In the Efficacy Assessments section, an X was added in the Early Termination column of the Video ileocolonoscopy row. |
| **Rationale:** In order to specify the type of procedures that will not be included in the category of Crohn's disease-related hospitalization and surgeries, a new footnote was added to the Time and Events Schedules. | |
| Time and Events Schedule: Part I and Part II | To the event of 'Crohn's disease-related hospitalization and surgeries', a new footnote aa was added:<br><br>aa. Hospitalization for ileocolonoscopy is not included in this category. |
| **Rationale:** In countries where tuberculin is not available for the skin test, this method of testing will not be required. QuantiFERON-TB Gold and chest radiograph must still be done if tuberculosis (TB) is suspected, along with referral to a TB specialist if possible. | |
| Time and Events Schedule: Part I and Part II | Footnote g was revised to read:<br><br>g. All subjects will undergo QuantiFERON-TB Gold testing. In countries where the QuantiFERON-TB Gold test is not registered/approved, TB skin testing will also be required (recommended but not required for study centers in countries where tuberculin is not available). The QuantiFERON-TB Gold test is not required at screening for subjects with a history of latent TB and appropriate treatment (as described in Inclusion Criterion 11a). |
| Time and Events Schedule: Part I and Part II | Footnote l was revised to read:<br><br>l. If TB is suspected at any time during the study, a chest radiograph and QuantiFERON-TB Gold test should be performed. In countries where the QuantiFERON-TB Gold test is |

| Applicable Sections | Description of Changes |
|---|---|
| | not registered/approved, TB skin testing should also be performed (recommended but not required for study centers in countries where tuberculin is not available). |
| 4.1 Inclusion Criteria | Inclusion Criterion 11d was revised to specify that a tuberculin skin test is recommended but not required for study centers in countries where tuberculin is not available, as follows:<br><br>Within 2 months before the first administration of study agent, either have negative QuantiFERON-TB Gold test (Attachment 3), or have a newly identified positive QuantiFERON-TB Gold test in which active TB has been ruled out, and for which appropriate treatment for latent TB has been initiated either before or simultaneously with the first administration of study agent (except in countries with high multidrug-resistant TB burden [eg, Brazil, China, India, the Russian Federation, and South Africa]), where subjects with a newly identified positive QuantiFERON-TB Gold test result are excluded). Indeterminate results should be handled as outlined in Section 9.1.2. A negative tuberculin skin test (Attachment 4) is additionally required if the QuantiFERON-TB gold test is not approved/registered in that country. A tuberculin skin test is recommended but not required for study centers in countries where tuberculin is not available. The QuantiFERON-TB Gold In-Tube test is not required at screening for subjects with a history of latent TB and appropriate treatment as described above in Inclusion Criterion 11a. |
| 9.1.2 Screening Period | The sixth paragraph was revised as follows:<br><br>Subjects with a negative QuantiFERON-TB Gold test result (and a negative tuberculin skin test result in countries in which the QuantiFERON-TB Gold test is not approved/registered or the tuberculin skin is mandated by local health authorities) are eligible to continue with prerandomization procedures. A tuberculin skin test is recommended but not required for study centers in countries where tuberculin is not available. Subjects with a newly identified positive QuantiFERON-TB Gold (or tuberculin skin) test result must undergo an evaluation to rule out active TB and initiate appropriate treatment for latent TB. Appropriate treatment for latent TB is defined according to local country guidelines for immunocompromised patients. If no local country guidelines for immunocompromised patients exist, US guidelines must be followed, or the subject will be excluded from the study. **Note:** Subjects in countries with high multidrug-resistant TB burden (eg, Brazil, China, India, the Russian Federation, and South Africa) identified with latent TB at screening will be excluded from participating in the study. |
| 9.7 Safety Evaluations | Under the subheading Early Detection of Active Tuberculosis, the last paragraph was revised as follows:<br><br>Subjects who experience close contact with an individual with active TB during the conduct of the study must have a repeat chest radiograph, a repeat QuantiFERON®-TB Gold test, a repeat tuberculin skin test in countries in which the QuantiFERON-TB Gold test is not approved/registered and, if possible, referral to a physician specializing in TB to determine the subject's risk of developing active TB and whether treatment for latent TB is warranted. A tuberculin skin test is recommended but not required for study centers in countries where tuberculin is not available. A positive QuantiFERON-TB Gold (or tuberculin skin) test result should be considered detection of latent TB. If the QuantiFERON-TB Gold test result is indeterminate, the test should be repeated as outlined in Section 9.1.2. If recommended, treatment for latent TB must be initiated before or simultaneously with the administration of further study agent. Subjects who discontinue treatment for latent TB prematurely or who are noncompliant with therapy must immediately discontinue further administration of study agent and be encouraged to return for all subsequent scheduled study visits. |

| Applicable Sections | Description of Changes |
|---|---|
| 10.2 Discontinuation of Study Treatment | The following TB screening criterion for subject ineligibility was revised to read:<br>– A subject undergoing continued evaluation has a chest radiograph with evidence of current active TB and/or a positive QuantiFERON-TB Gold test result and/or a positive tuberculin skin test result in countries in which the QuantiFERON-TB Gold is not approved/registered (recommended but not required for study centers in countries where tuberculin is not available) unless active TB can be ruled out and appropriate treatment for latent TB can be initiated either prior to or simultaneously with the next administration of study agent and continued to completion. (Note: Study agent must be discontinued for all subjects diagnosed with latent TB in countries with high multidrug-resistant TB burden [eg, Brazil, China, India, the Russian Federation, and South Africa]). |
| Attachment 3 QuantiFERON-TB Gold Testing | Under the subheading Adherence to Local Guidelines, the following sentence was added to the second (and final) paragraph:<br>In countries where the QuantiFERON-TB Gold test is not approved/registered, the tuberculin skin test is recommended but not required if tuberculin is not available. |
| Attachment 4 Tuberculin Skin Testing | The following sentence was added as the last sentence:<br>**NOTE:** The tuberculin skin test is recommended but not required in countries where tuberculin is not available. |
| **Rationale:** Language was added to clarify that an indeterminate result for SNP would not meet inclusion criteria for the study, and that the deoxyribonucleic acid (DNA) samples can be used for both the required and optional DNA research. | |
| 4.1 Inclusion Criteria | Inclusion Criterion 18 was revised to read:<br>DNA sample collection for SNP testing is required for all subjects in this study. Each subject must have a SNP status of either positive or negative. Each subject must sign a separate informed consent form (ICF) if he or she agrees to consent to additional optional DNA research where local regulations permit. Refusal to give consent for the optional DNA research does not exclude a subject from participation in the study. |
| **Rationale:** Additional details were added for maintaining the study blind during preparation of JNJ-64304500 and placebo. | |
| 5.2 Blinding | The first paragraph was updated to add the words ‘or syringe number’, and was revised to read:<br>To maintain the study blind, the study agent container will have a label containing the study name and medication number or syringe number. The label will not identify the study agent in the container. The medication number or syringe number will be entered in the case report form (CRF) when the drug is dispensed. The study agents will be identical in appearance and packaging. |
| **Rationale:** The text in the Prestudy and Concomitant Therapy section was updated for clarity. | |
| 8 Prestudy and Concomitant Therapy | The following (first) sentence was deleted for clarity:<br>Prestudy therapies administered up to 30 days before first dose of study drug must be recorded at screening. |

| Applicable Sections | Description of Changes |
|---|---|
| **Rationale:** The text pertaining to concomitant medications was updated to specify that medications are required to remain stable through the final study visit. | |
| 8.1 Concomitant Medications | The second paragraph was revised to specify that concomitant medications are required to remain stable through the final study visit as opposed to Week 24, as follows:<br><br>With the exception of oral corticosteroids, subjects who are receiving these medications for Crohn's disease at baseline should maintain a stable dose through the final efficacy and safety visit. Corticosteroids must be maintained at baseline doses through Week 12 (see Section 8.1.1). |
| **Rationale:** The third paragraph of Section 8.1.1 Corticosteroid Tapering was moved to Section 8.1 Concomitant Medications for alignment of flow of contents, and text was updated for clarity. | |
| 8.1 Concomitant Medications | The third paragraph of Section 8.1.1 Corticosteroid Tapering was moved to Section 8.1 Concomitant Medications. In addition, the text was revised to indicate that subjects may transiently use increased doses of corticosteroids for reasons other than worsening of Crohn's disease, but not for loss of response to treatment for Crohn's disease, as follows:<br><br>After Week 12, subjects may transiently use (ie, for <4 weeks) increased doses of corticosteroids for reasons other than worsening of Crohn's disease (eg, stress doses of corticosteroids for surgery, asthma, adrenocortical insufficiency). |
| **Rationale:** In Section 9.2.1 Crohn's Disease Activity Index, the text was updated to clarify that subjects will complete CDAI as a daily diary entry and will bring the diary to each study visit. | |
| 9.2.1 Crohn's Disease Activity Index | The section was revised to read:<br><br>The CDAI will be assessed by collecting information on 8 different Crohn's disease-related variables (Attachment 5):[1] extra-intestinal manifestations, abdominal mass, weight, hematocrit, total number of liquid stools, abdominal pain/cramping, use of antidiarrheal drug(s) and/or opiates, and general well-being. The last 4 variables are scored over 7 days by the subject on a diary card. The PRO-2 score is based on the CDAI components of the total number of liquid stools and abdominal pain/cramping. The PRO-3 score, which is also based on the CDAI, comprises the PRO-2 components plus general well-being. Subjects are to complete a daily diary entry and bring the diary to each visit. |
| **Rationale:** At the request of a health authority, the National Cancer Institute Common Terminology Criteria for Adverse Events definitions of adverse events and hepatotoxicity guidelines were incorporated. | |
| 9.7 Safety Evaluations | Under the subheading Clinical Laboratory Tests, the first paragraph was revised as follows:<br><br>Blood samples for serum chemistry and hematology will be collected. The investigator must review the laboratory results, document this review, and record any clinically relevant changes occurring during the study in the Adverse Event section of the CRF. Clinical hematology and chemistry laboratory test results will be graded by the National Cancer Institute Common Terminology Criteria for Adverse Events (NCI-CTCAE) toxicity criteria. The laboratory reports must be filed with the source documents. |
| 10.2 Discontinuation of Study Treatment | The fourth bullet was revised as follows:<br><br>• A serious adverse reaction occurs that is related to an injection or an infusion, including an injection-site or infusion reaction, resulting in bronchospasm with wheezing and/or dyspnea that requires ventilatory support **OR** that results in symptomatic hypotension with a decrease in systolic blood pressure >40 mm Hg or blood pressure <90/60 mm Hg. This may include events of NCI-CTCAE toxicity grade ≥3.<br><br>A new fifth bullet was added: |

| Applicable Sections | Description of Changes |
|---|---|
| | • Adverse events of NCI-CTCAE grade ≥3 potentially related to worsening of Crohn's disease will be evaluated by the investigator and the study medical monitor to make a determination on discontinuation of study agent. Discontinuation of study agent should be considered in subjects with worsening Crohn's disease where continuation of the study drug is not in the best interest of the subject. |
| 11.9 Safety Analyses | Under the subheading Clinical Laboratory Tests, the second bullet was revised to specify that the summary of maximum NCI-CTCAE toxicity grade for postbaseline hematology and chemistry laboratory values will be used to assess the safety of subjects, as follows:<br>• Summary of maximum NCI-CTCAE toxicity grade for postbaseline laboratory values (hematology and chemistry). |
| 12.1.3 Severity Criteria | The first sentence was revised as follows:<br>An assessment of severity grade will be made using the following general categorical descriptors as outlined in Section 12.1.3.1 and by NCI-CTCAE toxicity grade outlined in Section 12.1.3.2. |
| 12.1.3.1 Severity Criteria: General Categorical Descriptors | The following text was moved from Section 12.1.3 Severity Criteria to newly added Section 12.1.3.1 Severity Criteria: General Categorical Descriptors:<br>**Mild:** Awareness of symptoms that are easily tolerated, causing minimal discomfort and not interfering with everyday activities.<br><br>**Moderate:** Sufficient discomfort is present to cause interference with normal activity.<br><br>**Severe:** Extreme distress, causing significant impairment of functioning or incapacitation. Prevents normal everyday activities.<br><br>The investigator should use clinical judgment in assessing the severity of events not directly experienced by the subject (eg, laboratory abnormalities).<br><br>**Note:** Seriousness and severity should not be confused. A subject could experience a severe headache that would not qualify as a serious adverse event (SAE), while another might experience a mild stroke that, while not severe, would be considered serious. |
| 12.1.3.2 Severity Criteria Based on National Cancer Institute Common Terminology Criteria for Adverse Events Toxicity Grade | The following text was added to newly added Section 12.1.3.2 Severity Criteria Based on National Cancer Institute Common Terminology Criteria for Adverse Events Toxicity Grade:<br>An assessment of severity grade will also be made using the following NCI-CTCAE categorical descriptors. The NCI-CTCAE Grade refers to the severity of the AE. The NCI-CTCAE displays Grades 1 through 5 with unique clinical descriptions of severity for each AE based on this general guideline:<br><br>**Grade 1:** Mild; asymptomatic or mild symptoms; clinical or diagnostic observations only; intervention not indicated.<br><br>**Grade 2:** Moderate; minimal, local or noninvasive intervention indicated; limiting age-appropriate instrumental Activities of Daily Living (ADL)*.<br><br>**Grade 3:** Severe or medically significant but not immediately life-threatening; hospitalization or prolongation of hospitalization indicated; disabling; limiting self-care ADL**. |

| Applicable Sections | Description of Changes |
|---|---|
| | **Grade 4:** Life-threatening consequences; urgent intervention indicated.<br><br>**Grade 5:** Death related to AE.<br><br>*Instrumental ADL refer to preparing meals, shopping for groceries or clothes, using the telephone, managing money, etc.<br><br>**Self-care ADL refer to bathing, dressing and undressing, feeding self, using the toilet, taking medications, and not bedridden. |
| **Rationale:** At the request of a health authority, additional guidelines on the evaluation of abnormal liver function tests were added. | |
| 9.7 Safety Evaluations | Under subheading Clinical Laboratory Tests, the following new bullet was added:<br>• **Abnormal liver function tests:** If laboratory testing for a subject enrolled in the study and receiving study drug reveals an increase of serum aminotransferases (ALT or AST) to >3x the upper limit of normal (ULN) and an increase of bilirubin to >2x ULN, study agent should be suspended immediately. In addition, laboratory tests for ALT, AST, alkaline phosphatase, and total bilirubin should be confirmed by a retest within 24 hours if possible, but no later than 72 hours following notification of test results. See Attachment 6 (Liver Safety Monitoring and Assessment) for additional information on monitoring and assessment of abnormal liver function tests. |
| 10.2 Discontinuation of Study Treatment | The following new bullet was added:<br>• Severe hepatic function abnormalities as described in Section 9.7. |
| **Rationale:** An attachment was added to provide clarification regarding the definitions of inadequate response to or intolerance of corticosteroids or AZA/6-MP and corticosteroid dependence. | |
| Attachment 2: Definitions of Inadequate Response to or Intolerance of Corticosteroids or AZA/6-MP and Corticosteroid Dependence | The definitions of an inadequate response to or intolerance of corticosteroids or AZA/6-MP and corticosteroid dependence were added to the protocol as Attachment 2. |
| **Rationale:** At the request of a health authority, additional guidance on the evaluation of subjects who have evidence of hepatotoxicity is provided. | |
| Attachment 6: Liver Safety Monitoring and Assessment | Liver Safety Monitoring and Assessment was added to the protocol as Attachment 6. |
| **Rationale:** Minor errors were noted. | |
| Throughout the protocol | Minor grammatical, formatting, or spelling changes were made. |

### SYNOPSIS

**A Phase 2b, Randomized, Double-blind, Placebo-controlled, Parallel-group, Multicenter Protocol to Evaluate the Safety and Efficacy of JNJ-64304500 in Subjects with Moderately to Severely Active Crohn's Disease**

**Protocol Number:** 64304500CRD2001

**EudraCT Number:** 2016-000634-21

JNJ-64304500 (formerly known as NNC0142-002) is a human immunoglobulin G4 isotype monoclonal antibody that binds specifically to the natural killer group 2 member D (NKG2D) receptor, and blocks NKG2D ligand binding, thereby preventing the downstream-signaling events that otherwise lead to cell proliferation and release of proinflammatory cytokines and cytotoxic mediators. Several lines of evidence from patients with Crohn's disease support the hypothesis that NKG2D receptor activation plays a role in disease pathogenesis by mediating the production of local cytokines, activation of an immune response, and direct cytotoxicity of target intestinal cells. Collectively, preclinical and clinical data on the expression of NKG2D ligands or proinflammatory cytokines in the target tissue and abnormal expression and activation of the NKG2D receptor on CD8+ and CD4+ T cells provide a rationale for the clinical development of inhibitors of the NKG2D receptor.

Results from a Phase 2a study (conducted by Novo Nordisk) provided initial evidence for the efficacy of JNJ-64304500 in the treatment of Crohn's disease, particularly in subjects who had not previously failed conventional therapy.

This Phase 2b protocol will further evaluate JNJ-64304500 in subjects with moderately to severely active Crohn's disease who have failed biologic or conventional therapy.

The protocol is divided into 2 parts.

In Part I, the following 2 studies will be conducted:

- Study 1: A study in subjects who are biologic intolerant or refractory (Bio-IR);
- Study 2: A study in subjects who have not previously failed a biologic therapy (Biologic nonfailure [Bio-NF]).

In Part II, the following study will be conducted:

- Study 3: A dose-ranging study in subjects who are Bio-IR or Bio-NF

A long-term extension (LTE) for Part II of the study was added to the protocol as part of Protocol Amendment 5. After Protocol Amendment 5 is implemented, subjects who complete Part II Week 24 assessments and who may benefit from continued treatment in the opinion of the investigator, are eligible to enter the Part II LTE to receive up to 52 weeks of additional treatment. Subjects who have completed Week 24 assessments prior to the implementation of Protocol Amendment 5 will not be eligible to enroll in the Part II LTE.

#### OBJECTIVES, ENDPOINTS, AND HYPOTHESIS

##### Objectives

The objectives are the same in each of the 2 parts.

##### *Primary Objectives*

- To evaluate the efficacy of JNJ-64304500 to reduce the Crohn's Disease Activity Index (CDAI) score from baseline.
- To evaluate the safety of JNJ-64304500.

##### *Secondary Objectives*

- To evaluate the efficacy of JNJ-64304500 to induce clinical remission, clinical response, and endoscopic healing of the mucosa, and to maintain remission.
- To evaluate the relationship between efficacy and the presence of the NKG2D and/or MICB single nucleotide polymorphism (SNP) biomarkers.
- To evaluate the efficacy of JNJ-64304500 to improve general and disease-specific health-related quality of life and to reduce Crohn's disease-related hospitalizations and surgeries.
- To evaluate the pharmacokinetics (PK), immunogenicity, pharmacodynamics (PD), and biomarkers (eg, reductions in C-reactive protein [CRP], fecal calprotectin, and fecal lactoferrin) of JNJ-64304500 therapy.

#### Endpoints

The data from Study 1 and Study 2 in Part I will be pooled for analysis.

##### *Primary Endpoints*

The primary endpoint for Part I is: Change from baseline in the CDAI score at Week 8.

The primary endpoint for Part II is: Change from baseline in the CDAI score at Week 12.

##### *Major Secondary Endpoints*

The following endpoints will be evaluated as major secondary endpoints only in Part II (the dose-ranging portion of the study); these endpoints will be evaluated in Part I but are not specified as major secondary endpoints.

- Clinical remission at Week 12 as measured by CDAI (CDAI <150).
- Clinical response at Week 12 as measured by CDAI (≥100-point reduction from baseline in CDAI or CDAI <150).
- Change in PRO-2 (the sum of the abdominal pain and stool frequency subscores of the CDAI score) from baseline at Week 12.
- Clinical remission at Week 12 as measured by PRO-2 (PRO-2 <75).
- Clinical response at Week 12 as measured by PRO-2 (≥50-point reduction from baseline in PRO-2 or PRO-2 <75).
- Change in Simple Endoscopic Score for Crohn's Disease (SES-CD) from baseline at Week 12.

#### Hypothesis

The hypothesis for Part I and Part II is that JNJ-64304500 is superior to placebo in inducing a reduction from baseline in CDAI in subjects with moderately to severely active Crohn's disease.

#### OVERVIEW OF STUDY DESIGN

This protocol is comprised of 2 parts (Part I [Study 1: Bio-IR subjects and Study 2: Bio-NF subjects] and Part II [Bio-IR and Bio-NF subjects]). Each study is a randomized, double-blind, placebo-controlled, parallel-group, multicenter study of JNJ-64304500 in subjects with moderately to severely active Crohn's disease. A minimum of 120 subjects and a maximum of 170 subjects (Bio-IR and Bio-NF) will be randomized in Part I. An additional 250 subjects will be randomized in Part II. Part I will study the safety and efficacy of a high dose regimen of JNJ-64304500 compared with placebo in subjects who have failed biologic or conventional therapy (Bio-IR or Bio-NF subjects, respectively). Part II will study the safety and efficacy of multiple-dose regimens of JNJ-64304500 compared with placebo, with ustekinumab (STELARA®) as a reference arm, in Bio-IR and Bio-NF subjects.

The duration of the study will be 38 weeks in Part I and 36 weeks in Part II for subjects who do not enter the Part II LTE. The study duration includes study agent administration visits and a final efficacy and safety follow-up visit. Eligible subjects will only participate in Part I or Part II of the study.

After Protocol Amendment 5 is implemented, subjects who complete Part II of the study through Week 24 and who may benefit from continued treatment in the opinion of the investigator, will be eligible to enter the Part II LTE to receive up to 52 weeks of additional treatment.

The Time and Events Schedules summarize the frequency and timing of assessments for efficacy, PK, biomarkers, immunogenicity, PD, and safety in Part I and Part II of the study.

An interim analysis is planned in Part I when 100 randomized Part I subjects (Study 1: Bio-IR and Study 2: Bio-NF) have completed their Week 12 visit or have terminated their study participation before Week 12. If acceptable safety and efficacy are established in the combined Bio-IR and Bio-NF populations, Part II of the protocol, which consists of a dose-ranging study in subjects who are Bio-IR or Bio-NF, will be initiated. The interim analysis will allow for timely evaluation of safety and efficacy in Part I and uninterrupted patient enrollment through a seamless transition from Part I to Part II of the trial. Enrollment for Part I will continue until the sponsor makes a decision on whether to start Part II based on the interim analysis or when a maximum of 170 subjects have been randomized, whichever occurs first. Part II will be initiated if results from Part I demonstrate acceptable safety and efficacy, either at the interim analysis or when all Part I subjects have completed their Week 12 visit (or have terminated study participation prior to Week 12). If Part II is not initiated based on the interim analysis results, then the results through Week 12 for all subjects in Part I (ie, when all randomized subjects in Part I have either completed the Week 12 visit or terminated study participation prior to Week 12) will be examined to determine whether to start Part II. Under this scenario, a pause in enrollment between Part I and Part II will occur.

An external Data Monitoring Committee (DMC) will review unblinded safety data from all subjects periodically to monitor subject safety.

The end of the 64304500CRD2001 study is defined as the date on which the last subject completes the last efficacy and safety follow-up visit.

#### SUBJECT POPULATION

The subject population consists of men or women ≥18 years of age with moderately to severely active Crohn's disease (of at least 3 months' duration), defined as a CDAI score ≥220 and ≤450, with elevated CRP >0.3 mg/dL (>3.0 mg/L) and/or calprotectin >250 mg/kg. Subjects must have colitis, ileitis, or ileocolitis previously confirmed at any time in the past by radiography, histology, and/or endoscopy. Subjects also must have previously failed or been intolerant to 1 or more approved biologic agents (ie, tumor necrosis factor alpha antagonists or vedolizumab; Bio-IR subjects) or have demonstrated an inadequate response to or failed to tolerate corticosteroids or immunomodulators (ie, 6-mercaptopurine, azathioprine, and methotrexate) but not a biologic (Bio-NF subjects).

#### DOSAGE AND ADMINISTRATION

##### *Part I*

In Part I, a minimum of 120 subjects and a maximum of 170 subjects (Bio-IR and Bio-NF) will be randomly assigned to receive placebo or the JNJ-64304500 high dose in a 1:1 ratio using permuted block randomization, stratified by baseline CDAI score (≤300 or >300) and SNP-positive status (yes or no). The Bio-IR and Bio-NF populations will be randomized separately.

The treatment groups in Part I will be as follows:

- Placebo subcutaneously (SC) at Weeks 0, 2, 4, 6, 8, and 10; from Week 12, these subjects will receive additional doses as follows:
  - Placebo-treated subjects who are in clinical response at Week 12 (≥100-point reduction from baseline in CDAI or CDAI <150) will continue to receive placebo SC injections every 2 weeks (q2w) from Week 12 through Week 22.
  - Placebo-treated subjects who are not in clinical response at Week 12 will receive JNJ-64304500 400 mg SC at Week 12, and then JNJ-64304500 200 mg SC from Week 14 through Week 22.
- JNJ-64304500 400 mg SC at Week 0 then 200 mg SC q2w through Week 22.

##### *Part II*

In Part II of the study, subjects (Bio-IR and Bio-NF) will be randomly assigned in equal proportions to receive placebo, 1 of 3 dose regimens of JNJ-64304500, or ustekinumab, using permuted block randomization, stratified by baseline CDAI score (≤300 or >300), SNP-positive status (yes or no), and Bio-IR status (yes or no). The treatment groups are as follows:

- Placebo SC at Weeks 0, 2, 4, 8; from Week 12, these subjects will receive additional doses as follows:
  - Placebo-treated subjects who are in clinical response at Week 12 (≥100-point reduction from baseline in CDAI or CDAI <150) will continue to receive placebo SC injections at Weeks 12, 14, 16 and 20.
  - Placebo-treated subjects who are not in clinical response at Week 12 will receive JNJ-64304500 150 mg SC at Week 12 and JNJ-64304500 75 mg SC at Weeks 14, 16, and 20.
- JNJ-64304500 high dose: 400 mg SC at Week 0 and 200 mg SC at Week 2 and Week 4, then 200 mg every 4 weeks (q4w) through Week 20.
- JNJ-64304500 middle dose: 150 mg SC at Week 0 and 75 mg SC at Week 2 and Week 4, then 75 mg q4w through Week 20.
- JNJ-64304500 low dose: 50 mg SC at Week 0 and 25 mg SC at Week 2 and Week 4, then 25 mg q4w through Week 20.
- Ustekinumab: tiered doses approximating 6 mg/kg intravenously (IV) at Week 0 and 90 mg SC at Weeks 8 and 16.

##### *Long-term Extension*

After Protocol Amendment 5 is implemented, subjects who complete Part II Week 24 assessments, and who may benefit from continued treatment in the opinion of the investigator, are eligible to enter the Part II LTE. In the Part II LTE, each subject will be eligible to receive up to 52 weeks of additional study drug (for a total of up to 72 weeks of study drug in Part II). The first dose of study drug within the Part II LTE will be administered after the Week 24 assessments have been completed. The dose may be administered on the same day as the Week 24 assessments, or may be administered on a later date, as long as the dose is administered within the Week 24 visit window.

Subjects will continue to receive the same treatment regimen during the Part II LTE that they were receiving between Week 12 and Week 20 in the main study phase of Part II (placebo, high, middle, low dose JNJ-64304500 or ustekinumab). To maintain the study blind, all patients, investigators, and sites will remain blinded to treatment allocation during the Part II LTE until the last subject in the Part II main study phase has completed the Week 24 assessments and the Week 24 data analysis has been completed.

The timing of the Week 24 data analysis is dependent upon the timing and completion of Part II enrollment. An individual subject's unblinding during the Part II LTE will depend upon his/her Part II enrollment date. Therefore, a portion of subjects will complete the entire Part II LTE in a blinded fashion before study unblinding, while other subjects could be unblinded to treatment allocation during their participation in the Part II LTE.

Any subject who withdraws from the Part II LTE prior to study unblinding will return for a final safety follow-up visit 16 weeks after the last dose of study drug. For subjects that remain in the Part II LTE at the time of study unblinding, continuation of study drug during the Part II LTE will be dependent upon treatment allocation as follows:

- **JNJ-64304500:** Subjects will continue to receive the same dose of JNJ-64304500 administered during the Part II main study phase. Subjects will continue to receive study drug q4w, although any additional placebo injections used to maintain the study blind during the blinded phase of the Part II LTE will be discontinued after study unblinding. Following the last scheduled dose of study drug at Week 72, a final safety follow-up visit will be performed at approximately Week 88. For subjects who discontinue study participation prior to Week 72, a final safety follow-up visit will be performed 16 weeks after the last dose of study drug.

- **Placebo**: Subjects receiving placebo who remain in the Part II LTE at the time of study unblinding will have their final safety follow-up visit subsequent to unblinding and will be discontinued from the study. No further follow-up visits will be performed.

- **Ustekinumab**: Subjects receiving ustekinumab will continue to receive it during the Part II LTE until unblinding occurs. After unblinding, based on the treating physician's clinical judgment, subjects may receive ustekinumab in a manner dependent on the country in which they are located:

    - If a subject is not continuing on ustekinumab after unblinding, the subject will need to return for a final safety follow-up visit that will be performed 16 weeks after the final dose of study ustekinumab.

    - If a subject continues on ustekinumab in a country where commercial ustekinumab is available and approved for the treatment of adult Crohn's disease, the treating physician is responsible for providing the subject with commercial ustekinumab, which should be administered according to the approved dosing regimen in each country. The final safety follow-up visit should be performed after unblinding but before receiving the first dose of commercial ustekinumab.

- If a subject continues on ustekinumab in countries where commercial ustekinumab is not available or approved for adult Crohn's disease, ustekinumab will continue to be provided to subjects through Week 72, followed by a final safety follow-up visit 16 weeks after the last dose of ustekinumab.

For eligible Part II subjects, participation in the Part II LTE is entirely voluntary. Eligible subjects who do not wish to enter the Part II LTE will complete the Part II Week 24 safety and efficacy assessments, followed by the final efficacy and safety assessments at Week 36.

During the Part II LTE, all concomitant medications, including Crohn's disease-specific medications (except for the prohibited medications listed in Section 8.2) may be administered at the discretion of the investigator.

All subjects in the Part II LTE will be assessed according to the Time & Events Schedule (Table 3, Table 4), which includes assessments, adverse events (AEs), laboratory analyses, and PK and immunogenicity samples.

#### EFFICACY EVALUATIONS

The following efficacy evaluations will be performed in Part I and Part II of the main study phase.

- Crohn's Disease Activity Index (CDAI)
- C-reactive protein (CRP)
- Fecal lactoferrin and fecal calprotectin
- Inflammatory Bowel Disease Questionnaire (IBDQ)
- 36-Item Short Form Health Survey (SF-36)
- Fistula assessment
- Bristol stool form scale
- Abdominal pain Numerical Rating Scale (NRS)
- Patient's Global Impression of Severity (PGIS) of Crohn's disease
- Patient's Global Impression of Change (PGIC) of severity of Crohn's disease
- Ileocolonoscopy
- Histologic assessment

Efficacy evaluations will not be performed during the Part II LTE.

#### PHARMACOKINETIC, PHARMACODYNAMIC, AND IMMUNOGENICITY EVALUATIONS

Serum samples will be used to evaluate the PK, PD, and immunogenicity of JNJ-64304500 (and ustekinumab in Part II) and will be collected from each subject at the timepoints indicated in the Time and Events Schedules.

#### DNA AND BIOMARKER EVALUATIONS

Biomarker assessments will be made to examine the biologic response to treatment and to identify biomarkers that are relevant to JNJ-64304500 treatment and/or Crohn's disease. Blood samples for serum-based biomarker analyses will be collected from all subjects to assess proteins related to the NKG2D pathway or the pathogenesis of Crohn's disease. Whole blood samples will be collected from all subjects for the analysis of RNA expression and T-cell receptor repertoire. Mucosal biopsy samples will be collected during ileocolonoscopy for the analysis of gene and/or protein expression and the histologic assessment of disease and/or healing. Receptor occupancy assessments for NKG2D and immunophenotyping assessments (including natural killer [NK] cells and CD8+ T cells) will also be performed.

Whole blood will be collected from all subjects for SNP analysis (the NKG2D SNP rs2255336 and the MICB [NKG2D ligand]); subjects who have signed an optional pharmacogenomics consent form will undergo complete genomic testing.

#### SAFETY EVALUATIONS

Safety evaluations will include AEs; clinical laboratory tests (chemistry and hematology); vital signs and physical examinations; a screening electrocardiogram; allergic reactions, infusion reactions and injection-site reactions; infections; and early detection of active tuberculosis.

#### STATISTICAL METHODS

##### Sample Size Determination

Sample size calculations for Part I (Bio-IR subjects and Bio-NF subjects combined) were determined by the power to detect a significant difference in the change from baseline in the CDAI score at Week 8 between JNJ-64304500 and placebo using a 2-sample t-test. Sample size calculations for Part II (Bio-IR and Bio-NF subjects) were determined by the power to detect a dose-response signal for the change from baseline in the CDAI score at Week 12 (primary endpoint in Part II) using the Multiple Comparison Procedures with modeling techniques (MCP-Mod) method.

For Part I, assuming the mean CDAI change from baseline at Week 8 is -98 in the JNJ-64304500 group and -46 in the placebo group with a common standard deviation (SD) of 96, 60 subjects per treatment group will provide 84% power to detect a treatment difference between JNJ-64304500 and placebo at an overall Type 1 error rate of 0.05.

For Part II, assuming the mean CDAI change from baseline at Week 12 is -111 in the JNJ-64304500 group and -51 in the placebo group with a common SD of 102, 50 subjects per treatment group will provide a mean power of 85% to detect a dose-response signal based on 7 candidate dose-response models at an overall Type 1 error rate of 0.05.

##### Efficacy Analyses

This protocol is comprised of 2 separate parts (Part I [Study 1: Bio-IR subjects and Study 2: Bio-NF subjects] and Part II [Bio-IR and Bio-NF subjects]). Part I and Part II will be analyzed separately with separate Type I error control for the primary endpoint (at the 0.05 level of significance). The other endpoints within each part will not be controlled for multiplicity.

For each part, the analysis set is all randomized subjects who received study agent. Efficacy analyses will be based on a modified intent-to-treat principle. Therefore, the efficacy data for each subject who received study agent will be analyzed according to the assigned treatment regardless of the actual treatment received.

For Part I, the primary endpoint of change from baseline in the CDAI score at Week 8 will be compared between the JNJ-64304500 treatment group and the placebo treatment group by an analysis of covariance model on the van der Waerden normal scores with treatment as a fixed factor and baseline CDAI score, SNP-positive status (yes or no), and Bio-IR status (yes or no) as covariates. Part I will be considered to be positive if a significant improvement is detected in the change from baseline in the CDAI score at Week 8 in the JNJ-64304500 group compared with the placebo group at the 0.05 level of significance.

The primary endpoint for Part II is the change from baseline in the CDAI score at Week 12. A unified strategy that combines Multiple Comparison Procedures with modeling techniques, MCP-Mod, will be used to analyze the dose-response data over the JNJ-64304500 doses. This approach consists of 2 major steps. The first step consists of testing the dose-response signal via multiple contrast tests while controlling the overall Type 1 error rate. If a dose-response signal is detected, the second step is to select a model that best describes the observed data and use it to estimate adequate doses with associated precision. The study will be considered positive if a dose-response signal for the primary endpoint is detected. In addition to the dose-response analysis, pairwise comparisons of the JNJ-64304500 treatment groups versus the placebo group will be performed for the change from baseline in the CDAI score at Week 12; these comparisons will not be adjusted for multiplicity.

The major secondary endpoints are only specified in Part II. The major secondary endpoints that are dichotomous endpoints will be analyzed by the Cochran-Mantel-Haenszel chi-square test (2-sided) stratified by baseline CDAI score (≤300 or >300), SNP-positive status (yes or no), and Bio-IR status (yes or no); the major secondary endpoints that are continuous endpoints will be analyzed by an analysis of covariance model on the van der Waerden normal scores with treatment as a fixed factor and appropriate baseline level of the variable, SNP-positive status (yes or no), and Bio-IR status (yes or no) as covariates. The major secondary endpoints will not be controlled for multiplicity.

**Safety Analyses**

Safety will be assessed by analyses of the incidence and type of AEs, serious AEs, AEs considered by the investigator to be reasonably related to study drug, AEs leading to discontinuation of study agent, infections, infusion reactions, and injection-site reactions. Safety assessments will also include analyses of change from baseline in laboratory parameters (hematology and chemistry) and summary of maximum National Cancer Institute Common Terminology Criteria for Adverse Events toxicity grade for postbaseline laboratory values.

#### TIME AND EVENTS SCHEDULE: PART I

**Table 1: Time and Events Schedule, Part I**

| Period | Screening[w] | Main Study phase [x] | | | | | | | | | | | | | Early termination [c] | Final efficacy & safety follow-up [d] |
|---|---|---|---|---|---|---|---|---|---|---|---|---|---|---|---|---|
| Week | -5 weeks | 0 | 2 | 4 | 6 | 8 | 10 | 12 | 14 | 16 | 18 | 20 | 22 | 24 | | |
| **Study Procedures**[aa] | | | | | | | | | | | | | | | | |
| **Screening/Administrative** | | | | | | | | | | | | | | | | |
| Informed consent | X | | | | | | | | | | | | | | | |
| Inclusion/exclusion criteria | X | X | | | | | | | | | | | | | | |
| Medical history and demographics | X | | | | | | | | | | | | | | | |
| Chest radiograph[bb] | X | | | | | | | | | | | | | | | |
| QuantiFERON-TB Test [cc] | X | | | | | | | | | | | | | | | |
| Stool studies to evaluate for enteric pathogens [dd] | X | | | | | | | | | | | | | | | |
| Training on diary card completion | X | | | | | | | | | | | | | | | |
| **Study Agent Administration** | | | | | | | | | | | | | | | | |
| Randomization | | X | | | | | | | | | | | | | | |
| Administer study agent [aa] (SC study agent at a different location at each visit [Section 6]) | | X | X | X | X | X[e][e] | X | X | X | X | X | X | X | | | |
| **Safety Assessments** | | | | | | | | | | | | | | | | |
| Physical examination | X | | | | | | | | | | | | | | X | X |
| Weight | X | X | X | X | X | X | X | X | X | X | X | X | X | X | X | X |
| Height | X | | | | | | | | | | | | | | | |
| Vital signs [ff] | X | X | X | X | X | X | X | X | X | X | X | X | X | X | X | X |
| Baseline 12-lead ECG | X | | | | | | | | | | | | | | | |
| Serum pregnancy test | X | | | | | | | | | | | | | | | |
| Urine pregnancy test [gg] | | X | X | X | X | X | X | X | X | X | X | X | X | X | X | X |
| TB evaluation [hh]/other infection assessment | | X | X | X | X | X | X | X | X | X | X | X | X | X | X | X |
| Injection-site evaluation | | X | X | X | X | X | X | X | X | X | X | X | X | X | X | |
| **Efficacy Assessments** | | | | | | | | | | | | | | | | |
| Collect and review diary cards [ii] | | X | X | X | X | X | X | X | X | X | X | X | X | X | X | X |
| CDAI assessments [jj] | | X | X | X | X | X | X | X | X | X | X | X | X | X | X | X |

**Table 1: Time and Events Schedule, Part I**

| Period | Screening[w] | Main Study phase [x] | | | | | | | | | | | | | Early termination [c] | Final efficacy & safety follow-up [d] |
|---|---|---|---|---|---|---|---|---|---|---|---|---|---|---|---|---|
| **Week** | **-5 weeks** | **0** | **2** | **4** | **6** | **8** | **10** | **12** | **14** | **16** | **18** | **20** | **22** | **24** | | |
| **Study Procedures**[aa] | | | | | | | | | | | | | | | | |
| Fistula exam | | X | X | X | X | X | X | X | X | X | X | X | X | X | X | X |
| Bristol stool form scale [kk] | | X | X | X | X | X | X | X | | | | | | | | |
| Abdominal pain NRS assessment [ll] | | X | X | X | X | X | X | X | X | X | X | X | X | X | X | X |
| Stool sample (fecal lactoferrin and calprotectin) and microbiome | X [mm] | | X | | | X | | X [mm] | | | | | | X [mm] | X | |
| CRP | X | X | X | X | X | X | | X | | X | | X | | X | X | X |
| Video ileocolonoscopy [nn] | X | | | | | | | X | | | | | | X[oo] | X | |
| IBDQ [pp] | | X | | | | X | | X | | | | | | X | X | |
| SF-36 [pp] | | X | | | | X | | X | | | | | | X | X | |
| **Clinical Laboratory Assessments** | | | | | | | | | | | | | | | | |
| Hematology | X | G[Q] | X | X | X | X | X | X | X | X | X | X | X | X | X | X |
| Chemistry | X | ,[q] | | X | | X | | X | | X | | X | | X | X | |
| HBV and HCV testing | X | | | | | | | | | | | | | | | |
| HIV test | X | | | | | | | | | | | | | | | |
| **Pharmacokinetics/ Immunogenicity** | | | | | | | | | | | | | | | | |
| JNJ-64304500 serum concentration [rr, ss] | | X | X | X | X | X | X | X | | X | | X | | X | X | X |
| JNJ-64304500 random population PK sample [tt, ss] |  | ----X---- |  |  |  |  |  |  |  |  |  |  | ----X---- |  |  |  |
| Antibodies to JNJ-64304500 [rr, ss] | | X | X | X | | X | | X | | | | | | X | X | X |
| Receptor occupancy | | X | X | X | | X | | X | | X | | | | X | X | |
| Immunophenotyping (NK cells and CD8+ T cells) | | X | X | X | | X | | X | | X | | | | X | X | |
| **Pharmacodynamics/Biomarkers** | | | | | | | | | | | | | | | | |
| Blood sample collection (T-cell receptor repertoire) | | X | X | | | X | | X | | | | | | X | X | |
| Whole blood sample collection for RNA analysis [uu] | | X | X | | | X | | X | | | | | | X | X | |
| Serum proteins | | X | X | | | X | | X | | | | | | X | X | |
| Ileocolonoscopy biopsy sample | X | | | | | | | X | | | | | | X[oo] | X | |

**Table 1: Time and Events Schedule, Part I**

| Period | Screening[w] | Main Study phase [x] | | | | | | | | | | | | | Early termination [c] | Final efficacy & safety follow-up [d] |
|---|---|---|---|---|---|---|---|---|---|---|---|---|---|---|---|---|
| Week | -5 weeks | 0 | 2 | 4 | 6 | 8 | 10 | 12 | 14 | 16 | 18 | 20 | 22 | 24 | | |
| **Study Procedures[aa]** | | | | | | | | | | | | | | | | |
| collections for RNA and histology | | | | | | | | | | | | | | | | |
| **Pharmacogenomics (DNA)** | | | | | | | | | | | | | | | | |
| Whole blood sample collection for SNPs and DNA analysis [vv] | X | | | | | | | | | | | | | | | |
| **Ongoing Subject Review** | | | | | | | | | | | | | | | | |
| Concomitant therapy | X | X | X | X | X | X | X | X | X | X | X | X | X | X | X | X |
| Adverse events | X | X | X | X | X | X | X | X | X | X | X | X | X | X | X | X |
| Crohn's disease-related hospitalizations and surgeries [ww] | X | X | X | X | X | X | X | X | X | X | X | X | X | X | X | X |

Abbreviations: CDAI=Crohn's Disease Activity Index; CRP=C-reactive protein; ECG=electrocardiogram; HBV=hepatitis B virus; HCV=hepatitis C virus; HIV=human immunodeficiency virus; IBDQ=Inflammatory Bowel Disease Questionnaire; NK=natural killer; NKG2D=natural killer group 2 member D; PK=pharmacokinetics; PRO=patient-reported outcome; SF-36=36-Item Short Form Health Survey; SNP=single nucleotide polymorphism; TB=tuberculosis.

w. Screening period should be a minimum of 7 days to allow for collection of CDAI data. Subjects who are rescreened do not need to have a minimum of 7 days, provided that they have enough data to support CDAI calculation.
x. Visit windows should be ± 4 days for each visit.
y. Subjects who terminate study participation should complete an early termination visit. If a subject completed assessments at Week 24 and terminated after Week 24, the only assessments that should be performed at the early termination visit are those planned for the final efficacy and safety follow-up. Subjects who discontinue study agent administration before Week 8 (but have not terminated study participation) should complete the Week 8 and Week 12 assessments and return for a final efficacy and safety follow-up visit approximately 16 weeks after their last study agent administration. Subjects who discontinue study agent administration after Week 8 (but have not terminated study participation) should complete the Week 12 and Week 24 assessments and return for a final efficacy and safety follow-up visit approximately 16 weeks after their last study agent administration.
z. Subjects should complete a final efficacy and safety follow-up visit approximately 16 weeks after their last study agent administration. For subjects who complete all visits, this will occur at Week 38.
aa. All assessments are to be completed before study agent administration, unless otherwise specified. It is recommended that PRO assessments be completed first, starting with the PRO components of the CDAI.
bb. Chest radiograph (posterior-anterior and lateral views) may be obtained within 3 months before the Week 0 visit.
cc. All subjects will undergo QuantiFERON-TB testing. In countries where the QuantiFERON-TB test is not registered/approved, TB skin testing will also be required (recommended but not required for study centers in countries where tuberculin is not available). The QuantiFERON-TB test is not required at screening for subjects with a history of latent TB and appropriate treatment (as described in Inclusion Criterion 11a).

dd. Stool studies for enteric pathogens may be performed at either the central or a local laboratory and must include a stool culture and *Clostridium difficile* toxin assay. These must have been performed within 4 months before Week 0. Additional testing, such as ova and parasites or *Escherichia coli* O157:H7 assessment, may be performed at the investigator's clinical discretion.
ee. Administration of study agent at the Week 8 visit must occur after all other procedures and assessments specified for the Week 8 visit have been completed.
ff. Temperature, pulse/heart rate, respiratory rate, and blood pressure.
gg. Must be performed before every study agent administration in female subjects of childbearing potential.
hh. If TB is suspected at any time during the study, a chest radiograph and QuantiFERON-TB test should be performed. In countries where the QuantiFERON-TB test is not registered/approved, TB skin testing should also be performed (recommended but not required for study centers in countries where tuberculin is not available).
ii. For all visits up to Week 24, daily diary information should be collected for every day prior to each visit. For the final efficacy and safety visit, daily diary information should be collected for 14 days prior to the visit.
jj. The hematocrit value obtained during screening will be used to calculate CDAI at Week 0. For other visits, the most recent hematocrit value obtained before a visit will be used to calculate the CDAI for that visit.
kk. Subject will complete the Bristol stool form scale as a daily diary entry and bring the diary to each visit. Daily diary information should be collected for every day prior to each visit.
ll. Subject will complete the abdominal pain NRS assessment as a daily diary entry and bring the diary to each visit. For all visits up to Week 24, daily diary information should be collected for every day prior to the visits. For the final efficacy and safety visit, daily diary information should be collected for 14 days prior to the visit.
mm. Stool samples required for this visit should be obtained before the start of the bowel preparation for the video ileocolonoscopy that is also scheduled for the visit.
nn. Because these procedures must not interfere with the collection of CDAI data, if performed on the day of these visits, the 7 days before the initiation of the colonoscopy preparation should be used to calculate CDAI scores for these visits. If the video ileocolonoscopic examination is not performed on the day of the visit, it must be performed at least 8 days before the Week 0 visit and no more than 8 days before the Week 12 visit and Week 24 visit.
oo. The Week 24 video ileocolonoscopy and related endoscopic assessments are optional.
pp. The IBDQ and SF-36 survey should be administered before any clinical procedures or tests are performed.
qq. Laboratory tests at Week 0 are not required if screening laboratory tests were performed within 2 weeks before the Week 0 visit.
rr. Blood samples should be collected before SC injection of study agent. All reasonable attempts should be made to collect samples at the scheduled timepoints and record the actual times of PK sample collections.
ss. At visits where only serum concentration of study agent will be evaluated (ie, no antibodies to study agent will be evaluated), 1 venous blood sample of sufficient volume should be collected, and each serum sample should be divided into 2 aliquots (1 for serum concentration of study agent, and a back-up). At visits where serum concentration of study agent and antibodies to study agent will be evaluated, 1 venous blood sample of sufficient volume should be collected. Each serum sample will be divided into 3 aliquots (1 each for serum concentration of study agent, antibodies to study agent, and a back-up).
tt. Each random population PK sample should be collected at any time between Weeks 0 and 2 (after the Week 0 study agent administration) and between Weeks 22 and 24 (after the Week 22 study agent administration). The date for the random population PK sample collection will be at the subject's discretion. However, samples should not be drawn on or within 1 day of a scheduled visit (ie, the Week 0, 2, 22, or 24 visit) in this time period.
uu. Whole blood for RNA analysis will be collected from all subjects in the study.
vv. Whole blood for SNP analysis will be collected from all subjects. All subjects will be tested for the NKG2D SNP rs2255336 and the MICB (NKG2D ligand) SNP rs2239705 at screening. For subjects who have signed a separate informed consent form, DNA testing will be done to search for links of specific genes to disease or response.
ww. Hospitalization for ileocolonoscopy is not included in this category.

#### TIME AND EVENTS SCHEDULE: PART II MAIN STUDY PHASE

**Table 2: Time and Events Schedule, Part II**

| Period | Screening[a] | Main study phase[b] | | | | | | | | | | | Early termination [c] | Final efficacy & safety follow-up [d,f] |
|---|---|---|---|---|---|---|---|---|---|---|---|---|---|---|
| **Week** | **-5 weeks** | **0** | **2** | **4** | **6** | **8** | **10** | **12** | **14** | **16** | **20** | **24** | | |
| **Study Procedures[e]** | | | | | | | | | | | | | | |
| **Screening/Administrative** | | | | | | | | | | | | | | |
| Informed consent | X | | | | | | | | | | | | | |
| Inclusion/exclusion criteria | X | X | | | | | | | | | | | | |
| Medical history and demographics | X | | | | | | | | | | | | | |
| Chest radiograph [g] | X | | | | | | | | | | | | | |
| QuantiFERON-TB Test [h] | X | | | | | | | | | | | | | |
| Stool studies to evaluate for enteric pathogen [i] | X | | | | | | | | | | | | | |
| Training on diary card completion | X | | | | | | | | | | | | | |
| **Study Drug Administration** | | | | | | | | | | | | | | |
| Randomization | | X | | | | | | | | | | | | |
| Administer study agent [e] (SC study agent at a different location at each visit [Section 6]) | | X | X | X | | X | | X | X | X | X | | | |
| **Safety Assessments** | | | | | | | | | | | | | | |
| Physical examination | X | | | | | | | | | | | | X | X |
| Weight | X | X | X | X | X | X | X | X | X | X | X | X | X | X |
| Height | X | | | | | | | | | | | | | |
| Vital signs [j] | X | X [k] | X | X | | X | | X | X | X | X | X | X | X |
| Baseline 12-lead ECG | X | | | | | | | | | | | | | |
| Serum pregnancy test | X | | | | | | | | | | | | | |
| Urine pregnancy test [l] | | X | X | X | | X | | X | X | X | X | | X | X |
| TB evaluation [m]/other infection assessment | | X | X | X | X | X | X | X | X | X | X | X | X | X |
| Injection-site evaluation | | X | X | X | X | X | X | X | X | X | X | | X | |
| **Efficacy Assessments** | | | | | | | | | | | | | | |
| Collect and review diary cards [n] | | X | X | X | X | X | X | X | X | X | X | X | X | X |
| CDAI assessments [o] | | X | X | X | X | X | X | X | X | X | X | X | X | X |
| Fistula exam | | X | X | X | X | X | X | X | X | X | X | X | X | X |

**Table 2: Time and Events Schedule, Part II**

| Period | Screening[a] | Main study phase[b] | | | | | | | | | | | Early termination [c] | Final efficacy & safety follow-up [d,f] |
|---|---|---|---|---|---|---|---|---|---|---|---|---|---|---|
| **Week** | **-5 weeks** | **0** | **2** | **4** | **6** | **8** | **10** | **12** | **14** | **16** | **20** | **24** | | |
| **Study Procedures[e]** | | | | | | | | | | | | | | |
| Bristol stool form scale [p] | | X | X | X | X | X | X | X | | | | | | |
| Abdominal pain NRS assessment [q] | | X | X | X | X | X | X | X | X | X | X | X | X | X |
| PGIS of Crohn's disease assessment | | X | | X | | X | | X | | | | X | X | |
| PGIC of severity of Crohn's disease assessment | | | | X | | X | | X | | | | X | X | |
| Stool sample (fecal lactoferrin and calprotectin) and microbiome | X [r] | | X | | | X | | X [r] | | | | X [r] | X | |
| CRP | X | X | X | X | X | X | | X | | X | X | X | X | **X** |
| Video ileocolonoscopy [s] | X | | | | | | | X | | | | | X | |
| IBDQ [t] | | X | | | | X | | X | | | | X | X | |
| SF-36 [t] | | X | | | | X | | X | | | | X | X | |
| **Clinical Laboratory Assessments** | | | | | | | | | | | | | | |
| Hematology | X | X[u] | X | X | X | X | X | X | X | X | X | X | X | X |
| Chemistry | X | X[u] | | X | | X | | X | | X | X | X | X | |
| HBV and HCV testing | X | | | | | | | | | | | | | |
| HIV test | X | | | | | | | | | | | | | |
| **Pharmacokinetics/ Immunogenicity** | | | | | | | | | | | | | | |
| Study agent serum concentration[v, w] | | X | X | X | X | X | X | X | | X | X | X | X | X |
| JNJ-64304500 random population PK sample[x, w] |  | ----X---- |  |  |  |  |  |  |  |  | ----X---- |  |  |  |
| Assessment for antibody to study agent [v, w] | | X | X | X | | X | | X | | | | X | X | X |
| Receptor occupancy | | X | X | X | | X | | X | | X | | X | X | |
| Immunophenotyping (NK cells and CD8+ T cells) | | X | X | X | | X | | X | | X | | X | X | |

**Table 2: Time and Events Schedule, Part II**

| Period | Screening[a] | Main study phase[b] | | | | | | | | | | | Early termination [c] | Final efficacy & safety follow-up [d,f] |
|---|---|---|---|---|---|---|---|---|---|---|---|---|---|---|
| **Week** | **-5 weeks** | **0** | **2** | **4** | **6** | **8** | **10** | **12** | **14** | **16** | **20** | **24** | | |
| **Study Procedures[e]** | | | | | | | | | | | | | | |
| **Pharmacodynamics/Biomarkers** | | | | | | | | | | | | | | |
| Blood sample collection (T-cell receptor repertoire) | | X | X | | | X | | X | | | | X | X | |
| Whole blood sample collection for RNA analysis [y] | | X | X | | | X | | X | | | | X | X | |
| Serum proteins | | X | X | | | X | | X | | | | X | X | |
| Ileocolonoscopy biopsy sample collections for RNA and histology | X | | | | | | | X | | | | | X | |
| **Pharmacogenomics (DNA)** | | | | | | | | | | | | | | |
| Whole blood sample collection for SNPs and DNA analysis [z] | X | | | | | | | | | | | | | |
| **Ongoing Subject Review** | | | | | | | | | | | | | | |
| Concomitant therapy | X | X | X | X | X | X | X | X | X | X | X | X | X | X |
| Adverse events | X | X | X | X | X | X | X | X | X | X | X | X | X | X |
| Crohn's disease-related hospitalizations and surgeries [aa] | X | X | X | X | X | X | X | X | X | X | X | X | X | X |

Abbreviations: CDAI=Crohn's Disease Activity Index; CRP=C-reactive protein; ECG=electrocardiogram; HBV=hepatitis B virus; HCV=hepatitis C virus; HIV=human immunodeficiency virus; IBDQ=Inflammatory Bowel Disease Questionnaire; NK=natural killer; NKG2D=natural killer group 2 member D; PGIC=Patient's Global Impression of Change; PGIS=Patient's Global Impression of Severity; PK=pharmacokinetics; PRO=patient-reported outcome; SF-36=36-Item Short Form Health Survey; SNP=single nucleotide polymorphism; TB=tuberculosis.

a. Screening period should be a minimum of 7 days to allow for collection of CDAI data. Subjects who are rescreened do not need to have a minimum of 7 days, provided that they have enough data to support CDAI calculation.
b. Visit windows should be ±4 days for each visit up to and including Week 12; from Week 16 to end of study, visit window should be ± 7 days.
c. Subjects who terminate study participation should complete an early termination visit. If a subject completed assessments at Week 24 and terminated after Week 24, the only assessments that should be performed at the early termination visit are those planned for the final efficacy and safety follow-up visit. Subjects who discontinue study agent administration before Week 12 (but have not terminated study participation) should complete the Week 12 assessments and return for a final efficacy and safety follow-up visit approximately 16 weeks after their last study agent administration. Subjects who discontinue study agent administration after Week 12 (but have not terminated study participation) should complete the Week 24 assessments and return for a final efficacy and safety follow-up visit approximately 16 weeks after their last study agent administration.
d. Subjects should complete a final efficacy and safety follow-up visit approximately 16 weeks after their last study agent administration. For subjects who complete all visits and are not enrolling in the Part II LTE, this will occur at Week 36.

e. All assessments are to be completed before study agent administration, unless otherwise specified. It is recommended that PRO assessments be completed first, starting with the PRO components of the CDAI.
f. The final efficacy and safety follow-up visit in Part II will only occur for those subjects not enrolling into the Part II LTE.
g. Chest radiograph (posterior-anterior and lateral views) may be obtained within 3 months before the Week 0 visit.
h. All subjects will undergo QuantiFERON-TB testing. In countries where the QuantiFERON-TB test is not registered/approved, TB skin testing will also be required (recommended but not required for study centers in countries where tuberculin is not available). The QuantiFERON-TB test is not required at screening for subjects with a history of latent TB and appropriate treatment (as described in Inclusion Criterion 11a).
i. Stool studies for enteric pathogens may be performed at either the central or a local laboratory and must include a stool culture and *Clostridium difficile* toxin assay. These must have been performed within 4 months before Week 0. Additional testing, such as ova and parasites or *Escherichia coli* O157:H7 assessment, may be performed at the investigator's clinical discretion.
j. Temperature, pulse/heart rate, respiratory rate, and blood pressure.
k. At Week 0, vital signs must be obtained before the IV infusion, approximately every 30 minutes during the infusion, and twice (at approximately 30-minute intervals) after completion of the infusion.
l. Must be performed before every study agent administration in female subjects of childbearing potential.
m. If TB is suspected at any time during the study, a chest radiograph and QuantiFERON-TB test should be performed. In countries where the QuantiFERON-TB test is not registered/approved, TB skin testing should also be performed (recommended but not required for study centers in countries where tuberculin is not available).
n. For all visits up to Week 24, daily diary information should be collected for every day prior to each visit. For the final efficacy and safety visit, daily diary information should be collected for 14 days prior to the visit.
o. The hematocrit value obtained during screening will be used to calculate CDAI at Week 0. For other visits, the most recent hematocrit value obtained before a visit will be used to calculate the CDAI for that visit.
p. Subject will complete the Bristol stool form scale as a daily diary entry and bring the diary to each visit. Daily diary information should be collected for every day prior to each visit.
q. Subject will complete the abdominal pain NRS assessment as a daily diary entry and bring the diary to each visit. For all visits up to Week 24, daily diary information should be collected for every day prior to the visits. For the final efficacy and safety visit, daily diary information should be collected for 14 days prior to the visit.
r. Stool samples required for this visit should be obtained before the start of the bowel preparation for the video ileocolonoscopy that is also scheduled for the visit.
s. Because these procedures must not interfere with the collection of CDAI data, if performed on the day of these visits, the 7 days before the initiation of the colonoscopy preparation should be used to calculate CDAI scores for these visits. If the video ileocolonoscopic examination is not performed on the day of the visit, it must be performed at least 8 days before the Week 0 visit and no more than 8 days before the Week 12 visit and Week 24 visit.
t. The IBDQ and SF-36 survey should be administered before any clinical procedures or tests are performed.
u. Laboratory tests at Week 0 are not required if screening laboratory tests were performed within 2 weeks before the Week 0 visit.
v. At all visits when study intervention will be administered, 1 blood sample should be collected prior to study intervention administration for evaluation of serum concentrations and/or antibodies to the study interventions. For the IV infusion related visit (Week 0), the SC study intervention should be administered first, followed by IV infusion; another blood draw should be taken approximately 60 minutes after completion of the infusion for serum concentration measurement.
w. At visits where only serum concentration of study agent will be evaluated (ie, no antibodies to study agent will be evaluated), 1 venous blood sample of sufficient volume should be collected, and each serum sample should be divided into 2 aliquots (1 for serum concentration of study agent, and a back-up). At

visits where serum concentration of study agent and antibodies to study agent will be evaluated, 1 venous blood sample of sufficient volume should be collected. Each serum sample will be divided into 3 aliquots (1 each for serum concentration of study agent, antibodies to study agent, and a back-up).

x. Each random population PK sample should be collected at any time between Weeks 0 and 2 (after the Week 0 study agent administration) and between Weeks 20 and 24 (after the Week 20 study agent administration). The date for the random population PK sample collection will be at the subject's discretion. However, samples should not be drawn on or within 1 day of a scheduled visit (ie, the Week 0, 2, 20, or 24 visit) in this time period.

y. Whole blood for RNA analysis will be collected from all subjects in the study.

z. Whole blood for SNP analysis will be collected from all subjects. All subjects will be tested for the NKG2D SNP rs2255336 and the MICB (NKG2D ligand) SNP rs2239705 at screening. For subjects who have signed a separate informed consent form, DNA testing will be done to search for links of specific genes to disease or response.

aa. Hospitalization for ileocolonoscopy is not included in this category.

#### TIME AND EVENTS SCHEDULE: PART II LONG-TERM EXTENSION

**Table 3: Time and Events Schedule, Long-term Extension Before Unblinding for JNJ-64304500, Ustekinumab, and Placebo and After Unblinding for JNJ-64304500[j]**

| Period | Long-term Extension | | | | | | |
|---|---|---|---|---|---|---|---|
| | Week 24[a,o] | Every 4 weeks[a,l] | Every 3 months[a,m] | Every 6 months[a,n] | Week 72[a] | Final safety follow-up[a,d] | Early termination[a] |
| **Study Procedures** | | | | | | | |
| **Study Drug Administration** | | | | | | | |
| Administer study agent (SC study agent at a different location at each visit [Section 6])[b,c] | X | X[i] | | | | | |
| **Safety Assessments** | | | | | | | |
| Symptom-driven physical examination | X | X | | | X | X | X |
| Vital signs[e] | X | X | | | X | X | X |
| Urine pregnancy test[f] | X | X | | | X | X | X |
| TB evaluation /other infection assessment[g] | | X | | | X | X | X |
| Injection-site evaluation | X | X | | | X | X | X |
| **Clinical Laboratory Assessments** | | | | | | | |
| Hematology | | | X | | X | X | X |
| Chemistry | | | | X | X | X | X |
| **Pharmacokinetics/ Immunogenicity[k]** | | | | | | | |
| Study agent serum concentration[h] | | | | | X | X | X |
| Assessment for antibody to study agent | | | | | X | X | X |
| **Ongoing Subject Review** | | | | | | | |
| Concomitant therapy | | X | | | X | X | X |
| Adverse events | X | X | | | X | X | X |
| Crohn's disease-related hospitalizations and surgeries | | X | | | X | X | X |

a. Visit window is ±7days for each visit.
b. All assessments are to be completed before study agent administration unless otherwise specified.
c. The first study agent administration in the Part II LTE will occur at Week 24. After the main study is unblinded to the investigative sites, subjects in the Part II LTE receiving placebo will be terminated from study participation, and subjects receiving ustekinumab will receive ustekinumab until commercial ustekinumab becomes available to them. Subjects receiving JNJ-64304500 will continue to receive JNJ-64304500 every 4 weeks. Those subjects still receiving ustekinumab will return to the clinic every 8 weeks to receive drug for SC ustekinumab injections.
d. Final safety visit at Week 88 or approximately 16 weeks after a subject's last administration of study agent (for subjects who have not terminated study participation). If a subject's treatment allocation is unblinded while the LTE is ongoing and the subject was receiving placebo, the site may schedule the final safety visit to occur earlier than 16 weeks after the subject's last administration of study drug.
e. Temperature, pulse/heart rate, respiratory rate, and blood pressure. At a study agent administration visit, vital signs should be obtained before and approximately 30 minutes after SC injection.

f. Urine pregnancy test must be performed before each study agent administration in female subjects of childbearing potential.
g. If TB is suspected at any time, a chest radiograph and QuantiFERON-TB test should be performed. In countries where the QuantiFERON is not registered/approved, TB skin testing should also be performed (recommended but not required for study centers in Ukraine if tuberculin is not available).
h. Serum concentration of study agent and antibodies to study agent will be evaluated. One venous blood sample of sufficient volume should be collected (at visits where study intervention will be administered, blood samples should be collected prior to study intervention administration). Each serum sample will be divided into 3 aliquots (1 each for serum concentration of study agent, antibodies to study agent and a back-up).
i. JNJ-64304500 dosing only after unblinding.
j. This Time & Events Schedule outlines the tasks that need to be completed during the Part II LTE prior to unblinding for all study subjects and after unblinding for study subjects receiving JNJ-64304500. Table 4 outlines the tasks that need to be completed during the Part II LTE after study unblinding for study subjects receiving ustekinumab in countries where it is not commercially available.
k. Pharmacokinetic and immunogenicity samples will be obtained only at the specified timepoints and when SAEs are reported by study subjects.
l. Visits every 4 weeks include scheduled visits at Weeks 28, 32, 36, 40, 44, 48, 52 56, 60, 64, 68, and 72.
m. Visits every 3 months include scheduled visits at Weeks 36, 48, 60, and 72.
n. Visits every 6 months include scheduled visits at Weeks 48 and 72.
o. Subjects may receive the first LTE dose of study drug at the Week 24 visit after the Week 24 assessments are completed or the subject may return to the study site for the first LTE does of study drug within the 7-day visit window.

#### TIME AND EVENTS SCHEDULE: PART II LONG-TERM EXTENSION

**Table 4: Time and Events Schedule, Long-term Extension Phase After Unblinding in Countries Where Ustekinumab Is Not Commercially Available**

| **Period** | **Long-term Extension** | | | |
|---|---|---|---|---|
| | **Every 8 Weeks[a,j]** | **Every 16 Weeks[a,k]** | **Final safety follow-up[a,d]** | **Early termination[a]** |
| **Study Procedures** | | | | |
| **Study Drug Administration** | | | | |
| Administer study agent[b,c] | X | | | |
| **Safety Assessments** | | | | |
| Symptom-driven physical examination | X | | X | X |
| Vital signs[e] | X | | X | X |
| Urine pregnancy test[f] | X | | X | X |
| TB evaluation /other infection assessment[g] | X | | X | X |
| Injection-site evaluation | X | | X | X |
| **Clinical Laboratory Assessments** | | | | |
| Hematology | | X | X | X |
| Chemistry | | X | X | X |
| **Pharmacokinetics/ Immunogenicity[i]** | | | | |
| Study agent serum concentration[h] | X | | X | X |
| Assessment for antibody to study agent | | | X | X |
| **Ongoing Subject Review** | | | | |
| Concomitant therapy | X | | X | X |
| Adverse events | X | | X | X |
| Crohn's disease-related hospitalizations and surgeries | X | | X | X |

a. Visit window is ±7 days for each visit.
b. All assessments are to be completed before study agent administration unless otherwise specified.
c. The first study agent administration in the Part II LTE will occur at Week 24. After the main study is unblinded to the investigative sites, subjects in the Part II LTE receiving placebo will be terminated from study participation, and subjects receiving ustekinumab will return to the clinic every 8 weeks for SC ustekinumab injections.
d. The final safety follow-up visit should occur after unblinding but before receiving the first dose of commercial ustekinumab.
e. Temperature, pulse/heart rate, respiratory rate, and blood pressure. At a study agent administration visit, vital signs should be obtained before and approximately 30 minutes after SC injection.
f. Urine pregnancy test must be performed before each study agent administration in female subjects of childbearing potential.
g. If TB is suspected at any time, a chest radiograph and QuantiFERON-TB test should be performed. In countries where the QuantiFERON is not registered/approved, TB skin testing should also be performed (recommended but not required for study centers in Ukraine if tuberculin is not available).
h. Serum concentration of study agent and antibodies to study agent will be evaluated. One venous blood sample of sufficient volume should be collected (at visits where study intervention will be administered, blood samples should be collected prior to study intervention administration). Each serum sample will be divided into 3 aliquots (1 each for serum concentration of study agent, antibodies to study agent and a back-up).

i. Pharmacokinetic and immunogenicity samples will be obtained only at the specified timepoints and when SAEs are reported by study subjects.
j. Visits every 8 weeks may include scheduled visits at Weeks 32, 40, 48, 56, 64, and 72, depending on when unblinding occurs.
k. Visits every 16 weeks may include scheduled visits at Weeks 40, 56, and 72, depending on when unblinding occurs.

### ABBREVIATIONS

| | |
|---|---|
| 5-ASA | 5-aminosalicylic acid |
| 6-MP | 6-mercaptopurine |
| 6-TG | 6-thioguanine |
| ADL | Activities of Daily Living |
| AE | adverse event |
| ALT | alanine aminotransferase |
| ANCOVA | analysis of covariance |
| anti-HBc | HBV core antibody |
| anti-HBs | HBV surface antibody |
| AST | aspartate aminotransferase |
| AUC | area under the serum concentration–time curve |
| AZA | azathioprine |
| BCG | Bacille Calmette-Guérin |
| β-hCG | β-human chorionic gonadotropin |
| Bio-IR | biologic intolerant or refractory |
| Bio-NF | biologic nonfailure, ie, inadequate response to or failed to tolerate corticosteroids or immunomodulators, but not a biologic |
| CDAI | Crohn's Disease Activity Index |
| $C_{max}$ | maximum concentration |
| CRF | case report form (paper or electronic, as appropriate for this study) |
| CRP | C-reactive protein |
| DBL | database lock |
| DMC | Data Monitoring Committee |
| DNA | deoxyribonucleic acid |
| ECG | electrocardiogram |
| eDC | electronic data capture |
| FDA | Food and Drug Administration |
| GCP | Good Clinical Practice |
| GHAS | Global Histology Activity Score |
| HBsAg | hepatitis B surface antigen |
| HBV | hepatitis B virus |
| HCV | hepatitis C virus |
| HIV | human immunodeficiency virus |
| IB | Investigator's Brochure |
| IBD | inflammatory bowel disease |
| IBDQ | Inflammatory Bowel Disease Questionnaire |
| ICF | informed consent form |
| ICH | International Conference on Harmonisation |
| IEC | Independent Ethics Committee |
| IL | interleukin |
| IRB | Institutional Review Board |
| IUD | intrauterine device |
| IUS | intrauterine hormone-releasing system |
| IV | intravenous |
| IWRS | interactive web response system |
| JAK | Janus kinase |
| LTE | long-term extension |
| MCP-Mod | Multiple Comparison Procedures with modeling |
| MCS | Mental Component Summary |
| MICA | major histocompatibility class I chain-related protein A |
| MICB | major histocompatibility class I chain-related protein B |
| MTX | methotrexate |
| NCI-CTCAE | National Cancer Institute Common Terminology Criteria for Adverse Events |
| NK | natural killer |
| NKG2D | natural killer group 2D |

| | |
|---|---|
| NRS | numerical rating scale |
| PCS | Physical Component Summary |
| PD | pharmacodynamic |
| PFS | prefilled syringe |
| PGIC | Patient’s Global Impression of Change |
| PGIS | Patient’s Global Impression of Severity |
| PK | pharmacokinetic(s) |
| PQC | Product Quality Complaint |
| PRO | patient-reported outcome(s) |
| q2w | every 2 weeks |
| q4w | every 4 weeks |
| RA | rheumatoid arthritis |
| RNA | ribonucleic acid |
| RO | receptor occupancy |
| SAE | serious adverse event |
| SAP | Statistical Analysis Plan |
| SC | subcutaneous |
| SD | standard deviation |
| SES-CD | Simple Endoscopic Score for Crohn's Disease |
| SF-36 | 36-item Short Form Health Survey |
| SNP | single nucleotide polymorphism |
| SUSAR | suspected unexpected serious adverse reaction |
| TB | tuberculosis |
| TNF | tumor necrosis factor |
| TNFα | tumor necrosis factor alpha |
| ULN | upper limit of normal |

### 1. INTRODUCTION

The natural killer group 2 member D (NKG2D) is an activating receptor expressed on the surface of natural killer (NK) cells, CD8+ T cells, and subsets of CD4+ T cells, invariant NK T cells, and gamma/delta (γδ) T cells. It signals via the noncovalently associated DNAX-activating protein of 10 kDa and a phosphoinositide 3-kinase. The receptor recognizes an extensive repertoire of self-ligands (ie, ligands encoded by the host's own genome as opposed to foreign antigens), encoded by at least 8 genes including the major histocompatibility class I chain-related proteins A and B (MICA and MICB), and proteins from the ULPB/RAET1 family (ULBP1-6), some of which have extensive allelic polymorphisms. Expression of NKG2D ligands is tightly regulated. All NKG2D ligands are expressed at very low levels in healthy adult tissues; however, cellular stress, cancer, or virus infection can upregulate the expression of these self-encoded NKG2D ligands. Recognition of these self-encoded NKG2D ligands on unhealthy cells by the NKG2D receptor ("induced self-recognition") results in the proliferation and the subsequent release of proinflammatory cytokines and cytotoxic mediators such as interferon gamma, Vav1 guanine nucleotide exchange factor, interleukin (IL)-9, IL-10, vascular endothelial growth factor-A, IL-13, and tumor necrosis factor alpha (TNFα) by these NKG2D receptor-bearing NK and T cells leading to a deoxyribonucleic acid (DNA) damage response and inflammation.

Several lines of evidence from patients with Crohn's disease support the hypothesis that NKG2D receptor activation plays a role in disease pathogenesis by mediating the production of local cytokines, activation of an immune response, and direct cytotoxicity of target intestinal cells. In Crohn's disease, CD4+ and CD8+ T cells expressing the NKG2D receptor accumulate at sites of intestinal inflammation. The level of circulating NKG2D+CD4+ T cells rapidly declines following surgical removal of inflamed intestinal tissue, and their concentration increases at the relapse of disease. Approximately 40% of patients with Crohn's disease have circulating NKG2D+CD4+ T cells, and these cells were found in virtually all samples of inflamed intestine. Expression of NKG2D ligands has been demonstrated to be upregulated in intestinal biopsies in patients with Crohn's disease compared with healthy subjects, further supporting a role for the involvement of the NKG2D receptor in inflammation. In addition, blockade of the NKG2D receptor is effective in preventing disease progression in preclinical animal models of human inflammatory diseases. Collectively, preclinical and clinical data on the expression of NKG2D-ligands or proinflammatory cytokines in the target tissue and abnormal expression and activation of the NKG2D receptor on CD8+ and CD4+ T cells provide a rationale for the clinical development of inhibitors of the NKG2D receptor.

JNJ-64304500 (formerly known as NNC0142-002) is a human immunoglobulin G4 isotype monoclonal antibody that binds specifically to the NKG2D receptor, and blocks NKG2D ligand binding, thereby preventing the downstream-signaling events that otherwise lead to cell proliferation and release of proinflammatory cytokines and cytotoxic mediators. Results from a Phase 2a study in subjects with moderately to severely active Crohn's disease who had failed or were intolerant to conventional therapy or TNFα antagonists for Crohn's disease (conducted by Novo Nordisk) provided evidence for the efficacy of JNJ-64304500 in the treatment of Crohn's disease.

For the most comprehensive nonclinical and clinical information regarding JNJ-64304500, refer to the latest version of the Investigator's Brochure (IB) for JNJ-64304500. The term "sponsor" used throughout this document refers to the entities listed in the Contact Information page(s), which will be provided as a separate document.

#### 1.1. Nonclinical Studies

For the most comprehensive nonclinical information regarding JNJ-64304500, refer to the latest version of the IB for JNJ-64304500.

#### 1.2. Clinical Studies

As of 11 November 2015, a total of 128 subjects had been exposed to JNJ-64304500 in 3 clinical studies: 88 subjects in 2 studies in rheumatoid arthritis (RA) and 40 subjects in a Phase 2a study in Crohn's disease.

##### 1.2.1. Rheumatoid Arthritis

Two studies with JNJ-64304500 were conducted in subjects with active RA.

In **NN8555-3618**, a first-in-humans, Phase 1, single ascending dose/multiple ascending dose study that included single-dose (0.0002 to 7.5 mg/kg) and multiple-dose (0.02 to 4 mg/kg every 2 weeks [q2w] for 4 administrations) parts, 13 dose levels were evaluated in 24 subjects exposed to JNJ-64304500 in the single ascending dose portion and 23 subjects exposed to JNJ-64304500 in the multiple ascending dose portion. Subcutaneous (SC) administration of JNJ-64304500 was well tolerated at the dose ranges investigated and no safety signals were associated with either the single- or multiple-dose regimens.

In **NN8555-3796**, a Phase 2a, randomized, single-dose, double-blind, placebo-controlled, parallel-group study, clinical efficacy was assessed in subjects with active RA concomitantly treated with methotrexate (MTX). In this study, a single SC injection of 4 mg/kg JNJ-64304500 in 41 subjects exposed to JNJ-64304500 did not result in a statistically significant reduction in disease activity at Weeks 6, 12, or 24 after treatment compared with placebo. JNJ-64304500 was well tolerated and no safety concerns were raised during the study.

Additional details about these studies are provided in the latest version of the IB.

##### 1.2.2. Crohn's Disease

**NN8555-3797** was a Phase 2a, multicenter, randomized, double-blind, placebo-controlled, parallel-group, single-dose study in subjects with moderately to severely active Crohn's disease who had failed or were intolerant to conventional therapy (corticosteroids or immunomodulators) or were intolerant or refractory to 1 TNFα antagonist therapy. Only subjects who had a Crohn's Disease Activity Index (CDAI) score ≥220 but ≤450 and inflammation confirmed by CRP ≥10 mg/dL or by endoscopy (endoscopic verification of active ulceration performed during screening and read by a blinded central imaging reader) were included in the study. The study enrolled and randomized 78 subjects at 32 investigational sites in North America, Europe, and Israel. All subjects were randomly assigned in a 1:1 ratio at Week 0 to receive placebo SC (n=38) or 2 mg/kg JNJ-64304500 SC (n=40). Among the 78 randomized subjects, the mean baseline CDAI score was 330.5, and 29.5% were intolerant or refractory to a maximum of 1 TNFα antagonist therapy.

Subjects were evaluated for the primary endpoint of change from baseline in the CDAI score at Week 4. Safety and efficacy evaluations were performed through Week 24. The observed 16-point greater reduction in CDAI in the JNJ-64304500 group at Week 4 compared with the placebo group was not significant (p=0.403). Based on a predefined significance level of 0.10, however, the reduction in CDAI score was significantly higher in the JNJ-64304500 group compared with the placebo group at Week 12 (55-point greater reduction in CDAI was observed in JNJ-64304500 compared with placebo, p=0.056). Based on the same predefined significance level of 0.10, reductions in CDAI scores were significantly higher in the predefined subgroup of "no prior failure to biologics" (Bio-NF, 71% of the study population) in the JNJ-64304500 group compared with placebo at all post baseline visits through Week 12 (Week 1, p=0.068; Week 2, p=0.048; Week 4, p=0.095; Week 8, p=0.015; and Week 12, p=0.025). The biologic intolerant or refractory (Bio-IR) population in the Novo Nordisk Phase 2a study constituted 29% of the study population (N=12 in the active treatment arm and N=11 in the placebo arm). In this subpopulation, a 45-point greater reduction in CDAI was observed at Week 12 in JNJ-64304500 compared with placebo.

As a part of this study, genetic polymorphisms in the genes for the NKG2D receptor and NKG2D ligands of subjects were evaluated. A post hoc analysis of efficacy data demonstrated greater efficacy in a subgroup of subjects with a specific single nucleotide polymorphism (SNP) in the NKG2D receptor and/or MICB ligand (SNP-positive cohort). The association between SNP-positive status and higher clinical efficacy will be tested prospectively in this Phase 2b study.

The mean duration of study participation was equivalent between the 2 treatment groups. No deaths or medical events of special interest were reported. Through Week 24, the proportions of subjects with 1 or more adverse events (AEs) were similar in the JNJ-64304500 and placebo groups. Gastrointestinal events were the most commonly reported AEs in both groups (17 and 14 subjects in the JNJ-64304500 and placebo groups, respectively). Serious AEs (SAEs) were uncommon and reported in 7 of 78 (9%) randomized subjects with 1 SAE each: 2 in the placebo group (1 Crohn's disease, 1 nephrolithiasis) compared with 5 in the JNJ-64304500 group (4 Crohn's disease, 1 *Clostridium difficile* infection). All SAEs were evaluated as unlikely related to treatment with study agent.

Collectively, these data support the further development of JNJ-64304500 in subjects with moderately to severely active Crohn's disease.

This protocol (64304500CRD2001) is comprised of 2 randomized, double-blind, placebo-controlled, parallel-group, multicenter parts designed to evaluate the safety and efficacy of JNJ-64304500 in subjects with moderately to severely active Crohn's disease who have previously failed or who were intolerant to 1 or more approved biologic agents (Bio-IR) or those who have demonstrated an inadequate response to or have failed to tolerate corticosteroids or immunomodulators (Bio-NF).

In Part I, the following 2 studies will be conducted:

- Study 1: A study in subjects who are biologic intolerant or refractory (Bio-IR);
- Study 2: A study in subjects who have not previously failed a biologic therapy (Biologic nonfailure [Bio-NF]).

In Part II, the following study will be conducted:

- Study 3: A dose-ranging study in subjects who are biologic intolerant or refractory (Bio-IR) or Biologic nonfailure (Bio-NF)

In Study 1, a higher dose regimen than was used in the Phase 2a study will be employed supported by the acceptable safety margins demonstrated in Phase 1 (the Novo Nordisk Phase 1 single ascending/multiple ascending dose study NN8555-3618), and the high exposure margins relative to no-observed-adverse-effect level (NOAEL) exposure in cynomolgus monkeys. This higher dose will be used to provide additional information on safety and efficacy for the Bio-IR population, as proof of concept was not established in this population in the Novo Nordisk Phase 2a study (NN8555-3797). In addition, this higher dose will also be evaluated in Bio-NF subjects to determine if a more robust efficacy outcome than was shown in the Phase 2a study can be achieved while maintaining an acceptable safety profile. Combined with data from Phase 2a in Bio-NF subjects, Part I and Part II would provide valuable information for the assessment of dose/exposure-response and thus the selection of the optimal dose in Bio-NF population for the Phase 3 program. Study 1 and Study 2 comprise Part I of the protocol and these 2 components will be initiated in parallel. The decision to initiate the dose-ranging portion of the protocol (Part II, Bio-IR and Bio-NF) is dependent on demonstration of acceptable safety and efficacy in Part I (combined Bio-IR subjects and Bio-NF subjects). A ustekinumab reference arm will be included in Part II to provide context in the interpretation of results from the JNJ-64304500 and placebo groups and to assist in the overall selection of dose(s) to carry forward into the Phase 3 program.

A long-term extension (LTE) for Part II of the study was added to the protocol as part of Protocol Amendment 5. After Protocol Amendment 5 is implemented, subjects who complete Part II Week 24 assessments and who may benefit from continued treatment in the opinion of the investigator, are eligible to enter the Part II LTE to receive up to 52 weeks of additional treatment. Subjects who have completed Week 24 assessments prior to the implementation of Protocol Amendment 5 will not be eligible to enroll in the Part II LTE.

The inclusion of the 3 component studies in a single protocol is intended to facilitate both efficient trial conduct and program decision-making based on planned interim database locks (DBLs) as described in Section 3.1.1, Section 3.1.2, and Section 5.2.

### 2. OBJECTIVES, ENDPOINTS, AND HYPOTHESIS

#### 2.1. Objectives and Endpoints

##### 2.1.1. Objectives

The objectives are the same in each of the 2 parts.

**Primary Objectives**

- To evaluate the efficacy of JNJ-64304500 to reduce the CDAI score from baseline.
- To evaluate the safety of JNJ-64304500.

**Secondary Objectives**

- To evaluate the efficacy of JNJ-64304500 to induce clinical remission, clinical response, and endoscopic healing of the mucosa, and to maintain remission.
- To evaluate the relationship between efficacy and the presence of the NKG2D and/or MICB SNP biomarkers.
- To evaluate the efficacy of JNJ-64304500 to improve general and disease-specific health-related quality of life and to reduce Crohn's disease-related hospitalizations and surgeries.
- To evaluate the pharmacokinetics (PK), immunogenicity, pharmacodynamics (PD), and biomarkers (eg, reductions in CRP, fecal calprotectin, and fecal lactoferrin) of JNJ-64304500 therapy.

##### 2.1.2. Endpoints

The data from Study 1 and Study 2 in Part I will be pooled for analysis.

The primary endpoint for Part I is: Change from baseline in the CDAI score at Week 8.

The primary endpoint for Part II is: Change from baseline in the CDAI score at Week 12.

The following endpoints will be evaluated as major secondary endpoints only in Part II (the dose-ranging portion of the study); these endpoints will be evaluated in Part I but are not specified as major secondary endpoints.

- Clinical remission at Week 12 as measured by CDAI (CDAI <150).
- Clinical response at Week 12 as measured by CDAI (≥100-point reduction from baseline in CDAI or CDAI <150).
- Change in PRO-2 (the sum of the abdominal pain and stool frequency subscores of the CDAI score) from baseline at Week 12.
- Clinical remission at Week 12 as measured by PRO-2 (PRO-2 <75).
- Clinical response at Week 12 as measured by PRO-2 (≥50-point reduction from baseline in PRO-2 or PRO-2 <75).

- Change in Simple Endoscopic Score for Crohn's Disease (SES-CD) from baseline at Week 12.

The following efficacy endpoints will be evaluated in Part I and Part II (except for the Patient's Global Impression of Severity [PGIS] of Crohn's disease and the Patient's Global Impression of Change [PGIC] of severity of Crohn's disease, which will only be evaluated in Part II):

- Change in CDAI from baseline at all postbaseline visits.
- Clinical remission based on CDAI at all postbaseline visits.
- Clinical response based on CDAI at all postbaseline visits.
- Change in PRO-2 from baseline at all postbaseline visits.
- Change in abdominal pain score (daily average based on the CDAI assessment) from baseline at all postbaseline visits.
- Change in stool frequency score (daily average based on the CDAI assessment) from baseline at all postbaseline visits.
- Clinical remission based on PRO-2 at all postbaseline visits.
- Clinical response based on PRO-2 at all postbaseline visits.
- Change in PRO-3 (the sum of abdominal pain, stool frequency, and general well-being subscores of the CDAI score) from baseline at all postbaseline visits.
- Clinical remission based on CDAI at Week 24 among subjects in clinical response at Week 12.
- Clinical remission based on CDAI at Week 24 among subjects in clinical remission at Week 12.
- Change in SES-CD score from baseline at Weeks 12 and 24.
- Endoscopic improvement at Weeks 12 and 24 based on a reduction from baseline in SES-CD score ≥3. Endoscopy will only be performed during Part I.
- At least 50% improvement from baseline in SES-CD at Weeks 12 and 24.
- Endoscopic healing (defined as the absence of mucosal ulcerations) at Weeks 12 and 24.
- Endpoint(s) for histologic assessment based on the Global Histology Activity Score (GHAS; to be detailed in the Statistical Analysis Plan [SAP]).
- Fistula response at all postbaseline visits, defined as a ≥50% reduction from baseline in the number of draining fistulas.
- Endpoint(s) based on Bristol stool form scale (to be detailed in the SAP).
- Change in abdominal pain from baseline at all postbaseline visits based on a 0-10 Numerical Rating Scale (NRS).
- Change in PGIS of Crohn's disease from baseline at Weeks 4, 8, 12, and 24.
- A ≥1-point improvement in PGIS of Crohn's disease from baseline at Weeks 4, 8, 12, and 24.

- Improvement in PGIC of severity of Crohn's disease at Weeks 4, 8, 12, and 24.
- Change in Inflammatory Bowel Disease Questionnaire (IBDQ) score from baseline at Weeks 8, 12, and 24.
- Clinical remission based on IBDQ ($\geq$170) at Weeks 8, 12, and 24.
- A $\geq$16-point improvement in IBDQ from baseline at Weeks 8, 12, and 24.
- Change from baseline in the Physical Component Summary (PCS) and Mental Component Summary (MCS) scores of the 36-item Short Form Health Survey (SF-36) at Weeks 8, 12, and 24.
- A $\geq$5-point improvement in PCS or MCS scores of the SF-36 at Weeks 8, 12, and 24.
- Change in biomarkers (CRP, fecal calprotectin, fecal lactoferrin) from baseline at Weeks 8, 12, and 24.
- Clinical remission based on CDAI at Week 12 by SNP status. Subjects who are positive in at least 1 of 2 SNPs (NKG2D or MICB) will be considered to be SNP-positive.

  Other efficacy endpoints may be examined by SNP status (to be detailed in the SAP).

Refer to Section 9, Study Evaluations, for evaluations related to endpoints.

#### 2.2. Hypothesis

The hypothesis for Part I and Part II is that JNJ-64304500 is superior to placebo in inducing a reduction from baseline in CDAI in subjects with moderately to severely active Crohn's disease.

### 3. STUDY DESIGN AND RATIONALE

#### 3.1. Overview of Study Design

This protocol is comprised of 2 parts (Part I [Bio-IR subjects and Bio-NF subjects] and Part II [Bio-IR and Bio-NF subjects]) (Figure 1) that are designed to evaluate the safety and efficacy of JNJ-64304500 in subjects with moderately to severely active Crohn's disease.

In Part I, the following 2 studies will be conducted:

- Study 1: A study in subjects who are biologic intolerant or refractory (Bio-IR);
- Study 2: A study in subjects who have not previously failed a biologic therapy (Biologic nonfailure [Bio-NF]).

In Part II, the following study will be conducted:

- Study 3: A dose-ranging study in subjects who are Bio-IR or Bio-NF

The 2 studies in Part I serve to build on the original Phase 2a study findings by employing dedicated populations of both Bio-IR and Bio-NF subjects. If acceptable safety and efficacy are established in the combined Bio-IR and Bio-NF populations, Part II of the protocol, which consists of a dose-ranging study (Study 3) in subjects who are Bio-IR or Bio-NF, will be initiated.

After Protocol Amendment 5 is implemented, subjects who complete Part II Week 24 assessments and who may benefit from continued treatment in the opinion of the investigator, are eligible to enter the Part II LTE to receive up to 52 weeks of additional treatment. Subjects who have completed Week 24 prior to the implementation of Protocol Amendment 5 will not be eligible to enroll in the Part II LTE.

**Figure 1: Schematic representation of 64304500CRD2001**

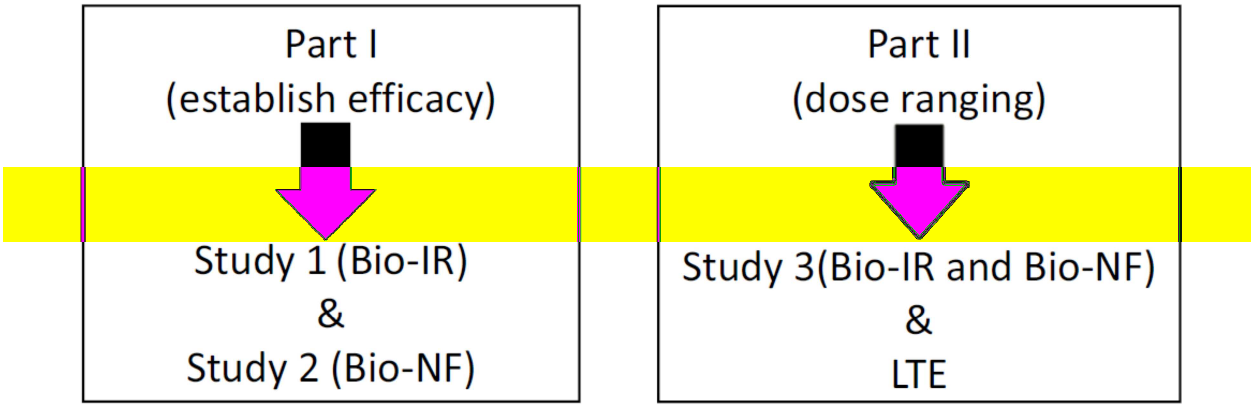

The target population for each of the studies consists of men or women ≥18 years of age with moderately to severely active Crohn's disease (of at least 3 months' duration), defined as a CDAI score ≥220 but ≤450 at Week 0, with elevated CRP >0.3 mg/dL (>3.0 mg/L) and/or calprotectin >250 mg/kg at screening. Subjects must have colitis, ileitis, or ileocolitis previously confirmed at any time in the past by radiography, histology, and/or endoscopy.

Additionally, subjects must have previously failed or been intolerant to 1 or more approved biologic agents (ie, TNFα-antagonists or vedolizumab, hereafter referred to as Bio-IR subjects) **or** have demonstrated an inadequate response to or failed to tolerate corticosteroids or immunomodulators (ie, 6-mercaptopurine [6-MP], azathioprine [AZA], and MTX) but not a biologic agent (hereafter referred to as biologic nonfailure subjects). These 2 populations are described below:

- **Biologic intolerant or refractory (Bio-IR) subjects** are defined as those who have received infliximab (or a biosimilar for infliximab), adalimumab (or a biosimilar for adalimumab), certolizumab pegol, or vedolizumab at a dose approved for the treatment of Crohn's disease, and either did not respond initially, responded initially but then lost response, or were intolerant to the medication (Attachment 1). Bio-IR subjects must allow a ≥8-week washout for prior TNFα antagonist use and a 16-week washout period for prior vedolizumab use.

- **Biologic nonfailure (Bio-NF) subjects** are defined as those who have demonstrated an inadequate response to or have failed to tolerate corticosteroids or the immunomodulators 6-MP, AZA, or MTX (Attachment 2). Subjects who have demonstrated corticosteroid dependence (ie, an inability to successfully taper corticosteroids without a return of the symptoms of Crohn's disease) are also eligible. Bio-NF subjects may also have received biologic therapy but only if it was discontinued for reasons other than lack of efficacy or intolerance (eg, drug holiday).

It is anticipated that approximately 370 to 420 subjects will be randomized overall across Part I and Part II:

- **Part I** will study the safety and efficacy of a high dose regimen of JNJ-64304500 compared with placebo and will enroll a minimum of 120 subjects and a maximum of 170 subjects (Bio-IR and Bio-NF).
- **Part II** will study the safety and efficacy of multiple-dose regimens of JNJ-64304500 compared with placebo, with ustekinumab (STELARA®) as a reference arm. Part II will enroll approximately 250 subjects (the maximum proportion of either Bio-NF or Bio-IR subjects will be 60%).

Schematic representations of Part I and Part II are shown in Figure 2 and Figure 3, respectively.

**Figure 2: Schematic Overview of Part I of Study 64304500CRD2001**

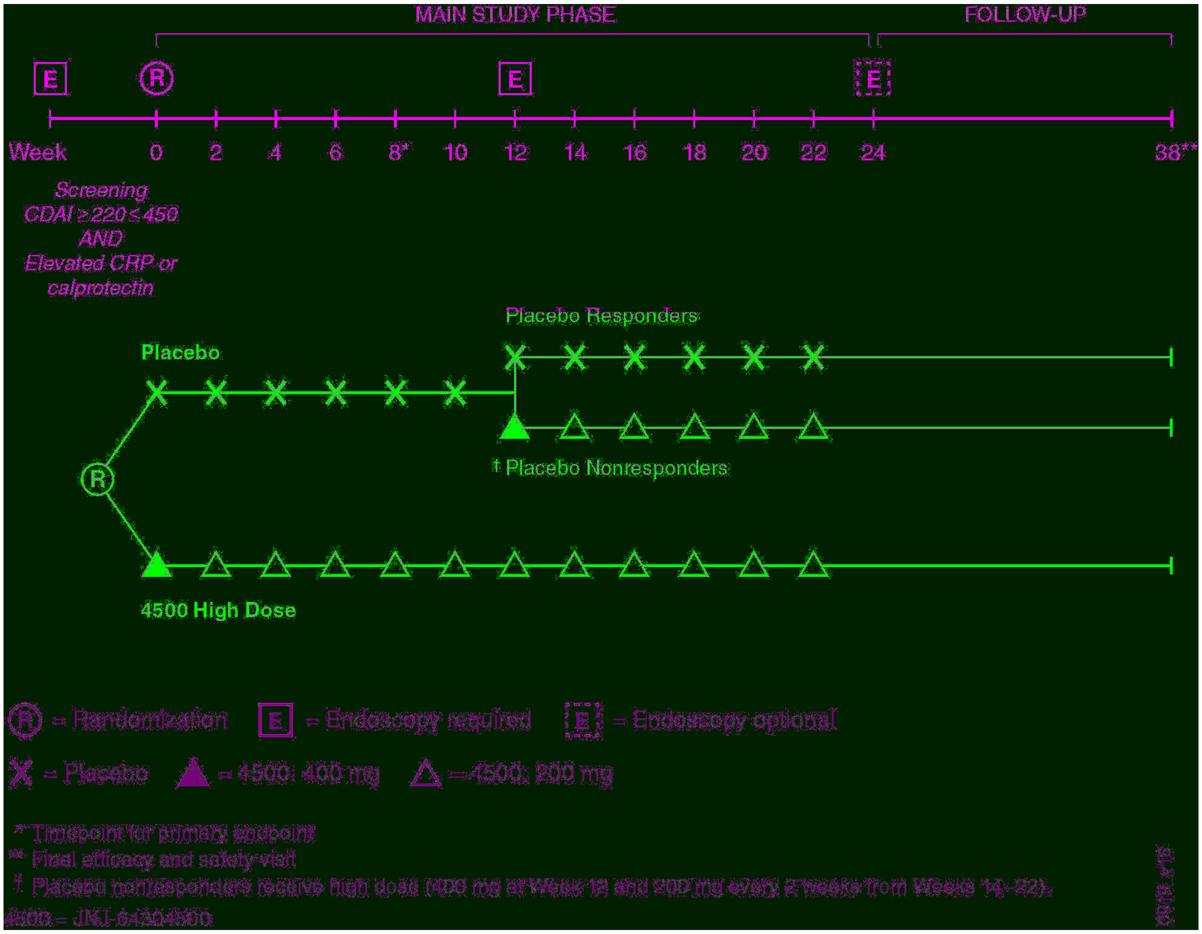

Throughout Part I and the main study phase of Part II efficacy, PK, PD, immunogenicity, biomarkers, and safety will be assessed at timepoints indicated in the appropriate Time and Events Schedules (Table 1 and Table 2). Efficacy evaluations will not be assessed in the Part II LTE.

Blood samples for pharmacogenomic analyses will be collected from subjects who consent separately to this component of the protocol (where local regulations permit). Subject participation in pharmacogenomic research is optional.

The primary endpoint for Part I is the change from baseline in the CDAI score at Week 8. The primary endpoint for Part II is the change from baseline in the CDAI score at Week 12.

After Protocol Amendment 5 is implemented, all subjects who complete Part II of the study through Week 24, and who may benefit from continued treatment in the opinion of the investigator, are eligible to enter the Part II LTE and continue to receive treatment for up to 52 weeks (Week 24 to Week 72), as described in Section 3.1.4. A schematic representation of the Part II LTE is shown in Figure 3.

**Figure 3: Schematic Overview of the 64304500CRD2001 Part II (Including the Long-term Extension)**

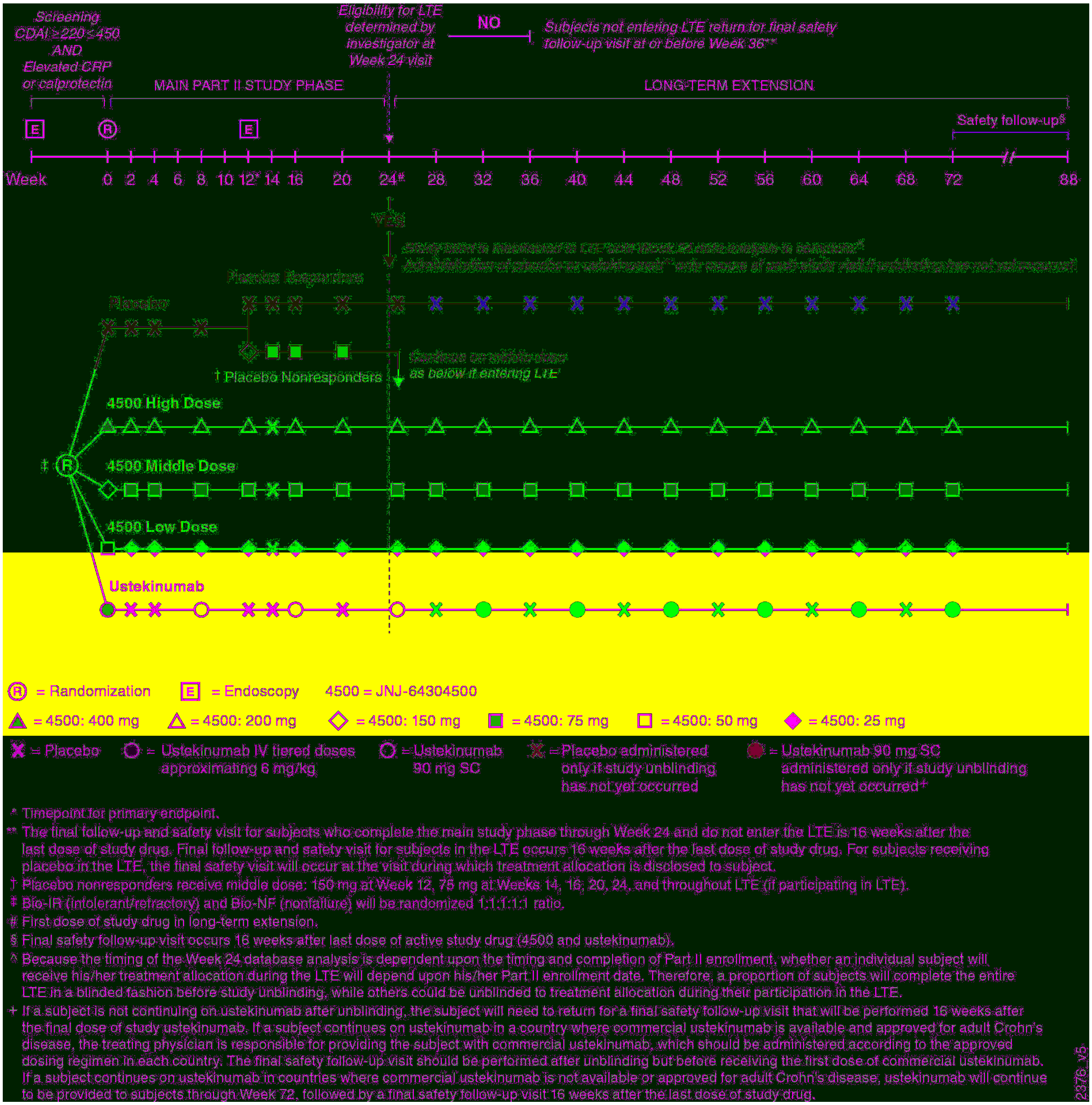

An external Data Monitoring Committee (DMC) will review unblinded safety data from all subjects periodically to monitor subject safety. The DMC will consist of at least one medical expert in the relevant therapeutic area and at least one statistician. The DMC responsibilities, authorities, and procedures will be documented in its charter. Refer to Section 11.11, Data Monitoring Committee, for details.

##### 3.1.1. Part I

In Part I, a minimum of 120 subjects and a maximum of 170 subjects (Bio-IR and Bio-NF) will be randomly assigned to receive placebo or the JNJ-64304500 high dose in a 1:1 ratio using permuted block randomization, stratified by baseline CDAI score (≤300 or >300) and SNP-positive status (yes or no). Separate randomizations will be used for the Bio-IR and Bio-NF populations.

The treatment groups for each study in Part I will be as follows:

- Placebo SC at Weeks 0, 2, 4, 6, 8, and 10; from Week 12, these subjects will receive additional doses as follows:
  - Placebo-treated subjects who are in clinical response at Week 12 (≥100-point reduction from baseline in CDAI or CDAI <150) will continue to receive placebo SC injections q2w from Week 12 through Week 22.
  - Placebo-treated subjects who are not in clinical response at Week 12 will receive JNJ-64304500 400 mg SC at Week 12 and then 200 mg SC q2w from Week 14 through Week 22.
- JNJ-64304500 400 mg SC at Week 0 then 200 mg SC q2w through Week 22.

An interim analysis is planned in Part I when 100 randomized subjects (Bio-IR and Bio-NF) have completed their Week 12 visit or have terminated their study participation before Week 12. The interim analysis will allow for uninterrupted patient enrollment through a seamless transition from Part I to Part II of the trial.

To allow for uninterrupted patient enrollment through a seamless transition from Part I to Part II of the trial, enrollment for Part I will continue until the sponsor makes a decision on whether to start Part II based on the interim analysis or when a maximum of 170 subjects have been randomized, whichever occurs first. If Part II is not initiated based on the interim analysis results, then the results through Week 12 for all subjects in Part I (ie, when all randomized subjects in Part I have either completed the Week 12 visit or terminated study participation prior to Week 12) will be examined to determine whether to start Part II. Under this scenario, a pause in enrollment between Part I and Part II will occur.

##### 3.1.2. Part II

In Part II, 250 additional Bio-IR or Bio-NF subjects will be randomly assigned to receive placebo or 1 of 3 dose levels of JNJ-64304500 or ustekinumab in a ratio of 1:1:1:1:1 using permuted block randomization, stratified by baseline CDAI score (≤300 or >300), SNP-positive status (yes or no), and Bio-IR status (yes or no). The maximum proportion of either Bio-NF or Bio-IR subjects will be 60%.

The treatment groups in Part II will be as follows:

- Placebo SC at Weeks 0, 2, 4, and 8; from Week 12, these subjects will receive additional doses as follows:

   - Placebo-treated subjects who are in clinical response at Week 12 (≥100-point reduction from baseline in CDAI or CDAI <150) will continue to receive placebo at Weeks 12, 14, 16, and 20.
   - Placebo-treated subjects who are not in clinical response at Week 12 will receive JNJ-64304500 150 mg SC at Week 12 and then JNJ-64304500 75 mg SC at Weeks 14, 16, and 20.
- High dose: JNJ-64304500 400 mg SC at Week 0 and 200 mg SC at Weeks 2 and 4, then 200 mg SC every 4 weeks (q4w) through Week 20.
- Middle dose: JNJ-64304500 150 mg SC at Week 0 and 75 mg SC at Weeks 2 and 4, then 75 mg SC q4w through Week 20.
- Low dose: JNJ-64304500 50 mg SC at Week 0 and 25 mg SC at Weeks 2 and 4, then 25 mg SC q4w through Week 20.
- Ustekinumab (tiered doses approximating 6 mg/kg intravenously [IV]) at Week 0 (as indicated in the bullets below), followed by 90 mg SC at Weeks 8 and 16.
   - Ustekinumab 260 mg (weight ≤55 kg).
   - Ustekinumab 390 mg (weight >55 kg and ≤85 kg).
   - Ustekinumab 520 mg (weight >85 kg);

As indicated in Figure 2, subjects will also receive placebo administrations, as necessary, to maintain the blind of Part II; see Section 6.2 for details.

##### 3.1.3. Interim Analysis

An interim analysis is planned in Part I when 100 randomized subjects (Bio-IR and Bio-NF) have completed their Week 12 visit or have terminated their study participation before Week 12.

This interim analysis will allow for uninterrupted patient enrollment through a seamless transition from Part I to Part II of the trial. As this interim analysis does not affect the conduct or completion of Part I, it will be considered administrative and will not require multiplicity adjustment for the final Part I analysis.

A sponsor committee independent of the study team will be established to review the interim data and formulate recommended decisions/actions in accordance with predefined decision rules (to be defined in the Interim Analysis Plan).

An interim analysis is not planned for Part II.

Other planned DBLs are described in Section 5.2.

##### 3.1.4. Part II Long-Term Extension

After Protocol Amendment 5 implementation, subjects who complete Part II Week 24 assessments, and who may benefit from continued treatment in the opinion of the investigator, are eligible to enter the Part II LTE. In the Part II LTE, each subject will be eligible to receive up to 52 weeks of additional study drug (for a total of up to 72 weeks of study drug in Part II). The first dose of study drug within the Part II LTE will be administered after the Week 24 assessments have been completed. The dose may be administered on the same day as the Week 24 assessments, or may be administered on a later date, as long as the dose is administered within the Week 24 visit window.

Subjects will continue to receive the same treatment regimen during the Part II LTE that they were receiving between Week 12 and Week 20 in the main study phase of Part II (placebo, high, middle, low dose JNJ-64304500 or ustekinumab). To maintain the study blind, all patients, investigators, and sites will remain blinded to treatment allocation during the Part II LTE until the last subject in the Part II main study phase has completed the Week 24 assessments and the Week 24 data analysis has been completed.

The timing of the Week 24 data analysis is dependent upon the timing and completion of Part II enrollment. Whether an individual subject will receive his/her treatment allocation during the Part II LTE will depend upon his/her Part II enrollment date. Therefore, a portion of subjects will complete the entire Part II LTE in a blinded fashion before study unblinding, while other subjects could be unblinded to treatment allocation during their participation in the Part II LTE.

Any subject who withdraws from the Part II LTE prior to study unblinding will return for a final safety follow-up visit 16 weeks after the last dose of study drug. For subjects that remain in the Part II LTE at the time of study unblinding, continuation of study drug during the Part II LTE will be dependent upon treatment allocation as follows:

- **JNJ-64304500**: Subjects will continue to receive the same dose of JNJ-64304500 administered during the Part II main study phase. Subjects will continue to receive study drug q4w, although any additional placebo injections used to maintain the study blind during the blinded phase of the Part II LTE will be discontinued after study unblinding. Following the last scheduled dose of study drug at Week 72, a final safety follow-up visit will be performed at approximately Week 88. For subjects who discontinue study participation prior to Week 72, a final safety follow-up visit will be performed 16 weeks after the last dose of study drug.

- **Placebo**: Subjects receiving placebo who remain in the Part II LTE at the time of study unblinding will have their final safety follow-up visit subsequent to unblinding and will be discontinued from the study. No further follow-up visits will be performed.

- **Ustekinumab**: Subjects receiving ustekinumab will continue to receive it during the Part II LTE until unblinding occurs. After unblinding, based on the treating physician's clinical judgment, subjects may receive ustekinumab in a manner dependent on the country in which they are located:

    - If a subject is not continuing on ustekinumab after unblinding, the subject will need to return for a final safety follow-up visit that will be performed 16 weeks after the final dose of study ustekinumab.

- If a subject continues on ustekinumab in a country where commercial ustekinumab is available and approved for adult Crohn's disease, the treating physician is responsible for providing the subject with commercial ustekinumab, which should be administered according to the approved dosing regimen in each country. The final safety follow-up visit should be performed after unblinding but before receiving the first dose of commercial ustekinumab.

- If a subject continues on ustekinumab in countries where commercial ustekinumab is not available or approved for adult Crohn's disease, ustekinumab will continue to be provided to subjects through Week 72, followed by a final safety follow-up visit 16 weeks after the last dose of study drug.

For eligible Part II subjects, participation in the Part II LTE is entirely voluntary. Eligible subjects who do not wish to enter the Part II LTE will complete the Part II Week 24 safety and efficacy assessments, followed by the final efficacy and safety assessments at Week 36.

During the Part II LTE, all concomitant medications, including Crohn's disease-specific medications (except for the prohibited medications listed in Section 8.2) may be administered at the discretion of the investigator.

All subjects in the Part II LTE will be assessed according to the Time & Events Schedule (Table 3, Table 4), which includes assessments, AEs, laboratory analyses, and PK and immunogenicity samples.

#### 3.2. Study Design Rationale

This protocol is comprised of 2 parts (Part I [Study 1: Bio-IR subjects and Study 2: Bio-NF subjects] and Part II [Bio-IR and Bio-NF subjects]) that are designed to evaluate the safety and efficacy of JNJ-64304500 in subjects with moderately to severely active Crohn's disease. Study 1 and Study 2 constitute Part I of the protocol. In this part, the safety and efficacy of a single dosing regimen of JNJ-64304500 in Bio-IR and Bio-NF subjects with moderately to severely active Crohn's disease is evaluated. If acceptable efficacy is established in Part I (for the combined Bio-IR and Bio-NF subjects), Part II (a dose-ranging study in Bio-IR and Bio-NF subjects) will be initiated. After Protocol Amendment 5 is implemented, subjects who complete Part II Week 24 assessments and who may benefit from continued treatment in the opinion of the investigator, are eligible to enter the Part II LTE to receive up to 52 weeks of additional treatment. Subjects who have completed Week 24 prior to the implementation of Protocol Amendment 5 will not be eligible to enroll in the Part II LTE.

##### 3.2.1. Choice of JNJ-64304500 Dose for Placebo Nonresponders

In Part I, the high dose of JNJ-64304500 (400 mg/200 mg) was chosen for placebo nonresponders because it is the only dose regimen studied in this part.

In Part II, the middle dose was chosen for placebo nonresponders as it is believed that this dose will be effective since it is higher than the dose studied in the prior Phase 2a study (where efficacy was shown). The middle dose also requires fewer injections compared with the high dose.

##### 3.2.2. Efficacy Assessments

The efficacy evaluations selected for both parts of the study (eg, CDAI, CRP, fecal biomarkers; Section 9.2) are well-established measures that are accepted by regulatory agencies as primary or supportive of clinically relevant effect of disease activity in Crohn's disease studies.

The primary endpoint for Part I is change from baseline in CDAI at Week 8. The primary endpoint for Part II is change from baseline in CDAI at Week 12.

Change in the CDAI is being used as the primary endpoint because this measure is more sensitive than remission (ie, the change in CDAI provides greater power than remission for the same sample size). Therefore, the study can be more efficient for Phase 2 using the change in CDAI.

To evaluate the level of efficacy after prolonged discontinuation of study drug, CDAI will be assessed at the final efficacy and safety visit in Part I and in Part II for those subjects who do not enter the Part II LTE. The Crohn's Disease Activity Index will be not assessed during the Part II LTE.

It is anticipated that endoscopic improvement will occur by Week 12. In order to have an appropriate comparison of JNJ-64304500 to placebo, the placebo-controlled period will continue to Week 12.

Efficacy evaluations will not be performed during the Part II LTE.

##### 3.2.3. Pharmacokinetic Assessments

Pharmacokinetic assessments will be used to further understand the disposition of JNJ-64304500 in subjects with Crohn's disease.

##### 3.2.4. Immunogenicity Assessments

Serum samples for the detection of antibodies to JNJ-64304500 will be collected to further evaluate the immunogenicity of JNJ-64304500 in subjects with Crohn's disease.

##### 3.2.5. Pharmacodynamic Assessments

Serum samples for the analysis of PD will be collected to further understand the response of subjects with Crohn's disease to treatment with JNJ-64304500.

##### 3.2.6. DNA and Biomarker Collection

It is recognized that genetic variation can be an important contributory factor to interindividual differences in drug distribution and response and can also serve as a marker for disease susceptibility and prognosis. Pharmacogenomic research may help to explain interindividual variability in clinical outcomes and may help to identify population subgroups that respond differently to a drug.

Whole blood will be collected from all subjects for SNP analysis (the NKG2D SNP rs2255336 and the MICB [NKG2D ligand] SNP rs2239705) to understand the association of these SNPs with response to JNJ-64304500 (refer to the latest version of the IB for more information). In addition, subjects who sign an optional pharmacogenomics consent form will undergo complete genomic testing.

The goal of this pharmacogenomic component is to collect DNA to allow the identification of genetic factors that may influence the PK, PD, efficacy, safety, or tolerability of JNJ-64304500 and to identify genetic factors associated with Crohn's disease.

Biomarker assessments will be made to examine the biologic response to treatment and to identify biomarkers that are relevant to JNJ-64304500 treatment and/or Crohn's disease. Blood samples for serum-based biomarker analyses will be collected from all subjects to assess proteins related to the NKG2D pathway or the pathogenesis of Crohn's disease. Whole blood samples will be collected from all subjects for the analysis of ribonucleic acid (RNA) expression and T-cell receptor repertoire. Mucosal biopsy samples will be collected during ileocolonoscopy for the analysis of gene and/or protein expression and the histologic assessment of disease and/or healing.

Receptor occupancy (RO) assessments for NKG2D and immunophenotyping assessments (including NK cells and CD8+ T cells) will also be performed. Immunophenotyping will be conducted using flow cytometry to assess the number of CD4, CD8, and NK cells before versus after dose administration.

##### 3.2.7. Control, Randomization, and Blinding

In both parts of the study, a placebo control will be used to establish the frequency and magnitude of changes in clinical endpoints that may occur in the absence of active treatment. In addition to placebo control, a ustekinumab reference arm will be used in Part II to determine the sensitivity of the clinical endpoints in this study.

Ustekinumab was chosen for use as a reference arm because the efficacy and safety profile of ustekinumab are well described. It is also recognized that use of other therapeutics (eg, TNFα antagonists) could potentially confound the population of Bio-IR subjects and introduce substantial patient burden to maintain blinding.

Randomization will be used to minimize bias in the assignment of subjects to treatment groups, to increase the likelihood that known and unknown subject attributes (eg, demographic and baseline characteristics) are evenly balanced across treatment groups, and to enhance the validity of statistical comparisons across treatment groups. Blinded treatment will be used to reduce potential bias during data collection and evaluation of clinical endpoints.

##### 3.2.8. Dose Selection

###### 3.2.8.1. JNJ-64304500

The results of PK, PD and efficacy analyses and safety data from previous clinical studies of JNJ-64304500 in subjects with RA and Crohn's disease were used to guide the dose selection for the 3 studies in this protocol. In study NN8555-3618, the highest single SC dose investigated was 7.5 mg/kg and the highest multiple SC dose regimen investigated was 4 mg/kg q2w for a total of 4 doses in RA subjects. JNJ-64304500 was well tolerated and no safety concerns were identified in subjects with RA or Crohn's disease from the previous clinical studies. In addition, a 52-week repeat-dose toxicology study has demonstrated a NOAEL of 100 mg/kg SC once weekly in cynomolgus monkeys. The exposures achieved in the last dosage interval (100 mg/kg) of the 52-week study were 10,900 and 8,480 µg/mL for maximum concentration ($C_{max}$), and 1,520,000 and 1,250,000 hr·µg/mL for $AUC_{0\text{-}168hr}$, for male and female monkeys, respectively. Based on these safety and toxicology findings, it is expected that the proposed dose regimens of JNJ-64304500 would have acceptable safety profiles.

Population PK/PD modeling and simulation was performed using JNJ-64304500 PK and NKG2D RO data from the previous clinical studies. The model-predicted JNJ-64304500 concentrations and NKG2D RO in the intestines were used to help guide the selection of the dose regimens for the present study. NKG2D RO in the intestines is predicted by assuming that the concentration of JNJ-64304500 in the intestines is approximately 10% of the concentration in the peripheral circulation because a 5%-15% antibody distribution coefficient between the general circulation and the intestines has been reported.[9]

A 16-week follow-up interval after the last administration of study agent has been employed based on the observed safety profile and PK profile of JNJ-64304500 in the completed clinical studies in RA and Crohn's disease subjects. The average serum concentration of JNJ-64304500 at 16 weeks after the last administration of study agent is predicted to be approximately 0.01 µg/mL, which is considered unlikely to produce drug-related effects or safety concerns.

**PART I**

The selected JNJ-64304500 dose regimen for Part I includes a loading dose of 400 mg SC at Week 0, followed by 200 mg SC q2 weeks through Week 22. The loading dose of 400 mg at Week 0 is intended to produce rapid onset of clinical response. The Part I dose regimen is predicted to achieve systemic exposures similar to the maximum systemic exposure that has been well tolerated in previous clinical studies. In the first-in-human study (NN8555-3618), the highest multiple-dose regimen of 4 mg/kg SC q2w (4 doses) provided a median (range) $C_{max}$ in serum of 79.3 (52.8 to 91.2) µg/mL. The predicted median JNJ-64304500 $C_{max}$ in serum is 64.64 µg/mL, median JNJ-64304500 concentration in serum at Week 8 is 38.94 µg/mL and median steady state trough JNJ-64304500 serum concentration is 38.31 µg/mL in subjects with Crohn's disease (Table 5).

Assuming the concentration of JNJ-64304500 in the intestines is approximately 10% of concentration of JNJ-64304500 in serum, the predicted median peak and trough concentrations of JNJ-64304500 in the intestines are 6.46 µg/mL and 3.83 µg/mL, respectively. Analysis of the *ex vivo* relationship between the NKG2D RO and the JNJ-64304500 serum concentration suggests that ≥ 90% RO is achieved when the JNJ-64304500 serum concentration is ≥3 µg/mL. As a result, the Part I dose regimen is expected to result in approximately 99% NKG2D RO in the intestine (Table 5). Thus, the Part I dose regimen is expected to increase the probability to achieve maximum clinical response while remaining within the acceptable safety margins in subjects with Crohn's disease.

**Table 5: Predicted median serum JNJ-64304500 concentrations and NKG2D receptor occupancy after administration of the selected dose regimens of JNJ-64304500**

| Dosing Regimen (SC) | JNJ-64304500 Serum Concentration (µg/mL) | | NKG2D Receptor Occupancy (%RO) | | |
|---|---|---|---|---|---|
| | Parameter | Value | Week | %RO in blood | %RO in intestine |
| 400 mg at Week 0 then 200 mg q2w | $C_{max}$ | 64.64 | - | - | - |
| | $C_{min}$ (Week 8) | 38.94 | 8 | 100 | 99 |
| | $C_{trough}$ (steady state) | 38.31 | 24 | 100 | 99 |
| 400 mg (Wk0), 200 mg (Wks 2 &4) then 200 mg q4w | $C_{max}$ | 53.99 | - | - | - |
| | $C_{min}$ (Week 8) | 18.95 | 8 | 100 | 97 |
| | $C_{trough}$ (steady state) | 12.00 | 24 | 100 | 96 |
| 150 mg (Wk0), 75 mg (Wks 2 &4) then 75 mg q4w | $C_{max}$ | 19.53 | - | - | - |
| | $C_{min}$ (Week 8) | 6.34 | 8 | 100 | 91 |
| | $C_{trough}$ (steady state) | 4.24 | 24 | 99 | 87 |
| 50 mg (Wk0), 25 mg (Wks 2 &4) then 25 mg q4w | $C_{max}$ | 6.46 | - | - | - |
| | $C_{min}$ (Week 8) | 1.77 | 8 | 97 | 76 |
| | $C_{trough}$ (steady state) | 0.74 | 24 | 94 | 64 |

$C_{max}$=maximum concentration; $C_{min}$ (Week 8)=concentration at Week 8; $C_{trough}$ (steady state)=steady state trough concentration; NKG2D=natural killer group 2D; q2w=every 2 weeks; q4w=every 4 weeks; %RO=percent receptor occupancy; SC=subcutaneous; Wk=Week

#### PART II

Three dose regimens of JNJ-64304500 (high, middle and low) have been selected for Part II which are expected to provide a wide range of systemic drug exposures in order to assess exposure-response relationship in subjects with Crohn's disease. The loading doses at Week 0, 2, and 4 are intended to produce rapid onset of clinical response. Since the apparent terminal half-life of JNJ-64304500 at the proposed dose regimens is about 2 to 3 weeks, SC administration of JNJ-64304500 at 4-week intervals from Week 4 through Week 20 is expected to produce median steady state trough serum JNJ-64304500 concentrations that are likely to maintain clinical response in Crohn's disease subjects. In addition, the PK/PD modeling results described below support the use of a 4-week dosing interval in Part II.

The high dose regimen for Part II is 400 mg at Week 0, 200 mg at Weeks 2 and 4, followed by 200 mg q4 weeks through Week 20. This dose regimen is predicted to result in a median JNJ-64304500 $C_{max}$ in serum of 53.99 µg/mL, a median JNJ-64304500 serum concentration at Week 8 of 18.95 µg/mL, and a median trough serum JNJ-64304500 concentration at steady state of 12.00 µg/mL in subjects with Crohn's disease. Simulations results suggest that 89% of subjects on this high dose regimen are expected to maintain trough serum JNJ-64304500 concentrations >3 µg/mL at steady state, and the predicted median intestinal NKG2D RO is >96% (Table 5).

The middle dose regimen for Part II is 150 mg SC at Week 0, 75 mg at Weeks 2 and 4, followed by 75 mg q4w through Week 20. This middle dose regimen is predicted to result in approximately 38% of the systemic exposure achieved with the high dose regimen. The predicted median intestinal NKG2D RO at steady state is 87%.

The low dose regimen for Part II is 50 mg SC at Week 0, 25 mg at Weeks 2 and 4, followed by 25 mg q4 weeks through Week 20. The low dose regimen is selected to explore the minimum effective dose of JNJ-64304500 in subjects with Crohn's disease. The predicted median serum trough concentration of JNJ-64304500 at steady state is 0.74 µg/mL which is predicted to result in a median NKG2D RO of 64% in the intestine. Furthermore, in the first-in-humans study in RA subjects, a decrease in NKG2D expression on NK cells was not observed until the dose of JNJ-64304500 was ≥0.3 mg/kg q2w, and decreases in NKG2D expression on both CD8+ T cells and NK cells were not observed until the dose was ≥1 mg/kg q2w. These observations suggest that a dose regimen of at least 0.3 mg/kg q2w may be required to produce pharmacological effects, and the selected low maintenance dose regimen of 25 mg q4w would likely produce treatment effect at the lower part of the exposure-response curve.

**Part I and Part II**

The selection of the 4 different dosing regimens of JNJ-64304500 in Part I and Part II was based on all available PK, efficacy and safety data from the previous clinical studies and from a 52-week repeat-dose toxicology study. It should be noted that the current available information on JNJ-64304500 has not established the relationship between the NKG2D RO and clinical effects of the drug. In addition, the currently reported antibody distribution coefficients for intestinal tissues may have limitations and the JNJ-64304500 concentrations in the intestine may not be accurately predicted. Nevertheless, the use of 4 different dose regimens of JNJ-64304500 in Part I and Part II, which will provide a wide range of drug exposures while remaining within the acceptable safety margins, is anticipated to provide a robust characterization of the exposure-response relationship of JNJ-64304500 in the treatment of Crohn's disease.

###### 3.2.8.2. Ustekinumab

Results from the 2 Phase 3 induction studies of IV ustekinumab (Study CNTO1275CRD3001 in subjects with Crohn's disease who failed 1 or more TNFα antagonists, and Study CNTO1275CRD3002 in subjects who failed conventional therapy with immunomodulators and/or corticosteroids) and 1 Phase 3 maintenance study with SC ustekinumab (Study CNTO1275CRD3003, which enrolled clinical responders from the 2 IV induction studies) were used to determine the appropriate dose regimen of ustekinumab for the treatment of Crohn's disease. As a result, a single IV induction dose of 6 mg/kg ustekinumab (administered as body weight-based tiered fixed doses, see Section 3.1.2 for details) at Week 0, followed by SC maintenance therapy of 90 mg every 8 weeks has been demonstrated to provide robust efficacy across a range of endpoints including patient-reported outcomes, objective measures of inflammation, and health-related quality of life measures, as well as a favorable safety profile.

### 4. SUBJECT POPULATION

Screening for eligible subjects will be performed within 5 weeks before administration of the study drug.

The inclusion and exclusion criteria for enrolling subjects in the 2 studies are described in the following 2 subsections. If there is a question about the inclusion or exclusion criteria, the investigator must consult with the appropriate sponsor representative and resolve any issues before enrolling a subject in the study. Waivers are not allowed.

For a discussion of the statistical considerations of subject selection, refer to Section 11.1, Sample Size Determination.

The following inclusion and exclusion criteria apply to Part I and Part II within this protocol.

#### 4.1. Inclusion Criteria

Each potential subject must satisfy all of the following criteria to be randomized in the study:

1. Be a man or woman ≥18 years of age.
2. Have Crohn's disease or fistulizing Crohn's disease of at least 3 months' duration, with colitis, ileitis, or ileocolitis, confirmed at any time in the past by radiography, histology, and/or endoscopy.
3. Have active Crohn's disease, defined as a baseline CDAI score of ≥220 but ≤450.
4. Have at least one of the following at screening:
   a. An abnormal CRP (>0.3 mg/dL [>3.0 mg/L])

      **OR**

   b. Calprotectin >250 mg/kg.
5. Criterion modified per Amendment 2:

   **In Study 1 of Part I**, meet the following requirements for prior or current medications for Crohn's disease:

   a. Has previously demonstrated inadequate response, loss of response, or intolerance to 1 or more approved biologic therapies (eg, infliximab, adalimumab, certolizumab pegol, or vedolizumab) as outlined in Attachment 1.

   **In Study 2 of Part I**, meet the following requirements for prior or current medications for Crohn's disease:

   b. Has failed conventional therapy:
      1) Is currently receiving corticosteroids and/or immunomodulators (ie, AZA, 6-MP, or MTX) at adequate therapeutic doses (Attachment 2).

         **OR**

      2) Has a history of failure to respond to or tolerate an adequate course of corticosteroids and/or immunomodulators (ie, AZA, 6-MP, or MTX) (Attachment 2).

         **OR**

      3) Is corticosteroid dependent or has had a history of corticosteroid dependency (Attachment 2).
6. **In Part II,** meet the following requirement for prior or current medications for Crohn's disease:
   a. Has previously demonstrated inadequate response, loss of response, or intolerance to 1 or more approved biologic therapies (eg, infliximab, adalimumab, certolizumab pegol, or vedolizumab) as outlined in Attachment 1.

      **OR**

   b. Has failed conventional therapy:

1) Is currently receiving corticosteroids and/or immunomodulators (ie, AZA, 6-MP, or MTX) at adequate therapeutic doses (Attachment 2).

**OR**

2) Has a history of failure to respond to or tolerate an adequate course of corticosteroids and/or immunomodulators (ie, AZA, 6-MP, or MTX) (Attachment 2).

**OR**

3) Is corticosteroid dependent or has had a history of corticosteroid dependency (Attachment 2).

7. Adhere to the following requirements for concomitant medication for the treatment of Crohn's disease, which are permitted provided that doses meeting these requirements are stable, or have been discontinued, for at least 3 weeks before baseline (Week 0), unless otherwise specified:

   a. Oral 5-aminosalicylic acid (5-ASA) compounds.

   b. Oral corticosteroids at a prednisone-equivalent dose at or below 40 mg/day, or 9 mg/day of budesonide, or 5 mg/day beclomethasone dipropionate.

   c. Antibiotics being used as a primary treatment of Crohn's disease.

   d. Conventional immunomodulators (ie, AZA, 6-MP, or MTX): subjects must have been taking them for at least 12 weeks and at a stable dose for at least 4 weeks before baseline.

8. A subject with a family history of colorectal cancer, personal history of increased colorectal cancer risk, age >50 years, or other known risk factor must be up-to-date on colorectal cancer surveillance (may be performed during screening). Adenomatous polyps must be removed before the first administration of study agent.

9. A subject who has had extensive colitis for ≥8 years, or disease limited to the left side of the colon for ≥12 years, must either have had a colonoscopy to assess for the presence of dysplasia within 1 year before the first administration of study agent or a colonoscopy to assess for the presence of malignancy at the screening visit, with no evidence of malignancy.

10. Have screening laboratory test results within the following parameters:

   a. Hemoglobin ≥8.0 g/dL.

   b. White blood cell count $\geq 3.0\times10^3/\mu L$.

   c. Neutrophils $\geq 1.5\times10^3/\mu L$.

   d. Platelets $\geq 100\times10^3/\mu L$.

   e. Serum creatinine <1.7 mg/dL.

   f. Alanine aminotransferase (ALT) and aspartate aminotransferase (AST) concentrations must be within 2 times the upper limit of the normal (ULN) range for the laboratory conducting the test.

   g. Direct (conjugated) bilirubin <1.0 mg/dL.

11. Are considered eligible according to the following tuberculosis (TB) screening criteria:

    a. Have no history of latent or active TB before screening. Exceptions are made for subjects currently receiving treatment for latent TB, if there is no evidence of active TB, or who have a history of latent TB and documentation of having completed adequate treatment for latent TB within 3 years before the first administration of study agent. It is the responsibility of the investigator to verify the adequacy of previous TB treatment and provide appropriate documentation.

       Note: The exceptions outlined above exclude subjects in countries with high multidrug-resistant TB burden (eg, Brazil, China, India, the Russian Federation, and South Africa), due to potential concerns for multi-drug-resistant TB.

    b. Have no signs or symptoms suggestive of active TB upon medical history and/or physical examination.

    c. Have had no recent close contact with a person with active TB or, if there has been such contact, will be referred to a physician specializing in TB to undergo additional evaluation and, if warranted, receive appropriate treatment for latent TB before or simultaneously with the first administration of study agent.

    d. Criterion modified per Amendment 1.

       d.1 Within 2 months before the first administration of study agent, either have negative QuantiFERON-TB test, or have a newly identified positive QuantiFERON-TB test in which active TB has been ruled out, and for which appropriate treatment for latent TB has been initiated either before or simultaneously with the first administration of study agent (except in countries with high multidrug-resistant TB burden [eg, Brazil, China, India, the Russian Federation, and South Africa]), where subjects with a newly identified positive QuantiFERON-TB test result are excluded). Indeterminate results should be handled as outlined in Section 9.1.2. A negative tuberculin skin test (Attachment 3) is additionally required if the QuantiFERON-TB test is not approved/registered in that country. A tuberculin skin test is recommended but not required for study centers in countries where tuberculin is not available. The QuantiFERON-TB test is not required at screening for subjects with a history of latent TB and appropriate treatment as described above in Inclusion Criterion 11a.

    e. Have a chest radiograph (posterior-anterior and lateral views), taken within 3 months before the first administration of study agent and read by a qualified radiologist, with no evidence of current active TB or old inactive TB.

12. A woman of childbearing potential must have a negative highly sensitive serum (β-human chorionic gonadotropin [β-hCG]) pregnancy test result at screening and a negative urine pregnancy test result at Week 0.

13. Before randomization, a female subject must be either:

    a. **Not of childbearing potential**, defined as:

       1) Premenarchal: A premenarchal state is one in which menarche has not yet occurred.

2) Postmenopausal: A postmenopausal state is defined as no menses for 12 months without an alternative medical cause. A high follicle-stimulating hormone level (>40 IU/L or mIU/mL) in the postmenopausal range may be used to confirm a postmenopausal state in women not using hormonal contraception or hormonal replacement therapy; however, in the absence of 12 months of amenorrhea, a single follicle-stimulating hormone measurement is insufficient.

3) Permanently sterile: Permanent sterilization methods include hysterectomy, bilateral salpingectomy, bilateral tubal occlusion/ligation procedures, and bilateral oophorectomy.

**OR**

b. **Of childbearing potential and**:

1) Practicing a highly effective method of contraception (failure rate of <1% per year when used consistently and correctly), consistent with local regulations regarding the use of contraceptive methods for subjects participating in clinical studies. Examples of highly effective contraceptives include user-independent methods such as implantable progestogen-only hormone contraception associated with inhibition of ovulation; intrauterine device (IUD); intrauterine hormone-releasing system (IUS); vasectomized partner; or sexual abstinence (considered a highly effective method only if defined as refraining from heterosexual intercourse during the entire period of risk associated with the study drug, and if in line with the preferred and usual lifestyle of the subject); or user-dependent methods such as combined (estrogen- and progestogen-containing) hormonal contraception associated with inhibition of ovulation (oral, intravaginal, transdermal); or progestogen-only hormone contraception associated with inhibition of ovulation (oral, injectable). If using a hormone birth control method, a second method of birth control, such as a condom or diaphragm, must be used.

2) Agrees to remain on a highly effective method of contraception throughout the study and for at least 16 weeks after the last administration of study agent.

Note: If a subject's childbearing potential changes after start of the study (eg, a premenarchal woman experiences menarche) or the risk of pregnancy changes (eg, a woman who is not heterosexually active becomes active), a woman must begin a highly effective method of contraception, as described throughout the inclusion criteria.

14. A woman must agree not to donate eggs (ova, oocytes) for the purposes of assisted reproduction during the study and for a period of 16 weeks after the last administration of study agent.

15. During the study and for at least 16 weeks after the last administration of study agent, a man

    a. who is sexually active with a woman of childbearing potential must agree to use a barrier method of contraception (eg, condom with spermicidal foam/gel/film/cream/suppository).

    b. who is sexually active with a pregnant woman must use a condom.

    c. must agree not to donate sperm.

16. Be willing and able to adhere to the prohibitions and restrictions specified in this protocol.

17. Must sign an informed consent form (ICF) indicating that he or she understands the purpose of and procedures required for the study and is willing to participate in the study.
18. Criterion modified per Amendment 1.
    18.1 DNA sample collection for SNP testing is required for all subjects in this study. Each subject must have a SNP status of either positive or negative. Each subject must sign a separate ICF if he or she agrees to consent to additional optional DNA research where local regulations permit. Refusal to give consent for the optional DNA research does not exclude a subject from participation in the study.

#### 4.2. Exclusion Criteria

Any potential subject who meets any of the following criteria will be excluded from participating in the study:

1. Has complications of Crohn's disease such as symptomatic strictures or stenoses, short gut syndrome, or any other manifestation that might be anticipated to require surgery, could preclude the use of the CDAI to assess response to therapy, or would possibly confound the ability to assess the effect of treatment with JNJ-64304500 or ustekinumab.
2. Currently has or is suspected to have an abscess. Recent cutaneous and perianal abscesses are not exclusionary if drained and adequately treated at least 3 weeks before baseline, or 8 weeks before baseline for intra-abdominal abscesses, provided that there is no anticipated need for any further surgery. Subjects with active fistulas may be included if there is no anticipation of a need for surgery and there are currently no abscesses identified.
3. Has had any kind of bowel resection within 6 months or any other intra-abdominal surgery within 3 months before baseline.
4. Has a draining (ie, functioning) stoma or ostomy.
5. Has received any of the following prescribed medications or therapies within the specified period:
    a. IV corticosteroids <3 weeks before baseline.
    b. Other oral immunomodulatory agents (eg, 6-thioguanine [6-TG], cyclosporine, tacrolimus, sirolimus, or mycophenolate mofetil, tofacitinib and other Janus kinase [JAK] inhibitors) <6 weeks or within 5 half-lives of agent before baseline, whichever is longer.
    c. Nonbiologic experimental or investigational agents <4 weeks or within 5 half-lives of agent before baseline, whichever is longer.
    d. Nonautologous stem cell therapy (eg, Prochymal), natalizumab, efalizumab, or biologic agents that deplete B or T cells (eg, rituximab, alemtuzumab, or visilizumab) <12 months before baseline.
    e. TNFα-antagonist biologic agents (eg, monoclonal antibody therapies) or other agents intended to suppress or eliminate TNFα <8 weeks before baseline.
    f. Vedolizumab <16 weeks before baseline.
    g. Other immunomodulatory biologic agents <12 weeks or within 5 half-lives of agent before baseline, whichever is longer.

h. Treatment with apheresis (eg, Adacolumn apheresis) <3 weeks before baseline.

i. Initiation of total (complete) or partial (supplemental) parenteral nutrition administered through any indwelling catheter <3 weeks before baseline or anticipated to require parenteral nutrition administered through an indwelling catheter during enrollment in the study.

j. Initiation of enteral therapy for Crohn's disease (liquid nutritional formula comprising ≥80% of total caloric intake administered through the gastrointestinal tract) <3 weeks before baseline. Subjects who are on a stable regimen of enteral feeds ≥3 weeks before baseline may be considered for enrollment if they plan to continue enteral feeds as treatment for Crohn's disease through the duration of the study (Week 24, Part I or Part II).

6. Has a stool culture or other examination positive for an enteric pathogen, including *Clostridium difficile* toxin, in the last 4 months unless a repeat examination is negative and there are no signs of ongoing infection with that pathogen.

7. Has previously received a biologic agent targeting IL-12 or IL-23, including but not limited to ustekinumab or briakinumab (ABT-874).

8. Has previously received JNJ-64304500 or NNC0142-002.

9. Has received a Bacille Calmette-Guérin (BCG) vaccination within 12 months or any other live bacterial or live viral vaccination within 12 weeks before baseline.

10. Has a history of, or ongoing, chronic or recurrent infectious disease, including but not limited to, chronic renal infection, chronic chest infection, recurrent urinary tract infection (eg, recurrent pyelonephritis or chronic nonremitting cystitis), or open, draining, or infected skin wounds or ulcers.

11. Has current signs or symptoms of infection. Established nonserious infections (eg, acute upper respiratory tract infection, simple urinary tract infection) need not be considered exclusionary at the discretion of the investigator.

12. Has a history of serious infection (eg, sepsis, pneumonia, or pyelonephritis), including any infection requiring hospitalization or IV antibiotics, for 8 weeks before baseline.

13. Has evidence of a *Herpes zoster* infection ≤8 weeks before baseline.

14. Has a history of latent or active granulomatous infection, including histoplasmosis or coccidioidomycosis, before screening. Refer to Inclusion Criteria 11a for information regarding eligibility with a history of latent TB.

15. Has evidence of current active infection, including TB, or a nodule suspicious for lung malignancy on screening or any other available chest radiograph, unless definitively resolved surgically or by additional imaging and with source document confirmation.

16. Has or ever has had a nontuberculous mycobacterial infection or serious opportunistic infection (eg, cytomegalovirus colitis, *Pneumocystis carinii*, aspergillosis).

17. Has a history of human immunodeficiency virus (HIV) antibody positivity, or tests positive for HIV at screening.

18. Are seropositive for antibodies to hepatitis C virus (HCV) without a history of successful treatment, defined as being negative for HCV RNA at least 24 weeks after completing antiviral treatment.

19. Subjects must undergo screening for hepatitis B virus (HBV). At a minimum, this includes testing for HBV surface antigen (HBsAg), HBV surface antibody (anti-HBs), and HBV core antibody (anti-HBc) total:
    a. Subjects who test negative for all HBV screening tests (ie, HBsAg-, anti-HBc-, and anti-HBs-) **are eligible** for this study.
    b. Subjects who test positive for surface antigen (HBsAg+) **are not eligible** for this study, regardless of the results of other hepatitis B tests.
    c. Subjects who test negative for surface antigen (HBsAg-) and test positive for core antibody (anti-HBc+) and surface antibody (anti-HBs+) **are eligible** for this study.
    d. Subjects who test positive only for surface antibody (anti-HBs+) **are eligible** for this study.
    e. Subjects who test positive only for core antibody (anti-HBc+) must undergo further testing for hepatitis B DNA acid (HBV DNA test). If the HBV DNA test is positive, the subject **is not eligible** for this study. If the HBV DNA test is negative, the subject **is eligibl**e for this study. In the event the HBV DNA test cannot be performed, the subject **is not eligible** for this study.

    **Note:** For subjects who **are not eligible** for this study due to HIV, HCV, and HBV test results, consultation with a physician with expertise in the treatment of those infections is recommended.
20. Has severe, progressive, or uncontrolled renal, hepatic, hematological, endocrine, pulmonary, cardiac, neurologic, cerebral, or psychiatric disease, or signs and symptoms thereof.
21. Has a transplanted organ (with exception of a corneal transplant >12 weeks before screening).
22. Has a known history of lymphoproliferative disease, including monoclonal gammopathy of unknown significance (MGUS), lymphoma, or signs and symptoms suggestive of possible lymphoproliferative disease, such as lymphadenopathy and/or splenomegaly.
23. Has any known malignancy or has a history of malignancy (with the exception of basal cell carcinoma; squamous cell carcinoma in situ of the skin; or cervical carcinoma in situ that has been treated with no evidence of recurrence; or squamous cell carcinoma of the skin that has been treated with no evidence of recurrence within 5 years before screening).
24. Is unable or unwilling to undergo multiple venipunctures because of poor tolerability or lack of easy access to veins.
25. Is known to have had a substance abuse (drug or alcohol) problem within the previous 12 months before baseline.
26. Has known allergies, hypersensitivity, or intolerance to JNJ-64304500 or ustekinumab or any of their excipients (refer to IBs).
27. Are currently or intending to participate in any other study using an investigational agent or procedure during participation in this study.
28. Is a woman who is pregnant, or breast-feeding, or planning to become pregnant or is a man who plans to father a child while randomized in this study or within 16 weeks after the last administration of study agent.

29. Has any condition that, in the opinion of the investigator, would make participation not be in the best interest (eg, compromise the well-being) of the subject or that could prevent, limit, or confound the protocol-specified assessments.
30. Is an employee of the investigator or study site, with direct involvement in the proposed study or other studies under the direction of that investigator or study site, as well as family members of the employees or the investigator.

**NOTE:** Investigators should ensure that all study enrollment criteria have been met at screening. If a subject's clinical status changes (including any available laboratory results or receipt of additional medical records) after screening but before the first dose of study drug is given, such that he or she no longer meets all eligibility criteria, then the subject should be excluded from participation in the study. Section 9.1.2 describes options for retesting. Section 17.4 describes the required documentation to support meeting the enrollment criteria.

**Long-term Extension Phase**

After Protocol Amendment 5 is implemented, subjects who complete Part II Week 24 assessments and who may benefit from continued treatment in the opinion of the investigator, are eligible to enter the Part II LTE to receive up to 52 weeks of additional treatment. Subjects must be excluded from LTE if they have developed Crohn’s disease complications which may be anticipated to require surgery in the next 12 months, if they have clinical evidence of a serious infection at Week 24, or if they intend to participate in any other study using an investigational agent or procedure during participation in the LTE. Investigators who have questions regarding subject eligibility for the LTE should contact the sponsor.

#### 4.3. Prohibitions and Restrictions

Potential subjects must be willing and able to adhere to the following prohibitions and restrictions during the course of the study to be eligible for participation:

1. Subjects must agree to follow the contraceptive requirements noted in the Inclusion Criteria.
2. Subjects must not receive ustekinumab outside of this protocol or participate in any other clinical study with an investigational agent while in this study, and must terminate study participation if they do. A subject who intends to participate in any other clinical study with an investigational agent should undergo an early termination visit before he or she withdraws from study participation.
3. Subjects must agree not to receive a live virus or live bacterial vaccination, including a BCG vaccination, during the study and for 12 months after receiving the last dose of study agent for BCG vaccination or 16 weeks for other live vaccines.

A complete list of prohibited therapies is provided in Section 8.2.

### 5. TREATMENT ALLOCATION AND BLINDING

#### 5.1. Treatment Allocation

Central randomization will be implemented in this study. Subjects will be randomly assigned to 1 of 2 treatment groups (1:1 ratio) in Part I and to 1 of 5 treatment groups (1:1:1:1:1 ratio) in Part II, based on a computer-generated randomization schedule prepared before the study by or under the supervision of the sponsor. Each of the 2 studies will have separate randomizations. Each randomization will be balanced by using randomly permuted blocks and will be stratified by baseline CDAI score (≤300 or >300) and SNP-positive status (yes or no) (Part II will also be stratified by Bio-IR status [yes or no]). The interactive web response system (IWRS) will assign a unique treatment code, which will dictate the treatment assignment and matching study drug kit for the subject. The requestor must use his or her own user identification and personal identification number when contacting the IWRS, and will then give the relevant subject details to uniquely identify the subject.

#### 5.2. Blinding

To maintain the study blind, the study agent container will have a label containing the study name and medication number or syringe number. The label will not identify the study agent in the container. The medication number or syringe number will be entered in the case report form (CRF) when the drug is dispensed. The study agents will be identical in appearance and packaging.

Planned efficacy and safety evaluations will be performed after the following planned DBLs (additional DBLs may occur and would be described in the SAP):

- **Interim analysis lock for Part I:** Occurs when 100 randomized subjects (Bio-IR and Bio-NF) in Part I have completed their Week 12 visit or have terminated their study participation before Week 12.
- **Week 12 DBL for Part I:** Occurs when all subjects in Part I have completed their Week 12 visit or have terminated their study participation before Week 12.
- **Final DBL for Part I:** Occurs when all Part I subjects have completed their final efficacy and safety visit or have terminated their study participation before the final efficacy and safety visit.
- **Week 12 DBL for Part II:** Occurs when all Part II subjects have completed their Week 12 visit or have terminated their study participation before Week 12.
- **Week 24 DBL for Part II:** Occurs when all Part II subjects have completed their Week 24 visit or have terminated their study participation before Week 24.
- **Final DBL for Part II:** Occurs when all Part II subjects who entered the Part II LTE have completed their final safety visit in the Part II LTE or have terminated their study participation before the final safety visit in the Part II LTE.

At the time of the interim analysis lock for Part I, a limited number of sponsor personnel will become unblinded to treatment assignment. At the time of the Week 12 DBL for Part I and the Week 12 DBL for Part II, the sponsor, except for site monitors, who have interactions with the investigative sites, will become unblinded to treatment assignment. Identification of sponsor personnel who will have access to subject-level data for the interim analysis lock for Part I will be documented before the unblinding. The study blind will be maintained for investigators, site personnel, subjects, and site monitors until the Week 24 analyses have been completed for Part II. This measure will mitigate the potential bias in the remaining investigator and subject assessments.

Data that may potentially unblind the treatment assignment (ie, study drug serum concentrations, anti-drug antibodies, treatment allocation, and study drug preparation/accountability data) will be handled with special care to ensure that the integrity of the blind is maintained and the potential for bias is minimized. In particular, before unblinding, this information will be available only to a limited number of data management staff for purposes of data cleaning, and if applicable, to quality assurance representatives for the purposes of conducting independent drug audits.

The SNP status and postbaseline results of CRP, fecal lactoferrin, and fecal calprotectin tests will be blinded to the investigative site. If an investigative site requests these data, it will be provided to them after the final analyses have been completed.

The designated pharmacists, or other appropriately licensed and authorized personnel, and independent drug monitors will be unblinded to study agent. Placebo infusions/injections will have the same appearance as the ustekinumab infusions/JNJ-64304500 injections. Under no circumstances should unblinded personnel reveal the treatment assignment for a subject.

For bioanalytical purposes, before the PK, anti-drug antibody, and PD bioanalyses are initiated, the unblinded data management team will provide the sponsor bioanalysts with the information about which treatment (JNJ-64304500, ustekinumab, or placebo) the subjects received, but not the dose level to which the subjects are randomized. For the purpose of performing PK, immunogenicity, and PD bioanalyses, bioanalysts in Biologics Clinical Pharmacology at Janssen will be unblinded to treatment-level data (JNJ-64304500, ustekinumab, or placebo) at the time of analyzing serum samples for the determination of drug concentrations, detection of antibodies to study agents, or PD assessments. Samples will be separated based on treatment administered; subject identification and dose given will not be disclosed.

Additionally, a given subject’s treatment assignment may be unblinded to the sponsor, Institutional Review Board/Independent Ethics Committee (IRB/IEC), and site personnel to fulfill regulatory reporting requirements for suspected unexpected serious adverse reactions (SUSARs).

The investigator will not be provided with randomization codes. The codes will be maintained within the IWRS, which has the functionality to allow the investigator to break the blind for an individual subject.

Under normal circumstances, the blind should not be broken until the final analyses have been completed for all subjects. Otherwise, the blind should be broken only if specific emergency treatment/course of action would be dictated by knowing the treatment status of the subject. In such cases, the investigator may in an emergency determine the identity of the treatment by contacting the IWRS. It is recommended that the investigator contact the sponsor or its designee if possible to discuss the particular situation, before breaking the blind. Telephone contact with the sponsor or its designee will be available 24 hours per day, 7 days per week. In the event the blind is broken, the sponsor must be informed as soon as possible. The date, time, and reason for the unblinding must be documented by the IWRS, in the appropriate section of the CRF, and in the source document. The documentation received from the IWRS indicating the code-break must be retained with the subject's source documents in a secure manner so as to not unblind the study site monitor. The investigators are advised not to reveal the study treatment assignment to the study site monitor or to sponsor personnel.

A separate code-break procedure will be available for use by the Janssen Global Medical Safety group to allow for unblinding of individual subjects to comply with specific requests from regulatory or health authorities.

Subjects who have had their treatment assignment unblinded will be discontinued from further study agent administration but should continue to return for scheduled evaluations (Section 10).

### 6. DOSAGE AND ADMINISTRATION

Subcutaneous injections of study agent should be administered at different locations at each visit at which study agent is administered to allow for assessment of potential injection-site reactions.

#### 6.1. Part I

In Part I of the study (Figure 2), all subjects will receive either placebo SC or JNJ-64304500 400 mg SC at Week 0 and placebo SC or JNJ-64304500 200 mg SC at Weeks 2, 4, 6, 8, 10, 12, 14, 16, 18, 20, and 22, with the exception of placebo nonresponders at Week 12. Placebo nonresponders will receive JNJ-64304500 400 mg SC at Week 12 and JNJ-64304500 200 mg SC at Weeks 14, 16, 18, 20, and 22. Study drug concentration in Part I is 100 mg/mL.

To maintain the blind in Part I, all subjects will receive 4 SC injections at Weeks 0 and 12, and 2 SC injections at Weeks 2, 4, 6, 8, 10, 14, 16, 18, 20, and 22.

#### 6.2. Part II

In Part II of the study (Figure 3), subjects will be randomly assigned in equal proportions to receive placebo, 1 of 3 dose regimens of JNJ-64304500, or ustekinumab, as follows:

- Placebo: Placebo SC at Weeks 0, 2, 4, 8, 12, 16, and 20. Placebo nonresponders at Week 12 will receive JNJ-64304500 150 mg SC at Week 12 and JNJ-64304500 75 mg SC at Weeks 14, 16, and 20.
- JNJ-64304500 high dose: 400 mg SC at Week 0 and 200 mg SC at Weeks 2, 4, 8, 12, 16, and 20 (study drug concentration=100 mg/mL).
- JNJ-64304500 middle dose: 150 mg SC at Week 0 and 75 mg SC at Weeks 2, 4, 8, 12, 16, and 20 (study drug concentration=75 mg/mL).
- JNJ-64304500 low dose: 50 mg SC at Week 0 and 25 mg SC at Weeks 2, 4, 8, 12, 16, and 20 (study drug concentration=25 mg/mL).
- Ustekinumab: tiered doses approximating 6 mg/kg IV (Section 3.1.2) at Week 0 and 90 mg SC at Weeks 8 and 16.

Administration of IV study agent at Week 0 should occur over a period of not less than 1 hour. The infusion should be completed within 5 hours of preparation. Detailed instructions on the administration of study agent are provided in the Study Reference Manual.

To maintain the blind in Part II, all subjects will receive 4 SC injections plus an IV infusion at Week 0, 2 SC injections at Weeks 2, 4, 12, and 20, 3 SC injections at Weeks 8 and 16, and one SC injection at Week 14.

#### 6.3. Part II Long-term Extension

After Protocol Amendment 5 is implemented, subjects who complete Part II Week 24 assessments, and who may benefit from continued treatment in the opinion of the investigator, are eligible to enter the Part II LTE. In the Part II LTE, each subject will be eligible to receive up to 52 weeks of additional study drug (for a total of up to 72 weeks of study drug in Part II). The first dose of study drug within the Part II LTE will be administered after the Week 24 assessments have been completed. The dose may be administered on the same day as the Week 24 assessments, or may be administered on a later date, as long as the dose is administered within the Week 24 visit window.

Subjects will continue to receive the same treatment regimen during the Part II LTE that they were receiving between Week 12 and Week 20 in the main study phase of Part II (placebo, high, middle, low dose JNJ-64304500 or ustekinumab). To maintain the study blind, all patients, investigators, and sites will remain blinded to treatment allocation during the Part II LTE until the last subject in the Part II main study phase has completed the Week 24 assessments and the Week 24 data analysis has been completed.

The timing of the Week 24 data analysis is dependent upon the timing and completion of Part II enrollment. An individual subject's unblinding during the Part II LTE will depend upon his/her Part II enrollment date. Therefore, a portion of subjects will complete the entire Part II LTE in a blinded fashion before study unblinding, while other subjects could be unblinded to treatment allocation during their participation in the Part II LTE.

Any subject who withdraws from the Part II LTE prior to study unblinding will return for a final safety follow-up visit 16 weeks after the last dose of study drug. For subjects that remain in the Part II LTE at the time of study unblinding, continuation of study drug during the Part II LTE will be dependent upon treatment allocation as follows:

- **JNJ-64304500:** Subjects will continue to receive the same dose of JNJ-64304500 administered during the Part II main study phase. Subjects will continue to receive study drug q4w, although any additional placebo injections used to maintain the study blind during the blinded phase of the Part II LTE will be discontinued after study unblinding. Following the last scheduled dose of study drug at Week 72, a final safety follow-up visit will be performed at approximately Week 88. For subjects who discontinue study participation prior to Week 72, a final safety follow-up visit will be performed 16 weeks after the last dose of study drug.
- **Placebo:** Subjects receiving placebo who remain in the Part II LTE at the time of study unblinding will have their final safety follow-up visit subsequent to unblinding and will be discontinued from the study. No further follow-up visits will be performed.

- **Ustekinumab:** Subjects receiving ustekinumab will continue to receive it during the Part II LTE until unblinding occurs. After unblinding, based on the treating physician's clinical judgment, subjects may receive ustekinumab in a manner dependent on the country in which they are located:
    - If a subject is not continuing on ustekinumab after unblinding, the subject will need to return for a final safety follow-up visit that will be performed 16 weeks after the final dose of study ustekinumab.
    - If a subject continues on ustekinumab in a country where commercial ustekinumab is available and approved for the treatment of adult Crohn's disease, the treating physician is responsible for providing the subject with commercial ustekinumab, which should be administered according to the approved dosing regimen in each country. The final safety follow-up visit should be performed after unblinding but before receiving the first dose of commercial ustekinumab.
    - If a subject continues on ustekinumab in countries where commercial ustekinumab is not available or approved for adult Crohn's disease, ustekinumab will continue to be provided to subjects through Week 72, followed by a final safety follow-up visit 16 weeks after the last dose of ustekinumab.

For eligible Part II subjects, participation in the Part II LTE is entirely voluntary. Eligible subjects who do not wish to enter the Part II LTE will complete the Part II Week 24 safety and efficacy assessments, followed by the final efficacy and safety assessments at Week 36.

During the Part II LTE, all concomitant medications, including Crohn's disease-specific medications (except for the prohibited medications listed in Section 8.2) may be administered at the discretion of the investigator.

All subjects in the Part II LTE will be assessed according to the Time & Events Schedule (Table 3, Table 4), which includes assessments, AEs, laboratory analyses, and PK and immunogenicity samples.

### 7. TREATMENT COMPLIANCE

Study agent will be administered as an IV infusion or SC injection by qualified staff. The details of each administration will be recorded in the CRF. For IV infusions, this will include date and start and stop times of the IV infusion and volume infused; for SC injections, this will include date and time of SC injection.

### 8. PRESTUDY AND CONCOMITANT THERAPY

Concomitant therapies must be recorded throughout the study, from signing of the informed consent to the last study visit.

All therapies (prescription or over-the-counter medications, including vaccines, vitamins, herbal supplements; non-pharmacologic therapies such as electrical stimulation, acupuncture, special diets, exercise regimens) different from the study drug must be recorded in the CRF. Recorded information will include a description of the type of therapy, treatment period, dosage, route of administration, and indication. Modification of an effective pre-existing therapy should not be made for the explicit purpose of entering a subject into the study.

#### 8.1. Concomitant Medications

Subjects who are receiving oral 5-ASA compounds, oral corticosteroids, conventional immunomodulators (ie, AZA, 6-MP, or MTX), enteral therapy, and/or antibiotics for the treatment of Crohn's disease at baseline should maintain a stable dose for a specified period before baseline, as defined in the Inclusion Criteria (Section 4.1).

With the exception of oral corticosteroids, subjects who are receiving these medications for Crohn's disease at baseline should maintain a stable dose through the final efficacy and safety visit. Corticosteroids must be maintained at baseline doses through Week 12 (see Section 8.1.1).

Randomized subjects **should not initiate** any of the following Crohn's disease-specific medical therapies during their participation in the study:

- Oral or rectal 5-ASA compounds.
- Immunomodulators (ie, AZA, 6-MP, or MTX).
- Oral, parenteral, or rectal corticosteroids.
- Antibiotics as a treatment for Crohn's disease.
- Total (complete) or partial (supplemental) parenteral nutrition administered through an indwelling catheter as a treatment for Crohn's disease.
- Enteral therapy (liquid nutritional formula comprising ≥80% of total caloric intake) as treatment for Crohn's disease.

If the above medications are initiated or medication doses are changed, subjects should continue to attend all study visits and have all assessments. If due to medical necessity in the opinion of the investigator, the above medications are initiated or medication doses are changed, this does not represent a deviation from the study protocol, but may be considered a treatment failure.

After Week 12, subjects may transiently use (ie, for <4 weeks) increased doses of corticosteroids for reasons other than worsening of Crohn's disease (eg, stress doses of corticosteroids for surgery, asthma, adrenocortical insufficiency).

Subjects participating in the Part II LTE may continue receiving concomitant medications they were receiving at the time of Part II Week 24, including aminosalicylates, immunomodulatory agents (including 6-MP/AZA, MTX), enteral therapy, and corticosteroids. During the Part II LTE, these medications may be initiated, discontinued, or dose adjusted at the investigator's discretion.

##### 8.1.1. Corticosteroid Tapering

In Part I and Part II, subjects who are receiving corticosteroids at Week 0 and who are in clinical response at Week 12 are encouraged to initiate corticosteroid tapering at the Week 12 visit, using the tapering schedules described below. Other subjects should remain on their original corticosteroid dose through Week 24 unless a change in corticosteroid dose is considered medically necessary (eg, for corticosteroid side effects).

For subjects who experience a worsening of disease activity while tapering, further dose decreases may be suspended and/or their oral corticosteroid dose increased if deemed necessary by the investigator. The oral corticosteroid dose, however, may not be increased above the baseline dose unless due to medical necessity (Section 8.1).

During the Part II LTE, corticosteroids may be initiated, discontinued, or dose adjusted at the investigator's discretion.

**Tapering Schedule for Oral Prednisone (or Equivalent)**

- Dose >15 mg/day prednisone or equivalent: taper daily dose by 5 mg/week until receiving 10 mg/day, then continue tapering at 2.5 mg/week until 0 mg/day.
- Dose 11 to 15 mg/day prednisone or equivalent: taper daily dose to 10 mg/day for 1 week, then continue tapering at 2.5 mg/week until 0 mg/day.
- Dose ≤10 mg/day prednisone or equivalent: taper daily dose by 2.5 mg/week until 0 mg/day.

**Tapering Schedule for Oral Budesonide or Beclomethasone**

- Tapering of budesonide or beclomethasone dipropionate should follow local clinical practice.

#### 8.2. Prohibited Medications

Randomized subjects must not initiate any of the following prohibited medications at any time during Part I, Part II, or the Part II LTE:

- Immunomodulatory agents other than 6-MP/AZA or MTX (including but not limited to 6-TG, cyclosporine, tacrolimus, sirolimus, mycophenolate mofetil, tofacitinib, and other JAK inhibitors).
- Immunomodulatory biologic agents (including but not limited to tumor necrosis factor [TNF] antagonists, natalizumab, abatacept, vedolizumab).
- Experimental Crohn's disease medications (including but not limited to thalidomide, briakinumab, traficet, brodalumab [AMG 827]). Ustekinumab is permitted in Part II of this protocol only in subjects randomly assigned to ustekinumab and only as stipulated per protocol.

Because protection of human research subjects is paramount, it is recognized that initiating such therapies may rarely be required due to medical necessity. However, initiation of these prohibited medications should be documented as a deviation from the study protocol, and subjects will be discontinued from receiving further study agent.

Subjects must not participate in any other clinical study with an investigational agent while in this study and must terminate study participation if they do. A subject who intends to participate in any other clinical study with an investigational agent should undergo an early termination visit before he or she withdraws from study participation.

The sponsor must be notified in advance (or as soon as possible thereafter) of any instances in which prohibited therapies are administered.

### 9. STUDY EVALUATIONS

#### 9.1. Study Procedures

##### 9.1.1. Overview

The Time and Events Schedules summarize the frequency and timing of efficacy, PK, immunogenicity, PD, pharmacogenomic, medical resource utilization, and safety measurements applicable to this study (Table 1, Table 2, Table 3, Table 4).

It is recommended that all visit-specific PRO assessments be conducted before any tests, procedures, or other consultations to prevent influencing subject perceptions.

The maximum amount of blood drawn from each subject in this study will be approximately 330 mL.

##### 9.1.2. Screening Period

At the screening visit, written informed consent must be obtained from the subject for this study by the principal investigator or designee before performing any protocol-specific procedure. Procedures to be performed at the screening visit are outlined in the Time and Events Schedules (Table 1, Table 2).

The CDAI diary will be completed by subjects during the screening period. The investigator or appropriate site personnel will use the hematocrit value obtained during screening to calculate the CDAI score at Week 0.

The screening period should be a minimum of 7 days to allow for collection of CDAI data. Subjects who are rescreened do not need to have a minimum of 7 days, provided that they have enough data to support CDAI calculation.

Women of childbearing potential must have a negative serum β-hCG pregnancy test result at screening. Subjects must be reminded that they are required to use a highly effective method of contraception during the study (as described in Inclusion Criterion 13) and must continue taking such precautions for 16 weeks after receiving the last administration of study agent. The method(s) of contraception used by each subject must be documented.

Subjects must undergo testing for TB (Attachment 3) and their medical history assessment must include specific questions about a history of TB or known occupational or other personal exposure to individuals with active TB. The subject should be asked about past testing for TB, including chest radiograph results and responses to tuberculin skin or other TB testing.

Subjects with a negative QuantiFERON-TB test result (and a negative tuberculin skin test result in countries in which the QuantiFERON-TB test is not approved/registered or the tuberculin skin is mandated by local health authorities) are eligible to continue with prerandomization procedures. A tuberculin skin test is recommended but not required for study centers in countries where tuberculin is not available. Subjects with a newly identified positive QuantiFERON-TB (or tuberculin skin) test result must undergo an evaluation to rule out active TB and initiate appropriate treatment for latent TB. Appropriate treatment for latent TB is defined according to local country guidelines for immunocompromised patients. If no local country guidelines for immunocompromised patients exist, US guidelines must be followed, or the subject will be excluded from the study. **Note:** Subjects in countries with high multidrug-resistant TB burden (eg, Brazil, China, India, the Russian Federation, and South Africa) identified with latent TB at screening will be excluded from participating in the study.

A subject whose first QuantiFERON-TB test result is indeterminate should have the test repeated. In the event that the second QuantiFERON-TB test result is also indeterminate, the subject may be randomized without treatment for latent TB if his/her chest radiograph shows no abnormality suggestive of TB (active or old, inactive TB) and the subject has no additional risk factors for TB as determined by the investigator and medical monitor.

**Note:** Subjects in countries with high multidrug-resistant TB burden (eg, Brazil, China, India, the Russian Federation, and South Africa) with a repeat indeterminate QuantiFERON-TB test result will be excluded from participating in the study, unless his/her chest radiograph shows no abnormality suggestive of TB (active or old, inactive TB), there are no additional risk factors for TB as determined by the investigator and medical monitor, and he/she has a negative tuberculin skin test result within the 2 months before baseline, in which case he/she may be randomized without treatment for latent TB.

An assessment of all screening laboratory test results, clinical data, and concomitant medication data will be made by the principal investigator or designee to confirm that the subject satisfies all inclusion criteria and does not violate any exclusion criteria.

##### 9.1.3. Double-Blind Treatment Period

In Part I, the visit window should be ±4 days for each visit. In Part II, the visit window should be ±4 days, from the Week 0 visit up to and including the Week 12 visit, and ±7 days from Week 16 until the end of the study. Procedures to be performed at each visit are outlined in the Time and Events Schedules (Table 1, Table 2). Unless otherwise specified, all procedures are to be conducted before study agent administration.

Study agent should not be administered to a subject with a clinically important, active infection.

Each subject must be instructed to complete the CDAI diary card daily during the study, except during the Part II LTE.

###### 9.1.4. Final Efficacy and Safety Follow-up Visit

Subjects in Part I should complete the final efficacy and safety follow-up visit approximately 16 weeks after the last study agent administration (Table 1). Subjects in Part II who are not continuing into the Part II LTE should complete the final efficacy and safety follow-up visit approximately 16 weeks after receiving the last dose of study drug (Table 2). Subjects in the Part II LTE should complete the final safety follow-up visit approximately 16 weeks after receiving the last dose of study drug (Table 3, Table 4). Placebo subjects who remain in the Part II LTE at the time of study unblinding will have their final safety follow-up visit subsequent to unblinding and will be discontinued from the study.

##### 9.2. Efficacy Evaluations

The CDAI will be the primary tool for assessing disease activity response to JNJ-64304500, along with PRO-2, PRO-3, Bristol stool form scale, abdominal pain based on NRS 0-10 scale, PGIS of Crohn's disease, and PGIC of severity of Crohn's disease. The degree of inflammation will be assessed by measuring serum CRP concentrations. Stool samples will be collected and analyzed to evaluate changes in markers that may reflect JNJ-64304500 or ustekinumab treatment. The well-being of subjects will be measured using the IBDQ and the SF-36. Endoscopic improvement will be assessed by ileocolonoscopy. For subjects with fistulizing disease, fistula closure will also be assessed.

###### 9.2.1. Crohn's Disease Activity Index

The CDAI will be assessed by collecting information on 8 different Crohn's disease-related variables (Attachment 4):[1] extra-intestinal manifestations, abdominal mass, weight, hematocrit, total number of liquid stools, abdominal pain/cramping, use of antidiarrheal drug(s) and/or opiates, and general well-being. The last 4 variables are scored over 7 days by the subject on a diary card. The PRO-2 score is based on the CDAI components of the total number of liquid stools and abdominal pain/cramping. The PRO-3 score, which is also based on the CDAI, comprises the PRO-2 components plus general well-being. Subjects are to complete a daily diary entry and bring the diary to each visit.

###### 9.2.2. Bristol Stool Form Scale

The Bristol stool form scale is a medical aid to classify the form (or consistency) of human feces into 7 categories.[8] It has been used as a research tool to evaluate the effectiveness of treatments for various diseases of the bowel (eg, irritable bowel syndrome).[4] Subjects will complete the Bristol stool form scale as a daily diary entry and bring the diary to each visit up to Week 12.

###### 9.2.3. Abdominal Pain Numerical Rating Scale

The NRS for pain is a unidimensional measure of pain intensity in adults.[5] An 11-point (0-10) NRS will be used to evaluate abdominal pain. The score of 0 represents "no abdominal pain" and the score of 10 represents the "worst possible abdominal pain", with greater scores indicating greater pain severity and intensity. Subjects will select only one number that best reflects their pain at its worst in the past 24 hours. The abdominal pain NRS will be assessed daily. Subjects will complete the NRS at approximately the same time each day before going to bed and bring the diary to each visit.

###### 9.2.4. Patient's Global Impression of Severity of Crohn's Disease

The PGIS of Crohn's disease is a 5-point scale ("None", "Mild", "Moderate", "Severe" and "Very Severe") to rate Crohn's disease intensity. Subjects will rate their PGIS of Crohn's disease at Weeks 0, 4, 8, 12, and 24.

###### 9.2.5. Patient's Global Impression of Change of Severity of Crohn's Disease

The PGIC of severity of Crohn's disease is a 7-point scale ranging from "a lot better now" to "a lot worse now" with a neutral center point ("neither better nor worse") to measure perceived change (improvement or deterioration) in severity of Crohn's disease. Subjects will assess their PGIC of severity of Crohn's disease at Weeks 4, 8, 12, and 24.

###### 9.2.6. C-Reactive Protein

C-reactive protein has been demonstrated to be useful as a marker of inflammation in patients with inflammatory bowel disease (IBD). In Crohn's disease, elevated CRP concentrations have been associated with severe clinical activity, elevated sedimentation rate, and active disease as detected by colonoscopy.[10,11] Blood samples for the measurement of CRP will be collected from all subjects at visits indicated in the Time and Events Schedule. CRP will be assayed using a validated, high sensitivity CRP assay. Results of postbaseline CRP measurement will not be released to the investigators by the central laboratory.

###### 9.2.7. Fecal Lactoferrin and Calprotectin

Fecal lactoferrin and fecal calprotectin have been demonstrated to be sensitive and specific markers in identifying intestinal inflammation and response to treatment in patients with IBD.[2,3,7] Stool samples for fecal lactoferrin and calprotectin concentrations will be collected from all subjects at visits indicated in the Time and Events Schedules. Assays for fecal lactoferrin and calprotectin concentrations will be performed using a validated method. Additional tests may also be performed on the stool samples for additional markers related to intestinal inflammation and treatment response such as the microbiome. Results of postbaseline fecal lactoferrin and calprotectin tests will not be released to the investigators by the central laboratory.

###### 9.2.8. Inflammatory Bowel Disease Questionnaire

The IBDQ[6] is a 32-item self-report questionnaire for subjects with IBD to evaluate the PROs across 4 dimensions: bowel symptoms (loose stools, abdominal pain), systemic symptoms (fatigue, altered sleep pattern), social function (work attendance, need to cancel social events), and emotional function (anger, depression, irritability). Scores range from 32 to 224 with higher scores indicating better outcomes.

##### 9.2.9. 36-Item Short Form Health Survey

The SF-36 was developed to measure the general health status with 8 functional domain scales.

- Limitations in physical functioning due to health problems.
- Limitations in usual role activities due to physical health problems.
- Bodily pain.
- General mental health (psychological distress and well-being).
- Limitations in usual role activities due to personal or emotional problems.
- Limitations in social functioning due to physical or mental health problems.
- Vitality (energy and fatigue).
- General health perception.

Based on the 8 scale scores, the PCS and the MCS can be derived. The scale scores and summary scores are converted into a score from 0 to 100 using a norm-based system where linear transformations are performed to transform scores to a mean of 50 and standard deviations (SD) of 10, based on general US population norms.[12] The concepts measured by the SF-36 are not specific to any age, disease, or treatment group, allowing comparison of relative burden of different diseases and the relative benefit of different treatments.[13]

##### 9.2.10. Fistula Assessment

Subjects will be assessed for fistulas. For subjects with fistulizing disease, fistula closure will be assessed. Enterocutaneous fistulas (eg, perianal and abdominal) will be considered no longer draining (ie, closed) when there is absence of drainage despite gentle compression. Rectovaginal fistulas will be considered closed based on either physical examination or absence of relevant symptoms (eg, passage of rectal material or flatus from the vagina).

##### 9.2.11. Endoscopic Endpoints

Endoscopic improvement will be assessed during endoscopy (ileocolonoscopy). A video ileocolonoscopic examination will be performed to determine the presence or absence of mucosal inflammation and ulceration at screening and Week 12, and Week 24 (optional in Part I), according to the Study Reference Manual provided to each site; if the video ileocolonoscopic examination is not performed on the day of the visit, it must be performed at least 8 days before the Week 0 visit and no more than 8 days before the Week 12 visit. The Week 24 video ileocolonoscopy in Part I is suggested but not required; if performed, it should occur not more than 8 days before the Week 24 visit. The video ileocolonoscopy will not be performed at Week 24 in Part II. Video endoscopies will be assessed by a central facility that will be blinded to treatment group and study visit. A complete video endoscopic examination does not require assessment of the terminal ileum if it cannot be visualized.

The SES-CD score is based on the evaluation of 4 endoscopic components (presence/size of ulcers, proportion of mucosal surface covered by ulcers, proportion of mucosal surface affected by any other lesions, and presence/type of narrowing/strictures) across 5 ileocolonic segments.

Each endoscopic component is scored from 0 to 3 for each segment, and a total score is derived from the sum of all the component scores (range, 0 to 56). The SES-CD score will be evaluated by a central reader.

In addition to the evaluation of the SES-CD score, endoscopic healing, which is traditionally defined as the resolution (absence) of mucosal ulcers in response to a therapeutic intervention, will also be assessed by the central reader.

###### 9.2.12. Histologic Assessment

Histologic assessment will be performed using biopsy samples collected during endoscopy. Biopsy samples will be collected at screening, Week 12, and Week 24 from each of 3 predefined anatomic locations: the terminal ileum, splenic flexure, and rectum. The biopsy samples collected at Week 12 and Week 24 will be obtained near where the biopsy samples at screening were collected from each of the 3 predefined locations. Histologic assessment will be conducted by a central reader who is blinded to treatment groups. The GHAS will be used to evaluate histologic improvement. Additional details will be provided in the Study Reference Manual.

##### 9.3. Pharmacokinetics and Immunogenicity Evaluations

###### 9.3.1. Evaluations

Serum samples used to evaluate the PK and immunogenicity of JNJ-64304500 and ustekinumab (antibodies to JNJ-64304500 and antibodies to ustekinumab) will be collected according to the Time and Events Schedules. Samples collected for analyses of serum concentration of JNJ-64304500 and ustekinumab and antibodies to JNJ-64304500 or ustekinumab may additionally be used to evaluate safety or efficacy aspects that address concerns arising during or after the study period, for further characterization of immunogenicity or for the evaluation of relevant biomarkers. Genetic analyses will not be performed on these serum samples. Subject confidentiality will be maintained. The exact dates and times of sample collection must be recorded on the laboratory requisition form.

At visits where only serum concentration of study agent will be evaluated (ie, no antibodies to study agent will be evaluated), 1 venous blood sample of sufficient volume should be collected, and each serum sample should be divided into 2 aliquots (1 for serum concentration of study agent, and a back-up). At visits where serum concentration of study agent and antibodies to study agent will be evaluated, 1 venous blood sample of sufficient volume should be collected. Each serum sample will be divided into 3 aliquots (1 each for serum concentration of study agent, antibodies to study agent, and a back-up).

###### 9.3.2. Serum Concentration

Serum samples will be analyzed to determine concentrations of JNJ-64304500 and ustekinumab using a validated, specific, and sensitive method by or under the supervision of the sponsor.

##### 9.3.3. Immunogenicity Assessments (Antibodies to Study Agent)

The detection and characterization of antibodies to JNJ-64304500 and ustekinumab will be performed using a validated assay method by or under the supervision of the sponsor. All samples collected for detection of antibodies to JNJ-64304500 or ustekinumab will also be evaluated for JNJ-64304500 or ustekinumab serum concentration to enable interpretation of the antibody data.

Serum samples will be screened for antibodies binding to JNJ-64304500 or ustekinumab and the titer of confirmed positive samples will be reported. Other analyses may be performed to verify the stability of antibodies to JNJ-64304500 or ustekinumab and/or further characterize the immunogenicity of JNJ-64304500 or ustekinumab. Antibodies to JNJ-64304500 or ustekinumab will be evaluated on blood drawn from all subjects according to the Time and Events Schedule. Additionally, samples should also be collected at the final visit for subjects who terminate from the study. These samples will be tested by the sponsor or sponsor's designee.

#### 9.4. Biomarker and Other Pharmacodynamic Evaluations

Biomarker assessments will be made to examine the biological response to treatment and to identify biomarkers that are relevant to JNJ-64304500 treatment and/or Crohn’s disease. Assessments will include the evaluation of relevant biomarkers in serum, whole blood, stool, and mucosal biopsy samples collected according to the Time and Events Schedule.

##### 9.4.1. Serum-based Biomarkers

Blood samples for serum-based biomarker analyses will be collected from all subjects. Assays to be performed may include the following: measurement of proteins associated with the NKG2D pathway, including but not limited to, MICA, MICB, and UBPs 1-6, as well as proteins associated with Crohn’s disease such as serum amyloid A, interferon gamma, or matrix metalloproteinases.

##### 9.4.2. Whole Blood-based Biomarkers

Whole blood samples will be collected from all subjects to study the effect of study agent on RNA expression. Whole blood analyses may also examine RNA expression associated with the pathogenesis of Crohn’s disease. An additional blood sample will be obtained for analysis of the T-cell receptor repertoire.

##### 9.4.3. Biopsy-based Biomarkers

Mucosal biopsy samples will be collected during video ileocolonoscopy to study the effect of study agent on gene and protein expression and for the histologic assessment of disease and healing (refer to Study Reference Manual for further details). Mucosal biopsy analyses may also examine gene and protein expression associated with the pathogenesis of Crohn’s disease.

##### 9.4.4. NKG2D Receptor Occupancy

NKG2D RO assessments will be performed at the time points specified in the Time and Events Schedule. NKG2D RO will be determined using a validated flow cytometry assay.

###### 9.4.5. Immunophenotyping

Immunophenotyping assessments (including NK cells and CD8+ T cells) will be performed at the time points specified in the Time and Events Schedule. Immunophenotyping will be performed using flow cytometry.

##### 9.5. Pharmacogenomic (DNA) Evaluations

All subjects will be tested for the NKG2D SNP rs2255336 and the MICB (NKG2D ligand) SNP rs2239705 at screening. For subjects who have signed a separate ICF, complete genomic testing will be done to search for links of specific genes to disease or response to drug. Only DNA research related to JNJ-64304500 or ustekinumab or to the diseases for which this drug is developed will be performed. A 10 mL blood sample will be collected from all subjects for this testing; in the event of DNA extraction failure, a replacement pharmacogenomic blood sample will be requested from the subject.

Further, a subject may withdraw his/her optional DNA consent for complete genomic testing at any time without affecting their participation in other aspects of the study, or their future participation in the study.

##### 9.6. Medical Resource Usage

Crohn's disease-related hospitalizations and Crohn's disease-related surgeries will be collected in the study.

##### 9.7. Safety Evaluations

Details regarding the DMC are provided in Section 11.11.

Any clinically relevant changes occurring during the study must be recorded on the Adverse Event section of the CRF.

Any clinically significant abnormalities persisting at the end of the study/early withdrawal will be followed by the investigator until resolution or until a clinically stable endpoint is reached.

The study will include the following evaluations of safety and tolerability according to the time points provided in the Time and Events Schedules:

**Adverse Events**

Adverse events will be reported by the subject (or, when appropriate, by a caregiver, surrogate, or the subject's legally acceptable representative) for the duration of the study. Adverse events will be followed by the investigator as specified in Section 12, Adverse Event Reporting.

**Clinical Laboratory Tests**

Blood samples for serum chemistry and hematology will be collected. The investigator must review the laboratory results, document this review, and record any clinically relevant changes occurring during the study in the Adverse Event section of the CRF. Clinical hematology and chemistry laboratory test results will be graded by the National Cancer Institute Common

Terminology Criteria for Adverse Events (NCI-CTCAE) toxicity criteria. The laboratory reports must be filed with the source documents.

The following tests will be performed by the central laboratory unless otherwise specified or approved by the medical monitor:

- **Hematology assessments:** hematocrit, hemoglobin, platelet count, total and differential white blood cell count.
- **Blood chemistry assessments:** ALT, AST, alkaline phosphatase, albumin, total and direct bilirubin, blood urea nitrogen, calcium, chloride, creatinine, phosphate, potassium, total protein, and sodium.

  A medical monitor or delegate and the clinical site will be notified if prespecified abnormal laboratory values defined in the Laboratory Manual are identified in any subject during the conduct of the study.

- **Pregnancy testing:** Female subjects of childbearing potential will undergo a serum β-hCG pregnancy test at screening, and a urine pregnancy test before each study agent administration visit, at an early termination visit, and at the final efficacy and safety visit.
- Serology (HIV antibody, HBsAg, and HCV antibody)
- **Abnormal liver function tests:** If laboratory testing for a subject randomized in the study and receiving study drug reveals an increase of serum aminotransferases (ALT or AST) to >3x the ULN and an increase of bilirubin to >2x ULN, study agent should be suspended immediately. In addition, laboratory tests for ALT, AST, alkaline phosphatase, and total bilirubin should be confirmed by a retest within 24 hours if possible, but no later than 72 hours following notification of test results. See Attachment 5 (Liver Safety Monitoring and Assessment) for additional information on monitoring and assessment of abnormal liver function tests.

##### Electrocardiogram (ECG)

A 12-lead ECG will be performed at screening.

##### Vital Signs

Vital signs including temperature, pulse/heart rate, respiratory rate, and blood pressure will be collected at all study visits before and 30 minutes after each SC study agent administration. In Part II, vital signs will also be measured at Week 0 before the IV infusion, approximately every 30 minutes during the infusion, and twice after completion of the infusion (at approximately 30-minute intervals).

##### Physical Examination

Physical examinations will be performed as specified in the Time and Events Schedules. Height and weight will be recorded at screening; weight will also be recorded at timepoints specified in the Time and Events Schedules.

**Early Detection of Active Tuberculosis**

To aid in the early detection of TB reactivation or new TB infection during study participation, subjects must be evaluated for signs and symptoms of active TB at scheduled visits (refer to Time and Events Schedule). The following series of questions is suggested for use during the evaluation:

- "Have you had a new cough of >14 days' duration or a change in a chronic cough?"
- "Have you had any of the following symptoms:
  - Persistent fever?
  - Unintentional weight loss?
  - Night sweats?"
- "Have you had close contact with an individual with active TB?" (If there is uncertainty as to whether a contact should be considered "close," a physician specializing in TB should be consulted.)

If the evaluation raises suspicion that a subject may have TB reactivation or new TB infection, an immediate investigation should be undertaken, including, where possible, consultation with a physician specializing in TB.

Investigators should be aware that TB reactivation in immunocompromised subjects may present as disseminated disease or with extrapulmonary features. Subjects with evidence of active TB must discontinue further study agent administrations and should be referred for appropriate treatment.

Subjects who experience close contact with an individual with active TB during the conduct of the study must have a repeat chest radiograph, a repeat QuantiFERON-TB test, a repeat tuberculin skin test in countries in which the QuantiFERON-TB test is not approved/registered, and, if possible, referral to a physician specializing in TB to determine the subject's risk of developing active TB and whether treatment for latent TB is warranted. A tuberculin skin test is recommended but not required for study centers in countries where tuberculin is not available. A positive QuantiFERON-TB (or tuberculin skin) test result should be considered detection of latent TB. If the QuantiFERON-TB test result is indeterminate, the test should be repeated as outlined in Section 9.1.2. If recommended, treatment for latent TB must be initiated before or simultaneously with the administration of further study agent. Subjects who discontinue treatment for latent TB prematurely or who are noncompliant with therapy must immediately discontinue further administration of study agent and be encouraged to return for all subsequent scheduled study visits.

**Concomitant Medication Review**

Concomitant medications will be reviewed at each visit.

**Infusion Reactions**

An infusion reaction (Part II of the study) is defined as an AE that occurs during or within 1 hour after the infusion of study agent, with the exception of laboratory abnormalities. Minor infusion reactions may be managed by slowing the rate of the IV infusion and/or treating with antihistamines and/or acetaminophen (paracetamol) as clinically indicated. If an IV infusion of study agent is stopped because of an infusion reaction and the reaction, in the opinion of the investigator, is not severe or does not result in a serious adverse event (SAE; Section 12.3.2), the infusion may be restarted with caution.

**Injection-Site Reaction**

An injection-site reaction (in Part I or Part II) is any adverse reaction at an SC study agent injection-site. Injection sites will be evaluated for reactions and any injection-site reactions will be recorded as an AE.

**Allergic Reactions**

Before any SC injection or IV infusion, appropriately trained personnel and medications must be available to treat allergic reactions, including anaphylaxis. Appropriate medical personnel must be in attendance at the time of the injection or infusion and for at least 30 minutes after the SC injection or for at least 1 hour after the start of the IV infusion.

Appropriate medical personnel must remain in close proximity to the infusion center for the remaining duration of the infusion, and for 1 hour after the end of the infusion in the event that emergency resuscitation is required. All subjects must be observed carefully for symptoms of an allergic reaction (eg, urticaria, itching, hives).

If a mild or moderate allergic reaction is observed, acetaminophen, nonsteroidal anti-inflammatory drugs (NSAIDs), and/or diphenhydramine may be administered. In the case of a severe allergic reaction (eg, anaphylaxis), SC aqueous epinephrine, corticosteroids, respiratory assistance, and other proper resuscitative measures are essential and must be available at the study site where the injections or infusions are being given.

For severe reactions related to an injection or infusion, the subject may be permanently discontinued from further study injections at the discretion of the investigator (Section 10.2).

Subjects who experience serious adverse reactions related to an injection or infusion should be discontinued from further study agent administrations (Section 10.2).

**Subjects who experience reactions after an injection or infusion (Part II) that result in bronchospasm with wheezing and/or dyspnea that requires ventilatory support OR that result in symptomatic hypotension with a decrease in systolic blood pressure >40 mm Hg or blood pressure <90/60 mm Hg will not be permitted to receive additional study agent.**

**Infections**

Study agent administration should not be given to a subject with a clinically important, active infection. Investigators are required to evaluate subjects for any signs or symptoms of infection, and also review subjects' diary cards for signs of infection, at scheduled visits (see Time and Events Schedule). If a subject develops a serious infection, including but not limited to sepsis or pneumonia, discontinuation of study treatment (ie, no further study agent administrations) must be considered.

#### 9.8. Sample Collection and Handling

The actual dates and times of sample collection must be recorded in the CRF or laboratory requisition form.

Refer to the Time and Events Schedule for the timing and frequency of all sample collections.

Instructions for the collection, handling, storage, and shipment of samples are found in the laboratory manual that will be provided. Collection, handling, storage, and shipment of samples must be under the specified, and where applicable, controlled temperature conditions as indicated in the laboratory manual.

### 10. SUBJECT COMPLETION/DISCONTINUATION OF STUDY TREATMENT/ WITHDRAWAL FROM THE STUDY

#### 10.1. Completion

**Main Study Phase (Part I and Part II)**

A subject will be considered to have completed the study if he or she has completed assessments through the final efficacy and safety visit (ie, Week 38 in Part I and Week 36 in Part II), as specified in the Time and Events Schedules for Part I or Part II.

Subjects who prematurely discontinue study treatment for any reason can be considered to have completed the study if they have completed scheduled assessments as follows:

- Subjects who discontinue study agent in Part I before Week 8 should return for the Week 8 and Week 12 visit and a final efficacy and safety follow-up visit, which should occur 16 weeks after receiving the last dose of study drug.
- Subjects who discontinue study agent at or after Week 8 in Part I should complete the Week 12 and Week 24 visits and a final efficacy and safety follow-up visit, which should occur 16 weeks after receiving the last dose of study drug.
- Subjects who discontinue study agent in Part II before Week 12 should return for the Week 12 visit and a final efficacy and safety follow-up visit, which should occur 16 weeks after receiving the last dose of study drug.
- Subjects who discontinue study agent at or after Week 12 in Part II should complete the Week 24 visit and a final efficacy and safety follow-up visit, which should occur 16 weeks after receiving the last dose of study drug.

The final efficacy and safety follow-up visit for subjects who discontinue study agent is approximately 16 weeks after the last administration of study agent in both Part I and Part II.

**Long-term Extension Phase**

A subject will be considered to have completed the long-term extension phase of the study if he or she has completed the final safety follow-up visit in the Part II LTE.

The final safety follow-up visit will occur approximately 16 weeks after last study agent administration for all subjects who received JNJ-64304500 or ustekinumab. If the investigator decides that the patient should continue ustekinumab treatment after completing the Part II LTE, the investigator or the subject's treating physician is responsible for providing the subject with commercial ustekinumab, which should be administered according to the approved dosing regimen in each country. Refer to Section 3.1.4, Part II Long-Term Extension, for details regarding the ustekinumab follow-up visit.

Subjects who received placebo will be considered to have completed the extension phase of the study if they complete 52 weeks of the Part II LTE and complete the final safety follow-up visit in the Part II LTE before study unblinding or if they have been discontinued from the study prior to the final safety visit due to study unblinding. After unblinding of the Part II Week 24 data, subjects who received placebo during the Part II LTE will have their final safety follow-up visit at the time of study unblinding and will then be discontinued from the study. Subjects who prematurely discontinue study treatment for any reason, other than study unblinding, before completion of the study extension will not be considered to have completed the extension phase of the study.

#### 10.2. Discontinuation of Study Treatment

A subject's study treatment must be discontinued if:

- The investigator believes that for safety reasons or tolerability reasons (eg, AE), it is in the best interest of the subject to discontinue study treatment.
- The subject becomes pregnant or plans a pregnancy within the study period or within 16 weeks after the last study agent administration.
- The subject is deemed ineligible according to the following TB screening criteria:
  - A diagnosis of active TB is made.
  - A subject has symptoms suggestive of active TB based on follow-up assessment questions and/or physical examination, or has had recent close contact with a person with active TB, and cannot or will not continue to undergo additional evaluation.
  - A subject undergoing continued evaluation has a chest radiograph with evidence of current active TB and/or a positive QuantiFERON-TB test result and/or a positive tuberculin skin test result in countries in which the QuantiFERON-TB is not approved/registered (recommended but not required for study centers in countries where tuberculin is not available) unless active TB can be ruled out and appropriate treatment for latent TB can be initiated either prior to or simultaneously with the next

administration of study agent and continued to completion. (Note: Study agent must be discontinued for all subjects diagnosed with latent TB in countries with high multidrug-resistant TB burden [eg, Brazil, China, India, the Russian Federation, and South Africa]).

  - A subject receiving treatment for latent TB discontinues this treatment prematurely or is noncompliant with the therapy.

- A serious adverse reaction occurs that is related to an injection or an infusion, including an injection-site or infusion reaction, resulting in bronchospasm with wheezing and/or dyspnea that requires ventilatory support **OR** that results in symptomatic hypotension with a decrease in systolic blood pressure >40 mm Hg or blood pressure <90/60 mm Hg. This may include events of NCI-CTCAE toxicity grade ≥3.
- Adverse events of NCI-CTCAE grade ≥3 will be evaluated by the investigator and the study medical monitor to make a determination on discontinuation of study agent. Discontinuation of study agent should be considered in subjects with worsening Crohn's disease where continuation of the study drug is not in the best interest of the subject.
- Malignancy including squamous cell skin cancer. Consideration may be given to allowing subjects who develop ≤2 basal cell skin cancers that are adequately treated with no evidence of residual disease to continue to receive study agent.
- Total (complete) or partial (supplemental) parenteral nutrition is initiated through an indwelling catheter at any time during the study.
- The initiation of the following protocol-prohibited medications at any time during the study:
  - Immunomodulatory agents other than 6-MP, AZA, or MTX (including but not limited to 6-TG, cyclosporine, tacrolimus, sirolimus, mycophenolate mofetil, tofacitinib, and other JAK inhibitors).
  - Immunomodulatory biologic agents (including but not limited to TNF antagonists, natalizumab, abatacept, vedolizumab).
  - Experimental Crohn's disease medications (including but not limited to thalidomide, briakinumab, traficet, AMG-827). Ustekinumab is permitted in Part II of this study only in subjects randomly assigned to ustekinumab and only as stipulated per protocol.
- A systemic opportunistic infection.
- Severe hepatic function abnormalities as described in Section 9.7.
- The subject (or the subject's representative) withdraws consent for administration of study agent.
- Crohn's disease-related surgeries that represent a lack of efficacy of study agent or will preclude the future ability to assess efficacy through the CDAI.
- Note: Other permitted Crohn's disease-related surgeries (eg, to resolve long-standing complications such as strictures or for symptomatic nonhealing fistulas, in subjects experiencing improvement on study agent) other than minor procedures (eg, placement of a seton or cutaneous drainage of an abscess) should be postponed until after the final efficacy and safety visit, unless necessary to ensure subject well-being and/or safety.

In the event of any serious infection, severe injection-site or infusion reaction, the study drug must be held and dosing may not be resumed until the investigator has discussed the case with the study medical monitor.

A subject will not be automatically withdrawn from the study if he or she has to discontinue treatment before the end of the treatment regimen. If a subject discontinues study treatment for any reason before the end of the treatment period, assessments should be obtained as specified in Section 10.1.

#### 10.3. Withdrawal From the Study

A subject will be withdrawn from the study for any of the following reasons:

- Lost to follow-up
- Withdrawal of consent
- Death
- Sponsor decision (eg, participating in any other clinical study with an investigational agent)

Subjects who terminate study participation will not be required to return for any follow-up assessments; however, these subjects should complete the safety and efficacy evaluations specified for the Early Termination Visit in the appropriate Time and Events Schedule at the time they terminate study participation.

If a subject is lost to follow-up, every reasonable effort must be made by the study site personnel to contact the subject and determine the reason for discontinuation/withdrawal. The measures taken to follow-up must be documented.

When a subject withdraws before completing the study, the reason for withdrawal is to be documented in the CRF and in the source document. Study agent assigned to the withdrawn subject may not be assigned to another subject. Subjects who withdraw will not be replaced.

#### 10.4. Withdrawal From the Use of Research Samples

A subject who withdraws from the study will have the following options regarding the optional research samples:

- The collected samples will be retained and used in accordance with the subject's original separate informed consent for optional research samples.
- The subject may withdraw consent for optional research samples, in which case the samples will be destroyed and no further testing will take place. To initiate the sample destruction process, the investigator must notify the sponsor study site contact of withdrawal of consent for the optional research samples and to request sample destruction. The sponsor study site contact will, in turn, contact the biomarker representative to execute sample destruction. If requested, the investigator will receive written confirmation from the sponsor that the samples have been destroyed.

**Withdrawal From the Optional Research Samples While Remaining in the Main Study**

The subject may withdraw consent for optional research samples while remaining in the study. In such a case, the optional research samples will be destroyed. The sample destruction process will proceed as described above.

**Withdrawal From the Use of Samples in Future Research**

The subject may withdraw consent for use of samples for research (refer to Section 16.2.5, Long-Term Retention of Samples for Additional Future Research). In such a case, samples will be destroyed after they are no longer needed for the clinical study. Details of the sample retention for research are presented in the main ICF and in the separate ICF for optional research samples.

#### 10.5. Withdrawal From Pharmacogenomic Research Only

The subject may withdraw from the optional consent for pharmacogenomic research related to the drug while remaining in the clinical study. As noted in Section 9.5, however, any testing to be performed on all subjects consented as part of the main study can only be withdrawn if the subject withdraws from the study. In the case of withdrawal from the optional pharmacogenomics testing and request for sample destruction, the investigator must notify the sponsor site contact to request sample destruction. The sponsor site contact will, in turn, contact the biomarker representative to execute sample destruction. If requested, the investigator will receive written confirmation from the sponsor that the sample has been destroyed.

### 11. STATISTICAL METHODS

Statistical analysis will be done by the sponsor or under the authority of the sponsor. A general description of the statistical methods to be used to analyze the efficacy and safety data is outlined below. Specific details will be provided in the SAP.

Descriptive statistics (eg, mean, median, SD, interquartile range, minimum, and maximum) will be used to summarize continuous variables. Counts and percentages will be used to summarize categorical variables. Graphic data displays (eg, line plots) may also be used to summarize the data.

Analyses suitable for categorical data (eg, chi-square tests or Cochran-Mantel-Haenszel chi-square tests as appropriate) will be used to compare the proportions of subjects achieving selected endpoints (eg, clinical remission). In cases of rare events, Fisher's exact test will be used for treatment comparisons. Continuous response parameters will be compared using an analysis of variance or covariance (ANCOVA) model on the van der Waerden normal scores.

All statistical testing will be performed at a significance level of 0.05 (2-sided) unless otherwise specified. Nominal p-values will be displayed for all treatment comparisons.

#### 11.1. Sample Size Determination

Sample size calculations for Part I were determined by the power to detect a significant difference in the change from baseline in the CDAI score at Week 8 (primary endpoint for Part I) between JNJ-64304500 and placebo using a 2-sample t-test. Sample size calculations for Part II were determined by the power to detect a dose-response signal for the change from baseline in CDAI at Week 12 (primary endpoint for Part II) using the Multiple Comparison Procedures with modeling techniques (MCP-Mod) method.

##### 11.1.1. Sample Size in Part I

Part I is powered based on the combined data from Study 1 and Study 2. Study 1 and Study 2 were not individually powered to detect differences between JNJ-64304500 and placebo.

The assumptions for the sample size calculations in Part I were based on data from CNTO1275CRD3001 (a study conducted by the sponsor in subjects with Crohn's disease who had failed or were intolerant to TNF-antagonist therapy) and CNTO1275CRD3002 (a study conducted by the sponsor in subjects with Crohn's disease who had failed or were intolerant to corticosteroids or immunomodulators but who had not failed TNF-antagonist therapy). In CNTO1275CRD3001, the mean CDAI change from baseline at Week 8 was -25.1 (SD=91.41) and -78.7 (SD=91.79) for the placebo and ustekinumab 6 mg/kg groups, respectively. These assumptions incorporated the impact of 6% of subjects being noncompleters (in CNTO1275CRD3001). In CNTO1275CRD3002, the mean CDAI change from baseline at Week 8 was -66.3 (SD=97.81) and -116.3 (SD=102.88) for the placebo and ustekinumab 6 mg/kg groups, respectively. These assumptions incorporated the impact of 4% of subjects being noncompleters (in CNTO1275CRD3002).

For Part I, assuming the mean CDAI change from baseline at Week 8 is -98 in the JNJ-64304500 group and -46 in the placebo group with a common SD of 96 (these values are derived assuming 1:1 ratio of Bio-IR and Bio-NF subjects), 60 subjects per treatment group (a total of 120 subjects) will provide 84% power to detect a treatment difference between JNJ-64304500 and placebo at an overall Type 1 error of 0.05 (2-sided; Table 6).

**Table 6: Power to detect a treatment difference and sample size combinations at an overall Type 1 error of 0.05 (2-sided) for Part I**

| Sample size per group | Placebo | JNJ-64304500 | Difference | Power* |
|---|---|---|---|---|
| **Mean change from baseline in CDAI score at Week 8** | | | | |
| Based on data from CNTO1275CRD3001 and CNTO1275CRD3002 (assuming 1:1 of Bio-IR and Bio-NF subjects) | | | | |
| 55 | -46 | -68 | 32 | 41% |
| 55 | -46 | -78 | 42 | 62% |
| 55 | -46 | -98 | 52 | 80% |
| 55 | -46 | -108 | 62 | 92% |
| 55 | -46 | -118 | 72 | 97% |
| 60 | -46 | -68 | 32 | 44% |
| 60 | -46 | -78 | 42 | 66% |
| 60 | -46 | -98 | 52 | 84% |
| 60 | -46 | -108 | 62 | 94% |
| 60 | -46 | -118 | 72 | 98% |

*Assuming a standard deviation of 96 (derived from CNTO1275CRD3001 and CNTO1275CRD3002) for each group.

##### 11.1.2. Sample Size in Part II

The assumptions for the sample size calculations in Part II were based on the Part I Week 12 analysis results. For Bio-IR subjects in Part I, the mean CDAI change from baseline at Week 12 was -30.9 (SD=85.15) and -77.4 (SD=103.68) for the placebo and JNJ-64304500 groups, respectively. For Bio-NF subjects in Part I, the mean CDAI change from baseline at Week 12 was -70.9 (SD=93.59) and -144.7 (SD=92.50) for the placebo and JNJ-64304500 groups, respectively.

For Part II, assuming the mean CDAI change from baseline at Week 12 is -111 in the JNJ-64304500 high dose group and -51 in the placebo group (these values are derived assuming a 1:1 ratio of Bio-IR and Bio-NF subjects) with a common SD of 102, 50 subjects per treatment group will provide a mean power of 85% to detect a dose-response signal for the change from baseline in CDAI at Week 12 based on 7 candidate dose-response models (to be detailed in the SAP) at an overall Type 1 error rate of 0.05 (2-sided). Fifty subjects per treatment group will also provide 83% power to detect a treatment difference between the JNJ-64304500 treatment group with the highest dose and the placebo treatment group for change from baseline in CDAI at Week 12 based on a 2-sample t-test at a Type I error rate of 0.05 (2-sided; Table 7). This will result in a total sample size of 250 subjects in Part II (incorporating an additional 50 subjects for the ustekinumab treatment group).

**Table 7: Power to detect a treatment difference and sample size combinations at an overall Type 1 error of 0.05 (2-sided) for Part II**

| Sample size per group | Placebo | JNJ-64304500 high dose | Difference | Power* |
|---|---|---|---|---|
| **Mean change from baseline in CDAI score at Week 12** | | | | |
| Based on data from Part I (assuming 1:1 of Bio-IR and Bio-NF subjects) | | | | |
| 45 | -51 | -91 | 40 | 45% |
| 45 | -51 | -101 | 50 | 63% |
| 45 | -51 | -111 | 60 | 79% |
| 45 | -51 | -121 | 70 | 90% |
| 45 | -51 | -131 | 80 | 96% |
| 50 | -51 | -91 | 40 | 49% |
| 50 | -51 | -101 | 50 | 68% |
| 50 | -51 | -111 | 60 | 83% |
| 50 | -51 | -121 | 70 | 92% |
| 50 | -51 | -131 | 80 | 97% |

*Assuming a standard deviation of 102 (based on JNJ-64304500 group at Week 12) for each group.

#### 11.2. Efficacy Analyses

This protocol is comprised of 2 separate parts (Part I [Study 1: Bio-IR subjects and Study 2: Bio-NF subjects] and Part II [Bio-IR and Bio-NF subjects]). Part I and Part II will be analyzed separately with separate Type I error control for the primary endpoint (at the 0.05 level of significance). The other endpoints within each part will not be controlled for multiplicity.

For each part, the analysis set is all randomized subjects who received study agent. Efficacy analyses will be based on a modified intent-to-treat principle. Therefore, the efficacy data for each subject who received study agent will be analyzed according to the assigned treatment regardless of the actual treatment received.

##### 11.2.1. Part I

###### 11.2.1.1. Primary Endpoint Analysis

The primary endpoint for Part I is the change from baseline in the CDAI score at Week 8.

The change from baseline in the CDAI score will be compared between the JNJ-64304500 treatment group and the placebo treatment group for all subjects in Part I. For the comparison, an ANCOVA model on the van der Waerden normal scores will be used with treatment as a fixed factor and baseline CDAI score, SNP-positive status (yes or no), and Bio-IR status (yes or no) as covariates. For this analysis, treatment failure rules and missing data rules as specified in Section 11.2.2.1 will be applied.

Part I will be considered to be positive if a significant improvement is detected in the change from baseline in the CDAI score at Week 8 in the JNJ-64304500 group compared with the placebo group at the 0.05 level of significance.

###### 11.2.1.2. Other Efficacy Endpoint Analyses

The following endpoints will be compared between the JNJ-64304500 treatment group and the placebo treatment group for all subjects in Part I and by Bio-IR status (yes or no):

- Change in CDAI from baseline at all postbaseline visits.
- Clinical remission based on CDAI at all postbaseline visits.
- Clinical response based on CDAI at all postbaseline visits.
- Change in PRO-2 from baseline at all postbaseline visits.
- Change in abdominal pain score (daily average based on the CDAI assessment) from baseline at all postbaseline visits.
- Change in stool frequency score (daily average based on the CDAI assessment) from baseline at all postbaseline visits.
- Clinical remission based on PRO-2 at all postbaseline visits.
- Clinical response based on PRO-2 at all postbaseline visits.
- Change in PRO-3 from baseline at all postbaseline visits.
- Clinical remission based on CDAI at Week 24 among subjects in clinical response at Week 12.
- Clinical remission based on CDAI at Week 24 among subjects in clinical remission at Week 12.
- Change in SES-CD score from baseline at Weeks 12 and 24.

- Endoscopic improvement at Weeks 12 and 24 based on a reduction from baseline in SES-CD score ≥3.
- At least 50% improvement from baseline in SES-CD at Weeks 12 and 24.
- Endoscopic healing (defined as the absence of mucosal ulcerations) at Weeks 12 and 24.
- Endpoint(s) for histologic assessment based on the GHAS (to be detailed in the SAP).
- Fistula response at all postbaseline visits, defined as a ≥50% reduction from baseline in the number of draining fistulas.
- Endpoint(s) based on Bristol stool form scale (to be detailed in the SAP).
- Change in abdominal pain from baseline at all postbaseline visits based on a 0-10 NRS.
- Change in IBDQ score from baseline at Weeks 8, 12, and 24.
- Clinical remission based on IBDQ (≥170) at Weeks 8, 12, and 24.
- A ≥16-point improvement in IBDQ from baseline at Weeks 8, 12, and 24.
- Change from baseline in the PCS and MCS scores of the SF-36 at Weeks 8, 12, and 24.
- A ≥5-point improvement in PCS or MCS scores of the SF-36 at Weeks 8, 12, and 24.
- Change in biomarkers (CRP, fecal calprotectin, fecal lactoferrin) from baseline at Weeks 8, 12, and 24.
- Clinical remission based on CDAI at Week 12 by SNP status. Subjects who are positive in at least 1 of 2 SNPs (NKG2D or MICB) will be considered to be SNP-positive.

  Other efficacy endpoints may be examined by SNP status (to be detailed in the SAP).

##### 11.2.2. Part II

###### 11.2.2.1. Primary Endpoint Analysis

The primary endpoint is the change from baseline in the CDAI score at Week 12.

A unified strategy that combines Multiple Comparison Procedures with modeling (MCP-Mod) techniques, will be used to analyze the dose-response relationship for the JNJ-64304500 doses (the efficacy measurement for the dose-response analysis is the change from baseline in the CDAI score at Week 12). This approach consists of 2 major steps. The first step consists of testing the dose-response signal via multiple contrast tests while controlling the overall Type 1 error. If a dose-response signal is detected, the second step is to select a model that best describes the observed data and use it to estimate adequate doses with associated precision. The details of the dose-response analysis will be provided in the SAP.

Part II will be considered positive if a dose-response signal for the primary endpoint is detected.

In addition to the dose-response analysis, pairwise comparisons of the JNJ-64304500 treatment groups versus the placebo group will be performed for the change from baseline in the CDAI score at Week 12; these comparisons will not be adjusted for multiplicity. For these comparisons, an ANCOVA model on the van der Waerden normal scores will be used with treatment as a fixed factor and baseline CDAI score, SNP-positive status (yes or no), and Bio-IR status (yes or no) as covariates. Pairwise comparisons of the ustekinumab treatment group with the JNJ-64304500 treatment groups or with placebo are not planned; however, summary statistics will be provided for the ustekinumab treatment group.

For the analyses described above, subjects who meet 1 or more treatment failure rules before Week 12 will have their baseline value for the CDAI score carried forward to Week 12. Subjects who have any of the following events before the Week 12 visit will be considered to be treatment failures for the primary endpoint analysis, regardless of the actual CDAI score:

- Specified changes in concomitant Crohn's disease medications (to be detailed in the SAP).
- A Crohn's disease-related surgery (with the exception of drainage of an abscess or seton placement).
- Discontinuation of study agent due to lack of efficacy or due to an AE of worsening Crohn's disease.

In addition, subjects who do not return for evaluation or have insufficient data to calculate their CDAI score at Week 12 (ie, <4 components of the CDAI are available) will have their last available CDAI score carried forward for Week 12.

To examine the robustness of the primary endpoint analysis, sensitivity analyses of the primary endpoint will be conducted using different missing data approaches; these analyses will be described in the SAP. In addition, sensitivity analyses excluding subjects who do not meet predefined threshold values of stool frequency and abdominal pain at study entry will be performed for the primary endpoint; the threshold values and analyses will also be described in the SAP.

To evaluate the consistency of the efficacy, subgroup analysis of the primary endpoint by Bio-IR status (yes or no) will be performed.

###### 11.2.2.2. Major Secondary Endpoint Analyses

The major secondary endpoints are:

- Clinical remission at Week 12 as measured by CDAI (CDAI <150).
- Clinical response at Week 12 as measured by CDAI (≥100-point reduction from baseline in CDAI or CDAI <150).
- Change in PRO-2 from baseline at Week 12.
- Clinical remission at Week 12 as measured by PRO-2 (PRO-2 <75).
- Clinical response at Week 12 as measured by PRO-2 (≥50-point reduction from baseline in PRO-2 or PRO-2 <75).
- Change in SES-CD from baseline at Week 12.

The major secondary endpoints of clinical remission and clinical response at Week 12 (defined by either CDAI or PRO-2) will be compared between each of the JNJ-64304500 treatment groups and the placebo group using the Cochran-Mantel-Haenszel chi-square test (2-sided) stratified by baseline CDAI score (≤300 or >300), SNP-positive status (yes or no), and Bio-IR status (yes or no). In addition, for the endpoint of clinical remission at Week 12 as measured by CDAI, the MCP-MOD strategy will be used to examine the dose-response relationship for the JNJ-64304500 doses.

Subjects who meet 1 or more treatment failure rules (as specified for the primary endpoint) before Week 12 will be considered not to be in clinical remission or clinical response. Subjects who have a missing CDAI score (ie, <4 components of the CDAI score) at Week 12 will be considered not to be in clinical remission or clinical response, as measured by the CDAI score. Subjects who have a missing PRO-2 score (ie, at least one component score of the PRO-2 is missing) at Week 12 will be considered not to be in clinical remission or clinical response as measured by the PRO-2 score.

The change in PRO-2 from baseline at Week 12 will be compared between each of the JNJ-64304500 treatment groups and the placebo group using an ANCOVA model on the van der Waerden normal scores with treatment as a fixed factor and baseline PRO-2 score, SNP-positive status (yes or no), and Bio-IR status (yes or no) as covariates.

Subjects who meet 1 or more treatment failure rules before Week 12 will have their baseline PRO-2 score carried forward to Week 12. Subjects who do not return for evaluation or who have a missing PRO-2 score at Week 12 will have their last available PRO-2 score carried forward to Week 12.

The change in SES-CD score from baseline at Week 12 will be compared between each of the JNJ-64304500 treatment groups and the placebo group using an ANCOVA model on the van der Waerden normal scores with treatment as a fixed factor and baseline SES-CD score, SNP-positive status (yes or no), and Bio-IR status (yes or no) as covariates. Data-handling rules for the SES-CD score will be provided in the SAP.

For the major secondary endpoints, pairwise comparisons of the ustekinumab treatment group with the JNJ-64304500 treatment groups or with placebo are not planned; however, summary statistics will be provided for the ustekinumab treatment group.

Sensitivity analyses excluding subjects who do not meet predefined threshold values of stool frequency and abdominal pain at study entry will be performed for the major secondary endpoints; the threshold values and analyses will be described in the SAP.

To evaluate the consistency of the efficacy, subgroup analyses of the major secondary endpoints by Bio-IR status (yes or no) will be performed.

No adjustments for multiple comparisons will be made for the major secondary endpoints.

###### 11.2.2.3. Other Efficacy Endpoint Analyses

Comparisons between each of the JNJ-64304500 treatment groups and the placebo treatment group will also be made for each of the endpoints specified in Section 11.2.1.2 and for the following endpoints.

- Change in PGIS of Crohn's disease from baseline at Weeks 4, 8, 12 and 24.
- A ≥1-point improvement in PGIS of Crohn's disease from baseline at Weeks 4, 8, 12, and 24.
- Improvement in PGIC of severity of Crohn's disease at Weeks 4, 8, 12, and 24.

Pairwise comparisons of the ustekinumab treatment group with the JNJ-64304500 treatment groups or with placebo are not planned for these endpoints, however summary statistics will be provided for the ustekinumab group.

#### 11.3. Pharmacokinetic Analyses

Descriptive statistics of the serum JNJ-64304500 and ustekinumab concentrations will be calculated at each sampling time point. Serum JNJ-64304500 and ustekinumab concentrations over time will be summarized for each treatment group.

Concentrations below the lowest quantifiable concentration will be treated as zero in the summary statistics.

A population PK analysis approach for JNJ-64304500 using nonlinear mixed-effects modeling will be used to evaluate PK parameters. The influence of important covariates on the population PK parameter estimates may be evaluated. Details will be provided in a population PK analysis plan and the results of the population PK analysis will be presented in a separate technical report.

#### 11.4. Immunogenicity Analyses

The incidence and titers of antibodies to JNJ-64304500 and antibodies to ustekinumab will be summarized for all subjects who receive a dose of JNJ-64304500 or ustekinumab and have appropriate samples for detection of antibodies to JNJ-64304500 or antibodies to ustekinumab (ie, subjects with at least 1 sample obtained after their first dose of JNJ-64304500 or ustekinumab).

A listing of subjects who are positive for antibodies to JNJ-64304500 or ustekinumab will be provided. The maximum titers of antibodies to JNJ-64304500 or ustekinumab will be provided for subjects who are positive for antibodies to JNJ-64304500 or ustekinumab.

The incidence of neutralizing antibodies to JNJ-64304500 or ustekinumab will be summarized for subjects who are positive for antibodies to JNJ-64304500 or ustekinumab and have samples evaluable for neutralizing antibodies to JNJ-64304500 or ustekinumab.

#### 11.5. Biomarker Analyses

Biomarker analyses will characterize the effects of JNJ-64304500 on the measured biomarkers to identify biomarkers relevant to treatment and to determine if these biomarkers can predict response to JNJ-64304500. Biomarker analyses of ustekinumab will be performed as comparisons but not to identify novel biomarkers for ustekinumab.

Results of serum, whole blood analyses, stool, and mucosal biopsy analyses will be reported in separate technical reports.

#### 11.6. Pharmacokinetic/Pharmacodynamic Analyses

The relationship between serum concentrations of JNJ-64304500 and PD and/or clinical endpoints will be examined.

NKG2D RO (%) over time will be summarized for each treatment group.

The absolute numbers and percentages of peripheral blood NK cells and T cells (including CD4+ and CD8+) over time will be summarized for each treatment group.

#### 11.7. Pharmacogenomic Analyses

Exploratory genetic analyses on DNA collection from subjects who signed the optional DNA consent will be presented in a separate technical report.

#### 11.8. Medical Resource Utilization

Crohn's disease-related hospitalizations and Crohn's disease-related surgeries will be descriptively summarized by treatment group.

#### 11.9. Safety Analyses

**Adverse Events**

The verbatim terms used in the CRF by investigators to identify AEs will be coded using the Medical Dictionary for Regulatory Activities. Treatment-emergent AEs are AEs with onset during the treatment phase or that are a consequence of a pre-existing condition that has worsened since baseline. All reported treatment-emergent AEs will be included in the analysis. For each AE, the percentage of subjects who experience at least 1 occurrence of the given event will be summarized by treatment group.

The following analyses of AEs will be used to assess the safety of subjects:

- The incidence and type of AEs.
- The incidence and type of SAEs.
- The incidence and type of reasonably related AEs.
- The incidence and type of AEs leading to discontinuation of study agent.
- The incidence and type of infections.

- The incidence and type of infusion or injection-site reactions.

Summaries, listings, datasets, or subject narratives may be provided, as appropriate, for those subjects who die, who discontinue study agent due to an AE, or who experience a severe or a serious AE.

**Clinical Laboratory Tests**

The following summaries of clinical laboratory tests will be used to assess the safety of subjects:

- Laboratory parameters and change from baseline in laboratory parameters (hematology and chemistry).
- Summary of maximum NCI-CTCAE toxicity grade for postbaseline laboratory values (hematology and chemistry).

Listings of subjects with any abnormal postbaseline laboratory values of CTCAE grade ≥2 will also be provided.

#### 11.10. Interim Analysis

An interim analysis is planned in Part I when 100 randomized subjects (Study 1: Bio-IR and Study 2: Bio-NF) have completed their Week 12 visit or have terminated their study participation before Week 12. This interim analysis will allow for uninterrupted patient enrollment through a seamless transition from Part I to Part II of the trial. As this interim analysis does not affect the conduct or completion of Part I, it will be considered administrative and will not require multiplicity adjustment for the final Part I analysis.

The primary efficacy evaluation is the comparison between JNJ-64304500 and placebo with respect to the change in CDAI from baseline (in the combined Bio-IR and Bio-NF subjects). Other selected efficacy analyses (eg, clinical remission, clinical response, and change in SES-CD) will also been performed; details will be provided in the Interim Analysis Plan.

A sponsor committee independent of the study team will be established to review the interim data and formulate recommended decisions/actions in accordance with predefined decision rules that will be defined in the Interim Analysis Plan.

#### 11.11. Data Monitoring Committee

An external DMC will be established to monitor safety data on an ongoing basis to ensure the continuing safety of the subjects randomized in Part I and Part II. The committee will meet periodically to review interim data. After the review, the DMC will make recommendations regarding the continuation of these studies. The details will be provided in a separate DMC charter.

The DMC will consist of at least one medical expert in the relevant therapeutic area and at least one statistician. The DMC responsibilities, authorities, and procedures will be documented in its charter.

### 12. ADVERSE EVENT REPORTING

Timely, accurate, and complete reporting and analysis of safety information from clinical studies are crucial for the protection of subjects, investigators, and the sponsor, and are mandated by regulatory agencies worldwide. The sponsor has established Standard Operating Procedures in conformity with regulatory requirements worldwide to ensure appropriate reporting of safety information; all clinical studies conducted by the sponsor or its affiliates will be conducted in accordance with those procedures.

#### 12.1. Definitions

##### 12.1.1. Adverse Event Definitions and Classifications

**Adverse Event**

An AE is any untoward medical occurrence in a clinical study subject administered a medicinal (investigational or non-investigational) product. An AE does not necessarily have a causal relationship with the treatment. An AE can therefore be any unfavorable and unintended sign (including an abnormal finding), symptom, or disease temporally associated with the use of a medicinal (investigational or non-investigational) product, whether or not related to that medicinal (investigational or non-investigational) product (definition per International Conference on Harmonisation [ICH]).

This includes any occurrence that is new in onset or aggravated in severity or frequency from the baseline condition, or abnormal results of diagnostic procedures, including laboratory test abnormalities.

Note: The sponsor collects AEs starting with the signing of the ICF (refer to Section 12.3.1, All Adverse Events, for time of last AE recording).

**Serious Adverse Event**

An SAE based on ICH and European Union Guidelines on Pharmacovigilance for Medicinal Products for Human Use is any untoward medical occurrence that at any dose:

- Results in death
- Is life-threatening (ie, the subject was at risk of death at the time of the event; it does not refer to an event that hypothetically might have caused death if it were more severe)
- Requires inpatient hospitalization or prolongation of existing hospitalization
- Results in persistent or significant disability/incapacity
- Is a congenital anomaly/birth defect
- Is a suspected transmission of any infectious agent via a medicinal product
- Is medically important*

*Medical and scientific judgment should be exercised in deciding whether expedited reporting is also appropriate in other situations, such as important medical events that may not be immediately life-threatening or result in death or hospitalization but may jeopardize the subject or may require intervention to prevent one of the other outcomes listed in the definition above. These should usually be considered serious.

**Unlisted (Unexpected) Adverse Event/Reference Safety Information**

An AE is considered unlisted if the nature or severity is not consistent with the applicable product reference safety information. For JNJ-64304500 and for ustekinumab, the expectedness of an adverse event will be determined by whether or not it is listed in the IB for JNJ-64304500 or for ustekinumab, respectively.

**Adverse Event Associated With the Use of the Drug**

An AE is considered associated with the use of the drug if the attribution is possible, probable, or very likely by the definitions listed in Section 12.1.2, Attribution Definitions.

**Adverse Events Associated with the Study Population**

Anticipated events will be recorded and reported as described in Attachment 6.

##### 12.1.2. Attribution Definitions

**Not Related**

An AE that is not related to the use of the drug.

**Doubtful**

An AE for which an alternative explanation is more likely, eg, concomitant drug(s), concomitant disease(s), or the relationship in time suggests that a causal relationship is unlikely.

**Possible**

An AE that might be due to the use of the drug. An alternative explanation, eg, concomitant drug(s), concomitant disease(s), is inconclusive. The relationship in time is reasonable; therefore, the causal relationship cannot be excluded.

**Probable**

An AE that might be due to the use of the drug. The relationship in time is suggestive (eg, confirmed by dechallenge). An alternative explanation is less likely, eg, concomitant drug(s), concomitant disease(s).

**Very Likely**

An AE that is listed as a possible adverse reaction and cannot be reasonably explained by an alternative explanation, eg, concomitant drug(s), concomitant disease(s). The relationship in time is very suggestive (eg, it is confirmed by dechallenge and rechallenge).

##### 12.1.3. Severity Criteria

An assessment of severity grade will be made using the following general categorical descriptors as outlined in Section 12.1.3.1 and by NCI-CTCAE toxicity grade outlined in Section 12.1.3.2.

###### 12.1.3.1. Severity Criteria: General Categorical Descriptors

**Mild**: Awareness of symptoms that are easily tolerated, causing minimal discomfort and not interfering with everyday activities.

**Moderate**: Sufficient discomfort is present to cause interference with normal activity.

**Severe**: Extreme distress, causing significant impairment of functioning or incapacitation. Prevents normal everyday activities.

The investigator should use clinical judgment in assessing the severity of events not directly experienced by the subject (eg, laboratory abnormalities).

**Note**: Seriousness and severity should not be confused. A subject could experience a severe headache that would not qualify as a SAE, while another might experience a mild stroke that, while not severe, would be considered serious.

###### 12.1.3.2. Severity Criteria Based on National Cancer Institute Common Terminology Criteria for Adverse Events Toxicity Grade

An assessment of severity grade will also be made using the following NCI-CTCAE categorical descriptors. The NCI-CTCAE Grade refers to the severity of the AE. The NCI-CTCAE displays Grades 1 through 5 with unique clinical descriptions of severity for each AE based on this general guideline:

**Grade 1:** Mild; asymptomatic or mild symptoms; clinical or diagnostic observations only; intervention not indicated.

**Grade 2:** Moderate; minimal, local or noninvasive intervention indicated; limiting age-appropriate instrumental Activities of Daily Living (ADL)*.

**Grade 3:** Severe or medically significant but not immediately life-threatening; hospitalization or prolongation of hospitalization indicated; disabling; limiting self-care ADL**.

**Grade 4:** Life-threatening consequences; urgent intervention indicated.

**Grade 5:** Death related to AE.

*Instrumental ADL refer to preparing meals, shopping for groceries or clothes, using the telephone, managing money, etc.

**Self-care ADL refer to bathing, dressing and undressing, feeding self, using the toilet, taking medications, and not bedridden.

#### 12.2. Special Reporting Situations

Safety events of interest on a sponsor study drug that may require expedited reporting and/or safety evaluation include, but are not limited to:

- Overdose of a sponsor study drug
- Suspected abuse/misuse of a sponsor study drug
- Accidental or occupational exposure to a sponsor study drug
- Medication error involving a sponsor product (with or without subject/patient exposure to the sponsor study drug, eg, name confusion)
- Exposure to a sponsor study drug from breast-feeding

Special reporting situations should be recorded in the CRF. Any special reporting situation that meets the criteria of an SAE should be recorded on the SAE page of the CRF.

#### 12.3. Procedures

##### 12.3.1. All Adverse Events

All AEs and special reporting situations, whether serious or nonserious, will be reported from the time a signed and dated ICF is obtained until completion of the subject's last study-related procedure, which may include contact for follow-up of safety. Serious AEs, including those spontaneously reported to the investigator within 16 weeks after the last dose of study drug, must be reported using the Serious Adverse Event Form. The sponsor will evaluate any safety information that is spontaneously reported by an investigator beyond the time frame specified in this protocol.

All events that meet the definition of an SAE will be reported as SAEs, regardless of whether they are protocol-specific assessments. Anticipated events will be recorded and reported as described in Attachment 6.

All AEs, regardless of seriousness, severity, or presumed relationship to study drug, must be recorded using medical terminology in the source document and the CRF. Whenever possible, diagnoses should be given when signs and symptoms are due to a common etiology (eg, cough, runny nose, sneezing, sore throat, and head congestion should be reported as "upper respiratory infection"). Investigators must record in the CRF their opinion concerning the relationship of the AE to study therapy. All measures required for AE management must be recorded in the source document and reported according to sponsor instructions.

The sponsor assumes responsibility for appropriate reporting of AEs to the regulatory authorities. The sponsor will also report to the investigator (and the head of the investigational institute where required) all SUSARs. The investigator (or sponsor where required) must report SUSARs to the appropriate IEC/IRB that approved the protocol unless otherwise required and documented by the IEC/IRB. A SUSAR will be reported to regulatory authorities unblinded. Participating investigators and IEC/IRB will receive a blinded SUSAR summary, unless otherwise specified.

For all studies with an outpatient phase, including open-label studies, subjects must be provided with a "wallet (study) card" and instructed to carry this card with them for the duration of the study indicating the following:

- Study number
- Statement, in the local language(s), that the subject is participating in a clinical study
- Investigator's name and 24-hour contact telephone number
- Local sponsor's name and 24-hour contact telephone number (for medical staff only)
- Site number
- Subject number
- Any other information that is required to do an emergency breaking of the blind

##### 12.3.2. Serious Adverse Events

All SAEs occurring during the study must be reported to the appropriate sponsor contact person by study site personnel within 24 hours of their knowledge of the event.

Information regarding SAEs will be transmitted to the sponsor using the Serious Adverse Event Form, which must be completed and signed by a physician from the study site and transmitted to the sponsor within 24 hours. The initial and follow-up reports of an SAE should be made by facsimile (fax).

All SAEs that have not resolved by the end of the study, or that have not resolved upon discontinuation of the subject's participation in the study, must be followed until any of the following occurs:

- The event resolves
- The event stabilizes
- The event returns to baseline, if a baseline value/status is available
- The event can be attributed to agents other than the study drug or to factors unrelated to study conduct
- It becomes unlikely that any additional information can be obtained (subject or health care practitioner refusal to provide additional information, lost to follow-up after demonstration of due diligence with follow-up efforts)

Suspected transmission of an infectious agent by a medicinal product will be reported as an SAE.

Any event requiring hospitalization (or prolongation of hospitalization) that occurs during the course of a subject's participation in a study must be reported as an SAE, except hospitalizations for the following:

- Social reasons in the absence of an AE.
- Hospitalizations not intended to treat an acute illness or AE (eg, social reasons such as pending placement in long-term care facility).
- Surgery or procedure planned before entry into the study (must be documented in the CRF). Note: Hospitalizations that were planned before the signing of the ICF, and where the underlying condition for which the hospitalization was planned has not worsened, will not be considered SAEs. Any AE that results in a prolongation of the originally planned hospitalization is to be reported as a new SAE.

The cause of death of a subject in a study, whether or not the event is expected or associated with the study drug, is considered a SAE.

##### 12.3.3. Pregnancy

All initial reports of pregnancy in female subjects or partners of male subjects must be reported to the sponsor by the study site personnel within 24 hours of their knowledge of the event using the appropriate pregnancy notification form. Abnormal pregnancy outcomes (eg, spontaneous abortion, fetal death, stillbirth, congenital anomalies, ectopic pregnancy) are considered SAEs and must be reported using the Serious Adverse Event Form. Any subject who becomes pregnant during the study must discontinue further study treatment.

Because the effect of the study drug on sperm is unknown, pregnancies in partners of male subjects included in the study will be reported as noted above.

Follow-up information regarding the outcome of the pregnancy and any postnatal sequelae in the infant will be required.

##### 12.3.4. Events of Special Interest

Any newly identified malignancy or case of active TB occurring after the first administration of study agent in subjects participating in this clinical study must be reported by the investigator according to the procedures in Section 9.1.2. Investigators are also advised that active TB is considered a reportable disease in most countries. These events are to be considered serious only if they meet the definition of an SAE.

#### 12.4. Contacting Sponsor Regarding Safety

The names (and corresponding telephone numbers) of the individuals who should be contacted regarding safety issues or questions regarding the study are listed in the Contact Information page(s), which will be provided as a separate document.

### 13. PRODUCT QUALITY COMPLAINT HANDLING

A product quality complaint (PQC) is defined as any suspicion of a product defect related to manufacturing, labeling, or packaging, ie, any dissatisfaction relative to the identity, quality, durability, or reliability of a product, including its labeling or package integrity. A PQC may have an impact on the safety and efficacy of the product. Timely, accurate, and complete reporting and analysis of PQC information from studies are crucial for the protection of subjects, investigators, and the sponsor, and are mandated by regulatory agencies worldwide. The sponsor has established procedures in conformity with regulatory requirements worldwide to ensure appropriate reporting of PQC information; all studies conducted by the sponsor or its affiliates will be conducted in accordance with those procedures.

#### 13.1. Procedures

All initial PQCs must be reported to the sponsor by the study site personnel within 24 hours after being made aware of the event.

If the defect is combined with an SAE, the study site personnel must report the PQC to the sponsor according to the SAE reporting timelines (refer to Section 12.3.2, Serious Adverse Events). A sample of the suspected product should be maintained for further investigation if requested by the sponsor.

#### 13.2. Contacting Sponsor Regarding Product Quality

The names (and corresponding telephone numbers) of the individuals who should be contacted regarding product quality issues are listed in the Contact Information page(s), which will be provided as a separate document.

### 14. STUDY DRUG INFORMATION

#### 14.1. Physical Description of Study Drugs

In Part I and Part II, the JNJ-64304500 supplied for this study is a lyophilized drug product which, upon reconstitution with 1.1 mL of water for injection, contains 100 mg/mL JNJ-64304500 in 34 mM L-histidine, 8.6% (w/v) sucrose, and 0.03% (w/v) polysorbate 80, pH 6.0 in a 10 mL glass vial. It will be manufactured and provided under the responsibility of the sponsor. Refer to the IB for a list of excipients.

Placebo for JNJ-64304500 consists of a 9 mL solution of 34 mM L-histidine, 8.6% (w/v) sucrose, and 0.03% (w/v) polysorbate 80, pH 6.0 in a 10 mL glass vial.

In Part II, ustekinumab 5 mg/mL final vialed product for IV infusion and placebo to match will be supplied as a single-use, sterile solution in 30 mL vials with 1 dose strength (ie, 130 mg in 26 mL nominal volume).

Ustekinumab SC will be supplied as a sterile solution in a single-use prefilled syringe (PFS) at a volume of 1 mL (90 mg dose) that contains ustekinumab 90 mg, L-histidine, L-histidine monohydrochloride monohydrate, sucrose, and polysorbate 80 at pH 6.0 in 1.0 mL nominal volume. No preservatives are present. The needle cover on the PFS contains dry natural rubber (a derivative of latex), which may cause allergic reactions in individuals sensitive to latex. Liquid placebo will be supplied in a 1 mL PFS.

Placebo administrations will have the same appearance as the respective JNJ-64304500 or ustekinumab administrations.

#### 14.2. Packaging

The investigational supplies will be uniquely packaged to assure that they are appropriately managed throughout the supply chain process.

The study drug will be packaged in individual subject kits. Each kit will consist of one vial or one PFS of study agent packaged inside a protective outer carton. Each carton will be uniquely labeled with a medication identification number.

#### 14.3. Labeling

Study drug labels will contain information to meet the applicable regulatory requirements.

#### 14.4. Preparation, Handling, and Storage

All study drug must be stored at controlled temperatures ranging from 2°C to 8°C (36°F to 46°F).

Refer to the pharmacy manual/study site investigational product and procedures manual for additional guidance on study drug preparation, handling, and storage.

#### 14.5. Drug Accountability

The investigator is responsible for ensuring that all study drug received at the site is inventoried and accounted for throughout the study. The study drug administered to the subject must be documented on the drug accountability form. All study drug will be stored and disposed of according to the sponsor's instructions. Study site personnel must not combine contents of the study drug containers.

Study drug must be handled in strict accordance with the protocol and the container label and must be stored at the study site in a limited-access area or in a locked cabinet under appropriate environmental conditions. Unused study drug must be available for verification by the sponsor's study site monitor during on-site monitoring visits. The return to the sponsor of unused study drug will be documented on the drug return form. When the study site is an authorized destruction unit and study drug supplies are destroyed on-site, this must also be documented on the drug return form.

Potentially hazardous materials such as used ampules, needles, syringes, and vials containing hazardous liquids should be disposed of immediately in a safe manner and therefore will not be retained for drug accountability purposes. The immediate destruction of these drug supplies should be documented in the drug accountability records on-site.

Study drug should be dispensed under the supervision of the investigator or a qualified member of the study site personnel, or by a hospital/clinic pharmacist. Study drug will be supplied only to subjects participating in the study. Returned study drug must not be dispensed again, even to the same subject. Study drug may not be relabeled or reassigned for use by other subjects. The investigator agrees neither to dispense the study drug from, nor store it at, any site other than the study sites agreed upon with the sponsor.

### 15. STUDY-SPECIFIC MATERIALS

The investigator will be provided with the following supplies:

- Investigator Site File (includes protocol and IB)
- Investigational Product Binder
- Central Laboratory Manual
- CRF completion instructions
- Patient recruitment materials
- Subject study information
- Informed consent form
- CDAI diary
- Bristol stool form scale diary
- Abdominal pain NRS diary
- IWRS Manual
- Worksheets for data collection

### 16. ETHICAL ASPECTS

#### 16.1. Study-Specific Design Considerations

Potential subjects will be fully informed of the risks and requirements of the study and, during the study, subjects will be given any new information that may affect their decision to continue participation. They will be told that their consent to participate in the study is voluntary and may be withdrawn at any time with no reason given and without penalty or loss of benefits to which they would otherwise be entitled. Only subjects who are fully able to understand the risks, benefits, and potential AEs of the study, and provide their consent voluntarily will be randomized.

The total blood volume to be collected is considered to be an acceptable amount of blood to be collected over this time period from the population in this study based upon the standard of the American Red Cross (1 pint/473 mL of blood for donation).

#### 16.2. Regulatory Ethics Compliance

##### 16.2.1. Investigator Responsibilities

The investigator is responsible for ensuring that the study is performed in accordance with the protocol, current ICH guidelines on Good Clinical Practice (GCP), and applicable regulatory and country-specific requirements.

Good Clinical Practice is an international ethical and scientific quality standard for designing, conducting, recording, and reporting studies that involve the participation of human subjects. Compliance with this standard provides public assurance that the rights, safety, and well-being of study subjects are protected, consistent with the principles that originated in the Declaration of Helsinki, and that the study data are credible.

##### 16.2.2. Independent Ethics Committee or Institutional Review Board

Before the start of the study, the investigator (or sponsor where required) will provide the IEC/IRB with current and complete copies of the following documents (as required by local regulations):

- Final protocol and, if applicable, amendments
- Sponsor-approved ICF (and any other written materials to be provided to the subjects)
- Investigator's Brochure (or equivalent information) and amendments/addenda
- Sponsor-approved subject recruiting materials
- Information on compensation for study-related injuries or payment to subjects for participation in the study, if applicable
- Investigator's curriculum vitae or equivalent information (unless not required, as documented by the IEC/IRB)
- Information regarding funding, name of the sponsor, institutional affiliations, other potential conflicts of interest, and incentives for subjects
- Any other documents that the IEC/IRB requests to fulfill its obligation

This study will be undertaken only after the IEC/IRB has given full approval of the final protocol, amendments (if any, excluding the ones that are purely administrative, with no consequences for subjects, data or study conduct, unless required locally), the ICF, applicable recruiting materials, and subject compensation programs, and the sponsor has received a copy of this approval. This approval letter must be dated and must clearly identify the IEC/IRB and the documents being approved.

Approval for the collection of optional samples for research and for the corresponding ICF must be obtained from the IEC/IRB. Approval for the protocol can be obtained independent of this optional research component.

During the study the investigator (or sponsor where required) will send the following documents and updates to the IEC/IRB for their review and approval, where appropriate:

- Protocol amendments (excluding the ones that are purely administrative, with no consequences for subjects, data or study conduct)
- Revision(s) to ICF and any other written materials to be provided to subjects
- If applicable, new or revised subject recruiting materials approved by the sponsor

- Revisions to compensation for study-related injuries or payment to subjects for participation in the study, if applicable
- New edition(s) of the IB and amendments/addenda
- Summaries of the status of the study at intervals stipulated in guidelines of the IEC/IRB (at least annually)
- Reports of AEs that are serious, unlisted/unexpected, and associated with the study drug
- New information that may adversely affect the safety of the subjects or the conduct of the study
- Deviations from or changes to the protocol to eliminate immediate hazards to the subjects
- Report of deaths of subjects under the investigator's care
- Notification if a new investigator is responsible for the study at the site
- Development Safety Update Report and Line Listings, where applicable
- Any other requirements of the IEC/IRB

For all protocol amendments (excluding the ones that are purely administrative, with no consequences for subjects, data or study conduct), the amendment and applicable ICF revisions must be submitted promptly to the IEC/IRB for review and approval before implementation of the change(s).

At least once a year, the IEC/IRB will be asked to review and reapprove this study, where required.

At the end of the study, the investigator (or sponsor where required) will notify the IEC/IRB about the study completion.

##### 16.2.3. Informed Consent

Each subject must give written consent according to local requirements after the nature of the study has been fully explained. The ICF(s) must be signed before performance of any study-related activity. The ICF(s) that is/are used must be approved by both the sponsor and by the reviewing IEC/IRB and be in a language that the subject can read and understand. The informed consent should be in accordance with principles that originated in the Declaration of Helsinki, current ICH and GCP guidelines, applicable regulatory requirements, and sponsor policy.

Before enrollment in the study, the investigator or an authorized member of the study site personnel must explain to potential subjects the aims, methods, reasonably anticipated benefits, and potential hazards of the study, and any discomfort participation in the study may entail. Subjects will be informed that their participation is voluntary and that they may withdraw consent to participate at any time. They will be informed that choosing not to participate will not affect the care the subject will receive for the treatment of his or her Crohn’s disease. Subjects will be told that alternative treatments are available if they refuse to take part and that such refusal will not prejudice future treatment. Finally, they will be told that the investigator will maintain a subject identification register for the purposes of long-term follow-up if needed and that their records may be accessed by health authorities and authorized sponsor personnel without violating the confidentiality of the subject, to the extent permitted by the applicable law(s) or regulations. By signing the ICF, the subject is authorizing such access, and agrees to allow his or her study physician to recontact the subject for the purpose of obtaining consent for additional safety evaluations, if needed.

The subject will be given sufficient time to read the ICF and the opportunity to ask questions. After this explanation and before entry into the study, consent should be appropriately recorded by means of the subject's personally dated signature. After having obtained the consent, a copy of the ICF must be given to the subject.

Subjects will be asked for consent to provide optional samples for research where local regulations permit). After informed consent for the study is appropriately obtained, the subject will be asked to sign and personally date a separate ICF indicating agreement to participate in the optional research component. Refusal to participate in the optional research will not result in ineligibility for the study. A copy of this signed ICF will be given to the subject.

Subjects must be able to read and write and give informed consent without assistance. A subject who is unable to read or write is not eligible to participate in the study.

##### 16.2.4. Privacy of Personal Data

The collection and processing of personal data from subjects randomized in this study will be limited to those data that are necessary to fulfill the objectives of the study.

These data must be collected and processed with adequate precautions to ensure confidentiality and compliance with applicable data privacy protection laws and regulations. Appropriate technical and organizational measures to protect the personal data against unauthorized disclosures or access, accidental or unlawful destruction, or accidental loss or alteration must be put in place. Sponsor personnel whose responsibilities require access to personal data agree to keep the identity of subjects confidential.

The informed consent obtained from the subject includes explicit consent for the processing of personal data and for the investigator/institution to allow direct access to his or her original medical records (source data/documents) for study-related monitoring, audit, IEC/IRB review, and regulatory inspection. This consent also addresses the transfer of the data to other entities and to other countries.

The subject has the right to request through the investigator access to his or her personal data and the right to request rectification of any data that are not correct or complete. Reasonable steps will be taken to respond to such a request, taking into consideration the nature of the request, the conditions of the study, and the applicable laws and regulations.

Exploratory DNA, PD, biomarker, PK, and immunogenicity research is not conducted under standards appropriate for the return of data to subjects. In addition, the sponsor cannot make decisions as to the significance of any findings resulting from exploratory research. Therefore, exploratory research data will not be returned to subjects or investigators, unless required by law or local regulations. Privacy and confidentiality of data generated in the future on stored samples will be protected by the same standards applicable to all other clinical data.

###### 16.2.5. Long-Term Retention of Samples for Additional Future Research

Samples collected in this study may be stored for up to 15 years (or according to local regulations) for additional research. Samples will only be used to understand JNJ-64304500 and ustekinumab, to understand Crohn's disease, to understand differential drug responders, and to develop tests/assays related to JNJ-64304500 and ustekinumab and Crohn's disease. The research may begin at any time during the study or the post-study storage period.

Stored samples will be coded throughout the sample storage and analysis process and will not be labeled with personal identifiers. Subjects may withdraw their consent for their samples to be stored for research (refer to Section 10.4, Withdrawal From the Use of Research Samples).

###### 16.2.6. Country Selection

This study will only be conducted in those countries where the intent is to launch or otherwise help ensure access to the developed product if the need for the product persists, unless explicitly addressed as a specific ethical consideration in Section 16.1, Study-Specific Design Considerations.

#### 17. ADMINISTRATIVE REQUIREMENTS

##### 17.1. Protocol Amendments

Neither the investigator nor the sponsor will modify this protocol without a formal amendment by the sponsor. All protocol amendments must be issued by the sponsor and signed and dated by the investigator. Protocol amendments must not be implemented without prior IEC/IRB approval, or when the relevant competent authority has raised any grounds for non-acceptance, except when necessary to eliminate immediate hazards to the subjects, in which case the amendment must be promptly submitted to the IEC/IRB and relevant competent authority. Documentation of amendment approval by the investigator and IEC/IRB must be provided to the sponsor. When the changes involve only logistic or administrative aspects of the study, the IEC/IRB (where required) only needs to be notified.

During the course of the study, in situations where a departure from the protocol is unavoidable, the investigator or other physician in attendance will contact the appropriate sponsor representative listed in the Contact Information page(s), which will be provided as a separate document. Except in emergency situations, this contact should be made before implementing any departure from the protocol. In all cases, contact with the sponsor must be made as soon as possible to discuss the situation and agree on an appropriate course of action. The data recorded in the CRF and source documents will reflect any departure from the protocol, and the source documents will describe this departure and the circumstances requiring it.

#### 17.2. Regulatory Documentation

##### 17.2.1. Regulatory Approval/Notification

This protocol and any amendment(s) must be submitted to the appropriate regulatory authorities in each respective country, if applicable. A study may not be initiated until all local regulatory requirements are met.

##### 17.2.2. Required Prestudy Documentation

The following documents must be provided to the sponsor before shipment of study drug to the study site:

- Protocol and amendment(s), if any, signed and dated by the principal investigator
- A copy of the dated and signed (or sealed, where appropriate per local regulations), written IEC/IRB approval of the protocol, amendments, ICF, any recruiting materials, and if applicable, subject compensation programs. This approval must clearly identify the specific protocol by title and number and must be signed (or sealed, where appropriate per local regulations) by the chairman or authorized designee.
- Name and address of the IEC/IRB, including a current list of the IEC/IRB members and their function, with a statement that it is organized and operates according to GCP and the applicable laws and regulations. If accompanied by a letter of explanation, or equivalent, from the IEC/IRB, a general statement may be substituted for this list. If an investigator or a member of the study site personnel is a member of the IEC/IRB, documentation must be obtained to state that this person did not participate in the deliberations or in the vote/opinion of the study.
- Regulatory authority approval or notification, if applicable
- Signed and dated statement of investigator (eg, Form FDA 1572), if applicable
- Documentation of investigator qualifications (eg, curriculum vitae)
- Completed investigator financial disclosure form from the principal investigator, where required
- Signed and dated Clinical Trial Agreement, which includes the financial agreement
- Any other documentation required by local regulations

The following documents must be provided to the sponsor before enrollment of the first subject:

- Completed investigator financial disclosure forms from all subinvestigators
- Documentation of subinvestigator qualifications (eg, curriculum vitae)
- Name and address of any local laboratory conducting tests for the study, and a dated copy of current laboratory normal ranges for these tests, if applicable
- Local laboratory documentation demonstrating competence and test reliability (eg, accreditation/license), if applicable

#### 17.3. Subject Identification, Enrollment, and Screening Logs

The investigator agrees to complete a subject identification and enrollment log to permit easy identification of each subject during and after the study. This document will be reviewed by the sponsor study site contact for completeness.

The subject identification and enrollment log will be treated as confidential and will be filed by the investigator in the study file. To ensure subject confidentiality, no copy will be made. All reports and communications relating to the study will identify subjects by subject identification and date of birth. In cases where the subject is not randomized into the study, the date seen and date of birth will be used.

The investigator must also complete a subject screening log, which reports on all subjects who were seen to determine eligibility for inclusion in the study.

#### 17.4. Source Documentation

At a minimum, source documents consistent in the type and level of detail with that commonly recorded at the study site as a basis for standard medical care must be available for the following: subject identification, eligibility, and study identification; study discussion and date of signed informed consent; dates of visits; results of safety and efficacy parameters as required by the protocol; record of all AEs and follow-up of AEs; concomitant medication; drug receipt/dispensing/return records; study drug administration information; and date of study completion and reason for early discontinuation of study drug or withdrawal from the study, if applicable.

The author of an entry in the source documents should be identifiable.

At a minimum, the type and level of detail of source data available for a subject should be consistent with that commonly recorded at the study site as a basis for standard medical care. Specific details required as source data for the study and source data collection methods will be reviewed with the investigator before the study and will be described in the monitoring guidelines (or other equivalent document).

The minimum source documentation requirements for Section 4.1, Inclusion Criteria, and Section 4.2, Exclusion Criteria, that specify a need for documented medical history are as follows:

- Referral letter from treating physician or
- Complete history of medical notes at the site
- Discharge summaries

Inclusion and exclusion criteria not requiring documented medical history must be verified at a minimum by subject interview or other protocol required assessment (eg, physical examination, laboratory assessment) and documented in the source documents.

An electronic source system may be utilized, which contains data traditionally maintained in a hospital or clinic record to document medical care (eg, electronic source documents) as well as the clinical study-specific data fields as determined by the protocol. This data is electronically extracted for use by the sponsor. If an electronic source is utilized, references made to the CRF in the protocol include the electronic source system but information collected through the electronic source may not be limited to that found in the CRF. Data in this system may be considered source documentation.

#### 17.5. Case Report Form Completion

Case report forms are prepared and provided by the sponsor for each subject in electronic format. All data relating to the study must be recorded in the CRF. All CRF entries, corrections, and alterations must be made by the investigator or authorized study site personnel. The investigator must verify that all data entries in the CRF are accurate and correct.

The study data will be transcribed by study site personnel from the source documents onto an electronic CRF, if applicable. Study-specific data will be transmitted in a secure manner to the sponsor.

Worksheets may be used for the capture of some data to facilitate completion of the CRF. Any such worksheets will become part of the subject's source documents. Data must be entered into CRF in English. The CRF must be completed as soon as possible after a subject visit and the forms should be available for review at the next scheduled monitoring visit.

All subjective measurements (eg, pain scale information or other questionnaires) will be completed by the same individual who made the initial baseline determinations whenever possible.

If necessary, queries will be generated in the electronic data capture (eDC) tool. If corrections to a CRF are needed after the initial entry into the CRF, this can be done in either of the following ways:

- Investigator and study site personnel can make corrections in the eDC tool at their own initiative or as a response to an auto query (generated by the eDC tool).
- Sponsor or sponsor delegate can generate a query for resolution by the investigator and study site personnel.

#### 17.6. Data Quality Assurance/Quality Control

Steps to be taken to ensure the accuracy and reliability of data include the selection of qualified investigators and appropriate study sites, review of protocol procedures with the investigator and study site personnel before the study, periodic monitoring visits by the sponsor, and direct transmission of clinical laboratory data from a central laboratory into the sponsor's data base. Written instructions will be provided for collection, handling, storage, and shipment of samples.

Guidelines for CRF completion will be provided and reviewed with study site personnel before the start of the study.

The sponsor will review CRF for accuracy and completeness during on-site monitoring visits and after transmission to the sponsor; any discrepancies will be resolved with the investigator or designee, as appropriate. After upload of the data into the study database they will be verified for accuracy and consistency with the data sources.

#### 17.7. Record Retention

In compliance with the ICH/GCP guidelines, the investigator/institution will maintain all CRF and all source documents that support the data collected from each subject, as well as all study documents as specified in ICH/GCP Section 8, Essential Documents for the Conduct of a Clinical Trial, and all study documents as specified by the applicable regulatory requirement(s). The investigator/institution will take measures to prevent accidental or premature destruction of these documents.

Essential documents must be retained until at least 2 years after the last approval of a marketing application in an ICH region and until there are no pending or contemplated marketing applications in an ICH region or until at least 2 years have elapsed since the formal discontinuation of clinical development of the investigational product. These documents will be retained for a longer period if required by the applicable regulatory requirements or by an agreement with the sponsor. It is the responsibility of the sponsor to inform the investigator/institution as to when these documents no longer need to be retained.

If the responsible investigator retires, relocates, or for other reasons withdraws from the responsibility of keeping the study records, custody must be transferred to a person who will accept the responsibility. The sponsor must be notified in writing of the name and address of the new custodian. Under no circumstance shall the investigator relocate or dispose of any study documents before having obtained written approval from the sponsor.

If it becomes necessary for the sponsor or the appropriate regulatory authority to review any documentation relating to this study, the investigator/institution must permit access to such reports.

#### 17.8. Monitoring

The sponsor will perform on-site monitoring visits as frequently as necessary. The monitor will record dates of the visits in a study site visit log that will be kept at the study site, as allowed by local regulation. The first post-initiation visit will be made as soon as possible after enrollment has begun. At these visits, the monitor will compare the data entered into the CRF with the source documents (eg, hospital/clinic/physician’s office medical records). The nature and location of all source documents will be identified to ensure that all sources of original data required to complete the CRF are known to the sponsor and study site personnel and are accessible for verification by the sponsor study site contact. If electronic records are maintained at the study site, the method of verification must be discussed with the study site personnel.

Direct access to source documents (medical records) must be allowed for the purpose of verifying that the recorded data are consistent with the original source data. Findings from this review will be discussed with the study site personnel. The sponsor expects that, during monitoring visits, the relevant study site personnel will be available, the source documents will be accessible, and a suitable environment will be provided for review of study-related documents. The monitor will meet with the investigator on a regular basis during the study to provide feedback on the study conduct.

In addition to on-site monitoring visits, remote contacts can occur. It is expected that during these remote contacts, study site personnel will be available to provide an update on the progress of the study at the site.

#### 17.9. Study Completion/Termination

##### 17.9.1. Study Completion/End of Study

The study is considered completed with the last efficacy and safety follow-up visit for the last subject participating in the study. The final data from the study site will be sent to the sponsor (or designee) after completion of the final subject visit at that study site, in the time frame specified in the Clinical Trial Agreement.

##### 17.9.2. Study Termination

The sponsor reserves the right to close the study site or terminate the study at any time for any reason at the sole discretion of the sponsor. Study sites will be closed upon study completion. A study site is considered closed when all required documents and study supplies have been collected and a study site closure visit has been performed.

The investigator may initiate study site closure at any time, provided there is reasonable cause and sufficient notice is given in advance of the intended termination.

Reasons for the early closure of a study site by the sponsor or investigator may include but are not limited to:

- Failure of the investigator to comply with the protocol, the requirements of the IEC/IRB or local health authorities, the sponsor's procedures, or GCP guidelines
- Inadequate recruitment of subjects by the investigator
- Discontinuation of further study drug development

#### 17.10. On-Site Audits

Representatives of the sponsor's clinical quality assurance department may visit the study site at any time during or after completion of the study to conduct an audit of the study in compliance with regulatory guidelines and company policy. These audits will require access to all study records, including source documents, for inspection. Subject privacy must, however, be respected. The investigator and study site personnel are responsible for being present and available for consultation during routinely scheduled study site audit visits conducted by the sponsor or its designees.

Similar auditing procedures may also be conducted by agents of any regulatory body, either as part of a national GCP compliance program or to review the results of this study in support of a regulatory submission. The investigator should immediately notify the sponsor if he or she has been contacted by a regulatory agency concerning an upcoming inspection.

#### 17.11. Use of Information and Publication

All information, including but not limited to information regarding JNJ-64304500 or the sponsor's operations (eg, patent application, formulas, manufacturing processes, basic scientific data, prior clinical data, formulation information) supplied by the sponsor to the investigator and not previously published, and any data, including pharmacogenomic or exploratory biomarker research data, generated as a result of this study, are considered confidential and remain the sole property of the sponsor. The investigator agrees to maintain this information in confidence and use this information only to accomplish this study and will not use it for other purposes without the sponsor's prior written consent.

The investigator understands that the information developed in the study will be used by the sponsor in connection with the continued development of JNJ-64304500, and thus may be disclosed as required to other clinical investigators or regulatory agencies. To permit the information derived from the clinical studies to be used, the investigator is obligated to provide the sponsor with all data obtained in the study.

The results of the study will be reported in a Clinical Study Report generated by the sponsor and will contain data from all study sites that participated in the study as per protocol. Recruitment performance or specific expertise related to the nature and the key assessment parameters of the study will be used to determine a coordinating investigator. Results of pharmacogenomic or exploratory biomarker analyses performed after the Clinical Study Report has been issued will be reported in a separate report and will not require a revision of the Clinical Study Report. Study subject identifiers will not be used in publication of results. Any work created in connection with performance of the study and contained in the data that can benefit from copyright protection (except any publication by the investigator as provided for below) shall be the property of the sponsor as author and owner of copyright in such work.

Consistent with Good Publication Practices and International Committee of Medical Journal Editors guidelines, the sponsor shall have the right to publish such primary (multicenter) data and information without approval from the investigator. The investigator has the right to publish study site-specific data after the primary data are published. If an investigator wishes to publish information from the study, a copy of the manuscript must be provided to the sponsor for review at least 60 days before submission for publication or presentation. Expedited reviews will be arranged for abstracts, poster presentations, or other materials. If requested by the sponsor in writing, the investigator will withhold such publication for up to an additional 60 days to allow for filing of a patent application. In the event that issues arise regarding scientific integrity or regulatory compliance, the sponsor will review these issues with the investigator. The sponsor will not mandate modifications to scientific content and does not have the right to suppress information. For multicenter study designs and substudy approaches, secondary results generally should not be published before the primary endpoints of a study have been published. Similarly, investigators will recognize the integrity of a multicenter study by not submitting for publication data derived from the individual study site until the combined results from the completed study have been submitted for publication, within 12 months of the availability of the final data (tables, listings, graphs), or the sponsor confirms there will be no multicenter study publication. Authorship of publications resulting from this study will be based on the guidelines on authorship, such as those described in the Uniform Requirements for Manuscripts Submitted to Biomedical Journals, which state that the named authors must have made a significant contribution to the design of the study or analysis and interpretation of the data, provided critical review of the paper, and given final approval of the final version.

**Registration of Clinical Studies and Disclosure of Results**

The sponsor will register and disclose the existence of and the results of clinical studies as required by law.

### ATTACHMENTS

#### Attachment 1: Definition of Inadequate Initial Response, Loss of Response, or Intolerance to TNF-Antagonist Therapies (Infliximab, Adalimumab, or Certolizumab Pegol) or Vedolizumab

The criteria for inadequate initial response, response followed by loss of response, or intolerance to infliximab, adalimumab, certolizumab pegol, or vedolizumab are described in items I, II, and III, below.

**I. Inadequate initial response to current or prior therapy with infliximab, adalimumab, certolizumab pegol (primary nonresponse), or vedolizumab (primary nonresponse)**

Eligible subjects must satisfy criteria A, B, and C.

**A. Have received induction doses of:**

- Infliximab (2 or 3 doses of ≥ 5 mg/kg)

  or

- Adalimumab (at a dose of 160 mg followed by a dose ≥ 80 mg **or** at a dose of 80 mg followed by a dose ≥ 40 mg)

  or

- Certolizumab pegol (2 or 3 doses of ≥ 400 mg)

  or

- Vedolizumab (3 or 4 doses of 300 mg)

**AND**

**B. Did not initially respond to these induction doses of infliximab, adalimumab, certolizumab pegol, or vedolizumab, as evidenced by the presence of at least 1 of the following signs or symptoms related to persistence of Crohn's disease, as assessed by a treating physician:**

- Lack of improvement or worsening in stool frequency
- Lack of improvement or worsening in daily abdominal pain
- Occurrence, lack of improvement, or worsening of fever thought to be related to Crohn's disease
- Lack of improvement or worsening in a draining fistula or development of a new draining fistula
- Lack of improvement or worsening in rectal bleeding
- Initiation or increase in antidiarrheal medication

These signs and symptoms of Crohn's disease must have occurred ≥2 weeks after receiving the last induction dose of infliximab, adalimumab, certolizumab pegol, or vedolizumab, and are offered only as a benchmark of the minimally acceptable criteria required to designate a subject as having had an inadequate initial response to infliximab, adalimumab, certolizumab pegol, or vedolizumab therapy. This benchmark acknowledges that the CDAI is not routinely recorded in clinical practice.

**AND**

**C. Have documentation available to the investigator that meets the following 2 requirements:**

- Provide the dates and doses of the failed infliximab, adalimumab, certolizumab pegol, or vedolizumab induction therapy.
- Documents that the subject had persistence of disease activity following infliximab, adalimumab, certolizumab pegol, or vedolizumab induction therapy.

Examples of acceptable documents include: medical records, letter provided by a referring physician, or other "reason for referral" documents (eg, insurance authorization form).

**II. Initial response followed by loss of response to current or prior therapy with infliximab, adalimumab, certolizumab pegol, or vedolizumab (secondary nonresponse)**

Eligible subjects must satisfy criteria A, B, C, and D.

**A. Initially responded to induction therapy**

**AND**

**B. Have received at least 2 maintenance doses of:**

- Infliximab (at a dose of ≥5 mg/kg)

  or

- Adalimumab (at a dose of ≥40 mg)

  or

- Certolizumab pegol (at a dose of ≥400 mg)

  or

- Vedolizumab (at a dose of ≥300 mg)

**AND**

**C. Have or had at least 1 of the following signs or symptoms related to recurrence of Crohn's disease, as assessed by a treating physician:**

- Worsening in stool frequency
- Worsening in daily abdominal pain
- Occurrence or worsening in fever thought to be related to Crohn's disease
- Recurring drainage from a previously nondraining fistula or development of a new draining fistula
- Worsening in rectal bleeding
- Initiation or increase in antidiarrheal medication

These signs and symptoms of Crohn's disease must have occurred ≥2 weeks after receiving the last maintenance dose of infliximab, adalimumab, certolizumab pegol, or vedolizumab, and are offered only as a benchmark of the minimally acceptable criteria required to designate a subject as having lost response to infliximab, adalimumab, certolizumab pegol, or vedolizumab therapy. This benchmark acknowledges that the CDAI is not routinely recorded in clinical practice.

**AND**

**D. Have documentation available to the investigator that meets the following 2 requirements:**

- Provide the dates and doses of the failed infliximab, adalimumab, certolizumab pegol, or vedolizumab maintenance therapy.
- Documents that the subject had recurrence of disease activity despite infliximab, adalimumab, certolizumab pegol, or vedolizumab maintenance therapy.

Examples of acceptable documents include: medical records, letter provided by a referring physician, or other "reason for referral" documents (eg, insurance authorization form).

**III. Current or prior intolerance to therapy with infliximab, adalimumab, certolizumab pegol, or vedolizumab**

Eligible subjects must satisfy criteria A and B.

**A. Have had an adverse reaction that meets 1 of the following 3 criteria: 1) significant acute infusion/administration reaction; 2) significant delayed infusion/administration reaction (for example, delayed hypersensitivity or serum-sickness like reaction); or 3) significant injection-site reaction. Definitions of these 3 criteria are provided below. Adverse reactions also must have followed ≥1 dose of infliximab, adalimumab, certolizumab pegol,**

**or vedolizumab, and, in the treating physician's opinion, precluded continued use of the therapy.**

**1) A significant acute infusion/administration reaction is defined as an adverse reaction that was:**

- Manifested through ≥ 1 of the following symptoms.
  - a. Fever greater than 100°F (37.8°C)
  - b. Chills or rigors
  - c. Itching
  - d. Rash
  - e. Flushing
  - f. Urticaria or angioedema
  - g. Breathing difficulties (dyspnea, chest paint or tightness, shortness of breath, wheezing, stridor)
  - h. Clinical hypotension (pallor, diaphoresis, faintness, syncope), blood pressure less than 90 mm Hg systolic and 60 mm Hg diastolic, or a systemic or orthostatic drop in systolic blood pressure of greater than 20 mm Hg

  **AND**

- Occurred ≤24 hours after infusion/administration of infliximab, adalimumab, certolizumab pegol, or vedolizumab

  **AND**

- Was considered related to the infusion/administration of infliximab, adalimumab, certolizumab pegol, or vedolizumab.

**2) A significant delayed infusion/administration reaction is defined as an adverse reaction that:**

- Was manifested through 1 or more of the following symptoms:
  - a. Myalgias
  - b. Arthralgias
  - c. Fever greater than 100°F (37.8°C)
  - d. Malaise
  - e. Rash

  **AND**

- Occurred >24 hours and <15 days after infusion/administration of infliximab, adalimumab, certolizumab pegol, or vedolizumab

**AND**

- Was considered related to the infusion/administration of infliximab, adalimumab, certolizumab pegol, or vedolizumab.

**3) A significant injection-site reaction is defined as an adverse reaction that:**

- Was manifested through 1 or more of the following symptoms:
    a. Significant bruising
    b. Erythema
    c. Hemorrhage
    d. Irritation
    e. Pain
    f. Pruritus
    g. "Injection-site reaction"

**AND**

- Occurred within 24 hours of an SC injection of adalimumab or certolizumab pegol.

**AND**

- Was considered related to the injection.

**B. Have documentation available to the investigator that meets the following 2 requirements:**

- Provides the date of discontinuation of infliximab, adalimumab, certolizumab pegol, or vedolizumab.
- Documents that the subject had intolerance to infliximab, adalimumab, certolizumab pegol, or vedolizumab.

Examples of acceptable documents include: medical records, letter provided by a referring physician, or other "reason for referral" documents (eg, insurance authorization form).

**Attachment 2: Definitions of Inadequate Response to or Intolerance of Corticosteroids or AZA/6-MP/MTX and Corticosteroid Dependence**

#### CORTICOSTEROIDS

**Subjects have failed to respond to corticosteroids if** they have had evidence of an initial inadequate response, recurrent disease, or a relapse despite receiving at least 0.75 mg/kg/day or ≥40 mg/day of prednisone (or corticosteroid equivalent, given orally or intravenously) for 2 weeks; or ≥9 mg/day of budesonide or ≥5 mg/day of beclomethasone dipropionate given orally for at least 4 weeks.

**Subjects are intolerant of corticosteroids if:**

- They have developed clinically significant adverse events (eg, osteonecrosis or osteoporosis, psychosis, uncontrolled diabetes) unresponsive to dose reduction that, in the judgment of the investigator, precluded the use of corticosteroids to treat Crohn's disease.
- **OR**
- They have a medical condition that precludes the use of corticosteroids as a treatment for Crohn's disease.

**Subjects are corticosteroid dependent if** they have failed to successfully taper their corticosteroid (ie, had a flare of disease) within 3 months of starting therapy, or if a relapse occurs within 3 months after stopping corticosteroids or if they are unable to discontinue these agents without flare within 3 months after starting them.

#### 6-MERCAPTOPURINE (6-MP), AZATHIOPRINE (AZA), OR METHOTREXATE (MTX):

**Subjects have failed to respond to 6-MP, AZA, or MTX if** they have had evidence of an initial inadequate response, recurrent disease, or a relapse despite receiving:

- At least 3 months of therapy with 1 mg/kg/day of 6-MP, 2 mg/kg/day of AZA, or 25 mg/week (intramuscular or subcutaneous) of MTX.

  **OR**
- A lower dosage of 6-MP, AZA, or MTX when country or local guidelines specify a different treatment regimen. (In such an event, the country or local guidelines needs to be included in the source document).

  **OR**
- The dosage of 6-MP, AZA, or MTX confirmed to be therapeutic for the subject with whole blood thioguanine nucleotide levels >200 pmole/$8x10^8$ red blood cells.

  **OR**
- The highest tolerated dosage due to leukopenia, elevated liver enzymes, or nausea.

**Subjects are intolerant of 6-MP, AZA, or MTX if:**

- They have developed clinically significant adverse events (eg, pancreatitis, arthritis accompanied by high fever and/or rash, leukopenia, or persistently elevated liver enzymes) unresponsive to dose reduction that, in the judgment of the investigator, precluded the use of 6-MP, AZA, or MTX to treat Crohn's disease within the past 5 years.

  **OR**

- They have a medical condition that precludes the use of 6-MP, AZA, or MTX.

#### Attachment 3: Tuberculin Skin Testing

##### Administering the Mantoux Tuberculin Skin Test

The Mantoux tuberculin skin test (CDC, 2000) is the standard method of identifying persons infected with *Mycobacterium tuberculosis*. Multiple puncture tests (Tine and Heaf) should not be used to determine whether a person is infected because the amount of tuberculin injected intradermally cannot be precisely controlled. Tuberculin skin testing is both safe and reliable throughout the course of pregnancy. The Mantoux tuberculin test is performed by placing an intradermal injection of 0.1 mL of tuberculin into the inner surface of the forearm. The test must be performed with tuberculin that has at least the same strength as either 5 tuberculin units (TU) of standard purified protein derivative (PPD)-S or 2 TU of PPD-RT 23, Statens Seruminstitut, as recommended by the World Health Organization. PPD strengths of 1 TU or 250 TU are not acceptable (Menzies, 2000). Using a disposable tuberculin syringe with the needle bevel facing upward, the injection should be made just beneath the surface of the skin. This should produce a discrete, pale elevation of the skin (a wheal) 6 mm to 10 mm in diameter. To prevent needle-stick injuries, needles should not be recapped, purposely bent or broken, removed from disposable syringes, or otherwise manipulated by hand. After they are used, disposable needles and syringes should be placed in puncture-resistant containers for disposal. Institutional guidelines regarding universal precautions for infection control (eg, the use of gloves) should be followed. A trained health care worker, preferably the investigator, should read the reaction to the Mantoux test 48 to 72 hours after the injection. Subjects should never be allowed to read their own tuberculin skin test results. If a subject fails to show up for the scheduled reading, a positive reaction may still be measurable up to 1 week after testing. However, if a subject who fails to return within 72 hours has a negative test, tuberculin testing should be repeated. The area of induration (palpable raised hardened area) around the site of injection is the reaction to tuberculin. For standardization, the diameter of the induration should be measured transversely (perpendicular) to the long axis of the forearm. Erythema (redness) should not be measured. All reactions should be recorded in millimeters, even those classified as negative.

##### Interpreting the Tuberculin Skin Test Results

In the US and many other countries, the most conservative definition of positivity for the tuberculin skin test is reserved for immunocompromised patients, and this definition is to be applied in this study to maximize the likelihood of detecting latent TB, even though the subjects may not be immunocompromised at baseline.

In the US and Canada, an induration of 5 mm or greater in response to the intradermal tuberculin skin test is considered to be a positive result and evidence for either latent or active TB.

In countries outside the US and Canada, country-specific guidelines **for immunocompromised patients** should be consulted for the interpretation of tuberculin skin test results. If no local country guidelines for immunocompromised patients exist, US guidelines must be followed.

##### Treatment of Latent Tuberculosis

Local country guidelines **for immunocompromised patients** should be consulted for acceptable antituberculous treatment regimens for latent TB. If no local country guidelines for immunocompromised patients exist, US guidelines must be followed.

**NOTE:** The tuberculin skin test is recommended but not required in countries where tuberculin is not available.

#### Attachment 4: Crohn's Disease Activity Index

| DISEASE ACTIVITY INDEX | SUM | X FACTOR | | SUBTOTAL |
|---|---|---|---|---|
| Total number of liquid or very soft stools in the previous 7 days | _______ | x 2 | = | _______ |
| Sum abdominal pain/cramps ratings (total for previous 7 days):<br>0 = none 2 = moderate<br>1 = mild 3 = severe | _______ | x 5 | = | _______ |
| General well-being (total for previous 7 days):<br>0 = generally well 3 = very poor<br>1 = slightly under par 4 = terrible<br>2 = poor | _______ | x 7 | = | _______ |
| Categories currently present and <u>presumed to be related to Crohn's disease:</u> 0 = no; 1 = yes | | | | |
| □ = arthritis/arthralgia | _______ | x 20 | = | _______ |
| □ = iritis/uveitis | _______ | x 20 | = | _______ |
| □ = erythema nodosum/pyoderma gangrenosum/aphthous stomatitis | _______ | x 20 | = | _______ |
| □ = anal fissure, fistula or abscess | _______ | x 20 | = | _______ |
| □ = other fistula | _______ | x 20 | = | _______ |
| □ = fever over 100° F (37.8° C) during the previous 7 days. | _______ | x 20 | = | _______ |
| During the previous 7 days has subject received antidiarrheal therapy at least once:<br>**OR**<br>During the previous 7 days has subject received opiate therapy on each of the 7 days:<br>0 = no<br>1 = yes | _______ | x 30 | = | _______ |
| Abdominal mass:<br>0 = none 2 = questionable 5 = definite | _______ | x 10 | = | _______ |
| Hematocrit:<br>Males: (47-Hct) = SUM<br>Females: (42-Hct) = SUM | _______<br>(add or subtract by sign) | x 6 | = | _______ |
| (Standard Weight - Actual Body Weight) / Standard Weight x 100 = | * _______<br>(add or subtract by sign, round to 3 decimal places) | x 1 | = | _______ |

* If this value is less than -10 then enter -10 here.
Standard weight and actual weight must be in same units (kg or lb)

TOTAL = _______
(round total to integer)

##### Attachment 5: Liver Safety Monitoring and Assessment

If laboratory testing for a subject enrolled in study and receiving study drug reveals an increase of serum aminotransferases (ALT or AST) to >3X ULN and bilirubin >2X ULN, laboratory tests for ALT, AST, alkaline phosphatase, and total bilirubin should be confirmed by a retest within 24 hours if possible, but no later than 72 hours following notification of test results. Testing should be repeated within 24 hours but no later than 72 hours following notification of the test results. For studies for which a central laboratory is used, alerts will be generated by the central laboratory regarding moderate and marked liver abnormality to inform the investigator, study monitor, and study team. Subjects should be asked if they have any symptoms suggestive of hepatobiliary dysfunction.

#### DEFINITION OF LIVER ABNORMALITIES

Confirmed abnormalities will be characterized as moderate and marked where ULN:

| | ALT or AST | | Total Bilirubin |
|---|---|---|---|
| **Moderate** | >3x ULN | or | >2x ULN |
| **Marked** | >3x ULN | and | >2x ULN |

In addition, the subject should be considered to have marked hepatic abnormalities for any of the following:

- ALT or AST >8x ULN.
- ALT or AST >5x ULN for more than 2 weeks.
- ALT or AST >3x ULN and international normalized ratio (INR) >1.5.
- ALT or AST >3x ULN with the appearance of fatigue, nausea, vomiting, right upper quadrant pain or tenderness, fever, rash, and/or eosinophilia (>5%).

The investigator may determine that abnormal liver function results, other than as described above, may qualify as moderate or marked abnormalities and require additional monitoring and follow-up.

#### FOLLOW-UP PROCEDURES

Confirmed moderate and marked abnormalities in hepatic functions should be thoroughly characterized by obtaining appropriate expert consultations, detailed pertinent history, physical examination, and laboratory tests. The site should complete the Liver Abnormality Case Report Form (LA-CRF). Subjects with confirmed abnormal liver function testing should be followed as described below.

Confirmed moderately abnormal liver function tests (LFTs) should be repeated 2 to 3 times weekly then weekly or less if abnormalities stabilize or the trial drug has been discontinued and the subject is asymptomatic.

Marked hepatic liver function abnormalities, in the absence of another etiology, may be considered an important medical event and reported as an SAE. The medical monitor should be contacted and informed of all subjects for whom marked hepatic liver function abnormalities possibly attributable to study drug are observed.

To further assess abnormal hepatic laboratory findings, the investigator is expected to:

- Obtain a more detailed history of symptoms and prior or concurrent diseases. Symptoms and new onset-diseases should be recorded as 'adverse events' on the AE sheet. Illnesses and conditions such as hypotensive events, and decompensated cardiac disease than may lead to secondary liver abnormalities should be noted. Non-alcoholic steatohepatitis (NASH) is seen in obese hyperlipoproteinemic, and/or diabetic patients and may be associated with fluctuating aminotransferase levels. The investigator should ensure that the medical history form captures any illness that predates study enrollment that may be relevant in assessing hepatic function.
- Obtain a history of concomitant drug use (including nonprescription medication, complementary, and alternative medications), alcohol use, recreational drug use, and special diets. Medications, including dose, should be entered on the concomitant medication form. Information on alcohol, other substance use, and diet should be entered on the LA-CRF.
- Obtain a history of exposure to environmental chemical agents.
- Based on the subject's history, other testing may be appropriate including:
  - acute viral hepatitis (A, B, C, D, E or other infectious agents).
  - ultrasound or other imaging to assess biliary tract disease.
  - other laboratory tests including INR, direct bilirubin.
- Consider gastroenterology or hepatology consultations.
- Submit results for any additional testing and possible etiology on the LA-CRF.

**STUDY AGENT SUSPENSION and/or DISCONTINUATION**

If laboratory testing reveals an increase of serum aminotransferases (ALT or AST) to >3x ULN and an increase of bilirubin to >2x ULN, study agent should be suspended immediately. Suspension of study agent should also be considered if:

- ALT or AST >8x ULN.
- ALT or AST >5x ULN for more than 2 weeks.
- ALT or AST >3x ULN and INR >1.5.
- ALT or AST >3x ULN with the appearance of fatigue, nausea, vomiting, right upper quadrant pain or tenderness, fever, rash, and/or eosinophilia (>5%).

In the absence of an explanation for increased LFTs, such as viral hepatitis, pre-existing or acute liver disease, or exposure to other agents associated with liver injury, the subject may be discontinued from the study agent. The investigator may determine that it is not in the subject's best interest to continue study agent.

In addition, if close monitoring for a subject with moderate or marked hepatic laboratory tests is not possible, study agent should be discontinued.

Reference

Guidance for Industry titled "Drug-Induced Liver Injury: Premarketing Clinical Evaluation" issued by FDA on July 2009.

#### Attachment 6: Anticipated Events

##### Anticipated Event

An anticipated event is an adverse event (serious or nonserious) that commonly occurs as a consequence of the underlying disease or condition under investigation (disease-related) or background regimen.

For the purposes of this study the following events will be considered anticipated events:

- Adverse events related to symptoms of Crohn’s disease
- Adverse events related to worsening or progression of Crohn’s disease

##### Reporting of Anticipated Events

These events will be captured on the CRF and in the database, and will be reported to the sponsor as described in Section 12.3.1, All Adverse Events. Any event that meets serious adverse event criteria will be reported to the sponsor within the appropriate timeline as described in Section 12.3.2, Serious Adverse Events. These anticipated events are exempt from expedited reporting as individual single cases to Health Authorities. However, if based on an aggregate review, it is determined that an anticipated event is possibly related to study drug, the sponsor will report these events in an expedited manner.

##### Anticipated Event Review Committee (ARC)

An Anticipated Event Review Committee (ARC) will be established to perform reviews of prespecified anticipated events at an aggregate level. The ARC is a safety committee within the sponsor’s organization that is independent of the sponsor’s study team. The ARC will meet to aid in the recommendation to the sponsor’s study team as to whether there is a reasonable possibility that an anticipated event is related to the study drug.

##### Statistical Analysis

Details of statistical analysis of anticipated events, including the frequency of review and threshold to trigger an aggregate analysis of anticipated events will be provided in a separate Anticipated Events Safety Monitoring Plan (ASMP).

#### INVESTIGATOR AGREEMENT

I have read this protocol and agree that it contains all necessary details for carrying out this study. I will conduct the study as outlined herein and will complete the study within the time designated.

I will provide copies of the protocol and all pertinent information to all individuals responsible to me who assist in the conduct of this study. I will discuss this material with them to ensure that they are fully informed regarding the study drug, the conduct of the study, and the obligations of confidentiality.

**Coordinating Investigator (where required):**

Name (typed or printed): ____________________

Institution and Address: ____________________

____________________

____________________

____________________

Signature: ____________________ Date: ____________________
(Day Month Year)

**Principal (Site) Investigator:**

Name (typed or printed): ____________________

Institution and Address: ____________________

____________________

____________________

____________________

Telephone Number: ____________________

Signature: ____________________ Date: ____________________
(Day Month Year)

**Sponsor's Responsible Medical Officer:**

Name (typed or printed): Newman Yeilding, MD

Institution: Janssen Research & Development

Signature: ____________________ Date: 4-FEB-2020
(Day Month Year)

**Note:** If the address or telephone number of the investigator changes during the course of the study, written notification will be provided by the investigator to the sponsor, and a protocol amendment will not be required.

172

Approved, Date: 4 February 2020
